# Twelve-Month All-Cause Mortality after Initial COVID-19 Vaccination with Pfizer-BioNTech or mRNA-1273 among Adults Living in Florida

**DOI:** 10.1101/2025.04.25.25326460

**Authors:** Retsef Levi, Fahad Mansuri, Melissa M. Jordan, Joseph A. Ladapo

## Abstract

Comparative effects of the mRNA COVID-19 vaccines on mortality and other health outcomes are uncertain. Matched cohort analysis of 1,962,822 adults living in Florida who received a first dose of either BNT162b2 (Pfizer) or mRNA-1273 (Moderna) between December 18, 2020, and August 31, 2021, was used to examine 12-month risk of all-cause, cardiovascular, COVID-19, and non-COVID-19 mortality. Matching was performed on seven criteria, including census tract. Compared with mRNA-1273 recipients, BNT162b2 recipients had significantly higher risk for all-cause mortality (783.6 vs. 562.4 deaths per 100,000; odds ratio, OR [95% CI]: 1.407 [1.358, 1.457]), cardiovascular mortality, COVID-19 mortality and non-COVID-19 mortality. Negative control outcomes and robustness analyses did not show any indication of meaningful unobserved residual confounding that is likely to alter the results. The results of this study are consistent with cumulative evidence from prior literature showing worse outcomes in recipients of BNT162b2 compared to mRNA-1273.

**Primary Funding Source:** The study did not receive any external funding.

## INTRODUCTION

A growing number of both randomized and observational studies suggest vaccines may have beneficial or harmful non-specific effects, including potential impact on all-cause mortality, beyond their effect on the target disease.^1,2^ In the context of the initial messenger RNA (mRNA) COVID-19 vaccination series, little is known about their effect on all-cause mortality. Pooled data from the initial, pivotal randomized trials evaluating BNT162b2 (Pfizer BioNTech) and mRNA-1273 (Moderna) COVID-19 vaccines were underpowered to detect differences in overall mortality compared to placebo. Moreover, no trial was conducted to compare the relative effects of different vaccines. While findings from these trials suggested that both vaccines reduced deaths due to COVID-19, pooled all-cause deaths were not reduced. Cardiac deaths were more frequent, with a relative but statistically insignificant increase of 50% and 40% compared to the placebo groups among BNT162b2 and mRNA-1273 vaccine recipients, respectively.^3^

Nonspecific vaccine effects can be directly assessed in observational studies by evaluating a vaccine’s impact on all-cause mortality and other outcomes. However, the results of these studies are prone to multiple biases, including healthy vaccinee bias, i.e., observed and unobserved characteristics that have a confounding effect on the decision to vaccinate and an outcome.^4^ As a result, they often have relatively low reliability. Moreover, healthy vaccinee bias, likely the result of substantial unobserved differences between vaccinated and unvaccinated individuals, were demonstrated in observational studies of COVID-19 vaccination, even in studies using matching or risk adjustment.^4–7^

Nonspecific vaccine effects may also be indirectly assessed by evaluating health outcomes among patients who receive different vaccines for the same indication.^8^ This type of comparison is less likely to be vulnerable to healthy vaccinee bias and has already been demonstrated for COVID-19 vaccines. A study using a target trial simulation design with data from the U.S. Department of Veterans Affairs, showed that initial vaccination with BNT162b2 was associated with significantly higher risk of multiple cardiovascular events compared to initial vaccination with mRNA-1273, over a 266-day follow-up period.^9^ Another study based on Veterans Affairs data using a similar methodological approach found improved outcomes against COVID-19 infection, hospitalization and intensive care unit admission among U.S. veterans who received a first dose of mRNA-1273 versus BNT162b2, but no statistically significant difference in non-COVID-19 mortality.^10^ To further assess comparative effects of the BNT162b2 vaccine and mRNA-1273 vaccine on 12-month all-cause, cardiovascular, COVID-19, and non-COVID-19 mortality, this study evaluated mortality outcomes among a cohort of adult Florida residents.

## METHODS

### Data

This study analyzed data from Florida’s state-level public health databases with records about COVID-19 vaccination, sociodemographic characteristics of vaccine recipients, and vital statistics. Florida SHOTS is a statewide, centralized online immunization information system (IIS) with immunization records for patients of all ages.^11^ As a program of the Florida Health Immunization Section, it is authorized by Section 381.003, Florida Statutes. The Bureau of Vital Statistics collects vital records data (birth, deaths, marriages, etc.) at the Florida Department of Health. The vital records data for deaths includes primary and underlying cause of death, health-related and other information about individuals residing in Florida at the time of their death.^12^ This study was reviewed by the Florida Department of Health institutional review board and deemed exempt as public health practice.

### Study Population and Design

The study population included noninstitutionalized adult Florida residents (ages 18-99 years) receiving a first dose of either the BNT162b2 or mRNA-1273 mRNA vaccine between December 18, 2020, and August 31, 2021, inclusive. Institutionalized individuals living in nursing homes or correctional facilities, as well as those identified as homeless in the Florida SHOTS database or with missing or unknown sex, were excluded from the analysis (See Supplement for the method used to identify nursing home residents and correctional facility residents). To minimize baseline differences between BNT162b2 and mRNA-1273 vaccine recipients, a matched cohort was formed, with a 1:1 ratio and exact matching according to 5-year age bins, sex, race, ethnicity, vaccination site (9 types), calendar month of vaccination with first dose, and census tract of residential address. Vaccination sites included private pharmacies, county health departments, private physician clinics, federally qualified health centers, hospital sites, and other locations. The total number of census tracts was 4,832. The selected matching criteria were hypothesized to have a potential effect on the type of vaccine received or mortality risk.

### Outcomes

The primary outcome of the study was 12-month all-cause mortality in the matched cohort, defined as any death within 12 months after the first COVID-19 vaccine dose. The Florida Vital Statistics database was used to identify all deaths. A multistep algorithm was then used to match records from Florida SHOTS with death records from the Vital Statistics database. Details about the matching algorithm are available in the Supplement.

Secondary outcomes were cardiovascular, COVID-19, and non-COVID-19 deaths. The classification of subcategories of death was determined by the primary cause of the death on each decedent’s death certificate, based on International Classification of Diseases, Tenth Revision (ICD-10) codes (Supplement Table 1). Deaths were attributed to COVID-19 if a corresponding ICD-10 code was listed either as the primary or underlying cause of death on the death certificate. Decedents without an ICD-10 code corresponding to COVID-19 on their death certificates were classified as non-COVID-19 deaths.

### Negative Control Outcomes

To further evaluate the risk of residual confounding, deaths from suicide or homicide within 12 months of vaccination with the first dose were used a negative control outcome. This negative control outcome was chosen because it is unlikely to be affected by the type of vaccination an individual receives but may be affected by potential unobserved confounders of health outcomes. These deaths were defined based on their corresponding ICD-10 codes being listed as a primary cause of death on the death certificate. To further assess potential residual unobserved confounding factors, the occurrence of SARS-CoV-2 infection (based on documented positive tests) before December 20, 2020, the start of COVID-19 vaccination, was considered as an additional negative control outcome.

### Statistical Analysis

Kaplan–Meier cumulative incidence curves were used to compare the risk of mortality outcomes among BNT162b2 and mRNA-1273 vaccine recipients in the matched cohort. Further risk adjustment was conducted using logistic regression models with a binary outcome of 1 if death occurred within 12 months of vaccination and 0 otherwise. In addition to an indicator variable corresponding to the type of vaccine received (1 for BNT162b2 and 0 for mRNA-1273), the regression models included the covariates used for matching, as well as covariates corresponding to components of the census tract-level social vulnerability index scores (socioeconomic, housing and transportation components) and ZIP code-level five-year average mortality rate specific for the individual’s age, sex, and race. Further stratification was conducted with similar risk-adjusted regression models estimated on age groups 18-39 years-old, 40-59 years-old, and 60 years-old and older.

Negative control outcomes were assessed with similar logistic regressions. For the negative control outcome of SARS-CoV-2 infections, the standardized mean difference was calculated to assess balance among the matched cohort. As a post-hoc analysis, similar regression models with an additional covariate indicator that captures SARS-CoV-2 infection prior to receiving dose 1, were estimated. Additional post hoc robustness analyses excluded patients with SARS-CoV-2 infection prior to vaccination. To assess the plausibility of the hypothesis that differences in outcomes between the vaccines could be attributable to differential protective effects against SARS-CoV-2 infection, the analyses were repeated with a more expansive definition of COVID-19+ deaths. Specifically, COVID-19+ deaths were defined as deaths with COVID-19 listed either as the primary or underlying cause of death on the death certificate, or decedents with documented SARS-CoV-2 infection after their first vaccine dose. Decedents without COVID-19 on their death certificates or a SARS-CoV-2 infection after the first dose were classified as non-COVID-19+ deaths.

Additional sensitivity analysis was conducted to assess the robustness of the matched cohort analysis to the presence of residual unobserved confounding covariates. The sensitivity analysis estimates thresholds on the association levels (quantified through odds ratio) between a potential unobserved confounder and the exposure and outcome, respectively, that would render the test statistics of the study inference insignificant (i.e., p-value higher than 0.05). Upper bounds on the respective p-values were calculated using methods proposed by Rosenbaum for matched observational studies.^13^ (See Supplement for more detailed description.) We calculated confidence intervals using the Wald method. Statistical analyses were performed in SAS (version 9.4).

### Code Sharing

All codes and output files are available as a supplemental file.

### Patient and Public Involvement

No patients were directly involved in defining the research question, development of study methods, performance of the analyses, or reporting. However, discussions with community members raising concerns about differing safety and efficacy profiles of COVID-19 vaccines helped inform the research question and design.

## RESULTS

A total of 11,288,333 adult Florida residents met inclusion criteria with 6,575,206 BNT162b2 vaccine recipients and 4,713,127 mRNA-1273 vaccine recipients. Of those, a total of 1,962,822 vaccine recipients, including 981,411 recipients each for BNT162b2 and mRNA-1273, were matched (1:1) across all seven criteria (Figure 1).

**Figure 1.**
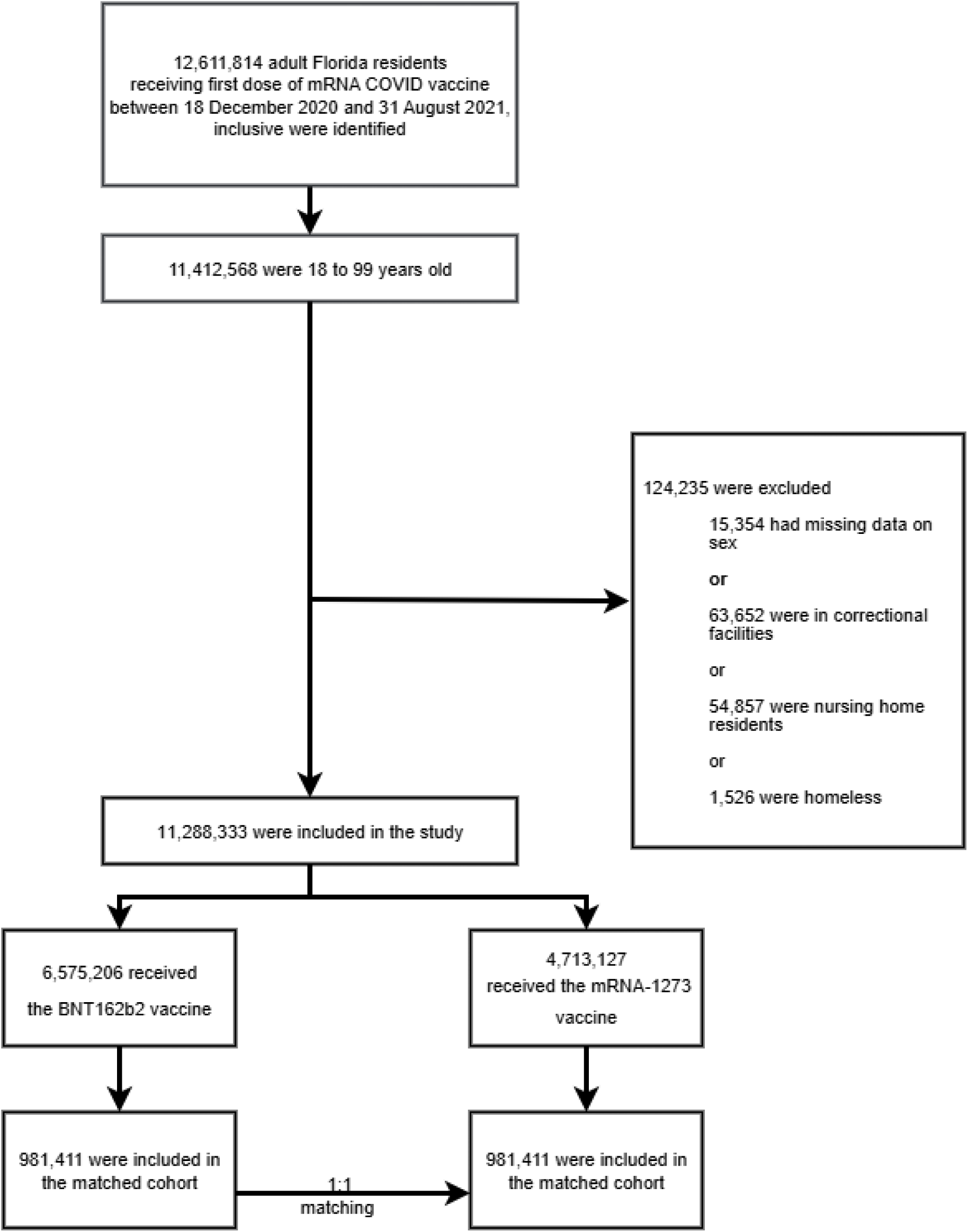
Selection of study participants.

Characteristics of vaccine recipients in the full and matched cohorts are shown in Table 1. In the full cohort, BNT162b2 recipients were younger, more likely to be Hispanic and had a higher social vulnerability index (SVI) than mRNA-1273 recipients. Compared to the full cohort, the matched cohort was slightly younger and had lower 12-month mortality rates.

**Table 1.** Demographic and vaccine characteristics of the study participants.

| Variables | Full cohort |  |  | Standardize<br>d Difference | Matched cohort |  |  | Standardize<br>d Difference |
| --- | --- | --- | --- | --- | --- | --- | --- | --- |
|  | BNT162b2<br>N (%) | mRNA-1273 N<br>(%) | Total N (%) |  | BNT162b2<br>N (%) | mRNA-1273<br>N (%) | Total N (%) |  |
| **Total** | 6,575,206<br>(58.3) | 4,713,127<br>(41.8) | 11,288,333 |  | 981,411 (50.0) | 981,411 (50.0) | 1,962,822 |  |
| **Age** |  |  |  | 0.2737 |  |  |  | 0.001 |
| Median (Q1,<br>Q3) | 52 (37, 66) | 60 (43, 71) | 56 (39, 69) |  | 56 (39, 67) | 56 (39, 67) | 56 (39, 67) |  |
| 18-39 | 1,906,280<br>(29.0) | 990,032 (21.0) | 2,896,312<br>(25.7) |  | 246,242 (25.1) | 246,242 (25.1) | 492,484 (25.1) |  |
| 40-49 | 1,050,560<br>(16.0) | 561,018 (11.9) | 1,611,578<br>(14.3) |  | 127,577 (13.0) | 127,577 (13.0) | 255,154 (13.0) |  |
| 50-59 | 1,186,370<br>(18.0) | 777,253 (16.5) | 1,963,623<br>(17.4) |  | 189,474 (19.3) | 189,474 (19.3) | 378,948 (19.3) |  |
| 60-69 | 1,152,961<br>(17.5) | 1,044,834<br>(22.2) | 2,197,795<br>(19.5) |  | 212,722 (21.7) | 212,722 (21.7) | 425,444 (21.7) |  |
| 70-79 | 844,380 (12.8) | 923,425 (19.6) | 1,767,805<br>(15.7) |  | 153,096 (15.6) | 153,096 (15.6) | 306,192 (15.6) |  |
| 80+ | 434,655 (6.6) | 416,565 (8.8) | 851,220 (7.5) |  | 52,300 (5.3) | 52,300 (5.3) | 104,600 (5.3) |  |
| **Sex** |  |  |  | 0.0215 |  |  |  | 0.0 |
| Female | 3,578,944<br>(54.4) | 2,514,975<br>(53.4) | 6,093,919<br>(54.0) |  | 522,702 (53.3) | 522,691 (53.3) | 1,045,393<br>(53.3) |  |
| Male | 2,996,262<br>(45.6) | 2,198,152<br>(46.6) | 5,194,414<br>(46.0) |  | 458,709 (46.7) | 458,720 (46.7) | 917,429 (46.7) |  |
| **Race** |  |  |  | 0.4275 |  |  |  | 0.0 |
| Black | 645,949 (9.8) | 353,416 (7.5) | 999,365 (8.9) |  | 62,607 (6.4) | 62,601 (6.4) | 125,208 (6.4) |  |
| Others | 1,459,955<br>(22.2) | 702,248 (14.9) | 2,162,203<br>(19.2) |  | 137,196 (14.0) | 137,190 (14.0) | 274,386 (14.0) |  |
| Unknown | 247,985 (3.8) | 720,500 (15.3) | 968,485 (8.6) |  | 30,111 (3.1) | 30,111 (3.1) | 60,222 (3.1) |  |
| White | 4,221,317<br>(64.2) | 2,936,963<br>(62.3) | 7,158,280<br>(63.4) |  | 751,497 (76.6) | 751,509 (76.6) | 1,503,006<br>(76.6) |  |
| **Ethnicity** |  |  |  | 0.4989 |  |  |  | 0.0 |
| Hispanic | 2,056,323<br>(31.3) | 685,958 (14.6) | 2,742,281<br>(24.3) |  | 194,063 (19.8) | 194,067 (19.8) | 388,130 (19.8) |  |
| Not Hispanic | 3,730,586<br>(56.7) | 2,769,752<br>(58.8) | 6,500,338<br>(57.6) |  | 688,449 (70.2) | 688,439 (70.2) | 1,376,888<br>(70.2) |  |

| Variables | Full cohort |  |  |  | Matched cohort |  |  |  |
| --- | --- | --- | --- | --- | --- | --- | --- | --- |
|  | BNT162b2<br>N (%) | mRNA-1273 N<br>(%) | Total N (%) | Standardize<br>d Difference | BNT162b2<br>N (%) | mRNA-1273<br>N (%) | Total N (%) | Standardize<br>d Difference |
| Unknown | 788,297 (12.0) | 1,257,417<br>(26.7) | 2,045,714<br>(18.1) |  | 98,899 (10.1) | 98,905 (10.1) | 197,804 (10.1) |  |
| **Vaccination**<br>**site** |  |  |  | 0.4654 |  |  |  | 0.0 |
| CHD OR DOH | 1,049,292<br>(16.0) | 826,335 (17.5) | 1,875,627<br>(16.6) |  | 115,927 (11.8) | 115,927 (11.8) | 231,854 (11.8) |  |
| Doctor's clinic | 1,235,108<br>(18.8) | 469,804 (10.0) | 1,704,912<br>(15.1) |  | 158,314 (16.1) | 158,314 (16.1) | 316,628 (16.1) |  |
| Fire Rescue<br>Or EMS | 347,376 (5.3) | 104,593 (2.2) | 451,969 (4.0) |  | 16,350 (1.7) | 16,350 (1.7) | 32,700 (1.7) |  |
| FQHC OR<br>RHC OR CHC | 237,351 (3.6) | 186,131 (4.0) | 423,482 (3.8) |  | 13,488 (1.4) | 13,488 (1.4) | 26,976 (1.4) |  |
| Hospital Clinic | 446,202 (6.8) | 366,792 (7.8) | 812,994 (7.2) |  | 52,674 (5.4) | 52,674 (5.4) | 105,348 (5.4) |  |
| Hospital ER | 42,689 (0.7) | 83,983 (1.8) | 126,672 (1.1) |  | 2,945 (0.3) | 2,945 (0.3) | 5,890 (0.3) |  |
| Other<br>Pharmacies | 188,232 (2.9) | 556,815 (11.8) | 745,047 (6.6) |  | 40,756 (4.2) | 40,756 (4.2) | 81,512 (4.2) |  |
| Others Or<br>Misc. | 388,330 (5.9) | 335,533 (7.1) | 723,863 (6.4) |  | 91,400 (9.3) | 91,400 (9.3) | 182,800 (9.3) |  |
| Publix, CVS,<br>or Walgreens | 2,640,626<br>(40.2) | 1,783,141<br>(37.8) | 4,423,767<br>(39.2) |  | 489,557 (49.9) | 489,557 (49.9) | 979,114 (49.9) |  |
| **Month of**<br>**receiving**<br>**COVID-19**<br>**Vaccine Dose**<br>**1** |  |  |  | 0.2792 |  |  |  | 0.0 |
| **Dec** | 127,641 (1.9) | 118,059 (2.5) | 245,700 (2.2) |  | 9,920 (1.0) | 9,920 (1.0) | 19,840 (1.0) |  |
| Jan | 736,922 (11.2) | 797,497 (16.9) | 1,534,419<br>(13.6) |  | 143,684 (14.6) | 143,684 (14.6) | 287,368 (14.6) |  |
| Feb | 628,580 (9.6) | 648,219 (13.8) | 1,276,799<br>(11.3) |  | 87,037 (8.9) | 87,037 (8.9) | 174,074 (8.9) |  |
| Mar | 1,609,563<br>(24.5) | 1,195,638<br>(25.4) | 2,805,201<br>(24.9) |  | 295,057 (30.1) | 295,057 (30.1) | 590,114 (30.1) |  |
| Apr | 1,463,665<br>(22.3) | 900,505 (19.1) | 2,364,170<br>(20.9) |  | 211,246 (21.5) | 211,246 (21.5) | 422,492 (21.5) |  |
| May | 585,561 (8.9) | 349,067 (7.4) | 934,628 (8.3) |  | 66,581 (6.8) | 66,581 (6.8) | 133,162 (6.8) |  |
| Jun | 359,369 (5.5) | 138,060 (2.9) | 497,429 (4.4) |  | 24,830 (2.5) | 24,830 (2.5) | 49,660 (2.5) |  |
| Jul | 454,750 (6.9) | 215,792 (4.6) | 670,542 (5.9) |  | 48,517 (4.9) | 48,517 (4.9) | 97,034 (4.9) |  |
| Aug | 609,155 (9.3) | 350,290 (7.4) | 959,445 (8.5) |  | 94,539 (9.6) | 94,539 (9.6) | 189,078 (9.6) |  |
| **SVI Score** |  |  |  |  |  |  |  |  |
|  | BNT162b2<br>N (%) | mRNA-1273 N<br>(%) | Total N (%) | Standardize<br>d Difference | BNT162b2<br>N (%) | mRNA-1273<br>N (%) | Total N (%) | Standardize<br>d Difference |
| Overall<br>(Median<br>(IQR)) | 46.9 (25.0,<br>71.8) | 43 (20.6, 66.4) | 46.3 (23.3,<br>69.6) | 0.1458 | 42.4 (20.3,<br>62.6) | 56 (39, 67) | 42.4 (20.3,<br>62.6) | 0.0 |
| Socioeconomi<br>c status<br>(Median<br>(IQR)) | 45.3 (22.8,<br>68.9) | 42.9 (20.6,<br>65.8) | 44.4 (21.8,<br>67.6) | 0.0702 | 41.2 (19.1,<br>60.5) | 42.4 (20.3,<br>62.6) | 41.2 (19.1,<br>60.5) | 0.0 |
| Housing and<br>transportation<br>(Median<br>(IQR)) | 47.4 (24.3,<br>72.5) | 44.1 (21, 69.2) | 47.1 (22.9,<br>71.8) | 0.1009 | 44.3 (20.4,<br>65.9) | 44.3 (20.4,<br>65.9) | 44.3 (20.4,<br>65.9) | 0.0 |

Notes: Q1= 25<sup>th</sup> percentile, Q3= 75<sup>th</sup> percentile, CHD = County Health Department, DOH = Department of Health, EMS= Emergency Medical Services, Hospital ER = Hospital Emergency Room, FQHC = Federally Qualified Health Center, CHC = Community health Center, RHC = Rural Health Clinic

Overall, BNT162b2 was the dominant vaccine for all age groups in every month with the exception of January for all age groups less than 80 years, December 2020 to April 2021 for the age group 70-79 years, and February to April, 2021 for age group 80 years and older (Supplement Figure 1). Vaccine allocation to different vaccination sites shifted over time. In January 2021, 15% of COVID-19 vaccines were provided by hospitals and 10% were provided in outpatient pharmacies. By August 2021, this distribution had shifted to less than 1% being in hospitals and about 80% in pharmacies.

### Mortality Outcomes

Table 2 shows the number of deaths in the matched cohort. BNT162b2 recipients had significantly higher odds of all-cause mortality compared to mRNA-1273 recipients (odds ratio, OR [95% CI]: 1.407 [1.358, 1.457]), with an excess per 100,000 persons of 221.2 deaths (95% CI: [198.3, 244.1]) (Table 3). The corresponding risk ratio was 1.393 (95% CI: [1.346, 1.442]). Similarly, BNT162b2 recipients had significantly higher odds of cardiovascular mortality compared to mRNA-1273 recipients (OR [95% CI]: 1.562 [1.461, 1.670]), with an excess of 80.4 deaths (95% CI: [68.4, 92.4]) per 100,000 persons. This corresponded with a risk ratio of 1.554 (95% CI: [1.454, 1.661]).

**Table 2.**
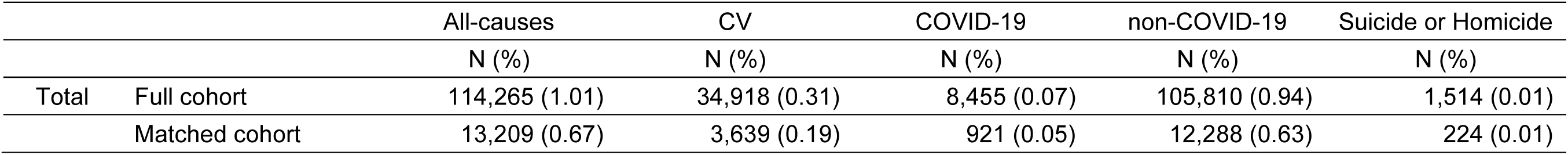
All-cause, Cardiovascular (CV), COVID-19, non-COVID-19, and Suicide or Homicide mortality in matched and full cohorts.

|  |  | All-causes | CV | COVID-19 | non-COVID-19 | Suicide or Homicide |
| --- | --- | --- | --- | --- | --- | --- |
|  |  | N (%) | N (%) | N (%) | N (%) | N (%) |
| Total | Full cohort | 114,265 (1.01) | 34,918 (0.31) | 8,455 (0.07) | 105,810 (0.94) | 1,514 (0.01) |
|  | Matched cohort | 13,209 (0.67) | 3,639 (0.19) | 921 (0.05) | 12,288 (0.63) | 224 (0.01) |

**Table 3.**
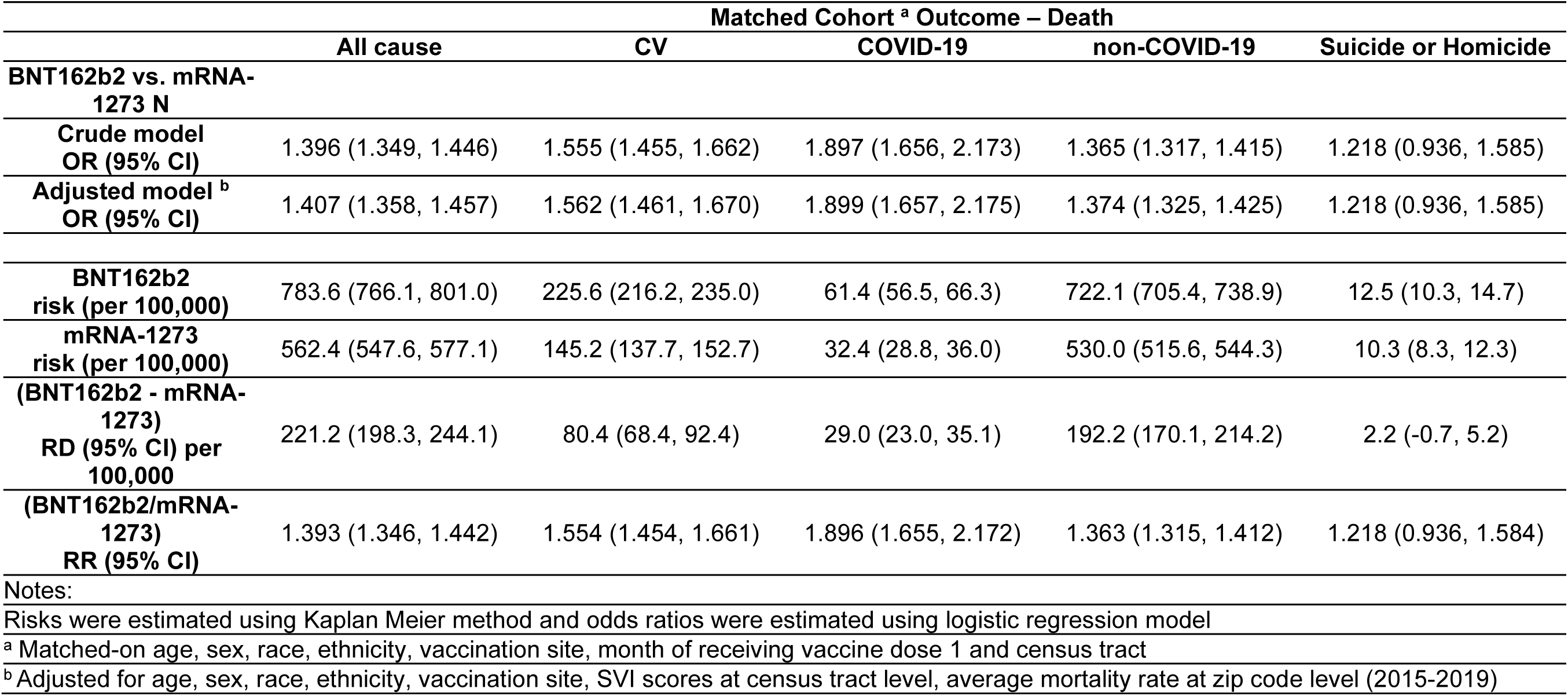
Risk, risk difference (RD), risk ratio (RR), and odds ratio (OR) of all-cause, cardiovascular, COVID-19, non-COVID-19, and suicide or homicide deaths in matched cohort.

The risk of COVID-19 mortality was higher in BNT162b2 recipients compared to mRNA-1273 recipients (OR [95% CI]: 1.899 [1.657, 2.175]), with an excess per 100,000 persons of 29.0 deaths (95% CI: [23.0, 35.1]). The corresponding risk ratio was 1.896 (95% CI: [1.655, 2.172]). Non-COVID-19 mortality risk was also higher in BNT162b2 recipients compared to mRNA-1273 recipients (OR [95% CI]: 1.374 [1.325, 1.425]), with an excess per 100,000 persons of 192.2 deaths (95% CI: [170.1, 214.2]). This corresponded with a risk ratio of 1.363 (95% CI: [1.315, 1.412]).

When stratifying by age group, the increase in mortality risk was highest in adults 60 years and older (Supplement Table 3).

### Cumulative Mortality Incidence Curves

Cumulative incidence curves for all-cause, cardiovascular, COVID-19, and non-COVID-19 mortality in the matched cohort are shown in Figure 2. For all-cause, cardiovascular, and non-COVID-19 deaths, the corresponding curves show clear separation starting 30-40 days post vaccination with dose 1. For COVID-19 mortality, there was a pronounced separation in incidence curves between recipients of BNT162b2 versus mRNA-1273 initially after vaccination and a second divergence that began approximately 200 days after vaccination. The latter period may correspond to the Delta wave which began in Florida around July 2021, when the Delta variant of severe acute respiratory syndrome coronavirus 2 (SARS-CoV-2) became the dominant variant in Florida.

**Figure 2.**
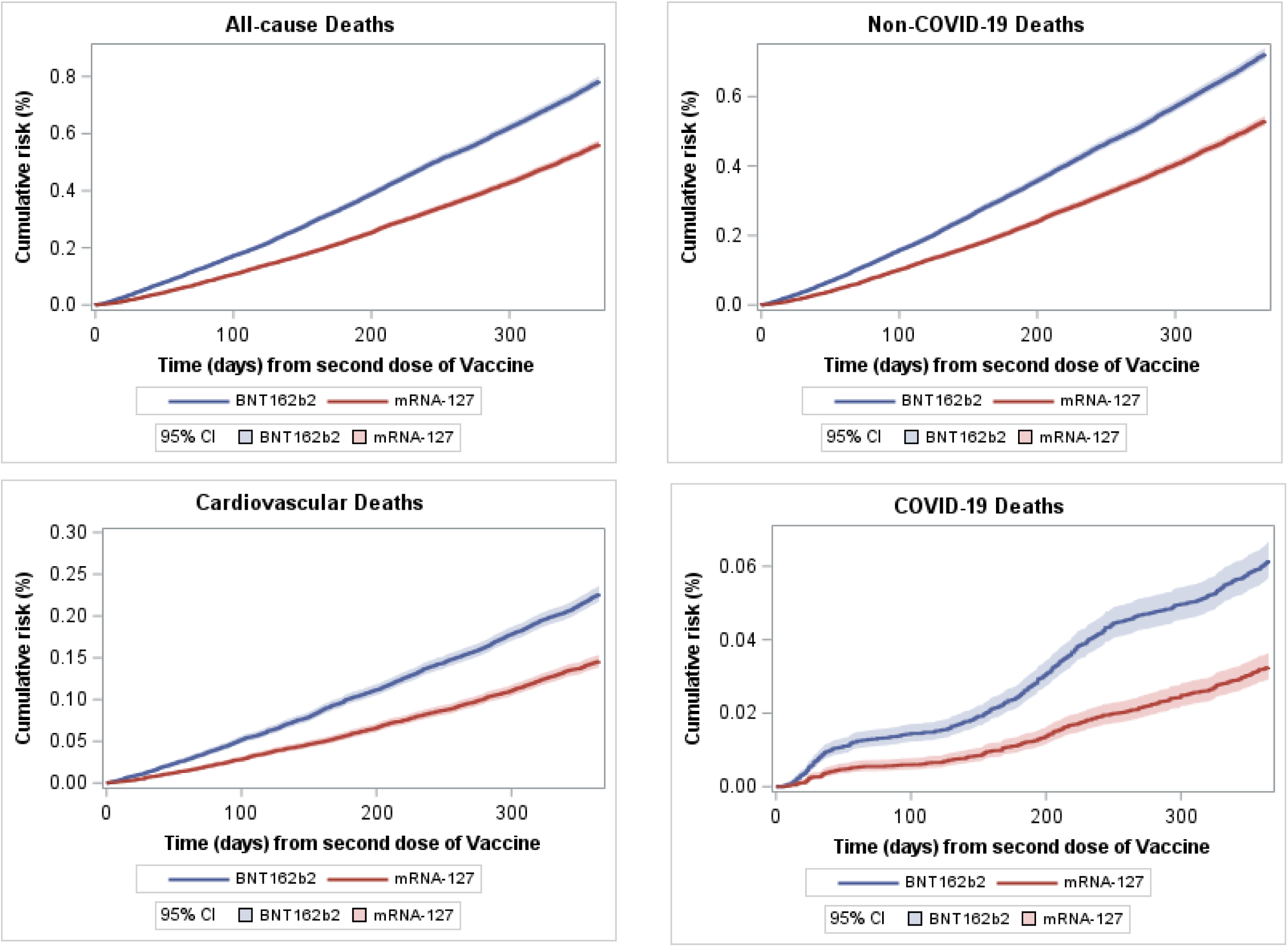
Cumulative incidence of all-cause, cardiovascular, COVID-19, and non-COVID-19 mortality in matched cohort. The cumulative incidence up to 365 days after the First dose of vaccine for all cause, cardiovascular, COVID-19, and non-COVID-19 are shown in the top left, top right, bottom left, and bottom right panels, respectively. Cumulative incidence was calculated using Kaplan Meier method and the follow-up ended either at the date of death or end of study observation period (365 days from first dose of vaccine). For cause-specific cumulative incidence, deaths from all other causes were censored at the time of death.

### Negative Control Outcomes

Similar cumulative incidence curves were constructed for the negative control outcome of homicide and suicide within the matched cohort (Supplement Figure 2). The difference between the curves is not significant. The odds ratio of homicide and suicide mortality in the matched cohort was 1.218 (95% CI: [0.936, 1.585]), and was not significant in any age groups. The prevalence of SARS-CoV-2 infection prior to December 20, 2020, was 3.55% for BNT162b2 recipients and 3.49% for mRNA-1273 recipients. The standardized mean difference was 0.008, suggesting very high level of balance. Logistic regression models, with SARS-CoV-2 infection as an outcome, yielded an odds ratio of 1.024 (95% CI: [1.001, 1.047]) for BNT162b2 as compared to mRNA-1273 recipients in the matched cohort. Stratification by age showed no meaningful difference for the 18-39 and 40–59-year-old groups and was significant only among individuals 60 years or older (OR [95% CI]: 1.096 [1.046, 1.149]) (Supplement Table 6). A post hoc analysis of mortality outcomes that adjusted for history of SARS-CoV-2 infection prior to vaccination (any infection before the first vaccine dose) showed similar results as the main analysis (Supplement Table 7). Additional post hoc analysis excluding patients with prior SARS-CoV-2 infection showed results similar to the primary analysis (Supplement Table 5).

### Sensitivity Analysis

The sensitivity analysis for unobserved confounders indicates that the findings from the analyses of the matched cohort are robust to unobserved confounders. For example, an unobserved binary covariate with odds ratio 1.6, i.e., 1.6-fold more likely odds of occurring among BNT162b2 recipients compared to mRNA-1273 recipients, will have to be associated with mortality outcome with an odds ratio of at least 5.2, for the results not to be statistically significant (at a level of 0.05). Complete results are provided in the Supplement (supplement Table 8). Repeating the analysis with COVID-19+ and non-COVID-19+ deaths yielded results similar to the primary analysis with non-COVID-19+ deaths having an odd ratio of 1.341 (95% CI: [1.293, 1.392) for BNT162b2 as compared to mRNA-1273 recipients in the matched cohort (Table 4 and Supplement Figure 4).

**Table 4.** COVID-19 and non-COVID-19 deaths, with (COVID-19+) or without (non-COVID-19+) prior SARS-CoV-2 infection, in matched cohort.

| Models | Matched Cohort<br>BNT162b2 vs. mRNA-1273 (First Dose) |  |
| --- | --- | --- |
|  | COVID-19 Death or non-COVID-19 Death<br>after prior SARS-CoV-2 infection<br>OR (95% CI) | Non-COVID-19 Death with<br>no prior SARS-CoV-2 infection<br>OR (95% CI) |
| **Unadjusted model** | 1.863 (1.683, 2.062) | 1.341 (1.293, 1.392) |
| **Adjusted model <sup>b</sup>** | 1.866 (1.685, 2.066) | 1.349 (1.300, 1.400) |
| **Adjusted Model by Age <sup>b</sup>** |  |  |
| 18-39 | 1.916 (0.953, 3.851) | 1.309 (1.021, 1.678) |
| 40-59 | 1.342 (1.014, 1.778) | 1.131 (1.013, 1.262) |
| 60+ | 1.967 (1.760, 2.198) | 1.383 (1.329, 1.440) |
| **BNT162b2**<br>**risk (per 100,000)** | 108.5 (102.0, 115.0) | 675.0 (658.8, 691.2) |
| **mRNA-1273**<br>**risk (per 100,000)** | 58.3 (53.5, 63.1) | 504.1 (490.1, 518.1) |
| **(BNT162b2 - mRNA-1273)**<br>**RD (95% CI) per 100,000** | 50.2 (42.2, 58.3) | 171.0 (149.6, 192.4) |
| **(BNT162b2/mRNA-1273)**<br>**RR (95% CI)** | 1.862 (1.682, 2.061) | 1.339 (1.291, 1.389) |
| Notes: |  |  |
| a Matched-on age, sex, race, ethnicity, vaccination site, month of receiving vaccine dose 1 and census tract |  |  |
| b Adjusted for age, sex, race, ethnicity, vaccination site, SVI scores at census tract level, average mortality rate at zip code level (2015-2019) |  |  |

## DISCUSSION

This study compared 12-month mortality outcomes of noninstitutionalized adult Florida residents who received a first dose of BNT162b2 or mRNA-1273 for initial vaccination against COVID-19. The findings demonstrate that vaccination with BNT162b2 was associated with higher 12-month all-cause, cardiovascular, COVID-19, and non-COVID-19 mortality compared to mRNA-1273. The increase in mortality risk was present in all age groups but was most pronounced among vaccine recipients over 60 years of age. Analysis of negative control outcomes did not demonstrate existence of meaningful residual bias, particularly compared to the observed effect size of increased mortality risk, and the robustness level indicated by the sensitivity analysis. More generally, the study results underscore the importance of considering non-specific vaccine effects, and specifically all-cause mortality, when evaluating vaccines.

Several prior studies compared different outcome endpoints between recipients of the BNT162b2 and mRNA-1273 vaccines. Three studies based on data from the U.S. Veterans Affairs system are the most closely related to the current study because they also used matching between BNT162b2 and mRNA-1273 recipients. The first of these studies compared COVID-19-related outcomes including infection, hospitalization, admission to the intensive care unit and death.^10^ It conducted up to 6-month follow-up on a matched cohort that included 219,842 individuals in each cohort after administration of the first vaccine dose. The matching was conducted based on calendar date (five-day bins), age (five-year bins), race, urbanicity of residence and coarse geographical region. The results showed that BNT162b2 recipients were at higher risk of documented SARS-CoV-2 infection, as well as COVID-19 hospitalization and death, although the study was not statistically powered with respect to the latter. One of the negative control outcomes used in this study were non-COVID-19 deaths, for which the study did not find statistically significant differences. A second study, also focused on COVID-19 outcomes, had similar results using Veterans Affairs and Medicare data, and applied a more nuanced matching scheme to match 902,235 BNT162b2 recipients with 656,736 mRNA-1273 recipients.^14^ An important finding of this analysis is that the difference between the BNT162b2 and mRNA-1273 cohorts increased with time, which is consistent with the finding of the current study with respect to COVID-19 deaths.

The third of these studies used a similar one-to-one matching method to conduct 266-day follow-up of multiple adverse event outcomes experienced after vaccination.^9^ It showed that BNT162b2 recipients were at a higher risk for several cardiovascular-related outcomes, such as myocardial infraction, ischemic stroke, thromboembolic disease, and myocarditis. Consistent with the studies from the Veterans Affairs, the current study detected increased cardiovascular mortality among recipients of the Pfizer-BioNTech’s BNT162b2 compared to Moderna’s mRNA-1273.

There are notable differences between the current study and the three Veterans Affairs studies that also compared between BNT162b2 and mRNA-1273 recipients. First, baseline characteristics of the adult population in Florida are likely to differ from the U.S. Veterans Affairs population. Second, the current study is more adequately statistically powered because it includes a substantially larger population than all prior studies, and a longer follow-up time of 12 months. Third, the matching used in the current study has significantly more refined adjustment for geographic characteristics

due to use of census tract and type of vaccination site, both of which likely capture important confounding factors. Notably, even the coarser matching used in the studies from the Veterans Affairs system formed cohorts that were balanced in terms of covariates associated with health status and health-seeking behavior.^10,14^ Indeed, in our study, the prevalence of SARS-CoV-2 infection prior to the start of the vaccination campaign was very balanced between BNT162b2 and mRNA-1273 recipients. Fourth, unlike other outcomes, detection of mortality outcomes is not likely dependent on health-seeking behaviors. Fifth, the two Veterans Affairs studies used a different definition of COVID-19 deaths, specifically counting any death within 30 days of confirmed infection, compared to using death certificates in the current study.^10,14^ These differences could potentially explain why the current study detected significant higher all-cause and non-COVID-19 mortality rates among recipients of BNT162b2 compared to mRNA-1273, whereas the studies from the Veterans Affairs system did not.^1,15-23^

The findings of this study are consistent with the reanalysis of the pivotal phase III randomized clinical trials conducted by Pfizer and Moderna.^24^ In their reanalysis, Fraiman et al found that BNT162b2 recipients experienced a 36% increase in the risk of serious adverse events compared to the placebo group, whereas mRNA-1273 recipients experienced a 6% increased risk, and only the former was statistically significant.

The biological mechanisms that may explain a potential difference in overall health outcomes after vaccination with BNT162b2 versus mRNA-1273 are still unclear. The two vaccines include different quantities of mRNA but there are also differences in the mRNA sequences, quality control of the manufacturing process, delivery systems including type and quantity of lipid nanoparticles, temperature storage requirements, and schedule for administration, in terms of time between the first and second dose.^25,26^ There may also be differences related to biodistribution and other pharmacokinetic and pharmacodynamic aspects of the vaccines.

The selection of appropriate negative controls in this analysis was complex due to uncertainty about the type and nature of non-specific effects associated with BNT162b2 and mRNA-1273 COVID-19 vaccines. A necessary condition of a negative outcome control is that it shares the same potential sources of bias as the primary outcome but is not causally related to the exposure of interest.^27^ Because the mechanisms of possible non-specific vaccine effects are unknown, negative controls such as bone fractures from falls or motor vehicle accidents may be inappropriate if, for example, the vaccines have differential cardiovascular effects that may influence the risk of syncope.

This study adds to a growing scientific understanding of the potential clinical significance of non-specific vaccines effects. While adverse reactions are the most widely recognized non-specific effects of vaccines, prior research indicates that non-specific effects also encompass longer term health effects that could be either beneficial or harmful. For example, multiple studies including randomized controlled trials have demonstrated that certain strains of the Bacille Calmette-Guérin (BCG) vaccine for tuberculosis are associated with a reduction in all-cause mortality among infants, with these benefits attributed to non-tuberculosis diseases.^1,15-17^ Similarly, randomized controlled trials (RCTs) reported that measles vaccination was associated with reductions in all-cause mortality, independent of its effects on measles infection or measles-induced immune amnesia.^18^ The live oral polio vaccine (OPV) has also been shown to likely convey survival benefits that cannot be solely attributed to prevention of polio.^19,20^ Some researchers hypothesize that these non-specific vaccine effects may be due to changes in immune system function.

However, non-specific vaccine effects may also be harmful to health, particularly in the context of non-live vaccines.^1^ For example, the diphtheria, tetanus, and pertussis (DTP) vaccine has been associated with an increased risk of all-cause mortality after childhood administration in multiple cohorts, with this adverse finding most pronounced in females. The sequence in which vaccines are administered may also affect the expression of adverse non-specific effects, as demonstrated in randomized clinical trials of the high-titer measles vaccine.^21–23^ Overall, this scientific area underscores the importance of considering vaccine effects on mortality that extend beyond the diseases targeted, both for unexpected benefits and unforeseen risks.

### Limitations

Like any observational study, there is always a concern regarding the existence of unobserved confounding that could bias the results. Although matching was performed on multiple characteristics, including age group, sex, race, ethnicity, calendar month, nine separate vaccination sites, and census tract, the matching did not include potentially important characteristics, such as comorbid conditions. Differences in these conditions could bias the analysis, although prior population studies using fewer criteria for matching have demonstrated that the omission of comorbid conditions does not appear to increase the risk of bias.^28^ Moreover, the robustness analysis suggests that only very substantial unobserved covariates could the alter the direction of the results.

Since this study does not compare vaccinated to unvaccinated adults, it is a priori possible that the differences in outcomes between BNT162b2 and mRNA-1273 recipients are explained by a higher protective effect of mRNA-1273 against infection-related outcomes. However, the sensitivity analysis that considered only deaths that occurred without prior documented infection, suggests that if such a mechanism exists, it would have to involve a highly nontrivial impact of asymptomatic infections.

Matching also necessarily reduced the size of the studied population, particularly because of the large number of characteristics used to match. It is therefore possible that, similar to clinical trials, the findings may not generalize to segments of the excluded population. Some of the health outcomes, such as COVID-19 deaths and cardiovascular deaths, could be subject to misclassification due to errors on death certificates. While misclassification would introduce error, it would not be expected to differ between recipients of BNT162b2 or mRNA-1273.

Despite these limitations, the large number of adults included in this analysis, rigorousness of the matching process, and consistency of our findings with prior studies support the validity of these findings. Florida adults who received the BNT162b2 COVID-19 vaccine in the initial series appear to have significantly higher risk of 12-month all-cause, cardiovascular, COVID-19, and non-COVID-19 mortality compared to mRNA-1273 recipients. These findings may have implications for public health recommendations and evaluation frameworks for vaccines in the context of nonspecific effects related to mortality.

## Supporting information

SAS Codes and Output File

## Data Availability

The study is based on data from the Florida Department of Health and is prohibited by statute from sharing in its raw form.

## Contributor statement

RL and JL conceived and designed the study, identified the study population, and defined the outcomes, MJ and FM obtained the data. RL, JL, and FM designed the study methods. FM conducted the statistical analyses. RL and JL drafted the manuscript, which was subsequently reviewed, revised and approved by all authors. RL and JL are the guarantors. The corresponding author attests that all listed authors meet authorship criteria and that no others meeting the criteria have been omitted.

## Ethical approval

This study was reviewed by the Florida Department of Health institutional review board and deemed exempt as public health practice.

## Dissemination to participants and related patient and public communities

The study findings will be disseminated through press releases by the institutions represented by the authors and through patient organizations.

## Transparency declaration

RL and JL affirm that the manuscript is an honest, accurate, and transparent account of the study being reported; that no important aspects of the study have been omitted; and that any discrepancies from the study as originally planned have been explained.

## Acknowledgement

We thank Tracy Beth Høeg, MD, PhD for assistance with study method development and writing during the early stage of this research.

## Supplement Tables and Figures

**Supplement Table 1.**
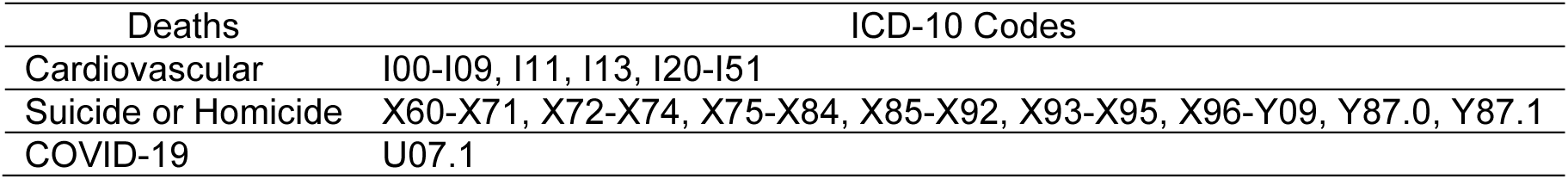
ICD-10 codes used to define COVID-19, Cardiovascular, and Suicide or Homicide Deaths.

**Supplement Table 2.** All-cause, Cardiovascular (CV), COVID-19, non-COVID-19, and Suicide or Homicide mortality by Age in matched and full cohorts.

|  |  | All-causes | CV | COVID-19 | non-COVID-19 | Suicide or Homicide |
| --- | --- | --- | --- | --- | --- | --- |
|  |  | N (%) | N (%) | N (%) | N (%) | N (%) |
| Age |  |  |  |  |  |  |
| 18-39 | Full cohort | 2,141 (0.07) | 202 (0.01) | 98 (<0.01) | 2,043 (0.07) | 307 (0.01) |
|  | Matched cohort | 289 (0.06) | 25 (0.01) | 13 (<0.00) | 276 (0.06) | 53 (0.01) |
| 40-49 | Full cohort | 2,804 (0.17) | 514 (0.03) | 176 (0.01) | 2,628 (0.16) | 197 (0.01) |
|  | Matched cohort | 369 (0.14) | 78 (0.03) | 30 (0.01) | 339 (0.13) | 23 (0.01) |
| 50-59 | Full cohort | 7,829 (0.40) | 1,854 (0.09) | 533 (0.03) | 7,296 (0.37) | 278 (0.01) |
|  | Matched cohort | 1,114 (0.29) | 240 (0.06) | 69 (0.02) | 1,045 (0.28) | 45 (0.01) |
| 60-69 | Full cohort | 18,477 (0.84) | 4,722 (0.21) | 1,280 (0.06) | 17,197 (0.78) | 307 (0.01) |
|  | Matched cohort | 2,765 (0.65) | 660 (0.16) | 192 (0.05) | 2,573 (0.60) | 47 (0.01) |
| 70-79 | Full cohort | 30,254 (1.71) | 8,318 (0.47) | 2,314 (0.13) | 27,940 (1.58) | 252 (0.01) |
|  | Matched cohort | 4,234 (1.38) | 1,103 (0.36) | 304 (0.10) | 3,930 (1.28) | 38 (0.01) |
| 80+ | Full cohort | 52,760 (6.20) | 19,308 (2.27) | 4,054 (0.48) | 48,706 (5.72) | 173 (0.02) |
|  | Matched cohort | 4,438 (4.24) | 1,533 (1.47) | 313 (0.30) | 4,125 (3.94) | 18 (0.02) |

**Supplement Table 3.** Odds ratios of mortality from All causes, Cardiovascular (CV), COVID-19, non-COVID-19, and Suicide or Homicide in matched cohort by Age group of participants receiving BNT162b2 as compared to mRNA-1273mRNA COVID vaccines.

| Models | Matched Cohort <sup>a</sup> BNT162b2 vs. mRNA-1273 N |  |  |  |  |
| --- | --- | --- | --- | --- | --- |
|  | All-cause<br>OR (95% CI) | CV<br>OR (95% CI) | COVID-19<br>OR (95% CI) | Non-COVID-19<br>OR (95% CI) | Suicide or Homicide<br>OR (95% CI) |
| **Results by Age <sup>b</sup>** |  |  |  |  |  |
| 18-39 | 1.369 (1.084, 1.729) | 1.500 (0.674, 3.340) | 1.599 (0.523, 4.888) | 1.359 (1.070, 1.726) | 1.304 (0.757, 2.245) |
| 40-59 | 1.157 (1.045, 1.282) | 1.150 (0.923, 1.434) | 1.201 (0.808, 1.784) | 1.154 (1.038, 1.283) | 1.061 (0.660, 1.707) |
| 60+ | 1.447 (1.393, 1.503) | 1.620 (1.509, 1.739) | 2.027 (1.750, 2.347) | 1.408 (1.354, 1.465) | 1.288 (0.873, 1.902) |
| Notes: |  |  |  |  |  |
| <sup>a</sup> Matched-on age, sex, race, ethnicity, vaccination site, month of receiving vaccine dose 1 and census tract |  |  |  |  |  |
| <sup>b</sup> Adjusted for age, sex, race, ethnicity, vaccination site, SVI scores at census tract level, average mortality rate at zip code level (2015-2019) |  |  |  |  |  |

**Supplement Table 4.** Odds ratios of mortality from All causes, Cardiovascular (CV), COVID-19, non-COVID-19, and Suicide or Homicide in full cohort of participants receiving MRNA-1273 as compared to BNT162b2 mRNA COVID-19 vaccines.

| Models | Full Cohort<br>BNT162b2 vs. mRNA-1273 |  |  |  |  |
| --- | --- | --- | --- | --- | --- |
|  | All-cause<br>OR (95% CI) | CV<br>OR (95% CI) | COVID-19<br>OR (95% CI) | Non-COVID-19<br>OR (95% CI) | Suicide or Homicide<br>OR (95% CI) |
| **Unadjusted model** | 0.947 (0.927, 0.968) | 1.323 (1.265, 1.384) | 0.868 (0.858, 0.879) | 0.795 (0.718, 0.879) | 0.947 (0.927, 0.968) |
| **Adjusted model <sup>b</sup>** | 1.506 (1.472, 1.541) | 2.096 (1.998, 2.199) | 1.367 (1.349, 1.385) | 1.060 (0.949, 1.184) | 1.506 (1.472, 1.541) |
| **Adjusted Model by Age <sup>b</sup>** |  |  |  |  |  |
| 18-39 | 0.904 (0.661, 1.236) | 0.847 (0.544, 1.319) | 0.899 (0.815, 0.992) | 0.915 (0.710, 1.180) | 0.904 (0.661, 1.236) |
| 40-59 | 0.986 (0.901, 1.079) | 1.499 (1.261, 1.781) | 0.892 (0.854, 0.932) | 0.997 (0.816, 1.219) | 0.986 (0.901, 1.079) |
| 60+ | 1.557 (1.520, 1.594) | 2.175 (2.068, 2.288) | 1.419 (1.399, 1.439) | 1.143 (0.978, 1.336) | 1.557 (1.520, 1.594) |
| Notes: |  |  |  |  |  |
| <sup>b</sup> Adjusted for age, sex, race, ethnicity, vaccination site, SVI scores at census tract level, average mortality rate at zip code level (2015-2019) |  |  |  |  |  |

**Supplement Table 5.** Odds ratios of mortality from All causes, Cardiovascular (CV), COVID-19, non-COVID-19, and Suicide or Homicide in matched cohort after excluding participants with history of COVID-19 infection before receiving BNT162b2 or mRNA-1273 COVID vaccine.

| Matched cohort (excluding COVID-19 infections)<sup>a, c</sup><br>BNT162b2 vs. mRNA-1273 |  |  |  |  |  |
| --- | --- | --- | --- | --- | --- |
|  | All-cause | CVD | COVID | Non-COVID | Suicide or Homicide |
|  | OR (95% CI) | OR (95% CI) | OR (95% CI) | OR (95% CI) | OR (95% CI) |
| **Overall results** |  |  |  |  |  |
| Crude model | 1.363 (1.315, 1.412) | 1.520 (1.420, 1.627) | 1.844 (1.606, 2.116) | 1.332 (1.284, 1.382) | 1.206 (0.922, 1.579) |
| Adjusted model<sup>b</sup> | 1.383 (1.334, 1.434) | 1.542 (1.440, 1.651) | 1.860 (1.620, 2.135) | 1.350 (1.301, 1.402) | 1.207 (0.922, 1.579) |
| **Results by Age<sup>b</sup>** |  |  |  |  |  |
| 18-39 | 1.396 (1.096, 1.780) | 1.550 (0.671, 3.582) | 1.401 (0.445, 4.416) | 1.396 (1.089, 1.789) | 1.329 (0.754, 2.340) |
| 40-59 | 1.165 (1.048, 1.295) | 1.157 (0.921, 1.455) | 1.151 (0.772, 1.716) | 1.166 (1.045, 1.302) | 0.996 (0.615, 1.614) |
| 60+ | 1.414 (1.360, 1.470) | 1.590 (1.479, 1.710) | 1.992 (1.717, 2.313) | 1.376 (1.321, 1.432) | 1.309 (0.880, 1.948) |

Notes:
<sup>a</sup> Matched-on age, sex, race, ethnicity, vaccination site, month of receiving vaccine dose 1 and census tract
<sup>b</sup> Adjusted for age, sex, race, ethnicity, vaccination site, SVI scores at census tract level, average mortality rate at zip code level (2015-2019)
<sup>c</sup> Sensitivity analysis was performed on matched cohort after excluding participants with a history of COVID-19 infection prior to the first dose of vaccine.

**Supplement Table 6.** Odds ratios of history of SARS-CoV-2 infections (before 20 December 2020) in Matched cohort.

| **Matched cohort<sup>a</sup>** |  |
| --- | --- |
| **BNT162b2 vs. mRNA-1273** |  |
|  | **SARS-CoV-2 infections** |
|  | **OR (95% CI)** |
| **Overall results** |  |
| Crude model | 1.016 (1.001, 1.032) |
| Adjusted model <sup>b</sup> | 1.024 (1.001, 1.047) |
| **Results by Age <sup>b</sup>** |  |
| 18-39 | 1.020 (0.984, 1.057) |
| 40-59 | 0.986 (0.950, 1.023) |
| 60+ | 1.096 (1.046, 1.149) |
| <sup>a</sup> Matched-on age, sex, race, ethnicity, vaccination site, month of receiving vaccine dose 1 and census tract |  |
| <sup>b</sup> Adjusted for age, sex, race, ethnicity, vaccination site, SVI scores at census tract level, average mortality rate at zip code level (2015-2019) |  |

**Supplement Table 7.** Odds ratios of mortality from All causes, Cardiovascular (CV), COVID-19, non-COVID-19, and Suicide or Homicide in Matched cohort adjusted for history of SARS-CoV-2 infection and other confounders.

| Matched cohort<sup>a</sup><br>BNT162b2 vs. mRNA-1273 |  |  |  |  |  |
| --- | --- | --- | --- | --- | --- |
|  | All-cause<br>OR (95% CI) | CVD<br>OR (95% CI) | COVID<br>OR (95% CI) | Non-COVID<br>OR (95% CI) | Suicide or Homicide<br>OR (95% CI) |
| **Overall results** |  |  |  |  |  |
| Crude model | 1.396 (1.349, 1.446) | 1.555 (1.455, 1.662) | 1.897 (1.656, 2.173) | 1.365 (1.317, 1.415) | 1.218 (0.936, 1.585) |
| Adjusted model <sup>b</sup> | 1.405 (1.356, 1.455) | 1.559 (1.458, 1.667) | 1.902 (1.660, 2.179) | 1.371 (1.323, 1.422) | 1.218 (0.936, 1.585) |
| **Results by Age <sup>b</sup>** |  |  |  |  |  |
| 18-39 | 1.367 (1.083, 1.727) | 1.499 (0.674, 3.337) | 1.600 (0.523, 4.892) | 1.357 (1.069, 1.724) | 1.303 (0.757, 2.244) |
| 40-59 | 1.157 (1.045, 1.282) | 1.150 (0.922, 1.434) | 1.197 (0.806, 1.778) | 1.154 (1.038, 1.283) | 1.058 (0.658, 1.703) |
| 60+ | 1.444 (1.390, 1.500) | 1.617 (1.506, 1.736) | 2.028 (1.751, 2.349) | 1.405 (1.351, 1.461) | 1.288 (0.873, 1.902) |

Notes:
<sup>a</sup> Matched-on age, sex, race, ethnicity, vaccination site, month of receiving vaccine dose 1 and census tract
<sup>b</sup> Adjusted for age, sex, race, ethnicity, vaccination site, SVI scores at census tract level, average mortality rate at zip code level (2015-2019) and history of SARS-CoV-2 infection before vaccination

**Supplement Table 8.**
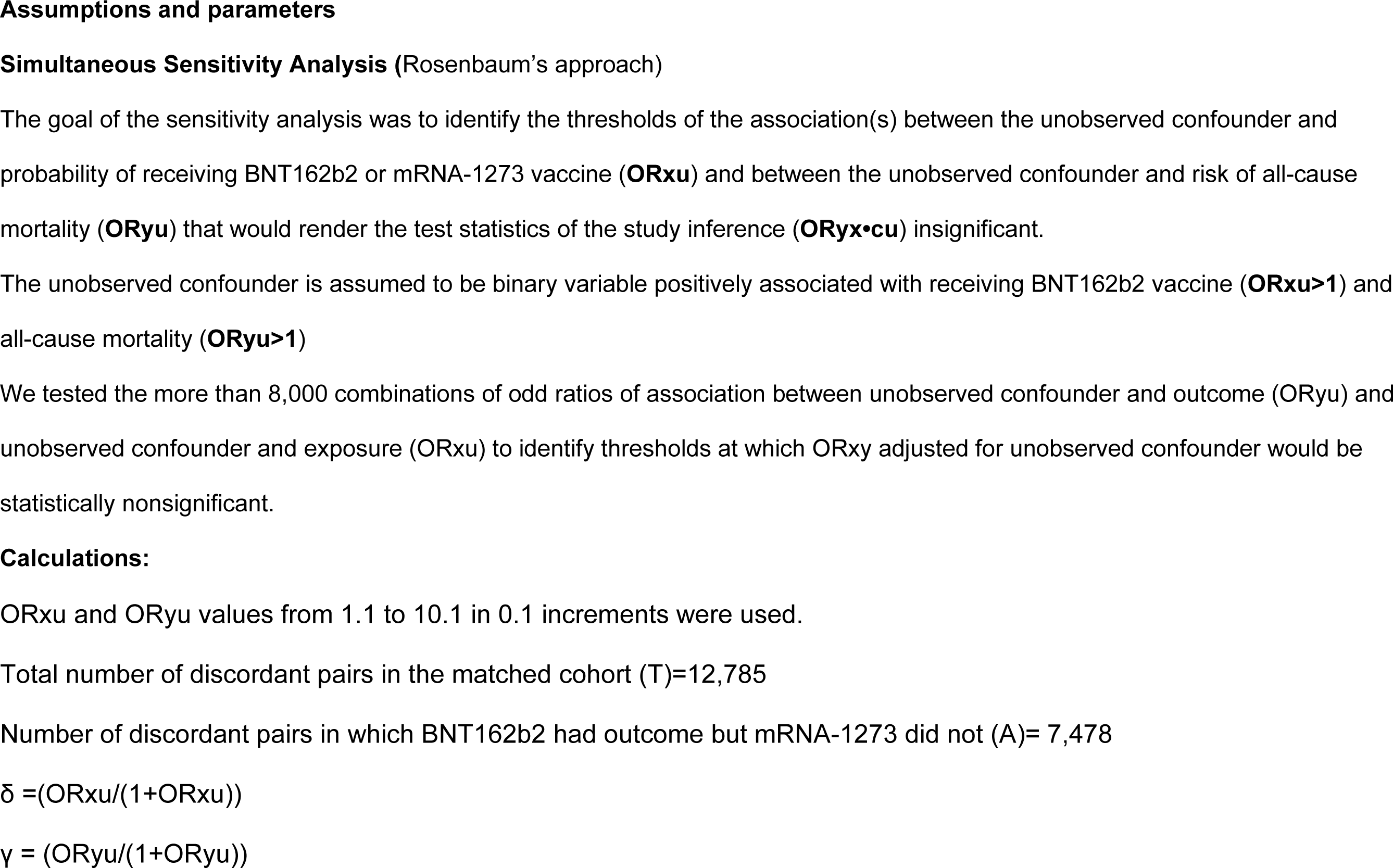
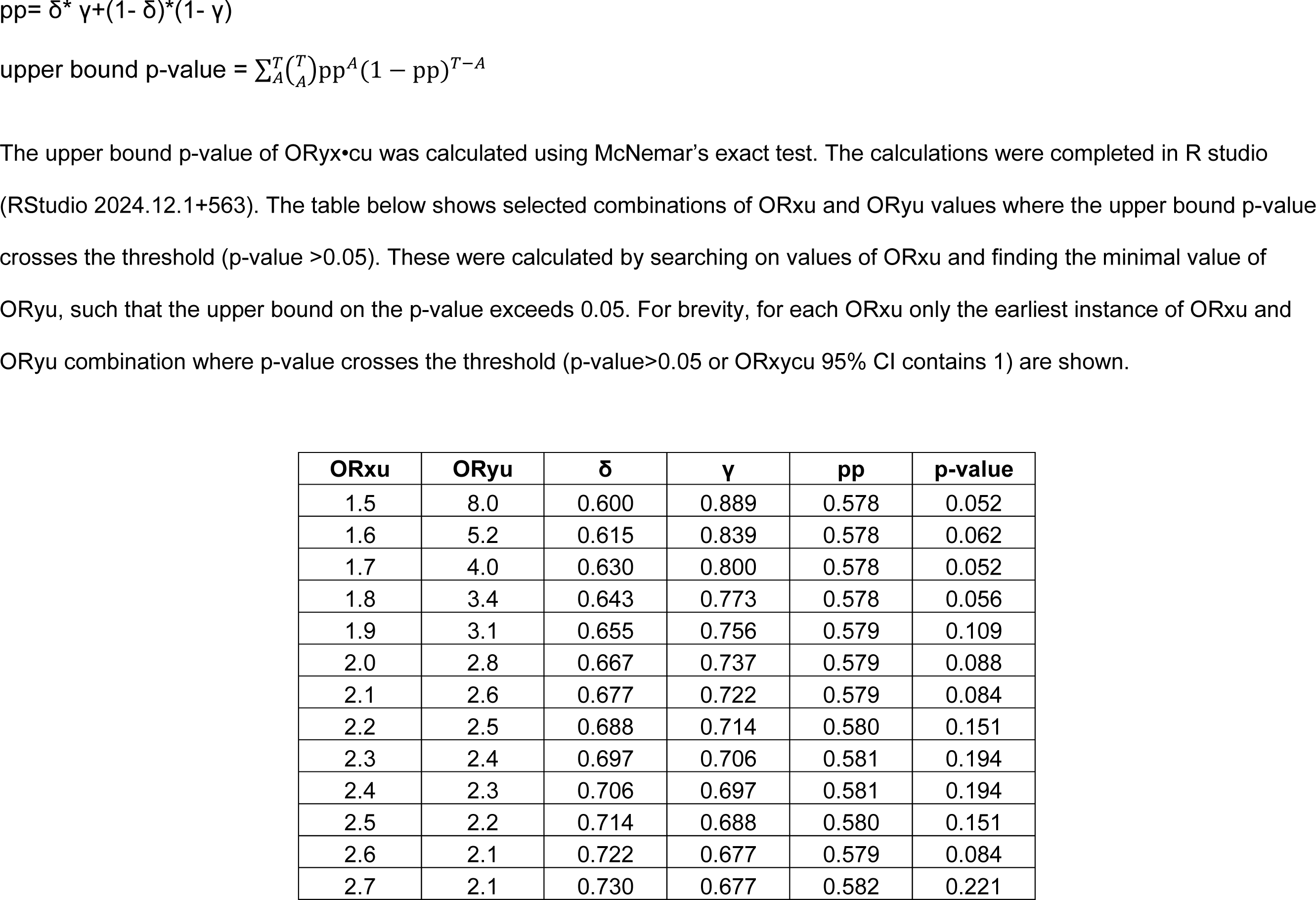
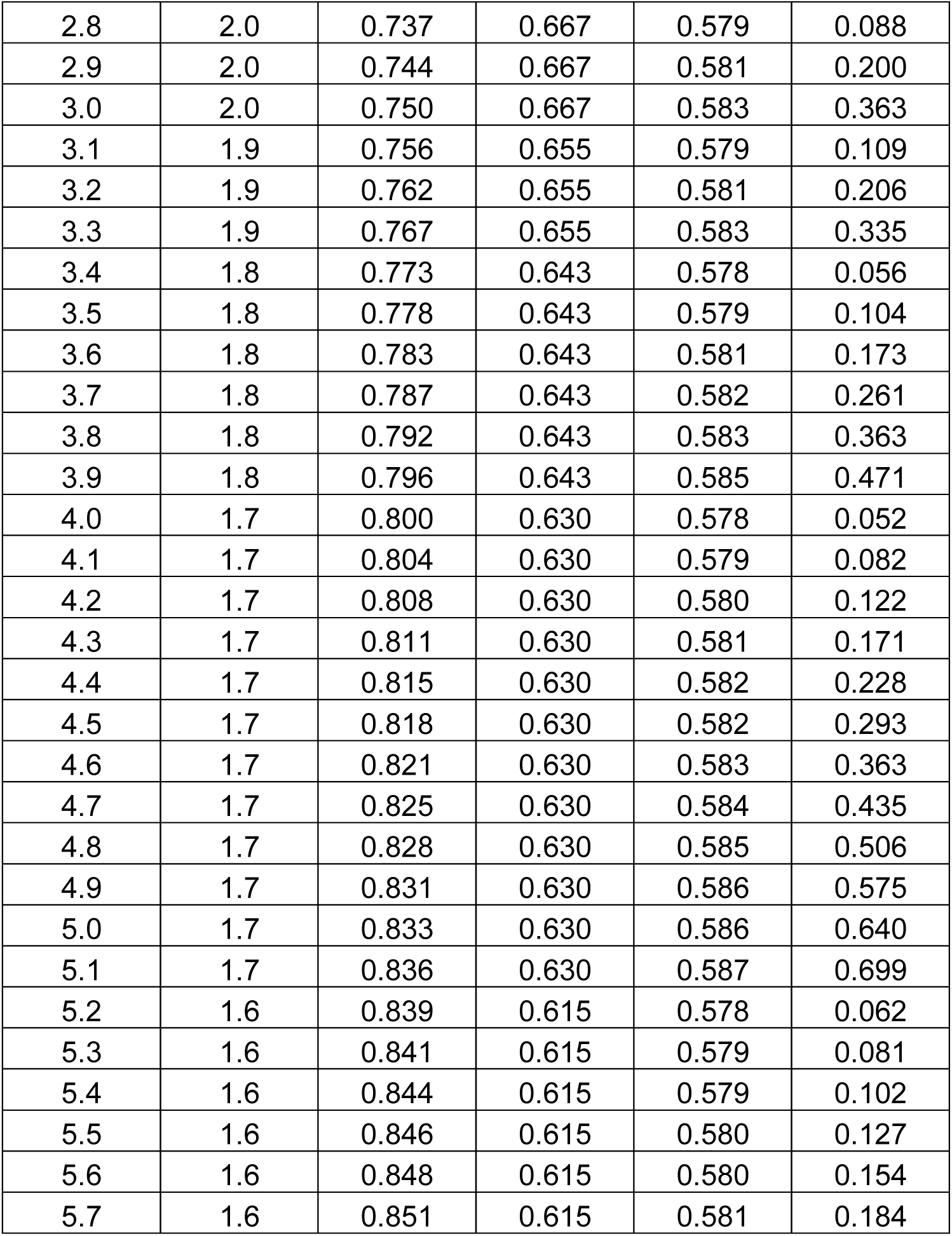
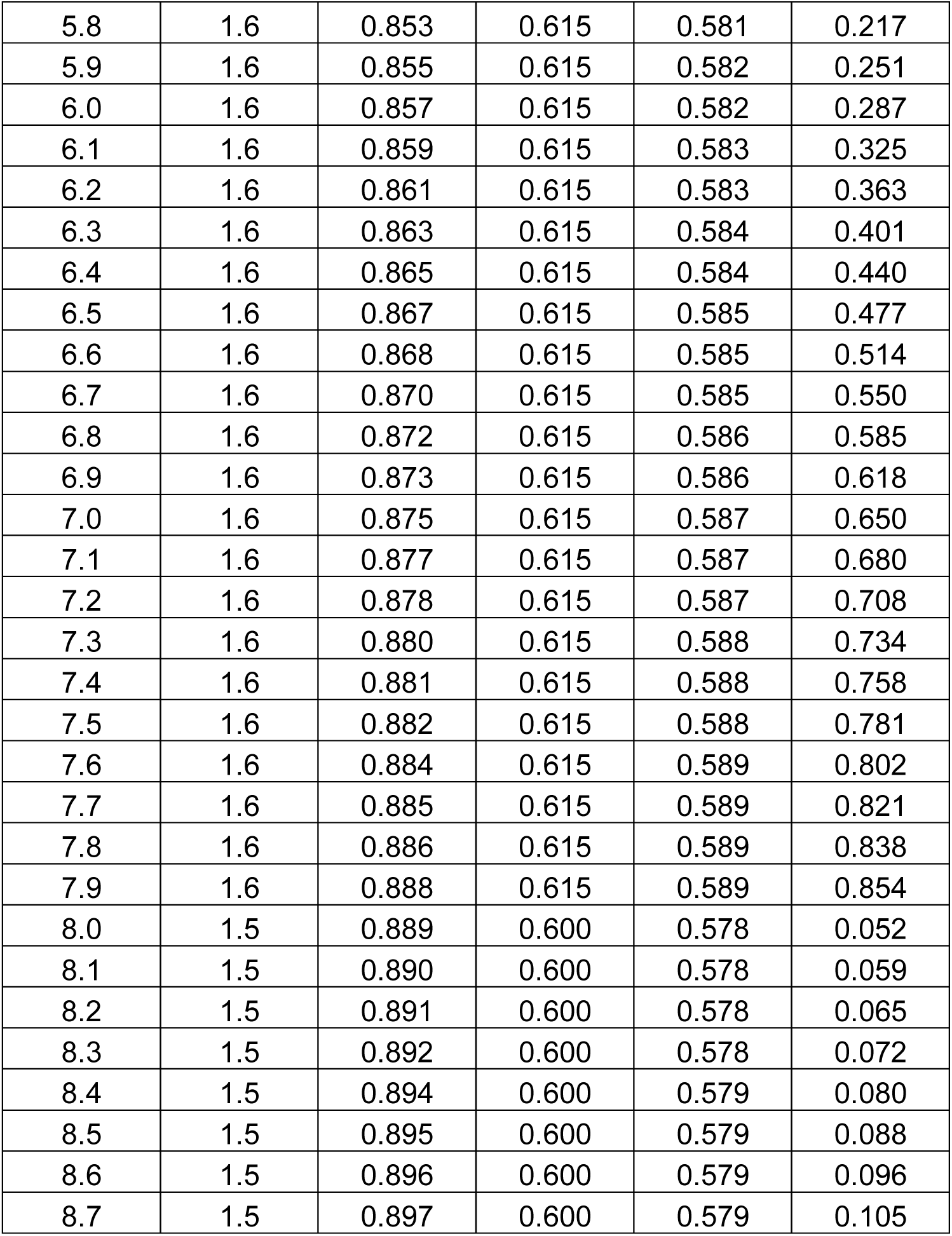
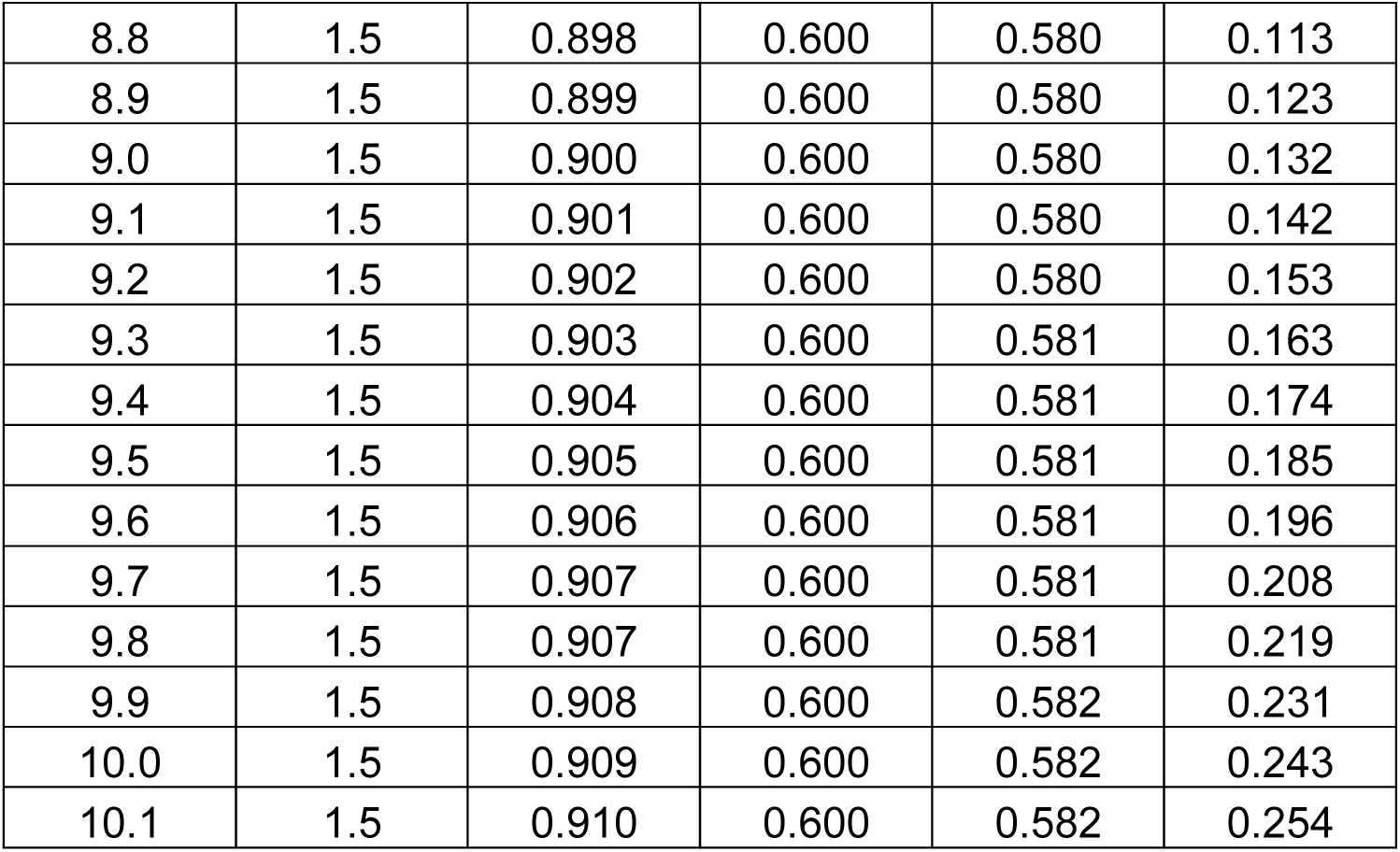
Sensitivity Analysis of Odds ratios of mortality from All causes in the Matched cohort to a binary unobserved confounder.

**Supplement Figure 1.**
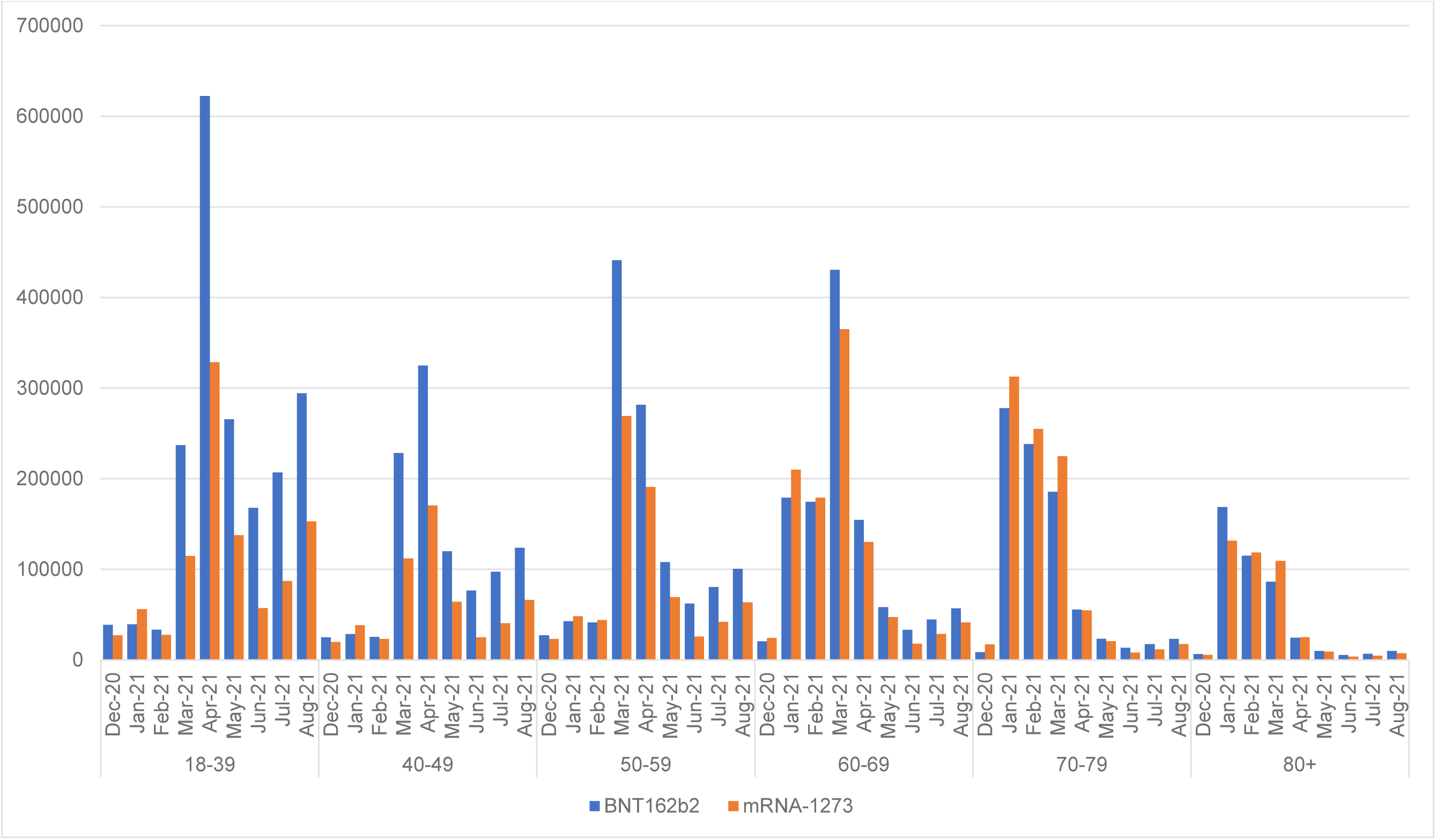
mRNA COVID-19 vaccination by age group and month of receiving dose 1 of vaccine

**Supplement Figure 2.**
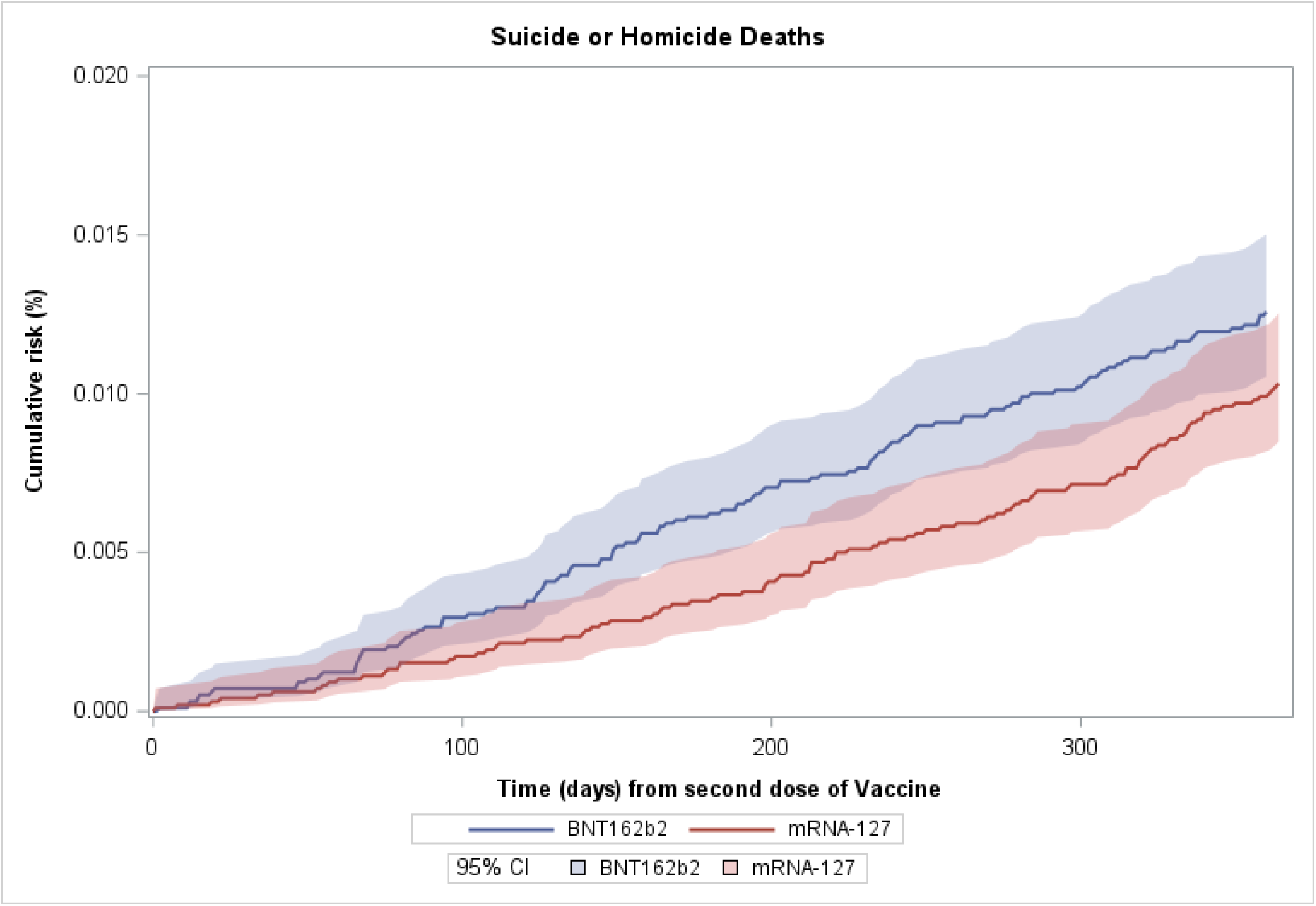
Cumulative incidence of Suicide or Homicide mortality in the matched cohort. The figure shows the cumulative incidence up to 365 days after the first dose of vaccine. Cumulative incidence was calculated using Kaplan Meier method and the follow-up ended either at the date of death or end of study observation period (365 days from first dose of vaccine). Deaths from all other causes were censored at the time of death.

**Supplement Figure 3.**
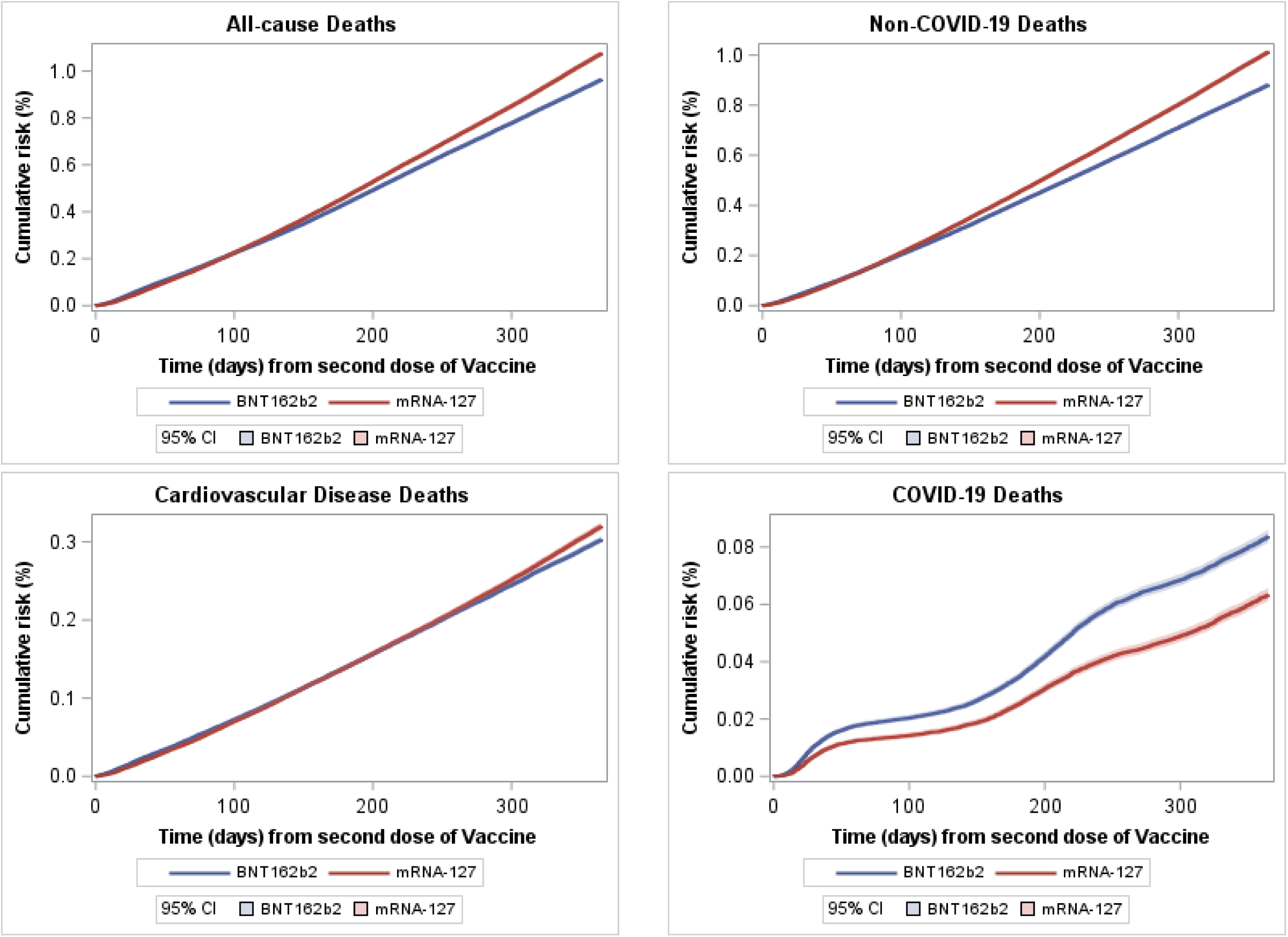
Cumulative incidence of all-cause, cardiovascular, COVID-19, and non-COVID-19 mortality in full cohort. The figure shows the cumulative incidence up to 365 days after the first dose of vaccine. Cumulative incidence is calculated using Kaplan Meier method and the follow-up ended either at the date of death or end of study observation period (365 days from first dose of vaccine). For cause-specific cumulative incidence, deaths from all other causes were censored at the time of death.

**Supplement Figure 4.**
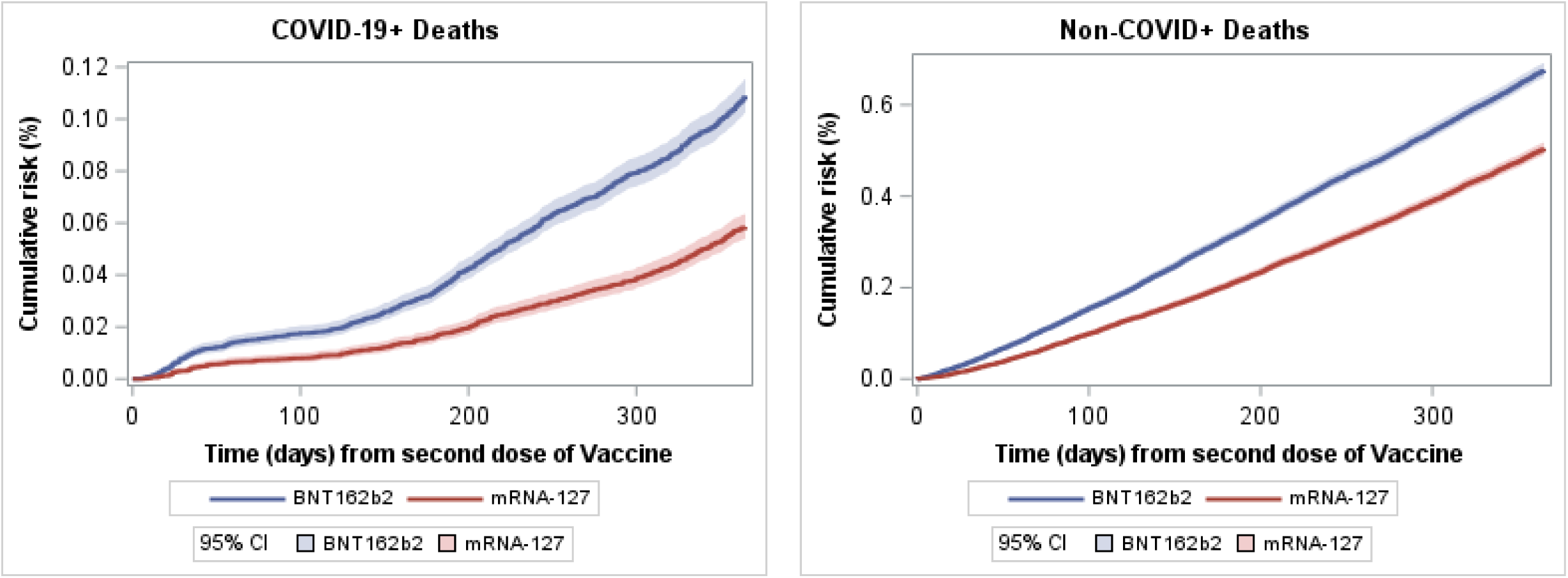
Cumulative incidence of COVID-19 +, and non-COVID-19 + mortality in matched cohort. The figure shows the cumulative incidence up to 365 days after the first dose of vaccine. Cumulative incidence is calculated using Kaplan Meier method and the follow-up ended either at the date of death or end of study observation period (365 days from first dose of vaccine). For cause-specific cumulative incidence, deaths from all other causes were censored at the time of death.

## Supplement Additional Methods. Identification of vaccinations in nursing homes, correctional facilities, and in homeless

Florida SHOTS records information on current residential address, site/location of vaccination, and vaccination provider. We utilized this information to identify vaccinees that are potentially residing in nursing homes, correctional facilities or were homeless. Vaccinees were identified as nursing home residents if their current address coincided with location of licensed nursing home, or their vaccination site was identified as a nursing home. Vaccinees with vaccination site identified as correctional facility were flagged as potential correctional facility residents. Lastly, vaccinees with homeless recorded in their current address in Florida shots were flagged as potentially homeless. These vaccinees represent institutionalized adults and hence were excluded from our analyses.

## Identification of deaths in Vaccinees

Florida Vital records were queried to identify potential deaths in Vaccinees. A multi-step algorithm was used to identify deaths based on demographic information from FL SHOTS and Florida Vital records. The records matched during each step were removed from the dataset and were not available in subsequent steps. The steps of algorithm are listed below.

FL SHOTS and Vital records matching algorithm:

1. Exact match on last name, first name, and date of birth, and fuzzy match on street address and city.
2. Exact match on last name, first name, and date of birth, and fuzzy match on city.
3. Exact match on last name, first name, and date of birth.
4. Exact match on last name, and date of birth
5. Exact match on last name and date of birth and fuzzy matching on city.
6. Exact match on last name and date of birth and fuzzy matching on first name.
7. Exact match on First name and date of birth and fuzzy matching on street address, city and last name.
8. Exact match on First name and date of birth and fuzzy matching on city and last name.
9. Exact match on First name and date of birth and fuzzy matching on last name.

