## Supplementary material for "Twelve-Month All-Cause Mortality after Initial COVID-19 Vaccination with Pfizer-BioNTech or mRNA-1273 among Adults Living in Florida": SAS Codes and Output File

### Table of Contents

| | |
|---|---|
| SAS CODE DOSE 1 combined | 13 |
| 1 Varible info | 14 |
| 1.1 The Contents Procedure | 14 |
| 1.1.1 WORK.DOSE1MATCHEDDATAFINAL | 14 |
| 1.1.1.1 Attributes | 14 |
| 1.1.1.2 Engine/Host Information | 14 |
| 1.1.1.3 Varnum | 15 |
| 1.1.1.4 Sortedby | 17 |
| 2 Dose 1 Descriptive Tables and Figures | 18 |
| 3 Dose 1 regression models main analysis | 86 |
| 4 Dose 1 regression models with and without covid | 173 |
| Matched cohort Risk of death Dose 1 Logistic All-cause 25JUN2025 | 265 |
| 1 The Logistic Procedure | 265 |
| 1.1 Model Information | 265 |
| 1.2 Observations Summary | 265 |
| 1.3 Response Profile | 265 |
| 1.4 Class Level Information | 265 |
| 1.5 Convergence Status | 265 |
| 1.6 Fit Statistics | 265 |
| 1.7 Global Tests | 265 |
| 1.8 Type 3 Tests | 266 |
| 1.9 Parameter Estimates | 266 |
| 1.10 Odds Ratios | 266 |
| 1.11 Association Statistics | 266 |
| Matched cohort Risk of death Dose 1 Logistic All-cause adjusted model (cat age) 25JUN2025 | 267 |
| 1 The Logistic Procedure | 267 |
| 1.1 Model Information | 267 |
| 1.2 Observations Summary | 267 |
| 1.3 Response Profile | 267 |
| 1.4 Class Level Information | 267 |
| 1.5 Convergence Status | 268 |
| 1.6 Fit Statistics | 268 |
| 1.7 Global Tests | 268 |
| 1.8 Type 3 Tests | 268 |
| 1.9 Parameter Estimates | 269 |
| 1.10 Odds Ratios | 269 |
| 1.11 Association Statistics | 270 |
| Matched cohort Risk of death Dose 1 logistic All-cause adjusted model (cat age) by Age (3 Age groups) 25JUN2025 | 271 |
| 1 The Logistic Procedure | 271 |
| 1.1 agenew=18-39 years | 271 |
| 1.1.1 Model Information | 271 |
| 1.1.2 Observations Summary | 271 |

1.1.3 Response Profile 271
1.1.4 Class Level Information 271
1.1.5 Convergence Status 272
1.1.6 Fit Statistics 272
1.1.7 Global Tests 272
1.1.8 Type 3 Tests 272
1.1.9 Parameter Estimates 273
1.1.10 Odds Ratios 273
1.1.11 Association Statistics 274
1.2 agenew=40-59 years 275
1.2.1 Model Information 275
1.2.2 Observations Summary 275
1.2.3 Response Profile 275
1.2.4 Class Level Information 275
1.2.5 Convergence Status 276
1.2.6 Fit Statistics 276
1.2.7 Global Tests 276
1.2.8 Type 3 Tests 276
1.2.9 Parameter Estimates 277
1.2.10 Odds Ratios 277
1.2.11 Association Statistics 278
1.3 agenew=60+ years 279
1.3.1 Model Information 279
1.3.2 Observations Summary 279
1.3.3 Response Profile 279
1.3.4 Class Level Information 279
1.3.5 Convergence Status 280
1.3.6 Fit Statistics 280
1.3.7 Global Tests 280
1.3.8 Type 3 Tests 280
1.3.9 Parameter Estimates 281
1.3.10 Odds Ratios 281
1.3.11 Association Statistics 282
Matched cohort Risk of death Dose 1 Logistic CVD DEATHS 25JUN2025 283
1 The Logistic Procedure 283
1.1 Model Information 283
1.2 Observations Summary 283
1.3 Response Profile 283
1.4 Class Level Information 283
1.5 Convergence Status 283
1.6 Fit Statistics 283
1.7 Global Tests 283
1.8 Type 3 Tests 284
1.9 Parameter Estimates 284
1.10 Odds Ratios 284

1.11 Association Statistics 284
Matched cohort Risk of death Dose 1 Logistic CVD Deaths adjusted model (cat age) 25JUN2025 285
1 The Logistic Procedure 285
1.1 Model Information 285
1.2 Observations Summary 285
1.3 Response Profile 285
1.4 Class Level Information 285
1.5 Convergence Status 286
1.6 Fit Statistics 286
1.7 Global Tests 286
1.8 Type 3 Tests 286
1.9 Parameter Estimates 287
1.10 Odds Ratios 287
1.11 Association Statistics 288
Matched cohort Risk of death Dose 1 logistic CVD DEATHS adjusted model (cat age) by Age (3 Age groups) 25JUN2025 289
1 The Logistic Procedure 289
1.1 agenew=18-39 years 289
1.1.1 Model Information 289
1.1.2 Observations Summary 289
1.1.3 Response Profile 289
1.1.4 Class Level Information 289
1.1.5 Convergence Status 290
1.1.6 Fit Statistics 290
1.1.7 Global Tests 290
1.1.8 Type 3 Tests 290
1.1.9 Parameter Estimates 291
1.1.10 Odds Ratios 291
1.1.11 Association Statistics 292
1.2 agenew=40-59 years 293
1.2.1 Model Information 293
1.2.2 Observations Summary 293
1.2.3 Response Profile 293
1.2.4 Class Level Information 293
1.2.5 Convergence Status 294
1.2.6 Fit Statistics 294
1.2.7 Global Tests 294
1.2.8 Type 3 Tests 294
1.2.9 Parameter Estimates 295
1.2.10 Odds Ratios 295
1.2.11 Association Statistics 296
1.3 agenew=60+ years 297
1.3.1 Model Information 297
1.3.2 Observations Summary 297
1.3.3 Response Profile 297

1.3.4 Class Level Information 297
1.3.5 Convergence Status 298
1.3.6 Fit Statistics 298
1.3.7 Global Tests 298
1.3.8 Type 3 Tests 298
1.3.9 Parameter Estimates 299
1.3.10 Odds Ratios 299
1.3.11 Association Statistics 300
Matched cohort Risk of death Dose 1 Logistic COVID DEATHS 25JUN2025 301
1 The Logistic Procedure 301
1.1 Model Information 301
1.2 Observations Summary 301
1.3 Response Profile 301
1.4 Class Level Information 301
1.5 Convergence Status 301
1.6 Fit Statistics 301
1.7 Global Tests 301
1.8 Type 3 Tests 302
1.9 Parameter Estimates 302
1.10 Odds Ratios 302
1.11 Association Statistics 302
Matched cohort Risk of death Dose 1 Logistic COVID Deaths adjusted model (cat age) 25JUN2025 303
1 The Logistic Procedure 303
1.1 Model Information 303
1.2 Observations Summary 303
1.3 Response Profile 303
1.4 Class Level Information 303
1.5 Convergence Status 304
1.6 Fit Statistics 304
1.7 Global Tests 304
1.8 Type 3 Tests 304
1.9 Parameter Estimates 305
1.10 Odds Ratios 305
1.11 Association Statistics 306
Matched cohort Risk of death Dose 1 logistic COVID DEATHS adjusted model (cat age) by Age (3 Age groups) 25JUN2025 307
1 The Logistic Procedure 307
1.1 agenew=18-39 years 307
1.1.1 Model Information 307
1.1.2 Observations Summary 307
1.1.3 Response Profile 307
1.1.4 Class Level Information 307
1.1.5 Convergence Status 308
1.1.6 Fit Statistics 308
1.1.7 Global Tests 308

1.1.8 Type 3 Tests 308
1.1.9 Parameter Estimates 309
1.1.10 Odds Ratios 309
1.1.11 Association Statistics 310
1.2 agenew=40-59 years 311
1.2.1 Model Information 311
1.2.2 Observations Summary 311
1.2.3 Response Profile 311
1.2.4 Class Level Information 311
1.2.5 Convergence Status 312
1.2.6 Fit Statistics 312
1.2.7 Global Tests 312
1.2.8 Type 3 Tests 312
1.2.9 Parameter Estimates 313
1.2.10 Odds Ratios 313
1.2.11 Association Statistics 314
1.3 agenew=60+ years 315
1.3.1 Model Information 315
1.3.2 Observations Summary 315
1.3.3 Response Profile 315
1.3.4 Class Level Information 315
1.3.5 Convergence Status 316
1.3.6 Fit Statistics 316
1.3.7 Global Tests 316
1.3.8 Type 3 Tests 316
1.3.9 Parameter Estimates 317
1.3.10 Odds Ratios 317
1.3.11 Association Statistics 318
Matched cohort Risk of death Dose 1 Logistic COVID Deaths adjusted model (cat age) 25JUN2025 319
1 The Logistic Procedure 319
1.1 Model Information 319
1.2 Observations Summary 319
1.3 Response Profile 319
1.4 Class Level Information 319
1.5 Convergence Status 320
1.6 Fit Statistics 320
1.7 Global Tests 320
1.8 Type 3 Tests 320
1.9 Parameter Estimates 321
1.10 Odds Ratios 321
1.11 Association Statistics 322
Matched cohort Risk of death Dose 1 Logistic NON COVID DEATHS 25JUN2025 323
1 The Logistic Procedure 323
1.1 Model Information 323
1.2 Observations Summary 323

1.3 Response Profile 323
1.4 Class Level Information 323
1.5 Convergence Status 323
1.6 Fit Statistics 323
1.7 Global Tests 323
1.8 Type 3 Tests 324
1.9 Parameter Estimates 324
1.10 Odds Ratios 324
1.11 Association Statistics 324
Matched cohort Risk of death Dose 1 logistic Non COVID DEATHS adjusted model (cat age) by Age (3 Age groups) 25JUN2025 325
1 The Logistic Procedure 325
1.1 agenew=18-39 years 325
1.1.1 Model Information 325
1.1.2 Observations Summary 325
1.1.3 Response Profile 325
1.1.4 Class Level Information 325
1.1.5 Convergence Status 326
1.1.6 Fit Statistics 326
1.1.7 Global Tests 326
1.1.8 Type 3 Tests 326
1.1.9 Parameter Estimates 327
1.1.10 Odds Ratios 327
1.1.11 Association Statistics 328
1.2 agenew=40-59 years 329
1.2.1 Model Information 329
1.2.2 Observations Summary 329
1.2.3 Response Profile 329
1.2.4 Class Level Information 329
1.2.5 Convergence Status 330
1.2.6 Fit Statistics 330
1.2.7 Global Tests 330
1.2.8 Type 3 Tests 330
1.2.9 Parameter Estimates 331
1.2.10 Odds Ratios 331
1.2.11 Association Statistics 332
1.3 agenew=60+ years 333
1.3.1 Model Information 333
1.3.2 Observations Summary 333
1.3.3 Response Profile 333
1.3.4 Class Level Information 333
1.3.5 Convergence Status 334
1.3.6 Fit Statistics 334
1.3.7 Global Tests 334
1.3.8 Type 3 Tests 334
1.3.9 Parameter Estimates 335

1.3.10 Odds Ratios 335
1.3.11 Association Statistics 336
Matched cohort Risk of death Dose 1 Logistic COVID + 26JUN2025 337
1 The Logistic Procedure 337
1.1 Model Information 337
1.2 Observations Summary 337
1.3 Response Profile 337
1.4 Class Level Information 337
1.5 Convergence Status 337
1.6 Fit Statistics 337
1.7 Global Tests 337
1.8 Type 3 Tests 338
1.9 Parameter Estimates 338
1.10 Odds Ratios 338
1.11 Association Statistics 338
Matched cohort Risk of death Dose 1 Logistic COVID + adjusted model (cat age) 26JUN2025 339
1 The Logistic Procedure 339
1.1 Model Information 339
1.2 Observations Summary 339
1.3 Response Profile 339
1.4 Class Level Information 339
1.5 Convergence Status 340
1.6 Fit Statistics 340
1.7 Global Tests 340
1.8 Type 3 Tests 340
1.9 Parameter Estimates 341
1.10 Odds Ratios 341
1.11 Association Statistics 342
Matched cohort Risk of death Dose 1 logistic COVID + adjusted model (cat age) by Age (3 Age groups) 26JUN2025 343
1 The Logistic Procedure 343
1.1 agenew=18-39 years 343
1.1.1 Model Information 343
1.1.2 Observations Summary 343
1.1.3 Response Profile 343
1.1.4 Class Level Information 343
1.1.5 Convergence Status 344
1.1.6 Fit Statistics 344
1.1.7 Global Tests 344
1.1.8 Type 3 Tests 344
1.1.9 Parameter Estimates 345
1.1.10 Odds Ratios 345
1.1.11 Association Statistics 346
1.2 agenew=40-59 years 347
1.2.1 Model Information 347

1.2.2 Observations Summary 347
1.2.3 Response Profile 347
1.2.4 Class Level Information 347
1.2.5 Convergence Status 348
1.2.6 Fit Statistics 348
1.2.7 Global Tests 348
1.2.8 Type 3 Tests 348
1.2.9 Parameter Estimates 349
1.2.10 Odds Ratios 349
1.2.11 Association Statistics 350
1.3 agenew=60+ years 351
1.3.1 Model Information 351
1.3.2 Observations Summary 351
1.3.3 Response Profile 351
1.3.4 Class Level Information 351
1.3.5 Convergence Status 352
1.3.6 Fit Statistics 352
1.3.7 Global Tests 352
1.3.8 Type 3 Tests 352
1.3.9 Parameter Estimates 353
1.3.10 Odds Ratios 353
1.3.11 Association Statistics 354
Matched cohort Risk of death Dose 1 Logistic COVID + 30JUN2025 355
1 The Logistic Procedure 355
1.1 Model Information 355
1.2 Observations Summary 355
1.3 Response Profile 355
1.4 Class Level Information 355
1.5 Convergence Status 355
1.6 Fit Statistics 355
1.7 Global Tests 355
1.8 Type 3 Tests 356
1.9 Parameter Estimates 356
1.10 Odds Ratios 356
1.11 Association Statistics 356
Matched cohort Risk of death Dose 1 Logistic NON COVID + adjusted model (cat age) 30JUN2025 357
1 The Logistic Procedure 357
1.1 Model Information 357
1.2 Observations Summary 357
1.3 Response Profile 357
1.4 Class Level Information 357
1.5 Convergence Status 358
1.6 Fit Statistics 358
1.7 Global Tests 358
1.8 Type 3 Tests 358

1.9 Parameter Estimates 359
1.10 Odds Ratios 359
1.11 Association Statistics 360
Matched cohort Risk of death Dose 1 logistic NON COVID + adjusted model (cat age) by Age (3 Age groups) 30JUN2025 361
1 The Logistic Procedure 361
1.1 agenew=18-39 years 361
1.1.1 Model Information 361
1.1.2 Observations Summary 361
1.1.3 Response Profile 361
1.1.4 Class Level Information 361
1.1.5 Convergence Status 362
1.1.6 Fit Statistics 362
1.1.7 Global Tests 362
1.1.8 Type 3 Tests 362
1.1.9 Parameter Estimates 363
1.1.10 Odds Ratios 363
1.1.11 Association Statistics 364
1.2 agenew=40-59 years 365
1.2.1 Model Information 365
1.2.2 Observations Summary 365
1.2.3 Response Profile 365
1.2.4 Class Level Information 365
1.2.5 Convergence Status 366
1.2.6 Fit Statistics 366
1.2.7 Global Tests 366
1.2.8 Type 3 Tests 366
1.2.9 Parameter Estimates 367
1.2.10 Odds Ratios 367
1.2.11 Association Statistics 368
1.3 agenew=60+ years 369
1.3.1 Model Information 369
1.3.2 Observations Summary 369
1.3.3 Response Profile 369
1.3.4 Class Level Information 369
1.3.5 Convergence Status 370
1.3.6 Fit Statistics 370
1.3.7 Global Tests 370
1.3.8 Type 3 Tests 370
1.3.9 Parameter Estimates 371
1.3.10 Odds Ratios 371
1.3.11 Association Statistics 372
Matched cohort Risk of death Dose 1 Logistic SUICHOM 27JUN2025 373
1 The Logistic Procedure 373
1.1 Model Information 373
1.2 Observations Summary 373

1.3 Response Profile 373
1.4 Class Level Information 373
1.5 Convergence Status 373
1.6 Fit Statistics 373
1.7 Global Tests 373
1.8 Type 3 Tests 374
1.9 Parameter Estimates 374
1.10 Odds Ratios 374
1.11 Association Statistics 374
Matched cohort Risk of death Dose 1 Logistic SUICHOM adjusted model (cat age) 27JUN2025 375
1 The Logistic Procedure 375
1.1 Model Information 375
1.2 Observations Summary 375
1.3 Response Profile 375
1.4 Class Level Information 375
1.5 Convergence Status 376
1.6 Fit Statistics 376
1.7 Global Tests 376
1.8 Type 3 Tests 376
1.9 Parameter Estimates 377
1.10 Odds Ratios 377
1.11 Association Statistics 378
Matched cohort Risk of death Dose 1 logistic SUICHOM adjusted model (cat age) by Age (3 Age groups) 27JUN2025 379
1 The Logistic Procedure 379
1.1 agenew=18-39 years 379
1.1.1 Model Information 379
1.1.2 Observations Summary 379
1.1.3 Response Profile 379
1.1.4 Class Level Information 379
1.1.5 Convergence Status 380
1.1.6 Fit Statistics 380
1.1.7 Global Tests 380
1.1.8 Type 3 Tests 380
1.1.9 Parameter Estimates 381
1.1.10 Odds Ratios 381
1.1.11 Association Statistics 382
1.2 agenew=40-59 years 383
1.2.1 Model Information 383
1.2.2 Observations Summary 383
1.2.3 Response Profile 383
1.2.4 Class Level Information 383
1.2.5 Convergence Status 384
1.2.6 Fit Statistics 384
1.2.7 Global Tests 384

1.2.8 Type 3 Tests 384
1.2.9 Parameter Estimates 385
1.2.10 Odds Ratios 385
1.2.11 Association Statistics 386
1.3 agenew=60+ years 387
1.3.1 Model Information 387
1.3.2 Observations Summary 387
1.3.3 Response Profile 387
1.3.4 Class Level Information 387
1.3.5 Convergence Status 388
1.3.6 Fit Statistics 388
1.3.7 Global Tests 388
1.3.8 Type 3 Tests 388
1.3.9 Parameter Estimates 389
1.3.10 Odds Ratios 389
1.3.11 Association Statistics 390
Matched cohort Risk of death Dose 1 Logistic All-cause 24JUN2025 391
1 The Logistic Procedure 391
1.1 Model Information 391
1.2 Observations Summary 391
1.3 Response Profile 391
1.4 Class Level Information 391
1.5 Convergence Status 391
1.6 Fit Statistics 391
1.7 Global Tests 391
1.8 Type 3 Tests 392
1.9 Parameter Estimates 392
1.10 Odds Ratios 392
1.11 Association Statistics 392
Matched cohort Risk of death Dose 1 Logistic All-cause adjusted model (cat age) 24JUN2025 393
1 The Logistic Procedure 393
1.1 Model Information 393
1.2 Observations Summary 393
1.3 Response Profile 393
1.4 Class Level Information 393
1.5 Convergence Status 394
1.6 Fit Statistics 394
1.7 Global Tests 394
1.8 Type 3 Tests 394
1.9 Parameter Estimates 395
1.10 Odds Ratios 395
1.11 Association Statistics 396
Matched cohort Risk of death Dose 1 logistic All-cause adjusted model (cat age) by Age (3 Age groups) 24JUN2025 397
1 The Logistic Procedure 397

1.1 agenew=18-39 years 397
1.1.1 Model Information 397
1.1.2 Observations Summary 397
1.1.3 Response Profile 397
1.1.4 Class Level Information 397
1.1.5 Convergence Status 398
1.1.6 Fit Statistics 398
1.1.7 Global Tests 398
1.1.8 Type 3 Tests 398
1.1.9 Parameter Estimates 399
1.1.10 Odds Ratios 399
1.1.11 Association Statistics 400
1.2 agenew=40-59 years 401
1.2.1 Model Information 401
1.2.2 Observations Summary 401
1.2.3 Response Profile 401
1.2.4 Class Level Information 401
1.2.5 Convergence Status 402
1.2.6 Fit Statistics 402
1.2.7 Global Tests 402
1.2.8 Type 3 Tests 402
1.2.9 Parameter Estimates 403
1.2.10 Odds Ratios 403
1.2.11 Association Statistics 404
1.3 agenew=60+ years 405
1.3.1 Model Information 405
1.3.2 Observations Summary 405
1.3.3 Response Profile 405
1.3.4 Class Level Information 405
1.3.5 Convergence Status 406
1.3.6 Fit Statistics 406
1.3.7 Global Tests 406
1.3.8 Type 3 Tests 406
1.3.9 Parameter Estimates 407
1.3.10 Odds Ratios 407
1.3.11 Association Statistics 408

### Table of Contents

| | |
|---|---|
| Varible info | 2 |
| 1 The Contents Procedure | 2 |
| 1.1 WORK.DOSE1MATCHEDDATAFINAL | 2 |
| 1.1.1 Attributes | 2 |
| 1.1.2 Engine/Host Information | 2 |
| 1.1.3 Varnum | 3 |
| 1.1.4 Sortedby | 5 |
| Dose 1 Descriptive Tables and Figures | 6 |
| Dose 1 regression models main analysis | 74 |
| Dose 1 regression models with and without covid | 161 |

### Final Dataset with Variables information

#### The CONTENTS Procedure

| | | | |
|---|---|---|---|
| **Data Set Name** | WORK.DOSE1MATCHEDDATAFINAL | **Observations** | 11288333 |
| **Member Type** | DATA | **Variables** | 80 |
| **Engine** | V9 | **Indexes** | 0 |
| **Created** | 07/18/2025 12:41:53 | **Observation Length** | 608 |
| **Last Modified** | 07/18/2025 12:41:53 | **Deleted Observations** | 0 |
| **Protection** | | **Compressed** | NO |
| **Data Set Type** | | **Sorted** | YES |
| **Label** | | | |
| **Data Representation** | WINDOWS_64 | | |
| **Encoding** | wlatin1 Western (Windows) | | |

| Engine/Host Dependent Information | |
|---|---|
| **Data Set Page Size** | 65536 |
| **Number of Data Set Pages** | 105499 |
| **First Data Page** | 1 |
| **Max Obs per Page** | 107 |
| **Obs in First Data Page** | 92 |
| **Number of Data Set Repairs** | 0 |
| **ExtendObsCounter** | YES |
| **Filename** | C:\Users\MansuriF\AppData\Local\Temp\SAS Temporary Files_TD16304_HSD00MLT057133_\dose1matcheddatafinal.sas7bdat |
| **Release Created** | 9.0401M7 |
| **Host Created** | X64_10PRO |
| **Owner Name** | DOHUSERS\MansuriF |
| **File Size** | 6GB |
| **File Size (bytes)** | 6914048000 |

#### The CONTENTS Procedure

| Variables in Creation Order | | | | | | |
|---|---|---|---|---|---|---|
| # | Variable | Type | Len | Format | Informat | Label |
| 1 | DWHPatient | Num | 8 | 11. | 11. | DWHPatient |
| 2 | VaccinationDate1 | Num | 8 | DATE9. | DATE9. | VaccinationDate |
| 3 | VaccinationDate2 | Num | 8 | DATE9. | DATE9. | VaccinationDate |
| 4 | CurrentStateIMMId | Char | 10 | $10. | $10. | CurrentStateIMMId |
| 5 | timebetweendose | Num | 8 | | | |
| 6 | vaccinedose2 | Num | 8 | | | |
| 7 | ageatdose1 | Num | 8 | | | |
| 8 | firstdose | Char | 9 | | | |
| 9 | geoidnew | Num | 8 | | | |
| 10 | seconddose | Char | 9 | | | |
| 11 | timeokay | Num | 8 | | | |
| 12 | homelessflag | Num | 8 | | | |
| 13 | ETHNICITY3 | Char | 20 | | | |
| 14 | RACE4 | Char | 7 | | | |
| 15 | sex2 | Char | 1 | | | |
| 16 | agegroupnew | Char | 5 | | | |
| 17 | Vaccinationsite | Char | 26 | | | |
| 18 | VS DOD | Num | 8 | DATE9. | DATE9. | VS DOD |
| 19 | TRANSAX1 | Char | 5 | $5. | $5. | TRANSAX1 |
| 20 | TRANSAX2 | Char | 8 | $8. | $8. | TRANSAX2 |
| 21 | TRANSAX3 | Char | 5 | $5. | $5. | TRANSAX3 |
| 22 | TRANSAX4 | Char | 5 | $5. | $5. | TRANSAX4 |
| 23 | TRANSAX5 | Char | 5 | $5. | $5. | TRANSAX5 |
| 24 | TRANSAX6 | Char | 5 | $5. | $5. | TRANSAX6 |
| 25 | TRANSAX7 | Char | 4 | $4. | $4. | TRANSAX7 |
| 26 | TRANSAX8 | Char | 4 | $4. | $4. | TRANSAX8 |
| 27 | TRANSAX9 | Char | 4 | $4. | $4. | TRANSAX9 |
| 28 | TRANSAX10 | Char | 4 | $4. | $4. | TRANSAX10 |
| 29 | TRANSAX11 | Char | 4 | $4. | $4. | TRANSAX11 |
| 30 | TRANSAX12 | Char | 4 | $4. | $4. | TRANSAX12 |
| 31 | TRANSAX13 | Char | 4 | $4. | $4. | TRANSAX13 |
| 32 | TRANSAX14 | Char | 4 | $4. | $4. | TRANSAX14 |
| 33 | TRANSAX15 | Char | 4 | $4. | $4. | TRANSAX15 |
| 34 | ICD_CODE | Char | 4 | $4. | $4. | ICD_CODE |
| 35 | daystodeath | Num | 8 | | | |

#### The CONTENTS Procedure

| Variables in Creation Order | | | | | | |
|---|---|---|---|---|---|---|
| # | Variable | Type | Len | Format | Informat | Label |
| 36 | death1yr | Num | 8 | | | |
| 37 | daystodeathd1 | Num | 8 | | | |
| 38 | death1yrd1 | Num | 8 | | | |
| 39 | mortalityratepct | Num | 8 | | | |
| 40 | agenew | Num | 8 | AGE10YGROUP. | | |
| 41 | SVISCORE | Num | 8 | 8.2 | | |
| 42 | SVISCORE1 | Num | 8 | 8.2 | | |
| 43 | SVISCORE2 | Num | 8 | 8.2 | | |
| 44 | SVISCORE3 | Num | 8 | 8.2 | | |
| 45 | SVISCORE4 | Num | 8 | 8.2 | | |
| 46 | nursinghomevaccinationfinal | Num | 8 | | | |
| 47 | monthdose1 | Num | 8 | | | |
| 48 | matchedpairs | Num | 8 | | | |
| 49 | decile_sviscore1 | Num | 8 | | | Rank for Variable SVISCORE1 |
| 50 | decile_sviscore4 | Num | 8 | | | Rank for Variable SVISCORE4 |
| 51 | ELD | Char | 8 | | | NAME OF FORMER VARIABLE |
| 52 | ELD1 | Num | 8 | DATE9. | | |
| 53 | ELD2 | Num | 8 | DATE9. | | |
| 54 | ELD3 | Num | 8 | DATE9. | | |
| 55 | ELD4 | Num | 8 | DATE9. | | |
| 56 | ELD5 | Num | 8 | DATE9. | | |
| 57 | ELD6 | Num | 8 | DATE9. | | |
| 58 | noseconddose | Num | 8 | | | |
| 59 | EXPOS | Num | 8 | | | |
| 60 | CVD1YRD1 | Num | 8 | | | |
| 61 | overdose1yrD1 | Num | 8 | | | |
| 62 | homicide1yrD1 | Num | 8 | | | |
| 63 | SUICIDE1YRD1 | Num | 8 | | | |
| 64 | SUICIDE1YR | Num | 8 | | | |
| 65 | UNINTENTIONAL1YRD1 | Num | 8 | | | |
| 66 | NOFALLUNINTENTIONAL1YRD1 | Num | 8 | | | |
| 67 | unnatural1yrD1 | Num | 8 | | | |
| 68 | suicidehomicide1yrD1 | Num | 8 | | | |
| 69 | NOFALLunnatural1yrD1 | Num | 8 | | | |
| 70 | mva1yrD1 | Num | 8 | | | |

#### The CONTENTS Procedure

| Variables in Creation Order | | | | | | |
|---|---|---|---|---|---|---|
| **#** | **Variable** | **Type** | **Len** | **Format** | **Informat** | **Label** |
| **71** | count | Num | 8 | | | |
| **72** | COVIDDEATH | Num | 8 | | | |
| **73** | COVIDDEATH1YRD1 | Num | 8 | | | |
| **74** | NONCOVIDDEATH1YRD1 | Num | 8 | | | |
| **75** | counteld | Num | 8 | | | |
| **76** | COVIDDEATHPLUS1YRD1 | Num | 8 | | | |
| **77** | NONCOVIDDEATHPLUS1YRD1 | Num | 8 | | | |
| **78** | covinftodeathdys | Num | 8 | | | |
| **79** | countcovinf | Num | 8 | | | |
| **80** | countcovinfbf20 | Num | 8 | | | |

| Sort Information | |
|---|---|
| **Sortedby** | firstdose |
| **Validated** | YES |
| **Character Set** | ANSI |

```
libname store 'C:\Users\MansuriF\OneDrive - Florida Department of
Health\Documents\SAS Datasets';

run;

LIBNAME Deaths ODBC NOPROMPT="driver={ODBC Driver 17 for SQL Server};

Server=CHARTSDATA.DOH.AD.STATE.FL.US;

Database=Vitalstat;

Trusted_Connection=Yes"  schema=dbo stringdates=NO ;

run;

LIBNAME COVVAC ODBC NOPROMPT="Driver={ODBC Driver 17 for SQL Server};

        Server=merlindbreport.doh.ad.state.fl.us;

        Database=CovidVaccine;Trusted_Connection=Yes" schema=dbo stringdates=Yes;

run;

LIBNAME COVID ODBC NOPROMPT="Driver={ODBC Driver 17 for SQL Server};

        Server=merlindbreport.doh.ad.state.fl.us;

        Database=MerlinCOVID;Trusted_Connection=Yes" schema=dbo stringdates=Yes;

run;

options validvarname=any;

%include "H:\EPI\Surveillance Section\Fahad Mansuri\SURVIVAL ANALYSIS 2024\SAS
CODE\formats for analysis.sas";
```

```
%include "H:\EPI\Surveillance Section\Fahad Mansuri\SURVIVAL ANALYSIS 2024\SAS
CODE\relrisk9.sas";

proc means data=store.dose1matcheddata nmiss;
	var sviscore:;
run;

proc stdize data=store.dose1matcheddata reponly method=mean missing=mean
out=dose1matcheddata;
	var sviscore:;
run;

proc means data=dose1matcheddata nmiss;
	var sviscore:;
run;

/*** Descriptives for matched data vs all data ***/

proc rank data=dose1matcheddata out=dose1matcheddata groups=10;
	var sviscore1 sviscore2 sviscore3 sviscore4 sviscore SESPOV
	SESUNEMP SESINCOME	SESNOHIGH	HTMUS	HTMOBILEHOME
	HTNOVEHICLE	HTCROWDING	HTGQ;
	ranks decile_sviscore1 decile_sviscore2 decile_sviscore3 decile_sviscore4
decile_sviscore
	decile_SESPOV	decile_SESUNEMP decile_SESINCOME	decile_SESNOHIGH
	decile_HTMUS	decile_HTMOBILEHOME
	decile_HTNOVEHICLE	decile_HTCROWDING	decile_HTGQ;
```

```
run;

proc contents data=store.dose1matcheddata order=varnum;
run;

proc sort data=dose1matcheddata nodupkey out=test;
	by DWHPatient;
run;

LIBNAME COVID ODBC NOPROMPT="Driver={ODBC Driver 17 for SQL Server};
	Server=merlindbreport.doh.ad.state.fl.us;
	Database=MerlinCOVID;Trusted_Connection=Yes" schema=dbo stringdates=no;
run;

proc contents data=covid.cases order=varnum;
run;

proc contents data=COVVAC.COVIDPatientIdentity order=varnum;
run;

data covidcases;
	set covid.cases;
	keep 'Case ID'n 'Profile ID'n 'shots id'n 'Publish case'n Jurisdiction
	'Earliest lab event date'n 'Date became case'n 'Dx status'n;
	rename 'shots id'n =shotsid2 'Publish case'n =Publishcase
	'Earliest lab event date'n=ELD
```

```
	'Date became case'n =casedate
	'Dx status'n=dxstatus 'Profile ID'n=profileid 'Case ID'n=caseid;
	where upcase('Publish case'n)="YES" and  "01Mar2020"d le 'Earliest lab event 
date'n le "31dec2022"D
	and	Jurisdiction="FL resident";
run;

proc sql;
	select count(distinct profileid)
	from covidcases
	where shotsid2 ne " ";
quit;

proc sql;
	select count (profileid)
	from covidcases
	where shotsid2 ne " ";
quit;

proc sql;
	select count(distinct caseid)
	from covidcases	where shotsid2 ne " ";
quit;

proc sql;
	select count(caseid)
```

```
        from covidcases        where shotsid2 ne " ";
quit;

proc sql;
        select count (distinct shotsid2)
                from covidcases        where shotsid2 ne " ";
quit;

proc sql;
        select count (shotsid2)
                from covidcases        where shotsid2 ne " ";
quit;

proc sort data=work.covidcases;
        by shotsid2;
run;

proc transpose data=covidcases out=covidcases2 name=ELD prefix=ELD;
        by shotsid2;
        var eld;
        where shotsid2 ne " ";
run;

proc freq data=covidcases;
        table Jurisdiction dxstatus;
run;
```

```
proc contents data=store.dose1matcheddata;
run;

proc datasets lib=work;
run;

proc sort data=dose1matcheddata;
	by shotsid2;
run;

proc sort data=covidcases2;
	by shotsid2;
run;

proc sql;
	create table dose1matchedatawithcovid as
	select a.*,b.*
	from dose1matcheddata as a
	left join covidcases2 as b
	on a.CurrentStateIMMId=b.shotsid2;
quit;

options validvarname=any;
```

```
proc contents data=store.covidvaccination5 order=varnum;
run;

data dose1matcheddatafinal;
	set dose1matchedatawithcovid (keep= ageatdose1 agegroupnew sex2 race4
ethnicity3 firstdose agenew
		decile_sviscore1 decile_sviscore4 mortalityratepct Vaccinationsite
		vaccinedose2 seconddose DWHPatient CurrentStateIMMId
VaccinationDate1
		DWHPatient firstdose seconddose agenew sex2 race4 ethnicity3
Vaccinationsite
		sviscore: VaccinationDate: monthdose1 timebetweendose timeokay
mortalityratepct
		matchedpairs icd_code geoidnew vaccinedose2 daystodeath daystodeathd1
death1yrd1 death1yr
		'vs dod'n nursinghomevaccinationfinal HOMELESSFLAG
		TRANSAX1-TRANSAX15 eld:);
	where SEX2 ne "U" and
	Vaccinationsite not in ("CORRECTIONAL FACILITY","NURSING HOME")
	and nursinghomevaccinationfinal ne 1 AND HOMELESSFLAG=0;
	if seconddose=" " then noseconddose=1;
	else noseconddose=0;
	format agenew age10ygroup.;
	mortalityratepct=round(mortalityratepct,1);
	IF FIRSTDOSE="1. PFIZER" THEN EXPOS=1;
	ELSE EXPOS=0;
```

```
if death1yrd1=1 and substr(icd_code,1,2) in ("I0","I1","I2","I3","I4","I5","I6") then CVD1YRD1=1;
	else if death1yrD1=1 and substr(icd_code,1,3) in ("I71","I72","I73","I74","I75","I76","I77") then CVD1YRD1=1;
	else CVD1YRD1=0;
	if SUBSTR(icd_code,1,3) IN ("X40","X41","X42","X43","X44",
	"X60","X61","X62","X62","X64","X85","Y10","Y11","Y12","Y13","Y14")
	and death1yrD1=1 then overdose1yrD1=1;
	else overdose1yrD1=0;
	if SUBSTR(icd_code,1,3) in ("X85","X86","X87","X88","X89","X90","X91","X92"
			,"X93","X94","X95","X96","X97","X98","X99","Y00","Y01","Y02","Y03"
			,"Y04","Y05","Y06","Y07","Y08","Y09")
	and DEATH1YRD1=1 then homicide1yrD1=1;
	else if substr(icd_code,1,4)="Y871" and death1yrD1=1 then homicide1yrD1=1;
	else homicide1yrD1=0;
	if SUBSTR (ICD_CODE,1,2) IN ("X6","X7") AND death1yrD1=1 THEN SUICIDE1YRD1=1;
	ELSE IF SUBSTR (ICD_CODE,1,3) IN ("X80","X81","X82","X83","X84") AND DEATH1YRD1=1 THEN SUICIDE1YRD1=1;
	ELSE IF substr(icd_code,1,4)="Y870" and death1yr=1 THEN SUICIDE1YR=1;
	ELSE SUICIDE1YRD1=0;
	if SUBSTR (ICD_CODE,1,2) IN ("V0","V1","V2","V3","V4","V5","V6","V7","V8","V9"
	,"W0","W1","W2","W3","W4","W5","W6","W7","W8","W9","X0","X1","X2","X3","X4")
	AND DEATH1YRD1=1 THEN UNINTENTIONAL1YRD1=1;
	ELSE IF SUBSTR(ICD_CODE,1,3) IN ("X50","X51","X52","X53","X54","X55","X56","X57","X58","X59"
			,"Y85","Y86") AND DEATH1YRD1=1 THEN UNINTENTIONAL1YRD1=1;
```

```
ELSE UNINTENTIONAL1YRD1=0;

if SUBSTR (ICD_CODE,1,2) IN ("V0","V1","V2","V3","V4","V5","V6","V7","V8","V9"

,"W2","W3","W4","W5","W6","W7","W8","W9","X0","X1","X2","X3","X4")

AND DEATH1YRD1=1 THEN NOFALLUNINTENTIONAL1YRD1=1;

ELSE IF SUBSTR(ICD_CODE,1,3) IN
("X50","X51","X52","X53","X54","X55","X56","X57","X58","X59"

,"Y85","Y86") AND DEATH1YRD1=1 THEN
NOFALLUNINTENTIONAL1YRD1=1;

ELSE NOFALLUNINTENTIONAL1YRD1=0;

if homicide1yrD1=1 or UNINTENTIONAL1YRD1=1 or SUICIDE1YRD1=1 or
overdose1yrD1=1 then unnatural1yrD1=1;

else unnatural1yrD1=0;

if homicide1yrD1=1 or SUICIDE1YRD1=1 then suicidehomicide1yrD1=1;

else suicidehomicide1yrD1=0;

if homicide1yrD1=1 or NOFALLUNINTENTIONAL1YRD1=1 or SUICIDE1YRD1=1 or
overdose1yrD1=1

then NOFALLunnatural1yrD1=1;

else NOFALLunnatural1yrD1=0;

if SUBSTR(ICD_CODE,1,2) in ("V2","V3","V4","V5","V6","V7") and death1yrD1=1 then
mva1yrD1=1;

else if SUBSTR(ICD_CODE,1,3) in
("V02","V03","V04","V12","V13","V14","V83","V84","V85","V86")

and death1yrD1=1 then mva1yrD1=1;

if icd_code in
("V090","V092","V190","V191","V192","V194","V195","V196","V803","V804",
```

```
	"V805","V810","V811","V820","V821","V870","V871","V872","V873","V874","V875",
	"V876","V877","V878","V881","V882","V883","V884","V885","V886","V887","V888","V890"
,"V892")
	AND DEATH1YRD1=1 THEN MVA1YRD1=1;
	ELSE MVA1YRD1=0;
	array test {*} transax: ; /*** Select all columns with ACME prefix ***/
	count =0;
	do j= 1 to 15;
		if test{j} ne " " then do;
		count + (test{j} in: ("U071"));
		end;
	end;
	if count>0 then COVIDDEATH=1;
	else COVIDDEATH=0;
	if COVIDDEATH=1 and DEATH1YRD1=1 then COVIDDEATH1YRD1=1;
	ELSE COVIDDEATH1YRD1=0;
	if COVIDDEATH=0 and DEATH1YRD1=1 then NONCOVIDDEATH1YRD1=1;
	ELSE NONCOVIDDEATH1YRD1=0;
	counteld=0;
	array new ELD1-ELD6;
		do i =1 to dim(new);
		if new{i} ne . and 'vs dod'n ne . then do;
		counteld+(vaccinationdate1 le new{i} <= 'vs dod'n);
		end;
	end;
	if DEATH1YRD1=1 and (COVIDDEATH1YRD1=1 or counteld >0) then
COVIDDEATHPLUS1YRD1=1;
```

```
ELSE COVIDDEATHPLUS1YRD1=0;

if DEATH1YRD1=1 and (COVIDDEATHPLUS1YRD1=0) then
NONCOVIDDEATHPLUS1YRD1=1;
ELSE NONCOVIDDEATHPLUS1YRD1=0;

if counteld>0 then do;
    if eld6 ne . and 'vs dod'n >=eld6 then covinftodeathdys='vs dod'n-eld6;
    else if eld5 ne . and 'vs dod'n >=eld5 then covinftodeathdys='vs dod'n-eld5;
    else if eld4 ne . and 'vs dod'n >=eld4 then covinftodeathdys='vs dod'n-eld4;
    else if eld3 ne . and 'vs dod'n >=eld3 then covinftodeathdys='vs dod'n-eld3;
    else if eld2 ne . and 'vs dod'n >=eld2 then covinftodeathdys='vs dod'n-eld2;

    else if eld1 ne . and 'vs dod'n >=eld1 then covinftodeathdys='vs dod'n-eld1;
end;
if daystodeathd1=. then daystodeathd1=366;
countcovinf=0;
countcovinfbf20=0;

array covinf ELD1-ELD6;
    do k =1 to dim(covinf);
    if covinf{k} ne . then do;
    countcovinf+(covinf{k} le vaccinationdate1);
    countcovinfbf20+(covinf{k} le "20dec2020"d);
    end;
end;
drop i j k;
```

```
run;

proc freq data=work.dose1matcheddatafinal;
	table countcovinf:;
run;

proc means data=work.dose1matcheddatafinal;
	var daystodeath daystodeathd1;
run;

proc freq data=work.dose1matcheddatafinal;
	table counteld*death1yrd1;
	table covinftodeathdys;
run;

proc print data=work.dose1matcheddatafinal (obs= 100);
	where COVIDDEATHPLUS1YRD1=1 and covinftodeathdys<0;
	var DWHPatient VaccinationDate: eld: counteld 'vs dod'n covinftodeathdys;
run;

proc freq data=dose1matcheddatafinal;
	tABLE daystodeath;
RUN;

proc freq data=dose1matcheddatafinal;
	tABLE death1yrd1 CVD: covid: NONCOVID: suicidehomicide1yrD1;
```

```
RUN;

/*** For Table 1 ***/

title;
proc freq data=dose1matcheddatafinal;
	table ageatdose1 sex2 race4 ethnicity3 firstdose seconddose noseconddose
	vaccinedose2/missing;
	format ageatdose1 age10ygroup.;
run;

title;
proc freq data=dose1matcheddatafinal;
	table ageatdose1 sex2 race4 ethnicity3 firstdose seconddose noseconddose
	vaccinedose2/missing;
	format ageatdose1 age10ygroup.;
	where matchedpairs=1;
run;

proc freq data=dose1matcheddatafinal;
	table Vaccinationsite VaccinationDate1;
	format VaccinationDate1 month7.;
run;
```

```
proc freq data=dose1matcheddatafinal;
	table Vaccinationsite VaccinationDate1;
	format VaccinationDate1 month7.;
	where matchedpairs=1;
run;

/**** For Sup Figure bar graph ***/

proc freq data=work.dose1matcheddatafinal;
	table firstdose*ageatdose1*VaccinationDate1/crosslist;
	format ageatdose1 age10ygroup. VaccinationDate1 monyy.;
run;

/*** DESCRIPTIVES BY EXPOSURE ***/

proc freq data=dose1matcheddatafinal;
	table FIRSTDOSE*(ageatdose1 sex2 race4 ethnicity3 seconddose noseconddose
vaccinedose2)/missing CROSSLIST;
	format ageatdose1 age10ygroup.;
run;

proc freq data=dose1matcheddatafinal;
	table firstdose*(Vaccinationsite VaccinationDate1)/missing CROSSLIST;
	format VaccinationDate1 month7.;
run;
```

```
proc freq data=dose1matcheddatafinal;
	table FIRSTDOSE*(ageatdose1 sex2 race4 ethnicity3 seconddose noseconddose vaccinedose2)/missing CROSSLIST;
	format ageatdose1 age10ygroup.;
	where matchedpairs=1;
run;

proc freq data=dose1matcheddatafinal;
	table firstdose*(Vaccinationsite VaccinationDate1)/missing CROSSLIST;
	format VaccinationDate1 month7.;
	where matchedpairs=1;
run;

proc freq data=dose1matcheddatafinal;
	tABLE death1yrd1 CVD: covid: NONCOVID: suicidehomicide1yrD1;
RUN;

proc freq data=dose1matcheddatafinal;
	TABLE death1yrd1 CVD: covid: NONCOVID: suicidehomicide1yrD1;
	where matchedpairs=1;
RUN;

proc freq data=dose1matcheddatafinal;
	tABLE (death1yrd1 CVD: covid: NONCOVID: suicidehomicide1yrD1)*ageatdose1/crosslist;
	format ageatdose1 age10ygroup.;
RUN;
```

```
proc freq data=dose1matcheddatafinal;
	tABLE (death1yrd1 CVD: covid: NONCOVID:
suicidehomicide1yrD1)*ageatdose1/crosslist;
	format ageatdose1 age10ygroup.;
	where matchedpairs=1;
RUN;

proc means data=work.dose1matcheddatafinal mean std median q1 q3;
	var ageatdose1 sviscore:;
	format ageatdose1 age10ygroup.;
run;

proc means data=work.dose1matcheddatafinal mean std median q1 q3 maxdec=1;
	var ageatdose1 sviscore:;
	format ageatdose1 age10ygroup.;
	class firstdose;
run;

proc means data=work.dose1matcheddatafinal mean std median q1 q3 maxdec=1;
	var ageatdose1 sviscore:;
	format ageatdose1 age10ygroup.;
	where matchedpairs=1;
run;
```

```
proc means data=work.dose1matcheddatafinal mean std median q1 q3 maxdec=1;
	var ageatdose1 sviscore:;
	format ageatdose1 age10ygroup.;
	class firstdose;
	where matchedpairs=1;
run;

proc format;
	value deaths
	1="1. Yes"
	0="2. No";
run;

proc template;
	edit Base.Freq.RiskDiff;
		define risk;
		format=12.8;
		end;
		define LowerCL;
		format=12.8;
		end;
		define UpperCL;
		format=12.8;
```

```
                    end;
            end;
run;

ods output RelativeRisks=RRMatched;
ods output RiskDiffCol1=RDmatched;
proc freq data=dose1matcheddatafinal order=formatted;
            table firstdose*(death1yrd1 CVD: covid: NONCOVID: suicidehomicide1yrD1)
            /relrisk riskdiff;
            where matchedpairs=1;
            format death1yrd1 CVD: covid: NONCOVID: suicidehomicide1yrD1 deaths.;
run;
ods output close;

ods output RelativeRisks=RRall;
ods output RiskDiffCol1=RDall;
proc freq data=dose1matcheddatafinal order=formatted;
            table firstdose*(death1yrd1 CVD: covid: NONCOVID: suicidehomicide1yrD1)
            /relrisk riskdiff;
            format death1yrd1 CVD: covid: NONCOVID: suicidehomicide1yrD1 deaths.;
run;
ods output close;

proc print data=rrall;
run;
```

```
proc print data=rdall;
run;

proc print data=rrmatched;
run;

proc print data=rdmatched;
run;

/*** All cause deaths ***/
proc sort data=dose1matcheddatafinal;
	by firstdose;
run;

ods graphics on;
proc lifetest data=dose1matcheddatafinal outsurv=allcause_Surv_matched method=km
	timelist=1 to 365 by 1 plots=none STDERR;
	time daystodeathd1*death1yrd1(0);
	strata firstdose;
	where matchedpairs=1;
run;

proc options option=config;
run;
```

```
data allcause_Surv_matched;
	set allcause_Surv_matched;
	risk=100*(1-survival);
	riskUCL=100*(1-SDF_LCL);
	riskLCL=100*(1-SDF_UCL);
run;

proc sql;
	select firstdose,max(risk) as risk,
	100-calculated risk as surv,
	calculated risk*calculated surv as var
	from allcause_Surv_matched
	group by firstdose;
quit;

ods graphics on;
proc lifetest data=dose1matcheddatafinal plots=all outsurv=allcause_Surv_full
method=km
	timelist=1 to 365 by 1 plots=none;
	time daystodeathd1*death1yrd1(0);
	strata firstdose;
run;

proc options option=config;
run;
```

```
data allcause_Surv_full;
	set allcause_Surv_full;
	risk=100*(1-survival);
	riskUCL=100*(1-SDF_LCL);
	riskLCL=100*(1-SDF_UCL);
run;

data allcause;
	set allcause_Surv_matched (in=a) allcause_Surv_full (in=b);
	if a=1 then matched="Matched observations";
	else matched="All observations";
	outcome="1. All cause";
	if firstdose="1. PFIZER" then vaccinegroup2="BNT162b2";
	else if firstdose="2. MODERN" then vaccinegroup2="mRNA-1273";
run;

proc sort data=allcause nodupkey;
	by vaccinegroup2 matched daystodeathd1 _CENSOR_;
run;

proc print data=allcause;
	where daystodeath>=365;
run;

proc sql;
	select ceil(10*max(riskUCL))/10 into :risk
```

```
    from work.allcause;
run;

ods graphics on/width=9in height=7in;
proc sgpanel data=allcause;
    title "All-cause Deaths" bold font='arial';
    panelby matched  /novarname headerbackcolor=white spacing=5
    HEADERATTRS=(Color=black Family=Arial Size=10 Weight=Bold) noheaderborder;
    series x= daystodeathd1 y=risk/group=vaccinegroup2 LINEATTRS=(pattern=1
thickness=2)
    legendlabel="Estimate" name="plot1";
    band x=daystodeathd1 upper=riskUCL lower=riskLCL/group=vaccinegroup2
TRANSPARENCY=0.7 NOOUTLINE
    legendlabel="95% CI" NAME="Confidence Interval";
    rowaxis max=&risk label="Cumulative risk (%)"     VALUEATTRS=(Color=black
Family=Arial Size=10)
    labelattrs=(Color=black Family=Arial Size=10 weight=bold);
    colaxis max=365 label="Time (days) from second dose of Vaccine"
    VALUEATTRS=(Color=black Family=Arial Size=10) labelattrs=(Color=black
Family=Arial Size=10 weight=bold);
    keylegend "plot1"/ title=" ";
    keylegend "Confidence Interval"/title="95% CI";
run;
ods graphics off;

ods graphics on/width=4in height=3in;
```

```
proc sgplot data=allcause;
	title "All-cause Deaths" bold font='arial';
	step x= daystodeathd1 y=risk/group=vaccinegroup2 LINEATTRS=(pattern=1
thickness=2)
	legendlabel="Estimate" name="plot1";
	band x=daystodeathd1 upper=riskUCL lower=riskLCL/group=vaccinegroup2
TRANSPARENCY=0.7 NOOUTLINE
	legendlabel="95% CI" NAME="Confidence Interval";
	yaxis max=&risk label="Cumulative risk (%)"	VALUEATTRS=(Color=black
Family=Arial Size=10)
	labelattrs=(Color=black Family=Arial Size=10 weight=bold);
	xaxis max=365 label="Time (days) from second dose of Vaccine"
	VALUEATTRS=(Color=black Family=Arial Size=10) labelattrs=(Color=black
Family=Arial Size=10 weight=bold);
	keylegend "plot1"/ title=" " position=bottom;
	keylegend "Confidence Interval"/title="95% CI" position=bottom;
	where matched="All observations";
run;
ods graphics off;

proc sql;
	select ceil(10*max(riskUCL))/10 into :risk
	from work.allcause
	where matched="Matched observations";
run;
```

```
ods graphics on/width=4in height=3in;

proc sgplot data=allcause;

    title "All-cause Deaths" bold font='arial';

    step x= daystodeathd1 y=risk/group=vaccinegroup2 LINEATTRS=(pattern=1
thickness=2)

    legendlabel="Estimate" name="plot1";

    band x=daystodeathd1 upper=riskUCL lower=riskLCL/group=vaccinegroup2
TRANSPARENCY=0.7 NOOUTLINE

    legendlabel="95% CI" NAME="Confidence Interval";

    yaxis max=&risk label="Cumulative risk (%)"      VALUEATTRS=(Color=black
Family=Arial Size=10)

    labelattrs=(Color=black Family=Arial Size=10 weight=bold);

    xaxis max=365 label="Time (days) from second dose of Vaccine"

    VALUEATTRS=(Color=black Family=Arial Size=10) labelattrs=(Color=black
Family=Arial Size=10 weight=bold);

    keylegend "plot1"/ title=" " position=bottom;

    keylegend "Confidence Interval"/title="95% CI" position=bottom;

    where matched="Matched observations";

run;

ods graphics off;

proc sql;

    select ceil(10*max(riskUCL))/10 into :risk

    from work.allcause

    where daystodeathd1 le 100;

run;
```

```
/*** Graph only uptil 100 days ***/

ods html on;

ods listing;

ods graphics on/width=9in height=7in;

proc sgpanel data=allcause;

	title "All-cause Deaths" bold font='arial';

	panelby matched  /novarname headerbackcolor=white spacing=5

	HEADERATTRS=(Color=black Family=Arial Size=10 Weight=Bold) noheaderborder;

	series x= daystodeath y=risk/group=vaccinegroup2 LINEATTRS=(pattern=1
thickness=2)

	legendlabel="Estimate" name="plot1";

	band x=daystodeath upper=riskUCL lower=riskLCL/group=vaccinegroup2
TRANSPARENCY=0.7 NOOUTLINE

	legendlabel="95% CI" NAME="Confidence Interval";

	rowaxis max=&risk label="Cumulative risk (%)"     VALUEATTRS=(Color=black
Family=Arial Size=10)

	labelattrs=(Color=black Family=Arial Size=10 weight=bold);

	colaxis max=100 label="Time (days) from second dose of Vaccine"

	VALUEATTRS=(Color=black Family=Arial Size=10) labelattrs=(Color=black
Family=Arial Size=10 weight=bold);

	keylegend "plot1"/ title=" ";

	keylegend "Confidence Interval"/title="95% CI";

run;

ods graphics off;

title;
```

```
data store.allcause;
	set work.allcause;
run;

/*** CVD Deaths ***/

ods graphics on;
proc lifetest data=work.dose1matcheddatafinal plots=all outsurv=CVD_Surv_matched
method=km
	timelist=1 to 365 by 1 plots=none notable;
	time daystodeathd1*cvd1yrd1(0);
	where matchedpairs=1;
	strata firstdose;
run;

data CVD_Surv_matched;
	set CVD_Surv_matched;
	risk=100*(1-survival);
	riskUCL=100*(1-SDF_LCL);
	riskLCL=100*(1-SDF_UCL);
run;

ods graphics on;
proc lifetest data=dose1matcheddatafinal plots=all outsurv=CVD_Surv_full method=km
	timelist=1 to 365 by 1 plots=none notable;
	time daystodeathd1*cvd1yrd1(0);
```

```
    strata firstdose;
run;

data CVD_Surv_full;
    set CVD_Surv_full;
    risk=100*(1-survival);
    riskUCL=100*(1-SDF_LCL);
    riskLCL=100*(1-SDF_UCL);
run;

data cvd;
    set CVD_Surv_matched (in=a) CVD_Surv_full (in=b);
    if a=1 then matched="Matched observations";
    else matched="All observations";
    length outcome $30.;
    outcome="2. Cardiovascular deaths";
    if Firstdose="1. PFIZER" then vaccinegroup2="BNT162b2";
    else if Firstdose="2. MODERN" then vaccinegroup2="mRNA-1273";
run;

proc sort data=cvd nodupkey;
    by vaccinegroup2 matched daystodeathd1 _CENSOR_;
run;

proc print data=CVD;
    where daystodeathd1>=365;
```

```
run;

proc sql;
	select ceil(100*max(riskUCL))/100 into :risk
	from work.cvd;
run;

ods html on;
ods listing;
ods graphics on/width=9in height=7in;

proc sgpanel data=cvd;
	title "CVD deaths" bold font='arial';
	panelby matched  /novarname headerbackcolor=white spacing=5
	HEADERATTRS=(Color=black Family=Arial Size=10 Weight=Bold) noheaderborder;
	series x= daystodeathd1 y=risk/group=vaccinegroup2 LINEATTRS=(pattern=1
thickness=2)
	legendlabel="Estimate" name="plot1";
	band x=daystodeathd1 upper=riskUCL lower=riskLCL/group=vaccinegroup2
TRANSPARENCY=0.7 NOOUTLINE
	legendlabel="95% CI" NAME="Confidence Interval";
	rowaxis max=&risk label="Cumulative risk (%)"     VALUEATTRS=(Color=black
Family=Arial Size=10)
	labelattrs=(Color=black Family=Arial Size=10 weight=bold);
	colaxis max=365 label="Time (days) from second dose of Vaccine"
	VALUEATTRS=(Color=black Family=Arial Size=10) labelattrs=(Color=black
Family=Arial Size=10 weight=bold);
```

```
    keylegend "plot1"/ title=" ";

    keylegend "Confidence Interval"/title="95% CI";

run;

ods graphics off;

title;

ods graphics on/width=4in height=3in;

proc sgplot data=cvd;

    title "Cardiovascular Disease Deaths" bold font='arial';

    step x= daystodeathd1 y=risk/group=vaccinegroup2 LINEATTRS=(pattern=1
thickness=2)

    legendlabel="Estimate" name="plot1";

    band x=daystodeathd1 upper=riskUCL lower=riskLCL/group=vaccinegroup2
TRANSPARENCY=0.7 NOOUTLINE

    legendlabel="95% CI" NAME="Confidence Interval";

    yaxis max=&risk label="Cumulative risk (%)"     VALUEATTRS=(Color=black
Family=Arial Size=10)

    labelattrs=(Color=black Family=Arial Size=10 weight=bold);

    xaxis max=365 label="Time (days) from second dose of Vaccine"

    VALUEATTRS=(Color=black Family=Arial Size=10) labelattrs=(Color=black
Family=Arial Size=10 weight=bold);

    keylegend "plot1"/ title=" " position=bottom;

    keylegend "Confidence Interval"/title="95% CI" position=bottom;

    where matched="All observations";

run;

ods graphics off;
```

```
proc sql;
	select ceil(10*max(riskUCL))/10 into :risk
	from work.cvd
	where matched="Matched observations";
quit;

ods graphics on/width=4in height=3in;

proc sgplot data=CVD;
	title "Cardiovascular Deaths" bold font='arial';
	step x= daystodeathd1 y=risk/group=vaccinegroup2 LINEATTRS=(pattern=1 thickness=2)
	legendlabel="Estimate" name="plot1";
	band x=daystodeathd1 upper=riskUCL lower=riskLCL/group=vaccinegroup2 TRANSPARENCY=0.7 NOOUTLINE
	legendlabel="95% CI" NAME="Confidence Interval";
	yaxis max=&risk label="Cumulative risk (%)"	VALUEATTRS=(Color=black Family=Arial Size=10)
	labelattrs=(Color=black Family=Arial Size=10 weight=bold);
	xaxis max=365 label="Time (days) from second dose of Vaccine"
	VALUEATTRS=(Color=black Family=Arial Size=10) labelattrs=(Color=black Family=Arial Size=10 weight=bold);
	keylegend "plot1"/ title=" " position=bottom;
	keylegend "Confidence Interval"/title="95% CI" position=bottom;
	where matched="Matched observations";
```

```
run;

ods graphics off;

proc sql;

	select ceil(100*max(riskUCL))/100 into :risk

	from work.CVD

	where daystodeath le 100;

run;

ods html on;

ods listing;

ods graphics on/width=9in height=7in;

proc sgpanel data=cvd;

	title "CVD deaths" bold font='arial';

	panelby matched  /novarname headerbackcolor=white spacing=5

	HEADERATTRS=(Color=black Family=Arial Size=10 Weight=Bold) noheaderborder;

	series x= daystodeath y=risk/group=vaccinegroup2 LINEATTRS=(pattern=1
thickness=2)

	legendlabel="Estimate" name="plot1";

	band x=daystodeath upper=riskUCL lower=riskLCL/group=vaccinegroup2
TRANSPARENCY=0.7 NOOUTLINE

	legendlabel="95% CI" NAME="Confidence Interval";

	rowaxis max=&risk label="Cumulative risk (%)"     VALUEATTRS=(Color=black
Family=Arial Size=10)

	labelattrs=(Color=black Family=Arial Size=10 weight=bold);

	colaxis max=100 label="Time (days) from second dose of Vaccine"
```

```
	VALUEATTRS=(Color=black Family=Arial Size=10) labelattrs=(Color=black
Family=Arial Size=10 weight=bold);
	keylegend "plot1"/ title=" ";
	keylegend "Confidence Interval"/title="95% CI";
run;
ods graphics off;
title;

data store.cvd;
	set cvd;
run;

/*** COVID ***/

%let data=DOSE1MATCHEDDATAFINAL;
run;

ods graphics on;
proc lifetest data=&data outsurv=COV_Surv_matched method=km
	timelist=1 to 365 by 1 plots=none notable;
	time daystodeathd1*coviddeath1yrd1(0);
	where matchedpairs=1;
	strata firstdose;
run;

data COV_Surv_matched;
```

```
	set COV_Surv_matched;
	risk=100*(1-survival);
	riskUCL=100*(1-SDF_LCL);
	riskLCL=100*(1-SDF_UCL);
run;

ods graphics on;
proc lifetest data=&data outsurv=COV_Surv_full method=km
	timelist=1 to 365 by 1 plots=none notable;
	time daystodeathd1*coviddeath1yrd1(0);
	strata firstdose;
run;

data COV_Surv_full;
	set COV_Surv_full;
	risk=100*(1-survival);
	riskUCL=100*(1-SDF_LCL);
	riskLCL=100*(1-SDF_UCL);
run;

data COVID;
	set COV_Surv_matched (in=a) COV_Surv_full (in=b);
	if a=1 then matched="Matched observations";
	else matched="All observations";
	length outcome $30.;
```

```
        outcome="3. COVID-19 deaths";
        if firstdose="1. PFIZER" then vaccinegroup2="BNT162b2";
        else if firstdose="2. MODERN" then vaccinegroup2="mRNA-1273";
run;

proc sort data=COVID nodupkey;
        by vaccinegroup2 matched daystodeathd1 _CENSOR_;
run;

proc sql;
        select ceil(100*max(riskUCL))/100 into :risk
        from work.covid;
run;

ods html on;
ods listing;
ods graphics on/width=9in height=7in;

proc sgpanel data=COVID;
        title "COVID deaths" bold font='arial';
        panelby matched  /novarname headerbackcolor=white spacing=5
        HEADERATTRS=(Color=black Family=Arial Size=10 Weight=Bold) noheaderborder;
        series x= daystodeathd1 y=risk/group=vaccinegroup2 LINEATTRS=(pattern=1
thickness=2)
        legendlabel="Estimate" name="plot1";
        band x=daystodeathd1 upper=riskUCL lower=riskLCL/group=vaccinegroup2
TRANSPARENCY=0.7 NOOUTLINE
```

```
	legendlabel="95% CI" NAME="Confidence Interval";
	rowaxis max=&risk label="Cumulative risk (%)"     VALUEATTRS=(Color=black
Family=Arial Size=10)
	labelattrs=(Color=black Family=Arial Size=10 weight=bold);
	colaxis max=365 label="Time (days) from second dose of Vaccine"
	VALUEATTRS=(Color=black Family=Arial Size=10) labelattrs=(Color=black
Family=Arial Size=10 weight=bold);
	keylegend "plot1"/ title=" ";
	keylegend "Confidence Interval"/title="95% CI";
run;
ods graphics off;
title;

ods graphics on/width=4in height=3in;
proc sgplot data=COVID;
	title "COVID-19 Deaths" bold font='arial';
	step x= daystodeathd1 y=risk/group=vaccinegroup2 LINEATTRS=(pattern=1
thickness=2)
	legendlabel="Estimate" name="plot1";
	band x=daystodeathd1 upper=riskUCL lower=riskLCL/group=vaccinegroup2
TRANSPARENCY=0.7 NOOUTLINE
	legendlabel="95% CI" NAME="Confidence Interval";
	yaxis max=&risk label="Cumulative risk (%)"        VALUEATTRS=(Color=black
Family=Arial Size=10)
	labelattrs=(Color=black Family=Arial Size=10 weight=bold);
	xaxis max=365 label="Time (days) from second dose of Vaccine"
	VALUEATTRS=(Color=black Family=Arial Size=10) labelattrs=(Color=black
Family=Arial Size=10 weight=bold);
```

```
keylegend "plot1"/ title=" " position=bottom;
keylegend "Confidence Interval"/title="95% CI" position=bottom;
where matched="All observations";
run;
ods graphics off;

proc sql;
select ceil(100*max(riskUCL))/100 into :risk
from work.covid
where matched="Matched observations";
run;

ods graphics on/width=4in height=3in;

proc sgplot data=Covid;
title "COVID-19 Deaths" bold font='arial';
step x= daystodeathd1 y=risk/group=vaccinegroup2 LINEATTRS=(pattern=1
thickness=2)
legendlabel="Estimate" name="plot1";
band x=daystodeathd1 upper=riskUCL lower=riskLCL/group=vaccinegroup2
TRANSPARENCY=0.7 NOOUTLINE
legendlabel="95% CI" NAME="Confidence Interval";
yaxis max=&risk label="Cumulative risk (%)"     VALUEATTRS=(Color=black
Family=Arial Size=10)
labelattrs=(Color=black Family=Arial Size=10 weight=bold);
xaxis max=365 label="Time (days) from second dose of Vaccine"
```

```
	VALUEATTRS=(Color=black Family=Arial Size=10) labelattrs=(Color=black
Family=Arial Size=10 weight=bold);
	keylegend "plot1"/ title=" " position=bottom;
	keylegend "Confidence Interval"/title="95% CI" position=bottom;
	where matched="Matched observations";
run;
ods graphics off;

proc sql;
	select ceil(1000*max(riskUCL))/1000 into :risk
	from work.covid
	where daystodeath le 100;
run;

ods html on;
ods listing;
ods graphics on/width=9in height=7in;

proc sgpanel data=COVID;
	title "COVID deaths" bold font='arial';
	panelby matched  /novarname headerbackcolor=white spacing=5
	HEADERATTRS=(Color=black Family=Arial Size=10 Weight=Bold) noheaderborder;
	series x= daystodeath y=risk/group=vaccinegroup2 LINEATTRS=(pattern=1
thickness=2)
	legendlabel="Estimate" name="plot1";
```

```
	band x=daystodeath upper=riskUCL lower=riskLCL/group=vaccinegroup2 
TRANSPARENCY=0.7 NOOUTLINE

	legendlabel="95% CI" NAME="Confidence Interval";

	rowaxis max=&risk label="Cumulative risk (%)"     VALUEATTRS=(Color=black 
Family=Arial Size=10)

	labelattrs=(Color=black Family=Arial Size=10 weight=bold);

	colaxis max=100 label="Time (days) from second dose of Vaccine"

	VALUEATTRS=(Color=black Family=Arial Size=10) labelattrs=(Color=black 
Family=Arial Size=10 weight=bold);

	keylegend "plot1"/ title=" ";

	keylegend "Confidence Interval"/title="95% CI";

run;

ods graphics off;

data store.covid;

	set covid;

run;

/*** non-COVID ***/

ods graphics on;

proc lifetest data=&data plots=none outsurv=noCOV_Surv_matched method=km

	timelist=1 to 365 by 1 plots=none notable;

	time daystodeathd1*noncoviddeath1yrd1(0);

	where matchedpairs=1;

	strata Firstdose;
```

```
run;

data noCOV_Surv_matched;
	set noCOV_Surv_matched;
	risk=100*(1-survival);
	riskUCL=100*(1-SDF_LCL);
	riskLCL=100*(1-SDF_UCL);
run;

ods graphics on;
proc lifetest data=&data plots=all outsurv=noCOV_Surv_full method=km
	timelist=1 to 365 by 1 plots=none notable;
	time daystodeathd1*noncoviddeath1yrd1(0);
	strata firstdose;
run;

data noCOV_Surv_full;
	set noCOV_Surv_full;
	risk=100*(1-survival);
	riskUCL=100*(1-SDF_LCL);
	riskLCL=100*(1-SDF_UCL);
run;

data nonCOVID;
	set noCOV_Surv_matched (in=a) noCOV_Surv_full (in=b);
```

```
    if a=1 then matched="Matched observations";
    else matched="All observations";
    length outcome $30.;
    outcome="NON-COVID-19 deaths";
    if firstdose="1. PFIZER" then vaccinegroup2="BNT162b2";
    else if firstdose="2. MODERN" then vaccinegroup2="mRNA-1273";
run;

proc sort data=nonCOVID nodupkey;
    by vaccinegroup2 matched daystodeathd1 _CENSOR_;
run;

proc sql;
    select ceil(100*max(riskUCL))/100 into :risk
    from work.noncovid;
run;

ods html on;
ods listing;
ods graphics on/width=9in height=7in;
proc sgpanel data=nonCOVID;
    title "Non-COVID deaths" bold font='arial';
    panelby matched  /novarname headerbackcolor=white spacing=5
    HEADERATTRS=(Color=black Family=Arial Size=10 Weight=Bold)  noheaderborder;
```

```
	series x= daystodeathd1 y=risk/group=vaccinegroup2 LINEATTRS=(pattern=1
thickness=2)
	legendlabel="Estimate" name="plot1";
	band x=daystodeathd1 upper=riskUCL lower=riskLCL/group=vaccinegroup2
TRANSPARENCY=0.7 NOOUTLINE
	legendlabel="95% CI" NAME="Confidence Interval";
	rowaxis max=&risk label="Cumulative risk (%)"	VALUEATTRS=(Color=black
Family=Arial Size=10)
	labelattrs=(Color=black Family=Arial Size=10 weight=bold);
	colaxis max=365 label="Time (days) from second dose of Vaccine"
	VALUEATTRS=(Color=black Family=Arial Size=10) labelattrs=(Color=black
Family=Arial Size=10 weight=bold);
	keylegend "plot1"/ title=" ";
	keylegend "Confidence Interval"/title="95% CI";
run;
ods graphics off;

ods graphics on/width=4in height=3in;
proc sgplot data=nonCOVID;
	title "Non-COVID-19 Deaths" bold font='arial';
	step x= daystodeathd1 y=risk/group=vaccinegroup2 LINEATTRS=(pattern=1
thickness=2)
	legendlabel="Estimate" name="plot1";
	band x=daystodeathd1 upper=riskUCL lower=riskLCL/group=vaccinegroup2
TRANSPARENCY=0.7 NOOUTLINE
	legendlabel="95% CI" NAME="Confidence Interval";
	yaxis max=&risk label="Cumulative risk (%)"	VALUEATTRS=(Color=black
Family=Arial Size=10)
```

```
        labelattrs=(Color=black Family=Arial Size=10 weight=bold);
        xaxis max=365 label="Time (days) from second dose of Vaccine"
        VALUEATTRS=(Color=black Family=Arial Size=10) labelattrs=(Color=black
Family=Arial Size=10 weight=bold);
        keylegend "plot1"/ title=" " position=bottom;
        keylegend "Confidence Interval"/title="95% CI" position=bottom;
        where matched="All observations";
run;
ods graphics off;

proc sql;
        select ceil(100*max(riskUCL))/100 into :risk
        from work.noncovid
        where matched="Matched observations";
run;

ods graphics on/width=4in height=3in;

proc sgplot data=nonCovid;
        title "Non-COVID-19 Deaths" bold font='arial';
        step x= daystodeathd1 y=risk/group=vaccinegroup2 LINEATTRS=(pattern=1
thickness=2)
        legendlabel="Estimate" name="plot1";
        band x=daystodeathd1 upper=riskUCL lower=riskLCL/group=vaccinegroup2
TRANSPARENCY=0.7 NOOUTLINE
        legendlabel="95% CI" NAME="Confidence Interval";
```

```
    yaxis max=&risk label="Cumulative risk (%)"     VALUEATTRS=(Color=black
Family=Arial Size=10)
    labelattrs=(Color=black Family=Arial Size=10 weight=bold);
    xaxis max=365 label="Time (days) from second dose of Vaccine"
    VALUEATTRS=(Color=black Family=Arial Size=10) labelattrs=(Color=black
Family=Arial Size=10 weight=bold);
    keylegend "plot1"/ title=" " position=bottom;
    keylegend "Confidence Interval"/title="95% CI" position=bottom;
    where matched="Matched observations";
run;
ods graphics off;

proc sql;
    select ceil(100*max(RIsk))/100 into :risk
    from work.noncovid
    where daystodeath le 100;
run;

ods html on;
ods listing;
ods graphics on/width=9in height=7in;
proc sgpanel data=nonCOVID;
    title "Non-COVID deaths" bold font='arial';
    panelby matched  /novarname headerbackcolor=white spacing=5
    HEADERATTRS=(Color=black Family=Arial Size=10 Weight=Bold)  noheaderborder;
```

```
	series x= daystodeath y=risk/group=vaccinegroup2 LINEATTRS=(pattern=1 thickness=2)

	legendlabel="Estimate" name="plot1";

	band x=daystodeath upper=riskUCL lower=riskLCL/group=vaccinegroup2 TRANSPARENCY=0.7 NOOUTLINE

	legendlabel="95% CI" NAME="Confidence Interval";

	rowaxis max=&risk label="Cumulative risk (%)"     VALUEATTRS=(Color=black Family=Arial Size=10)

	labelattrs=(Color=black Family=Arial Size=10 weight=bold);

	colaxis max=100 label="Time (days) from second dose of Vaccine"

	VALUEATTRS=(Color=black Family=Arial Size=10) labelattrs=(Color=black Family=Arial Size=10 weight=bold);

	keylegend "plot1"/ title=" ";

	keylegend "Confidence Interval"/title="95% CI";

run;

ods graphics off;

title;

data store.noncovid;

	set noncovid;

run;

/*** Suicide or homicide ***/

ods graphics on;

proc lifetest data=&data plots=none outsurv=SUHOM_Surv_matched method=km
```

```
        timelist=1 to 365 by 1 plots=none notable;
        time daystodeathd1*suicidehomicide1yrd1(0);
        where matchedpairs=1;
        strata firstdose;
run;

data SUHOM_Surv_matched;
        set SUHOM_Surv_matched;
        risk=100*(1-survival);
        riskUCL=100*(1-SDF_LCL);
        riskLCL=100*(1-SDF_UCL);
run;

ods graphics on;
proc lifetest data=&data plots=none outsurv=SUHOM_Surv_full method=km
        timelist=1 to 365 by 1 plots=none notable;
        time daystodeathd1*suicidehomicide1yrd1(0);
        strata firstdose;
run;

data SUHOM_Surv_full;
        set SUHOM_Surv_full;
        risk=100*(1-survival);
        riskUCL=100*(1-SDF_LCL);
        riskLCL=100*(1-SDF_UCL);
```

```
run;

data SUHOM;
	set SUHOM_Surv_matched (in=a) SUHOM_Surv_full (in=b);
	if a=1 then matched="Matched cohort";
	else matched="Full cohort";
	length outcome $30.;
	outcome="Suicide or Homicide deaths";
	if firstdose="1. PFIZER" then vaccinegroup2="BNT162b2";
	else if firstdose="2. MODERN" then vaccinegroup2="mRNA-1273";
run;

proc sort data=suhom nodupkey;
	by vaccinegroup2 matched daystodeathd1 _CENSOR_;
run;

proc print data=suhom;
	where daystodeath ge 360 and riskLCL ne .;
run;

proc sql;
	select ceil(100*max(riskUCL))/100 into :risk
	from work.suhom;
quit;

ods html on;
```

```
ods listing;

ods graphics on/width=9in height=7in;

proc sgpanel data=suhom ;

    title "Suicide or Homicide deaths" bold font='arial';

    panelby matched  /novarname headerbackcolor=white spacing=5

    HEADERATTRS=(Color=black Family=Arial Size=10 Weight=Bold)  noheaderborder
sort=descending;

    step x= daystodeathd1 y=risk/group=vaccinegroup2 LINEATTRS=(pattern=1
thickness=2)

    legendlabel="Estimate" name="plot1";

    band x=daystodeathd1 upper=riskUCL lower=riskLCL/group=vaccinegroup2
TRANSPARENCY=0.7 NOOUTLINE

    legendlabel="95% CI" NAME="Confidence Interval";

    rowaxis max=&risk label="Cumulative risk (%)"     VALUEATTRS=(Color=black
Family=Arial Size=10)

    labelattrs=(Color=black Family=Arial Size=10 weight=bold);

    colaxis max=365 label="Time (days) from second dose of Vaccine"

    VALUEATTRS=(Color=black Family=Arial Size=10) labelattrs=(Color=black
Family=Arial Size=10 weight=bold);

    keylegend "plot1"/ title=" ";

    keylegend "Confidence Interval"/title="95% CI";

run;

ods graphics off;

title;
```

```
ods html on;

ods listing;

ods graphics on/width=4in height=5in;

proc sgplot data=suhom;

	title "Suicide or Homicide Deaths (Full cohort)" bold font='arial';

	step x= daystodeathd1 y=risk/group=vaccinegroup2 LINEATTRS=(pattern=1
thickness=2)

	legendlabel="Estimate" name="plot1";

	band x=daystodeathd1 upper=riskUCL lower=riskLCL/group=vaccinegroup2
TRANSPARENCY=0.7 NOOUTLINE

	legendlabel="95% CI" NAME="Confidence Interval";

	yaxis max=&risk label="Cumulative risk (%)"	VALUEATTRS=(Color=black
Family=Arial Size=10)

	labelattrs=(Color=black Family=Arial Size=10 weight=bold);

	xaxis max=365 label="Time (days) from second dose of Vaccine"

	VALUEATTRS=(Color=black Family=Arial Size=10) labelattrs=(Color=black
Family=Arial Size=10 weight=bold);

	keylegend "plot1"/ title=" " position=bottom;

	keylegend "Confidence Interval"/title="95% CI" position=bottom;

	where matched="Full cohort";

run;

ods graphics off;

title;

proc sql;

	select ceil(100*max(riskUCL))/100 into :risk
```

```
        from work.suhom
        where matched="Matched cohort";
run;

proc print data=suhom;
        where matched="Matched cohort" and daystodeath ge 360;
run;

ods html on;
ods listing;
ods graphics on/width=8in height=5.5in;
proc sgplot data=suhom;
        title "Suicide or Homicide Deaths" bold font='arial';
        series x= daystodeathd1 y=risk/group=vaccinegroup2 LINEATTRS=(pattern=1
thickness=2)
        legendlabel="Estimate" name="plot1";
        band x=daystodeathd1 upper=riskUCL lower=riskLCL/group=vaccinegroup2
TRANSPARENCY=0.7 NOOUTLINE
        legendlabel="95% CI" NAME="Confidence Interval";
        yaxis max=&risk label="Cumulative risk (%)"     VALUEATTRS=(Color=black
Family=Arial Size=10)
        labelattrs=(Color=black Family=Arial Size=10 weight=bold);
        xaxis max=365 label="Time (days) from second dose of Vaccine"
        VALUEATTRS=(Color=black Family=Arial Size=10) labelattrs=(Color=black
Family=Arial Size=10 weight=bold);
        keylegend "plot1"/ title=" " position=bottom;
        keylegend "Confidence Interval"/title="95% CI" position=bottom;
```

```
	where matched="Matched cohort";
run;
ods graphics off;
title;

proc sql;
	select ceil(1000*max(RIsk))/1000 into :risk
	from work.suhom
	where daystodeath le 100;
run;

ods html on;
ods listing;
ods graphics on/width=9in height=7in;
proc sgpanel data=suhom;
	title "Suicide or Homicide deaths" bold font='arial';
	panelby matched  /novarname headerbackcolor=white spacing=5
	HEADERATTRS=(Color=black Family=Arial Size=10 Weight=Bold)  noheaderborder;
	step x= daystodeath y=risk/group=vaccinegroup2 LINEATTRS=(pattern=1
thickness=2)
	legendlabel="Estimate" name="plot1";
	band x=daystodeath upper=riskUCL lower=riskLCL/group=vaccinegroup2
TRANSPARENCY=0.7 NOOUTLINE
	legendlabel="95% CI" NAME="Confidence Interval";
	rowaxis max=&risk label="Cumulative risk (%)"     VALUEATTRS=(Color=black
Family=Arial Size=10)
```

```
	labelattrs=(Color=black Family=Arial Size=10 weight=bold);
	colaxis max=100 label="Time (days) from second dose of Vaccine"
	VALUEATTRS=(Color=black Family=Arial Size=10) labelattrs=(Color=black
Family=Arial Size=10 weight=bold);
	keylegend "plot1"/ title=" ";
	keylegend "Confidence Interval"/title="95% CI";
run;
ods graphics off;

data store.suhom;
	set suhom;
run;

data store.alldeaths;
	length outcome $30.;
	set allcause cvd covid noncovid suhom;
run;

proc freq data=store.alldeaths;
table outcome;
run;

ods html;
ods listing;
ods graphics on/width=9in height=7in;
proc sgpanel data=store.Alldeaths;
```

```
	title "Matched" bold font='arial';
	panelby outcome /novarname headerbackcolor=white spacing=5
	HEADERATTRS=(Color=black Family=Arial Size=10 Weight=Bold)  noheaderborder;
	step x= daystodeath y=risk/group=vaccinegroup2 LINEATTRS=(pattern=1
thickness=2)
	legendlabel="Estimate" name="plot1";
	band x=daystodeath upper=riskUCL lower=riskLCL/group=vaccinegroup2
TRANSPARENCY=0.7 NOOUTLINE
	legendlabel="95% CI" NAME="Confidence Interval";
	rowaxis label="Cumulative risk (%)"		VALUEATTRS=(Color=black Family=Arial
Size=10)
	labelattrs=(Color=black Family=Arial Size=10 weight=bold);
	colaxis max=365 label="Time (days) from second dose of Vaccine"
	VALUEATTRS=(Color=black Family=Arial Size=10) labelattrs=(Color=black
Family=Arial Size=10 weight=bold);
	keylegend "plot1"/ title=" ";
	keylegend "Confidence Interval"/title="95% CI";
	where matched="Matched observations" and outcome ne "Suicide or Homicide
deaths";
run;
ods graphics off;

/*** COVID  plus deaths ***/

%let data=DOSE1MATCHEDDATAFINAL;
run;
```

```
proc contents data=work.dose1matcheddatafinal;
run;

ods graphics on;
proc lifetest data=&data outsurv=COV2_Surv_matched method=km
	timelist=1 to 365 by 1 plots=none notable;
	time daystodeathd1*COVIDDEATHPLUS1YRD1 (0);
	where matchedpairs=1;
	strata firstdose;
run;

data COV2_Surv_matched;
	set COV2_Surv_matched;
	risk=100*(1-survival);
	riskUCL=100*(1-SDF_LCL);
	riskLCL=100*(1-SDF_UCL);
run;

ods graphics on;
proc lifetest data=&data outsurv=COV2_Surv_full method=km
	timelist=1 to 365 by 1 plots=none notable;
	time daystodeathd1*COVIDDEATHPLUS1YRD1(0);
	strata firstdose;
```

```
run;

data COV2_Surv_full;
	set COV2_Surv_full;
	risk=100*(1-survival);
	riskUCL=100*(1-SDF_LCL);
	riskLCL=100*(1-SDF_UCL);
run;

data COVIDplus;
	set COV2_Surv_matched (in=a) COV2_Surv_full (in=b);
	if a=1 then matched="Matched observations";
	else matched="All observations";
	length outcome $30.;
	outcome="3. COVID-19 deaths";
	if firstdose="1. PFIZER" then vaccinegroup2="BNT162b2";
	else if firstdose="2. MODERN" then vaccinegroup2="mRNA-1273";
run;

proc sort data=COVIDplus nodupkey;
	by vaccinegroup2 matched daystodeathd1 _CENSOR_;
run;

proc sql;
	select ceil(100*max(riskUCL))/100 into :risk
	from work.covidplus;
```

```
run;

ods html on;

ods listing;

ods graphics on/width=9in height=7in;

proc sgpanel data=COVIDplus;

	title "COVID-19+ deaths" bold font='arial';

	panelby matched  /novarname headerbackcolor=white spacing=5

	HEADERATTRS=(Color=black Family=Arial Size=10 Weight=Bold) noheaderborder;

	series x= daystodeathd1 y=risk/group=vaccinegroup2 LINEATTRS=(pattern=1
thickness=2)

	legendlabel="Estimate" name="plot1";

	band x=daystodeathd1 upper=riskUCL lower=riskLCL/group=vaccinegroup2
TRANSPARENCY=0.7 NOOUTLINE

	legendlabel="95% CI" NAME="Confidence Interval";

	rowaxis max=&risk label="Cumulative risk (%)"     VALUEATTRS=(Color=black
Family=Arial Size=10)

	labelattrs=(Color=black Family=Arial Size=10 weight=bold);

	colaxis max=365 label="Time (days) from second dose of Vaccine"

	VALUEATTRS=(Color=black Family=Arial Size=10) labelattrs=(Color=black
Family=Arial Size=10 weight=bold);

	keylegend "plot1"/ title=" ";

	keylegend "Confidence Interval"/title="95% CI";

run;

ods graphics off;

title;
```

```
ods graphics on/width=4in height=3in;

proc sgplot data=COVIDplus;

	title "COVID-19+ Deaths" bold font='arial';

	step x= daystodeathd1 y=risk/group=vaccinegroup2 LINEATTRS=(pattern=1
thickness=2)

	legendlabel="Estimate" name="plot1";

	band x=daystodeathd1 upper=riskUCL lower=riskLCL/group=vaccinegroup2
TRANSPARENCY=0.7 NOOUTLINE

	legendlabel="95% CI" NAME="Confidence Interval";

	yaxis max=&risk label="Cumulative risk (%)"	VALUEATTRS=(Color=black
Family=Arial Size=10)

	labelattrs=(Color=black Family=Arial Size=10 weight=bold);

	xaxis max=365 label="Time (days) from second dose of Vaccine"

	VALUEATTRS=(Color=black Family=Arial Size=10) labelattrs=(Color=black
Family=Arial Size=10 weight=bold);

	keylegend "plot1"/ title=" " position=bottom;

	keylegend "Confidence Interval"/title="95% CI" position=bottom;

	where matched="All observations";

run;

ods graphics off;

proc sql;

	select ceil(100*max(riskUCL))/100 into :risk

	from work.covidplus

	where matched="Matched observations";

run;
```

```
ods graphics on/width=4in height=3in;

proc sgplot data=Covidplus;
	title "COVID-19+ Deaths" bold font='arial';
	step x= daystodeathd1 y=risk/group=vaccinegroup2 LINEATTRS=(pattern=1 thickness=2)
	legendlabel="Estimate" name="plot1";
	band x=daystodeathd1 upper=riskUCL lower=riskLCL/group=vaccinegroup2 TRANSPARENCY=0.7 NOOUTLINE
	legendlabel="95% CI" NAME="Confidence Interval";
	yaxis max=&risk label="Cumulative risk (%)"	VALUEATTRS=(Color=black Family=Arial Size=10)
	labelattrs=(Color=black Family=Arial Size=10 weight=bold);
	xaxis max=365 label="Time (days) from second dose of Vaccine"
	VALUEATTRS=(Color=black Family=Arial Size=10) labelattrs=(Color=black Family=Arial Size=10 weight=bold);
	keylegend "plot1"/ title=" " position=bottom;
	keylegend "Confidence Interval"/title="95% CI" position=bottom;
	where matched="Matched observations";
run;
ods graphics off;

proc sql;
```

```
        select ceil(1000*max(riskUCL))/1000 into :risk
        from work.covid
        where daystodeath le 100;
run;

ods html on;
ods listing;
ods graphics on/width=9in height=7in;

proc sgpanel data=COVIDplus;
        title "COVID+ deaths" bold font='arial';
        panelby matched  /novarname headerbackcolor=white spacing=5
        HEADERATTRS=(Color=black Family=Arial Size=10 Weight=Bold) noheaderborder;
        series x= daystodeath y=risk/group=vaccinegroup2 LINEATTRS=(pattern=1
thickness=2)
        legendlabel="Estimate" name="plot1";
        band x=daystodeath upper=riskUCL lower=riskLCL/group=vaccinegroup2
TRANSPARENCY=0.7 NOOUTLINE
        legendlabel="95% CI" NAME="Confidence Interval";
        rowaxis max=&risk label="Cumulative risk (%)"     VALUEATTRS=(Color=black
Family=Arial Size=10)
        labelattrs=(Color=black Family=Arial Size=10 weight=bold);
        colaxis max=100 label="Time (days) from second dose of Vaccine"
        VALUEATTRS=(Color=black Family=Arial Size=10) labelattrs=(Color=black
Family=Arial Size=10 weight=bold);
        keylegend "plot1"/ title=" ";
        keylegend "Confidence Interval"/title="95% CI";
```

```
run;
ods graphics off;

/*** COVID  plus deaths ***/

%let data=DOSE1MATCHEDDATAFINAL;
run;

proc contents data=work.dose1matcheddatafinal;
run;

ods graphics on;
proc lifetest data=&data outsurv=nonCOV2_Surv_matched method=km
	timelist=1 to 365 by 1 plots=none notable;
	time daystodeathd1*nonCOVIDDEATHPLUS1YRD1 (0);
	where matchedpairs=1;
	strata firstdose;
run;

data nonCOV2_Surv_matched;
	set nonCOV2_Surv_matched;
	risk=100*(1-survival);
	riskUCL=100*(1-SDF_LCL);
```

```
        riskLCL=100*(1-SDF_UCL);
run;

ods graphics on;
proc lifetest data=&data outsurv=nonCOV2_Surv_full method=km
        timelist=1 to 365 by 1 plots=none notable;
        time daystodeathd1*nonCOVIDDEATHPLUS1YRD1(0);
        strata firstdose;
run;

data nonCOV2_Surv_full;
        set nonCOV2_Surv_full;
        risk=100*(1-survival);
        riskUCL=100*(1-SDF_LCL);
        riskLCL=100*(1-SDF_UCL);
run;

data nonCOVIDplus;
        set nonCOV2_Surv_matched (in=a) nonCOV2_Surv_full (in=b);
        if a=1 then matched="Matched observations";
        else matched="All observations";
        length outcome $30.;
        outcome="3. COVID-19 deaths";
        if firstdose="1. PFIZER" then vaccinegroup2="BNT162b2";
        else if firstdose="2. MODERN" then vaccinegroup2="mRNA-1273";
```

```
run;

proc sort data=nonCOVIDplus nodupkey;
	by vaccinegroup2 matched daystodeathd1 _CENSOR_;
run;

proc sql;
	select ceil(100*max(riskUCL))/100 into :risk
	from work.noncovidplus;
run;

ods html on;
ods listing;
ods graphics on/width=9in height=7in;

proc sgpanel data=nonCOVIDplus;
	title "Non-COVID+ deaths" bold font='arial';
	panelby matched  /novarname headerbackcolor=white spacing=5
	HEADERATTRS=(Color=black Family=Arial Size=10 Weight=Bold) noheaderborder;
	series x= daystodeathd1 y=risk/group=vaccinegroup2 LINEATTRS=(pattern=1
thickness=2)
	legendlabel="Estimate" name="plot1";
	band x=daystodeathd1 upper=riskUCL lower=riskLCL/group=vaccinegroup2
TRANSPARENCY=0.7 NOOUTLINE
	legendlabel="95% CI" NAME="Confidence Interval";
	rowaxis max=&risk label="Cumulative risk (%)"     VALUEATTRS=(Color=black
Family=Arial Size=10)
```

```
labelattrs=(Color=black Family=Arial Size=10 weight=bold);
colaxis max=365 label="Time (days) from second dose of Vaccine"
VALUEATTRS=(Color=black Family=Arial Size=10) labelattrs=(Color=black Family=Arial Size=10 weight=bold);
keylegend "plot1"/ title=" ";
keylegend "Confidence Interval"/title="95% CI";
run;
ods graphics off;
title;

ods graphics on/width=4in height=3in;
proc sgplot data=nonCOVIDplus;
title "Non-COVID+ Deaths" bold font='arial';
step x= daystodeathd1 y=risk/group=vaccinegroup2 LINEATTRS=(pattern=1 thickness=2)
legendlabel="Estimate" name="plot1";
band x=daystodeathd1 upper=riskUCL lower=riskLCL/group=vaccinegroup2 TRANSPARENCY=0.7 NOOUTLINE
legendlabel="95% CI" NAME="Confidence Interval";
yaxis max=&risk label="Cumulative risk (%)"     VALUEATTRS=(Color=black Family=Arial Size=10)
labelattrs=(Color=black Family=Arial Size=10 weight=bold);
xaxis max=365 label="Time (days) from second dose of Vaccine"
VALUEATTRS=(Color=black Family=Arial Size=10) labelattrs=(Color=black Family=Arial Size=10 weight=bold);
keylegend "plot1"/ title=" " position=bottom;
keylegend "Confidence Interval"/title="95% CI" position=bottom;
```

```
        where matched="All observations";
run;
ods graphics off;

proc sql;
        select ceil(100*max(riskUCL))/100 into :risk
        from work.noncovidplus
        where matched="Matched observations";
run;

ods graphics on/width=4in height=3in;

proc sgplot data=nonCovidplus;
        title "Non-COVID+ Deaths" bold font='arial';
        step x= daystodeathd1 y=risk/group=vaccinegroup2 LINEATTRS=(pattern=1
thickness=2)
        legendlabel="Estimate" name="plot1";
        band x=daystodeathd1 upper=riskUCL lower=riskLCL/group=vaccinegroup2
TRANSPARENCY=0.7 NOOUTLINE
        legendlabel="95% CI" NAME="Confidence Interval";
        yaxis max=&risk label="Cumulative risk (%)"        VALUEATTRS=(Color=black
Family=Arial Size=10)
        labelattrs=(Color=black Family=Arial Size=10 weight=bold);
        xaxis max=365 label="Time (days) from second dose of Vaccine"
        VALUEATTRS=(Color=black Family=Arial Size=10) labelattrs=(Color=black
Family=Arial Size=10 weight=bold);
```

```
        keylegend "plot1"/ title=" " position=bottom;

        keylegend "Confidence Interval"/title="95% CI" position=bottom;

        where matched="Matched observations";

run;

ods graphics off;

/*** Not necessary ****

proc template;

        define statgraph inset;

        begingraph;

        entrytitle 'All cause deaths';

        layout datapanel classvars=(matched)/rows=1 headerlabeldisplay=value ;

         layout gridded / width=300px height=200px halign=right valign=top;

                layout prototype;

                seriesplot x=daystodeath y=risk/group=vaccinegroup2
LINEATTRS=(pattern=5 thickness=3);

                endlayout;

        endlayout;

        endgraph;

        end;

run;

proc sgrender data=work.allcause template=inset;
```

```
run;

proc template;
define statgraph inset;
begingraph;
entrytitle 'All cause deaths';
 layout overlay;
   histogram cholesterol /  binaxis=false datatransparency=0.5
             display=(fillpattern outline fill)      fillattrs=(color=lightred)
             fillpatternattrs=(pattern=l3 color=red);
   densityplot cholesterol /  datatransparency=0.5 lineattrs=(color=darkred);
 layout gridded / width=300px height=200px halign=right valign=top;
   layout overlay / yaxisopts=(label='Blood Pressure' linearopts=(thresholdmax=1))
             xaxisopts=(linearopts=(thresholdmax=1));
     regressionplot x=cholesterol y=diastolic / lineattrs=(color=blue pattern=2)
name='Diastolic';
     regressionplot x=cholesterol y=systolic / lineattrs=(color=green) name='Systolic';
     discretelegend 'Diastolic' 'Systolic' / across=2;
   endlayout;
 endlayout;
 endlayout;
endgraph;
end;
run;
```

```
proc sgrender data=sashelp.heart template=inset;
run;

***/

proc contents data=work.matcheddata1;
run;

proc freq data=work.matcheddata1;
table sexf;
run;

/*** Calculation for standardized differences ****/
%include "H:\EPI\Surveillance Section\Fahad Mansuri\SURVIVAL ANALYSIS 2024\SAS CODE\std diff macro.sas";

data matchedpairs;
	set work.DOSE1MATCHEDDATAFINAL;
	if firstdose="1. PFIZER" THEN MODERNA=0;
	ELSE MODERNA=1;
	WHERE MATCHEDPAIRS=1;
	IF SEX2='F' THEN SEXF=1;
	ELSE SEXF=0;
run;
```

```
data matchedALL;
	set work.DOSE1MATCHEDDATAFINAL;
	if firstdose="1. PFIZER" THEN MODERNA=0;
	ELSE MODERNA=1;
	IF SEX2='F' THEN SEXF=1;
	ELSE SEXF=0;
run;

proc freq data=
PROC CONTENTS DATA=DOSE1MATCHEDDATAFINAL ORDER=VARNUM;
RUN;

%stddiff( inds = matchedALL,
				groupvar =MODERNA,
				numvars = agenew sviscore sviscore1 sviscore4,
				charvars = race4 ethnicity3 monthdose1 sexf vaccinationsite,
				wtvar = ,
				stdfmt = 8.4,
				outds = stddiff_result );

%stddiff( inds = matchedpairs,
				groupvar =moderna,
				numvars = agenew sviscore sviscore1 sviscore4,
```

charvars = race4 ethnicity3 monthdose1 sexf vaccinationsite,

wtvar = ,

stdfmt = **8.4**,

outds = stddiff_result1);

```
libname store 'C:\Users\MansuriF\OneDrive - Florida Department of
Health\Documents\SAS Datasets';

run;

LIBNAME Deaths ODBC NOPROMPT="driver={ODBC Driver 17 for SQL Server};

Server=CHARTSDATA.DOH.AD.STATE.FL.US;

Database=Vitalstat;

Trusted_Connection=Yes"  schema=dbo stringdates=NO ;

run;

LIBNAME COVVAC ODBC NOPROMPT="Driver={ODBC Driver 17 for SQL Server};

        Server=merlindbreport.doh.ad.state.fl.us;

        Database=CovidVaccine;Trusted_Connection=Yes" schema=dbo stringdates=Yes;

run;

LIBNAME COVID ODBC NOPROMPT="Driver={ODBC Driver 17 for SQL Server};

        Server=merlindbreport.doh.ad.state.fl.us;

        Database=MerlinCOVID;Trusted_Connection=Yes" schema=dbo stringdates=Yes;

run;

options validvarname=any;

%include "H:\EPI\Surveillance Section\Fahad Mansuri\SURVIVAL ANALYSIS 2024\SAS
CODE\formats for analysis.sas";
```

```
%include "H:\EPI\Surveillance Section\Fahad Mansuri\SURVIVAL ANALYSIS 2024\SAS CODE\relrisk9.sas";

proc means data=store.dose1matcheddata nmiss;
	var sviscore:;
run;

proc stdize data=store.dose1matcheddata reponly method=mean missing=mean out=dose1matcheddata;
	var sviscore:;
run;

proc means data=dose1matcheddata nmiss;
	var sviscore:;
run;

/*** Descriptives for matched data vs all data ***/

proc rank data=dose1matcheddata out=dose1matcheddata groups=10;
	var sviscore1 sviscore2 sviscore3 sviscore4 sviscore SESPOV
	SESUNEMP SESINCOME	SESNOHIGH	HTMUS	HTMOBILEHOME
	HTNOVEHICLE	HTCROWDING	HTGQ;
	ranks decile_sviscore1 decile_sviscore2 decile_sviscore3 decile_sviscore4 decile_sviscore
	decile_SESPOV	decile_SESUNEMP decile_SESINCOME	decile_SESNOHIGH
	decile_HTMUS	decile_HTMOBILEHOME
	decile_HTNOVEHICLE	decile_HTCROWDING	decile_HTGQ;
```

```
run;

proc contents data=store.dose1matcheddata order=varnum;
run;

proc sort data=dose1matcheddata nodupkey out=test;
	by DWHPatient;
run;

LIBNAME COVID ODBC NOPROMPT="Driver={ODBC Driver 17 for SQL Server};
	Server=merlindbreport.doh.ad.state.fl.us;
	Database=MerlinCOVID;Trusted_Connection=Yes" schema=dbo stringdates=no;
run;

proc contents data=covid.cases order=varnum;
run;

proc contents data=COVVAC.COVIDPatientIdentity order=varnum;
run;

data covidcases;
	set covid.cases;
	keep 'Case ID'n 'Profile ID'n 'shots id'n 'Publish case'n Jurisdiction
	'Earliest lab event date'n 'Date became case'n 'Dx status'n;
	rename 'shots id'n =shotsid2 'Publish case'n =Publishcase
	'Earliest lab event date'n=ELD
```

```
    'Date became case'n =casedate
    'Dx status'n=dxstatus 'Profile ID'n=profileid 'Case ID'n=caseid;
    where upcase('Publish case'n)="YES" and  "01Mar2020"d le 'Earliest lab event
date'n le "31dec2022"D
    and     Jurisdiction="FL resident";
run;

proc sql;
    select count(distinct profileid)
    from covidcases
    where shotsid2 ne " ";
quit;

proc sql;
    select count (profileid)
    from covidcases
    where shotsid2 ne " ";
quit;

proc sql;
    select count(distinct caseid)
    from covidcases      where shotsid2 ne " ";
quit;

proc sql;
    select count(caseid)
```

```
        from covidcases        where shotsid2 ne " ";
quit;

proc sql;
        select count (distinct shotsid2)
                from covidcases        where shotsid2 ne " ";
quit;

proc sql;
        select count (shotsid2)
                from covidcases        where shotsid2 ne " ";
quit;

proc sort data=work.covidcases;
        by shotsid2;
run;

proc transpose data=covidcases out=covidcases2 name=ELD prefix=ELD;
        by shotsid2;
        var eld;
        where shotsid2 ne " ";
run;

proc freq data=covidcases;
        table Jurisdiction dxstatus;
run;
```

```
proc contents data=store.dose1matcheddata;
run;

proc datasets lib=work;
run;

proc sort data=vaccinated12;
	by shotsid2;
run;

proc sort data=covidcases2;
	by shotsid2;
run;

proc sql;
	create table dose1matchedatawithcovid as
	select a.*,b.*
	from dose1matcheddata as a
	left join covidcases2 as b
	on a.CurrentStateIMMId=b.shotsid2;
quit;

options validvarname=any;
```

```
proc contents data=store.covidvaccination5 order=varnum;
run;

data dose1matcheddatafinal;
	set dose1matchedatawithcovid (keep= ageatdose1 agegroupnew sex2 race4 ethnicity3 firstdose agenew
		decile_sviscore1 decile_sviscore4 mortalityratepct Vaccinationsite
		vaccinedose2 seconddose DWHPatient CurrentStateIMMId VaccinationDate1
		DWHPatient firstdose seconddose agenew sex2 race4 ethnicity3 Vaccinationsite
		sviscore: VaccinationDate: monthdose1 timebetweendose timeokay mortalityratepct
		matchedpairs icd_code geoidnew vaccinedose2 daystodeath daystodeathd1 death1yrd1 death1yr
		'vs dod'n nursinghomevaccinationfinal HOMELESSFLAG
		TRANSAX1-TRANSAX15 eld:);
	where SEX2 ne "U" and
	Vaccinationsite not in ("CORRECTIONAL FACILITY","NURSING HOME")
	and nursinghomevaccinationfinal ne 1 AND HOMELESSFLAG=0;
	if seconddose=" " then noseconddose=1;
	else noseconddose=0;
	format agenew age10ygroup.;
	mortalityratepct=round(mortalityratepct,1);
	IF FIRSTDOSE="1. PFIZER" THEN EXPOS=1;
	ELSE EXPOS=0;
```

```
if death1yrd1=1 and substr(icd_code,1,2) in ("I0","I1","I2","I3","I4","I5","I6") then CVD1YRD1=1;
else if death1yrD1=1 and substr(icd_code,1,3) in ("I71","I72","I73","I74","I75","I76","I77") then CVD1YRD1=1;
else CVD1YRD1=0;
if SUBSTR(icd_code,1,3) IN ("X40","X41","X42","X43","X44",
"X60","X61","X62","X62","X64","X85","Y10","Y11","Y12","Y13","Y14")
and death1yrD1=1 then overdose1yrD1=1;
else overdose1yrD1=0;
if SUBSTR(icd_code,1,3) in ("X85","X86","X87","X88","X89","X90","X91","X92"
        ,"X93","X94","X95","X96","X97","X98","X99","Y00","Y01","Y02","Y03"
        ,"Y04","Y05","Y06","Y07","Y08","Y09")
and DEATH1YRD1=1 then homicide1yrD1=1;
else if substr(icd_code,1,4)="Y871" and death1yrD1=1 then homicide1yrD1=1;
else homicide1yrD1=0;
if SUBSTR (ICD_CODE,1,2) IN ("X6","X7") AND death1yrD1=1 THEN SUICIDE1YRD1=1;
ELSE IF SUBSTR (ICD_CODE,1,3) IN ("X80","X81","X82","X83","X84") AND DEATH1YRD1=1 THEN SUICIDE1YRD1=1;
ELSE IF substr(icd_code,1,4)="Y870" and death1yr=1 THEN SUICIDE1YR=1;
ELSE SUICIDE1YRD1=0;
if SUBSTR (ICD_CODE,1,2) IN ("V0","V1","V2","V3","V4","V5","V6","V7","V8","V9"
,"W0","W1","W2","W3","W4","W5","W6","W7","W8","W9","X0","X1","X2","X3","X4")
AND DEATH1YRD1=1 THEN UNINTENTIONAL1YRD1=1;
ELSE IF SUBSTR(ICD_CODE,1,3) IN ("X50","X51","X52","X53","X54","X55","X56","X57","X58","X59"
        ,"Y85","Y86") AND DEATH1YRD1=1 THEN UNINTENTIONAL1YRD1=1;
```

ELSE UNINTENTIONAL1YRD1=**0**;

if SUBSTR (ICD_CODE,**1**,**2**) IN ("V0","V1","V2","V3","V4","V5","V6","V7","V8","V9"

,"W2","W3","W4","W5","W6","W7","W8","W9","X0","X1","X2","X3","X4")

AND DEATH1YRD1=**1** THEN NOFALLUNINTENTIONAL1YRD1=**1**;

ELSE IF SUBSTR(ICD_CODE,**1**,**3**) IN ("X50","X51","X52","X53","X54","X55","X56","X57","X58","X59"

,"Y85","Y86") AND DEATH1YRD1=**1** THEN NOFALLUNINTENTIONAL1YRD1=**1**;

ELSE NOFALLUNINTENTIONAL1YRD1=**0**;

if homicide1yrD1=**1** or UNINTENTIONAL1YRD1=**1** or SUICIDE1YRD1=**1** or overdose1yrD1=**1** then unnatural1yrD1=**1**;

else unnatural1yrD1=**0**;

if homicide1yrD1=**1** or SUICIDE1YRD1=**1** then suicidehomicide1yrD1=**1**;

else suicidehomicide1yrD1=**0**;

if homicide1yrD1=**1** or NOFALLUNINTENTIONAL1YRD1=**1** or SUICIDE1YRD1=**1** or overdose1yrD1=**1**

then NOFALLunnatural1yrD1=**1**;

else NOFALLunnatural1yrD1=**0**;

if SUBSTR(ICD_CODE,**1**,**2**) in ("V2","V3","V4","V5","V6","V7") and death1yrD1=**1** then mva1yrD1=**1**;

else if SUBSTR(ICD_CODE,**1**,**3**) in ("V02","V03","V04","V12","V13","V14","V83","V84","V85","V86")

and death1yrD1=**1** then mva1yrD1=**1**;

if icd_code in ("V090","V092","V190","V191","V192","V194","V195","V196","V803","V804",

```
"V805","V810","V811","V820","V821","V870","V871","V872","V873","V874","V875",
"V876","V877","V878","V881","V882","V883","V884","V885","V886","V887","V888","V890"
,"V892")
AND DEATH1YRD1=1 THEN MVA1YRD1=1;
ELSE MVA1YRD1=0;
array test {*} transax: ; /*** Select all columns with ACME prefix ***/
count =0;
do j= 1 to 15;
    if test{j} ne " " then do;
    count + (test{j} in: ("U071"));
    end;
end;
if count>0 then COVIDDEATH=1;
else COVIDDEATH=0;
if COVIDDEATH=1 and DEATH1YRD1=1 then COVIDDEATH1YRD1=1;
ELSE COVIDDEATH1YRD1=0;
if COVIDDEATH=0 and DEATH1YRD1=1 then NONCOVIDDEATH1YRD1=1;
ELSE NONCOVIDDEATH1YRD1=0;
counteld=0;
array new ELD1-ELD6;
    do i =1 to dim(new);
    if new{i} ne . and 'vs dod'n ne . then do;
    counteld+(vaccinationdate1 le new{i} <= 'vs dod'n);
    end;
end;
if DEATH1YRD1=1 and (COVIDDEATH1YRD1=1 or counteld >0) then
COVIDDEATHPLUS1YRD1=1;
```

```
	ELSE COVIDDEATHPLUS1YRD1=0;

	if DEATH1YRD1=1 and (COVIDDEATHPLUS1YRD1=0) then
NONCOVIDDEATHPLUS1YRD1=1;
	ELSE NONCOVIDDEATHPLUS1YRD1=0;

	if counteld>0 then do;
		if eld6 ne . and 'vs dod'n >=eld6 then covinftodeathdys='vs dod'n-eld6;
		else if eld5 ne . and 'vs dod'n >=eld5 then covinftodeathdys='vs dod'n-eld5;
		else if eld4 ne . and 'vs dod'n >=eld4 then covinftodeathdys='vs dod'n-eld4;
		else if eld3 ne . and 'vs dod'n >=eld3 then covinftodeathdys='vs dod'n-eld3;
		else if eld2 ne . and 'vs dod'n >=eld2 then covinftodeathdys='vs dod'n-eld2;

		else if eld1 ne . and 'vs dod'n >=eld1 then covinftodeathdys='vs dod'n-eld1;
	end;
run;

proc freq data=work.dose1matcheddatafinal;
	table counteld*death1yrd1;
	table covinftodeathdys;
run;

proc print data=work.dose1matcheddatafinal (obs= 100);
	where COVIDDEATHPLUS1YRD1=1 and covinftodeathdys<0;
	var DWHPatient VaccinationDate: eld: counteld 'vs dod'n covinftodeathdys;
run;
```

```
proc freq data=dose1matcheddatafinal;
	tABLE daystodeath;
RUN;

proc freq data=dose1matcheddatafinal;
	tABLE death1yrd1 CVD: covid: NONCOVID: suicidehomicide1yrD1;
RUN;

title;
proc freq data=dose1matcheddatafinal;
	table ageatdose1 sex2 race4 ethnicity3 firstdose seconddose noseconddose
	vaccinedose2/missing;
	format ageatdose1 age10ygroup.;
run;

title;
proc freq data=dose1matcheddatafinal;
	table ageatdose1 sex2 race4 ethnicity3 firstdose seconddose noseconddose
	vaccinedose2/missing;
	format ageatdose1 age10ygroup.;
	where matchedpairs=1;
```

```
run;

title;
proc freq data=dose1matcheddatafinal;
	table (ageatdose1 sex2 race4 ethnicity3 firstdose seconddose noseconddose
	vaccinedose2)*firstdose/missing crosslist;
	format ageatdose1 age10ygroup.;
	where matchedpairs=1;
run;

title;
proc freq data=dose1matcheddatafinal;
	table (ageatdose1 sex2 race4 ethnicity3 firstdose seconddose noseconddose
	vaccinedose2)*firstdose/missing crosslist;
	format ageatdose1 age10ygroup.;
run;

proc freq data=work.dose1matcheddatafinal;
	table mortalityratepct;
run;

proc freq data=dose1matcheddatafinal;
	table Vaccinationsite VaccinationDate1;
```

```
	format VaccinationDate1 month7.;
run;

proc freq data=dose1matcheddatafinal;
	table Vaccinationsite VaccinationDate1;
	format VaccinationDate1 month7.;
	where matchedpairs=1;
run;

proc freq data=dose1matcheddatafinal;
	table vaccinedose2*(ageatdose1 sex2 race4 ethnicity3)/nopercent crosslist;
	format ageatdose1 age10ygroup.;
run;

/*** DESCRIPTIVES BY EXPOSURE ***/

proc freq data=dose1matcheddatafinal;
	table FIRSTDOSE*(ageatdose1 sex2 race4 ethnicity3 seconddose noseconddose
vaccinedose2)/missing CROSSLIST;
	format ageatdose1 age10ygroup.;
run;

proc freq data=dose1matcheddatafinal;
	table firstdose*(Vaccinationsite VaccinationDate1)/missing CROSSLIST;
	format VaccinationDate1 month7.;
run;
```

```
proc freq data=dose1matcheddatafinal;
	table FIRSTDOSE*(ageatdose1 sex2 race4 ethnicity3 seconddose noseconddose vaccinedose2)/missing CROSSLIST;
	format ageatdose1 age10ygroup.;
	where matchedpairs=1;
run;

proc freq data=dose1matcheddatafinal;
	table firstdose*(Vaccinationsite VaccinationDate1)/missing CROSSLIST;
	format VaccinationDate1 month7.;
	where matchedpairs=1;
run;

/*** Risk of All-cause, CVD, COVID, and Non-COVID Deaths          ***/

/*** MATCHED COHORT          ***/

%let dataset=MATCHEDCOHORT;

%let todaysdate=%sysfunc(today(),date9.);

%put &todaysdate;

%let textadd=Matched cohort Risk of death Dose 1;
```

```
%let path=H:\EPI\Surveillance Section\Fahad Mansuri\SURVIVAL ANALYSIS 2024;

proc contents data=work.MATCHEDCOHORT;
run;

proc freq data=work.dose1matcheddatafinal;
        table firstdose agenew firstdose agenew sex2 race4 ethnicity3
                        decile_sviscore1 decile_sviscore4
                       mortalityratepct Vaccinationsite/ missing;
run;

/*** 1.a. SUIC or HOMICIDE  with categorical age ***/

ods pdf file="&path\SAS output\&textadd\SUICIDE or HOMICIDE\
&textadd Logistic SUICHOM &todaysdate..pdf";
ods output ODDSRATIOS=log_SUICHOMcrude;
proc logistic data=&dataset;
        title "&textadd SUICIDE OR HOMICIDE deaths" ;
        class firstdose (REF=last)/param=reference;
        model suicidehomicide1yrD1(ref='0')=firstdose/ parmlabel;
run;
ODS OUTPUT CLOSE;
ods pdf close;
title;
```

```
proc print;
run;

data log_SUICHOMcrude1;
	set log_SUICHOMcrude;
	ODDSRatio1=put(OddsRatioEst,8.3);
	LowerCL1=put(LowerCL,8.3);
	UpperCL1=put(UpperCL,8.3);
	OR=STRIP(ODDSRatio1)||" ("||STRIP(LowerCL1)||", "||STRIP(UpperCL1)||')';
	MODEL=1;
run;

proc print;
run;

ods pdf file="&path\SAS output\&textadd\SUICIDE OR HOMICIDE\
&textadd Logistic SUICHOM adjusted model (cat age) &todaysdate..pdf";
ods output ODDSRATIOS=Log_SUICHOMadjusted;
proc logistic data=&dataset;
	TITLE 'SUICIDE OR HOMICIDE death adjusted (logistic regression)';
	class agenew (ref=first) firstdose (REF=last) sex2 (ref="M")
	race4 (ref='WHITE')  ethnicity3
	(ref='NOT HISPANIC') Vaccinationsite (ref="PUBLIX OR CVS OR
WALGREENS")/param=reference;
	model suicidehomicide1yrD1(ref='0')= firstdose agenew sex2 race4 ethnicity3
```

```
                              decile_sviscore1 decile_sviscore4
                              mortalityratepct Vaccinationsite/parmlabel;
                   format agenew AGE10yGROUP.;
run;
ods output close;
ods pdf close;
title;

proc print;
run;

data Log_SUICHOMadjusted1;
     set Log_SUICHOMadjusted;
     ODDSRatio1=put(OddsRatioEst,8.3);
     LowerCL1=put(LowerCL,8.3);
     UpperCL1=put(UpperCL,8.3);
     OR=STRIP(ODDSRatio1)||" ("||STRIP(LowerCL1)||", "||STRIP(UpperCL1)||')';
     MODEL=2;
run;

proc print data=Log_SUICHOMadjusted1;
     where SCAN(upcase(EFFECT),1)='FIRSTDOSE';
run;

DATA LOGSUICHOM;
     SET lOG_SUICHOMCRUDE1 Log_SUICHOMadjusted1;
```

```
RUN;

PROC PRINT data= LOGSUICHOM;
	where SCAN(upcase(EFFECT),1)='FIRSTDOSE';
RUN;

proc sort data=&dataset;
	by agenew;
run;

ods pdf file="&path\SAS output\&textadd\SUICIDE OR HOMICIDE\
&textadd logistic SUICHOM adjusted model (cat age) by Age (3 Age groups)
&todaysdate..pdf";
ods output ODDSRATIOS=Log_ageSUICHOMadjusted3g;
proc logistic data=&dataset;
	title 'SUICIDE OR HOMICIDE deaths adjusted by Age';
	class agenew (ref=first) FIRSTDOSE (REF=LAST) sex2 (ref="M")
	race4 (ref='WHITE')  ethnicity3
	(ref='NOT HISPANIC') Vaccinationsite (ref="PUBLIX OR CVS OR
WALGREENS")/param=reference;
	model suicidehomicide1yrD1(ref='0')= FIRSTDOSE agenew sex2 race4 ethnicity3
				decile_sviscore1 decile_sviscore4
				mortalityratepct Vaccinationsite/parmlabel;
	by agenew;
	format agenew age3groups.;
run;
ods output close;
```

```
ods pdf close;

title;

proc print data=Log_ageSUICHOMadjusted3g;

run;

data Log_ageSUICHOMadjusted3g1;

	set Log_ageSUICHOMadjusted3g;

	ODDSRatio1=put(OddsRatioEst,8.3);

	LowerCL1=put(LowerCL,8.3);

	UpperCL1=put(UpperCL,8.3);

	OR=STRIP(ODDSRatio1)||" ("||STRIP(LowerCL1)||", "||STRIP(UpperCL1)||')';

	MODEL=2;

run;

proc print data=Log_ageSUICHOMadjusted3g1;

	where SCAN(upcase(EFFECT),1)='FIRSTDOSE';

run;

/*** output excel file for results ***/

libname results xlsx "&path\SAS output\&textadd\SUICIDE OR HOMICIDE\SUICIDE OR HOMICIDE.xlsx";

/* Export first dataset */

data results.sheet1;

   set work.logSUICHOM;
```

```
            where SCAN(upcase(EFFECT),1)='FIRSTDOSE';
run;

/* Export third dataset */
data results.sheet3;
   set work.Log_ageSUICHOMadjusted3g1;
            where SCAN(upcase(EFFECT),1)='FIRSTDOSE';
run;

libname results clear;

/*** 1.a. NON COVID +  with categorical age ***/

ods pdf file="&path\SAS output\&textadd\NON COVID PLUS\
&textadd Logistic COVID + &todaysdate..pdf";
ods output ODDSRATIOS=log_NONCOV2crude;
proc logistic data=&dataset;
            title "&textadd NON COVID + deaths" ;
            class firstdose (REF=last)/param=reference;
            model NONCOVIDDEATHPLUS1YRD1(ref='0')=firstdose/ parmlabel;
run;
ODS OUTPUT CLOSE;
ods pdf close;
title;

proc print;
```

```
run;

data Log_NONCOV2crude1;
	set Log_NONCOV2crude;
	ODDSRatio1=put(OddsRatioEst,8.3);
	LowerCL1=put(LowerCL,8.3);
	UpperCL1=put(UpperCL,8.3);
	OR=STRIP(ODDSRatio1)||" ("||STRIP(LowerCL1)||", "||STRIP(UpperCL1)||')';
	MODEL=1;
run;

proc print;
run;

ods pdf file="&path\SAS output\&textadd\NON COVID PLUS\
&textadd Logistic NON COVID + adjusted model (cat age) &todaysdate..pdf";
ods output ODDSRATIOS=Log_NONCOV2adjusted;
proc logistic data=&dataset;
	TITLE 'NON COVID + death adjusted (logistic regression)';
	class agenew (ref=first) firstdose (REF=last) sex2 (ref="M")
	race4 (ref='WHITE')  ethnicity3
	(ref='NOT HISPANIC') Vaccinationsite (ref="PUBLIX OR CVS OR
WALGREENS")/param=reference;
	model NONCOVIDDEATHPLUS1YRD1(ref='0')= firstdose agenew sex2 race4
ethnicity3
				decile_sviscore1 decile_sviscore4
```

```
                    mortalityratepct Vaccinationsite/parmlabel;
                format agenew AGE10yGROUP.;
run;
ods output close;
ods pdf close;
title;

proc print;
run;

data Log_NONCOV2adjusted1;
    set Log_NONCOV2adjusted;
    ODDSRatio1=put(OddsRatioEst,8.3);
    LowerCL1=put(LowerCL,8.3);
    UpperCL1=put(UpperCL,8.3);
    OR=STRIP(ODDSRatio1)||" ("||STRIP(LowerCL1)||", "||STRIP(UpperCL1)||')';
    MODEL=2;
run;

proc print data=Log_NONCOV2adjusted1;
    where SCAN(upcase(EFFECT),1)='VACCINEGROUP2';
run;

DATA LOGNONCOV2;
    SET lOG_NONCOV2CRUDE1 lOG_NONCOV2adjusted1;
RUN;
```

```
PROC PRINT data= LOGNONCOV2;
	where SCAN(upcase(EFFECT),1)='FIRSTDOSE';
RUN;

proc sort data=&dataset;
	by agenew;
run;

ods pdf file="&path\SAS output\&textadd\NON COVID PLUS\
&textadd logistic NON COVID + adjusted model (cat age) by Age (3 Age groups)
&todaysdate..pdf";
ods output ODDSRATIOS=Log_ageNONCOV2adjusted3g;
proc logistic data=&dataset;
	title 'NON COVID + deaths adjusted by Age';
	class agenew (ref=first) FIRSTDOSE (REF=LAST) sex2 (ref="M")
	race4 (ref='WHITE')  ethnicity3
	(ref='NOT HISPANIC') Vaccinationsite (ref="PUBLIX OR CVS OR
WALGREENS")/param=reference;
	model NONCOVIDDEATHPLUS1YRD1(ref='0')= FIRSTDOSE agenew sex2 race4
ethnicity3
				decile_sviscore1 decile_sviscore4
				mortalityratepct Vaccinationsite/parmlabel;
	by agenew;
	format agenew age3groups.;
run;
ods output close;
```

```
ods pdf close;

title;

proc print data=Log_ageNONCOV2adjusted3g;

run;

data Log_ageNONCOV2adjusted3g1;

	set Log_ageNONCOV2adjusted3g;

	ODDSRatio1=put(OddsRatioEst,8.3);

	LowerCL1=put(LowerCL,8.3);

	UpperCL1=put(UpperCL,8.3);

	OR=STRIP(ODDSRatio1)||" ("||STRIP(LowerCL1)||", "||STRIP(UpperCL1)||')';

	MODEL=2;

run;

proc print data=Log_ageNONCOV2adjusted3g1;

	where SCAN(upcase(EFFECT),1)='FIRSTDOSE';

run;

/*** output excel file for results ***/

libname results xlsx "&path\SAS output\&textadd\NON COVID PLUS\NON COVID
PLUS.xlsx";

/* Export first dataset */

data results.sheet1;

   set work.logNONCOV2;
```

```
            where SCAN(upcase(EFFECT),1)='FIRSTDOSE';
run;

/* Export third dataset */
data results.sheet3;
   set work.Log_ageNONCOV2adjusted3g1;
            where SCAN(upcase(EFFECT),1)='FIRSTDOSE';
run;

libname results clear;

/*** 1.a. COVID +  with categorical age ***/

ods pdf file="&path\SAS output\&textadd\COVID PLUS\
&textadd Logistic COVID + &todaysdate..pdf";
ods output ODDSRATIOS=log_COV2crude;
proc logistic data=&dataset;
            title "&textadd COVID + deaths" ;
            class firstdose (REF=last)/param=reference;
            model COVIDDEATHPLUS1YRD1(ref='0')=firstdose/ parmlabel;
run;
ODS OUTPUT CLOSE;
ods pdf close;
title;
```

```
proc print;
run;

data Log_COV2crude1;
	set Log_COV2crude;
	ODDSRatio1=put(OddsRatioEst,8.3);
	LowerCL1=put(LowerCL,8.3);
	UpperCL1=put(UpperCL,8.3);
	OR=STRIP(ODDSRatio1)||" ("||STRIP(LowerCL1)||", "||STRIP(UpperCL1)||')';
	MODEL=1;
run;

proc print;
run;

ods pdf file="&path\SAS output\&textadd\COVID PLUS\
&textadd Logistic COVID + adjusted model (cat age) &todaysdate..pdf";
ods output ODDSRATIOS=Log_COV2adjusted;
proc logistic data=&dataset;
	TITLE 'COVID + death adjusted (logistic regression)';
	class agenew (ref=first) firstdose (REF=last) sex2 (ref="M")
	race4 (ref='WHITE')  ethnicity3
	(ref='NOT HISPANIC') Vaccinationsite (ref="PUBLIX OR CVS OR
WALGREENS")/param=reference;
	model COVIDDEATHPLUS1YRD1(ref='0')= firstdose agenew sex2 race4 ethnicity3
```

```
                    decile_sviscore1 decile_sviscore4
                    mortalityratepct Vaccinationsite/parmlabel;
            format agenew AGE10yGROUP.;
run;
ods output close;
ods pdf close;
title;

proc print;
run;

data Log_COV2adjusted1;
    set Log_COV2adjusted;
    ODDSRatio1=put(OddsRatioEst,8.3);
    LowerCL1=put(LowerCL,8.3);
    UpperCL1=put(UpperCL,8.3);
    OR=STRIP(ODDSRatio1)||" ("||STRIP(LowerCL1)||", "||STRIP(UpperCL1)||')';
    MODEL=2;
run;

proc print data=Log_COV2adjusted1;
    where SCAN(upcase(EFFECT),1)='VACCINEGROUP2';
run;

DATA LOGCOV2;
    SET lOG_COV2CRUDE1 lOG_COV2adjusted1;
```

```
RUN;

PROC PRINT data= LOGCOV2;
	where SCAN(upcase(EFFECT),1)='FIRSTDOSE';
RUN;

proc sort data=&dataset;
	by agenew;
run;

ods pdf file="&path\SAS output\&textadd\COVID PLUS\
&textadd logistic COVID + adjusted model (cat age) by Age (3 Age groups)
&todaysdate..pdf";
ods output ODDSRATIOS=Log_ageCOV2adjusted3g;
proc logistic data=&dataset;
	title 'COVID + deaths adjusted by Age';
	class agenew (ref=first) FIRSTDOSE (REF=LAST) sex2 (ref="M")
	race4 (ref='WHITE')  ethnicity3
	(ref='NOT HISPANIC') Vaccinationsite (ref="PUBLIX OR CVS OR
WALGREENS")/param=reference;
	model COVIDDEATHPLUS1YRD1(ref='0')= FIRSTDOSE agenew sex2 race4 ethnicity3
			decile_sviscore1 decile_sviscore4
			mortalityratepct Vaccinationsite/parmlabel;
	by agenew;
	format agenew age3groups.;
run;
ods output close;
```

```
ods pdf close;
title;

proc print data=Log_ageCOV2adjusted3g;
run;

data Log_ageCOV2adjusted3g1;
	set Log_ageCOV2adjusted3g;
	ODDSRatio1=put(OddsRatioEst,8.3);
	LowerCL1=put(LowerCL,8.3);
	UpperCL1=put(UpperCL,8.3);
	OR=STRIP(ODDSRatio1)||" ("||STRIP(LowerCL1)||", "||STRIP(UpperCL1)||')';
	MODEL=2;
run;

proc print data=Log_ageCOV2adjusted3g1;
	where SCAN(upcase(EFFECT),1)='FIRSTDOSE';
run;

/*** output excel file for results ***/
libname results xlsx "&path\SAS output\&textadd\COVID PLUS\COVID PLUS.xlsx";

/* Export first dataset */
data results.sheet1;
   set work.logCOV2;
	where SCAN(upcase(EFFECT),1)='FIRSTDOSE';
```

```
run;

/* Export third dataset */
data results.sheet3;
   set work.Log_ageCOV2adjusted3g1;
        where SCAN(upcase(EFFECT),1)='FIRSTDOSE';
run;

libname results clear;

/*** 1.a. All deaths with categorical age ***/

ods pdf file="&path\SAS output\&textadd\All cause\
&textadd Logistic All-cause &todaysdate..pdf";
ods output ODDSRATIOS=log_allcrude;
proc logistic data=&dataset;
        title "&textadd All-cause deaths" ;
        class firstdose (REF=last)/param=reference;
        model DEATH1YRd1(ref='0')=firstdose/ parmlabel;
run;
ODS OUTPUT CLOSE;
ods pdf close;
title;

proc print;
run;
```

```
data Log_allcrude1;
	set Log_allcrude;
	ODDSRatio1=put(OddsRatioEst,8.3);
	LowerCL1=put(LowerCL,8.3);
	UpperCL1=put(UpperCL,8.3);
	OR=STRIP(ODDSRatio1)||" ("||STRIP(LowerCL1)||", "||STRIP(UpperCL1)||')';
	MODEL=1;
run;

proc print;
run;

ods pdf file="&path\SAS output\&textadd\All cause\
&textadd Logistic All-cause adjusted model (cat age) &todaysdate..pdf";
ods output ODDSRATIOS=Log_alladjusted;
proc logistic data=&dataset;
	TITLE 'All-cause death adjusted (logistic regression)';
	class agenew (ref=first) firstdose (REF=last) sex2 (ref="M")
	race4 (ref='WHITE')  ethnicity3
	(ref='NOT HISPANIC') Vaccinationsite (ref="PUBLIX OR CVS OR
WALGREENS")/param=reference;
	model DEATH1YRd1(ref='0')= firstdose agenew sex2 race4 ethnicity3
				decile_sviscore1 decile_sviscore4
				mortalityratepct Vaccinationsite/parmlabel;
```

```
                    format agenew AGE10yGROUP.;
run;
ods output close;
ods pdf close;
title;

proc print;
run;

data Log_alladjusted1;
          set log_alladjusted;
          ODDSRatio1=put(OddsRatioEst,8.3);
          LowerCL1=put(LowerCL,8.3);
          UpperCL1=put(UpperCL,8.3);
          OR=STRIP(ODDSRatio1)||" ("||STRIP(LowerCL1)||", "||STRIP(UpperCL1)||')';
          MODEL=2;
run;

proc print data=Log_alladjusted1;
          where SCAN(upcase(EFFECT),1)='VACCINEGROUP2';
run;

DATA LOGALL;
          SET lOG_ALLCRUDE1 lOG_alladjusted1;
RUN;
```

```
PROC PRINT data= logall;
	where SCAN(upcase(EFFECT),1)='FIRSTDOSE';
RUN;

proc sort data=&dataset;
	by agenew;
run;

ods pdf file="&path\SAS output\&textadd\All cause\
&textadd logistic All-cause adjusted model (cat age) by Age (3 Age groups) &todaysdate..pdf";
ods output ODDSRATIOS=Log_agealladjusted3g;
proc logistic data=&dataset;
	title 'All-cause deaths adjusted by Age';
	class agenew (ref=first) FIRSTDOSE (REF=LAST) sex2 (ref="M")
	race4 (ref='WHITE')  ethnicity3
	(ref='NOT HISPANIC') Vaccinationsite (ref="PUBLIX OR CVS OR WALGREENS")/param=reference;
	model DEATH1YRd1(ref='0')= FIRSTDOSE agenew sex2 race4 ethnicity3
				decile_sviscore1 decile_sviscore4
				mortalityratepct Vaccinationsite/parmlabel;
	by agenew;
	format agenew age3groups.;
run;
ods output close;
ods pdf close;
title;
```

```
proc print data=Log_agealladjusted3g;
run;

data Log_agealladjusted3g1;
	set Log_agealladjusted3g;
	ODDSRatio1=put(OddsRatioEst,8.3);
	LowerCL1=put(LowerCL,8.3);
	UpperCL1=put(UpperCL,8.3);
	OR=STRIP(ODDSRatio1)||" ("||STRIP(LowerCL1)||", "||STRIP(UpperCL1)||')';
	MODEL=2;
run;

proc print data=Log_agealladjusted3g1;
	where SCAN(upcase(EFFECT),1)='FIRSTDOSE';
run;

/*** output excel file for results ***/
libname results xlsx "&path\SAS output\&textadd\All cause\allcause.xlsx";

/* Export first dataset */
data results.sheet1;
   set work.logall;
	where SCAN(upcase(EFFECT),1)='FIRSTDOSE';
run;
```

```
/* Export third dataset */
data results.sheet3;
   set work.log_agealladjusted3g1;
           where SCAN(upcase(EFFECT),1)='FIRSTDOSE';
run;

libname results clear;

/*** 1.a. CVD deaths with categorical age ***/

ods pdf file="&path\SAS output\&textadd\CVD DEATHS\
&textadd Logistic CVD DEATHS &todaysdate..pdf";
ods output ODDSRATIOS=log_cvdcrude;
proc logistic data=&dataset;
           title "&textadd CVD deaths" ;
           class firstdose (REF=last)/param=reference;
           model CVD1YRD1(ref='0')=firstdose/ parmlabel;
run;
ODS OUTPUT CLOSE;
ods pdf close;
title;

proc print;
run;

data log_cvdcrude1;
```

```
        set log_cvdcrude;
        ODDSRatio1=put(OddsRatioEst,8.3);
        LowerCL1=put(LowerCL,8.3);
        UpperCL1=put(UpperCL,8.3);
        OR=STRIP(ODDSRatio1)||" ("||STRIP(LowerCL1)||", "||STRIP(UpperCL1)||')';
        MODEL=1;
run;

proc print;
run;

ods pdf file="&path\SAS output\&textadd\CVD DEATHS\
&textadd Logistic CVD Deaths adjusted model (cat age) &todaysdate..pdf";
ods output ODDSRATIOS=Log_cvdadjusted;
proc logistic data=&dataset;
        TITLE 'CVD Death adjusted (logistic regression)';
        class agenew (ref=first) firstdose (REF=last) sex2 (ref="M")
        race4 (ref='WHITE')  ethnicity3
        (ref='NOT HISPANIC') Vaccinationsite (ref="PUBLIX OR CVS OR
WALGREENS")/param=reference;
        model CVD1YRD1(ref='0')= firstdose agenew sex2 race4 ethnicity3
                         decile_sviscore1 decile_sviscore4
                        mortalityratepct Vaccinationsite/parmlabel;
                format agenew AGE10yGROUP.;
run;
```

```
ods output close;

ods pdf close;

title;

proc print;

run;

data Log_cvdadjusted1;

	set Log_cvdadjusted;

	ODDSRatio1=put(OddsRatioEst,8.3);

	LowerCL1=put(LowerCL,8.3);

	UpperCL1=put(UpperCL,8.3);

	OR=STRIP(ODDSRatio1)||" ("||STRIP(LowerCL1)||", "||STRIP(UpperCL1)||')';

	MODEL=2;

run;

proc print data=Log_cvdadjusted1;

	where SCAN(upcase(EFFECT),1)='FIRSTDOSE';

run;

DATA LOGCVD;

	SET log_cvdcrude1 Log_cvdadjusted1;

RUN;

PROC PRINT data= LOGCVD;

	where SCAN(upcase(EFFECT),1)='FIRSTDOSE';
```

```
RUN;

proc sort data=&dataset;
	by agenew;
run;

ods pdf file="&path\SAS output\&textadd\CVD DEATHS\
&textadd logistic CVD DEATHS adjusted model (cat age) by Age (3 Age groups) &todaysdate..pdf";
ods output ODDSRATIOS=Log_agecvdadjusted3g;
proc logistic data=&dataset;
	title 'CVD deaths adjusted by Age';
	class agenew (ref=first) FIRSTDOSE (REF=LAST) sex2 (ref="M")
	race4 (ref='WHITE')  ethnicity3
	(ref='NOT HISPANIC') Vaccinationsite (ref="PUBLIX OR CVS OR WALGREENS")/param=reference;
	model CVD1YRD1(ref='0')= FIRSTDOSE agenew sex2 race4 ethnicity3
				decile_sviscore1 decile_sviscore4
				mortalityratepct Vaccinationsite/parmlabel;
	by agenew;
	format agenew age3groups.;
run;
ods output close;
ods pdf close;
title;

proc print data=Log_agecvdadjusted3g;
```

```
run;

data Log_agecvdadjusted3g1;
	set Log_agecvdadjusted3g;
	ODDSRatio1=put(OddsRatioEst,8.3);
	LowerCL1=put(LowerCL,8.3);
	UpperCL1=put(UpperCL,8.3);
	OR=STRIP(ODDSRatio1)||" ("||STRIP(LowerCL1)||", "||STRIP(UpperCL1)||')';
	MODEL=2;
run;

proc print data=Log_agecvdadjusted3g1;
	where SCAN(upcase(EFFECT),1)='FIRSTDOSE';
run;

/*** output excel file for results ***/
libname results xlsx "&path\SAS output\&textadd\CVD DEATHS\CVD DEATHS.xlsx";

/* Export first dataset */
data results.sheet1;
  set work.logcvd;
	where SCAN(upcase(EFFECT),1)='FIRSTDOSE';
run;

/* Export third dataset */
data results.sheet3;
```

```
   set work.log_ageCVDadjusted3g1;
        where SCAN(upcase(EFFECT),1)='FIRSTDOSE';
run;

libname results clear;

/*** COVID DEATHS ***/

/*** 1.a. COVID deaths with categorical age ***/

ods pdf file="&path\SAS output\&textadd\COVID DEATHS\
&textadd Logistic COVID DEATHS &todaysdate..pdf";
ods output ODDSRATIOS=log_COVIDcrude;
proc logistic data=&dataset;
        title "&textadd COVID deaths" ;
        class firstdose (REF=last)/param=reference;
        model COVIDDEATH1YRD1(ref='0')=firstdose/ parmlabel;
run;
ODS OUTPUT CLOSE;
ods pdf close;
title;

proc print;
run;
```

```
data log_COVIDcrude1;
	set log_COVIDcrude;
	ODDSRatio1=put(OddsRatioEst,8.3);
	LowerCL1=put(LowerCL,8.3);
	UpperCL1=put(UpperCL,8.3);
	OR=STRIP(ODDSRatio1)||" ("||STRIP(LowerCL1)||", "||STRIP(UpperCL1)||')';
	MODEL=1;
run;

proc print;
run;

ods pdf file="&path\SAS output\&textadd\COVID DEATHS\
&textadd Logistic COVID Deaths adjusted model (cat age) &todaysdate..pdf";
ods output ODDSRATIOS=Log_COVIDadjusted;
proc logistic data=&dataset;
	TITLE 'COVID Death adjusted (logistic regression)';
	class agenew (ref=first) firstdose (REF=last) sex2 (ref="M")
	race4 (ref='WHITE')  ethnicity3
	(ref='NOT HISPANIC') Vaccinationsite (ref="PUBLIX OR CVS OR
WALGREENS")/param=reference;
	model COVIDDEATH1YRD1(ref='0')= firstdose agenew sex2 race4 ethnicity3
				decile_sviscore1 decile_sviscore4
				mortalityratepct Vaccinationsite/parmlabel;
		format agenew AGE10yGROUP.;
```

```
run;
ods output close;
ods pdf close;
title;

proc print;
run;

data Log_COVIDadjusted1;
	set Log_COVIDadjusted;
	ODDSRatio1=put(OddsRatioEst,8.3);
	LowerCL1=put(LowerCL,8.3);
	UpperCL1=put(UpperCL,8.3);
	OR=STRIP(ODDSRatio1)||" ("||STRIP(LowerCL1)||", "||STRIP(UpperCL1)||')';
	MODEL=2;
run;

proc print data=Log_COVIDadjusted1;
	where SCAN(upcase(EFFECT),1)='FIRSTDOSE';
run;

DATA LOGcovid;
	SET log_covidcrude1 Log_covidadjusted1;
RUN;

PROC PRINT data= LOGcovid;
```

```
        where SCAN(upcase(EFFECT),1)='FIRSTDOSE';
RUN;

proc sort data=&dataset;
        by agenew;
run;

ods pdf file="&path\SAS output\&textadd\COVID DEATHS\
&textadd logistic COVID DEATHS adjusted model (cat age) by Age (3 Age groups)
&todaysdate..pdf";
ods output ODDSRATIOS=Log_ageCOVIDadjusted3g;
proc logistic data=&dataset;
        title 'COVID deaths adjusted by Age';
        class agenew (ref=first) FIRSTDOSE (REF=LAST) sex2 (ref="M")
        race4 (ref='WHITE')  ethnicity3
        (ref='NOT HISPANIC') Vaccinationsite (ref="PUBLIX OR CVS OR
WALGREENS")/param=reference;
        model COVIDDEATH1YRD1(ref='0')= FIRSTDOSE agenew sex2 race4 ethnicity3
                        decile_sviscore1 decile_sviscore4
                        mortalityratepct Vaccinationsite/parmlabel;
        by agenew;
        format agenew age3groups.;
run;
ods output close;
ods pdf close;
title;
```

```
proc print data=Log_ageCOVIDadjusted3g;
run;

data Log_ageCOVIDadjusted3g1;
	set Log_ageCOVIDadjusted3g;
	ODDSRatio1=put(OddsRatioEst,8.3);
	LowerCL1=put(LowerCL,8.3);
	UpperCL1=put(UpperCL,8.3);
	OR=STRIP(ODDSRatio1)||" ("||STRIP(LowerCL1)||", "||STRIP(UpperCL1)||')';
	MODEL=2;
run;

proc print data=Log_ageCOVIDadjusted3g1;
	where SCAN(upcase(EFFECT),1)='FIRSTDOSE';
run;

/*** output excel file for results ***/
libname results xlsx "&path\SAS output\&textadd\COVID DEATHS\COVID DEATHS.xlsx";

/* Export first dataset */
data results.sheet1;
   set work.logCOVID;
	where SCAN(upcase(EFFECT),1)='FIRSTDOSE';
run;

/* Export third dataset */
```

```
data results.sheet3;
   set work.log_ageCOVIDadjusted3g1;
        where SCAN(upcase(EFFECT),1)='FIRSTDOSE';
run;

libname results clear;

/*** NON COVID DEATHS ***/

ods pdf file="&path\SAS output\&textadd\NON COVID DEATHS\
&textadd Logistic NON COVID DEATHS &todaysdate..pdf";
ods output ODDSRATIOS=log_NONCOVIDcrude;
proc logistic data=&dataset;
        title "&textadd NON COVID deaths" ;
        class firstdose (REF=last)/param=reference;
        model NONCOVIDDEATH1YRD1(ref='0')=firstdose/ parmlabel;
run;
ODS OUTPUT CLOSE;
ods pdf close;
title;

proc print;
run;
```

```
data log_nonCOVIDcrude1;
	set log_NONCOVIDcrude;
	ODDSRatio1=put(OddsRatioEst,8.3);
	LowerCL1=put(LowerCL,8.3);
	UpperCL1=put(UpperCL,8.3);
	OR=STRIP(ODDSRatio1)||" ("||STRIP(LowerCL1)||", "||STRIP(UpperCL1)||')';
	MODEL=1;
run;

proc print;
run;

ods pdf file="&path\SAS output\&textadd\NON COVID DEATHS\
&textadd Logistic COVID Deaths adjusted model (cat age) &todaysdate..pdf";
ods output ODDSRATIOS=Log_NONCOVIDadjusted;
proc logistic data=&dataset;
	TITLE 'NON COVID Death adjusted (logistic regression)';
	class agenew (ref=first) firstdose (REF=last) sex2 (ref="M")
	race4 (ref='WHITE')  ethnicity3
	(ref='NOT HISPANIC') Vaccinationsite (ref="PUBLIX OR CVS OR
WALGREENS")/param=reference;
	model NONCOVIDDEATH1YRD1(ref='0')= firstdose agenew sex2 race4 ethnicity3
	                 decile_sviscore1 decile_sviscore4
	                mortalityratepct Vaccinationsite/parmlabel;
```

```
                    format agenew AGE10yGROUP.;
run;
ods output close;
ods pdf close;
title;

proc print;
run;

data Log_NONCOVIDadjusted1;
          set Log_NONCOVIDadjusted;
          ODDSRatio1=put(OddsRatioEst,8.3);
          LowerCL1=put(LowerCL,8.3);
          UpperCL1=put(UpperCL,8.3);
          OR=STRIP(ODDSRatio1)||" ("||STRIP(LowerCL1)||", "||STRIP(UpperCL1)||')';
          MODEL=2;
run;

proc print data=Log_NONCOVIDadjusted1;
          where SCAN(upcase(EFFECT),1)='FIRSTDOSE';
run;

DATA LOGNONcovid;
          SET log_NONcovidcrude1 Log_NONcovidadjusted1;
RUN;
```

```
PROC PRINT data= LOGNONcovid;
	where SCAN(upcase(EFFECT),1)='FIRSTDOSE';
RUN;

proc sort data=&dataset;
	by agenew;
run;

ods pdf file="&path\SAS output\&textadd\NON COVID DEATHS\
&textadd logistic Non COVID DEATHS adjusted model (cat age) by Age (3 Age groups) &todaysdate..pdf";
ods output ODDSRATIOS=Log_ageNONCOVIDadjusted3g;
proc logistic data=&dataset;
	title 'NON COVID deaths adjusted by Age';
	class agenew (ref=first) FIRSTDOSE (REF=LAST) sex2 (ref="M")
	race4 (ref='WHITE')  ethnicity3
	(ref='NOT HISPANIC') Vaccinationsite (ref="PUBLIX OR CVS OR WALGREENS")/param=reference;
	model NONCOVIDDEATH1YRD1(ref='0')= FIRSTDOSE agenew sex2 race4 ethnicity3
				decile_sviscore1 decile_sviscore4
				mortalityratepct Vaccinationsite/parmlabel;
	by agenew;
	format agenew age3groups.;
run;
ods output close;
ods pdf close;
title;
```

```
proc print data=Log_ageNONCOVIDadjusted3g;
run;

data Log_ageNONCOVIDadjusted3g1;
	set Log_ageNONCOVIDadjusted3g;
	ODDSRatio1=put(OddsRatioEst,8.3);
	LowerCL1=put(LowerCL,8.3);
	UpperCL1=put(UpperCL,8.3);
	OR=STRIP(ODDSRatio1)||" ("||STRIP(LowerCL1)||", "||STRIP(UpperCL1)||')';
	MODEL=2;
run;

proc print data=Log_ageNONCOVIDadjusted3g1;
	where SCAN(upcase(EFFECT),1)='FIRSTDOSE';
run;

/*** output excel file for results ***/
libname results xlsx "&path\SAS output\&textadd\NON COVID DEATHS\NON COVID DEATHS.xlsx";

/* Export first dataset */
data results.sheet1;
   set work.logNONCOVID;
	where SCAN(upcase(EFFECT),1)='FIRSTDOSE';
run;
```

```
/* Export third dataset */

data results.sheet3;

   set work.log_ageNONCOVIDadjusted3g1;

            where SCAN(upcase(EFFECT),1)='FIRSTDOSE';

run;

libname results clear;

/*** FULL COHORT                ***/

%let dataset=dose1matcheddatafinal;

%let todaysdate=%sysfunc(today(),date9.);

%put &todaysdate;

%let textadd=Matched cohort Risk of death Dose 1;

%let path=H:\EPI\Surveillance Section\Fahad Mansuri\SURVIVAL ANALYSIS 2024;
```

```
proc contents data=work.dose1matcheddatafinal;
run;

proc freq data=work.dose1matcheddatafinal;
	table firstdose;
run;

/*** 1.a. NON COVID +  with categorical age ***/

ods pdf file="&path\SAS output\&textadd\NON COVID PLUS FULL COHORT\
&textadd Logistic COVID + &todaysdate..pdf";
ods output ODDSRATIOS=log_NONCOV2crude;
proc logistic data=&dataset;
	title "&textadd NON COVID + deaths" ;
	class firstdose (REF=last)/param=reference;
	model NONCOVIDDEATHPLUS1YRD1(ref='0')=firstdose/ parmlabel;
run;
ODS OUTPUT CLOSE;
ods pdf close;
title;

proc print;
run;

data Log_NONCOV2crude1;
	set Log_NONCOV2crude;
```

```
        ODDSRatio1=put(OddsRatioEst,8.3);
        LowerCL1=put(LowerCL,8.3);
        UpperCL1=put(UpperCL,8.3);
        OR=STRIP(ODDSRatio1)||" ("||STRIP(LowerCL1)||", "||STRIP(UpperCL1)||')';
        MODEL=1;
run;

proc print;
run;

ods pdf file="&path\SAS output\&textadd\NON COVID PLUS FULL COHORT\
&textadd Logistic NON COVID + adjusted model (cat age) &todaysdate..pdf";
ods output ODDSRATIOS=Log_NONCOV2adjusted;
proc logistic data=&dataset;
        TITLE 'NON COVID + death adjusted (logistic regression)';
        class agenew (ref=first) firstdose (REF=last) sex2 (ref="M")
        race4 (ref='WHITE')  ethnicity3
        (ref='NOT HISPANIC') Vaccinationsite (ref="PUBLIX OR CVS OR
WALGREENS")/param=reference;
        model NONCOVIDDEATHPLUS1YRD1(ref='0')= firstdose agenew sex2 race4
ethnicity3
                         decile_sviscore1 decile_sviscore4
                        mortalityratepct Vaccinationsite/parmlabel;
                format agenew AGE10yGROUP.;
run;
ods output close;
```

```
ods pdf close;

title;

proc print;

run;

data Log_NONCOV2adjusted1;

	set Log_NONCOV2adjusted;

	ODDSRatio1=put(OddsRatioEst,8.3);

	LowerCL1=put(LowerCL,8.3);

	UpperCL1=put(UpperCL,8.3);

	OR=STRIP(ODDSRatio1)||" ("||STRIP(LowerCL1)||", "||STRIP(UpperCL1)||')';

	MODEL=2;

run;

proc print data=Log_NONCOV2adjusted1;

	where SCAN(upcase(EFFECT),1)='VACCINEGROUP2';

run;

DATA LOGNONCOV2;

	SET lOG_NONCOV2CRUDE1 lOG_NONCOV2adjusted1;

RUN;

PROC PRINT data= LOGNONCOV2;

	where SCAN(upcase(EFFECT),1)='FIRSTDOSE';

RUN;
```

```
proc sort data=&dataset;
	by agenew;
run;

ods pdf file="&path\SAS output\&textadd\NON COVID PLUS FULL COHORT\
&textadd logistic NON COVID + adjusted model (cat age) by Age (3 Age groups) &todaysdate..pdf";
ods output ODDSRATIOS=Log_ageNONCOV2adjusted3g;
proc logistic data=&dataset;
	title 'NON COVID + deaths adjusted by Age';
	class agenew (ref=first) FIRSTDOSE (REF=LAST) sex2 (ref="M")
	race4 (ref='WHITE')  ethnicity3
	(ref='NOT HISPANIC') Vaccinationsite (ref="PUBLIX OR CVS OR WALGREENS")/param=reference;
	model NONCOVIDDEATHPLUS1YRD1(ref='0')= FIRSTDOSE agenew sex2 race4 ethnicity3
				decile_sviscore1 decile_sviscore4
				mortalityratepct Vaccinationsite/parmlabel;
	by agenew;
	format agenew age3groups.;
run;
ods output close;
ods pdf close;
title;

proc print data=Log_ageNONCOV2adjusted3g;
```

```
run;

data Log_ageNONCOV2adjusted3g1;
	set Log_ageNONCOV2adjusted3g;
	ODDSRatio1=put(OddsRatioEst,8.3);
	LowerCL1=put(LowerCL,8.3);
	UpperCL1=put(UpperCL,8.3);
	OR=STRIP(ODDSRatio1)||" ("||STRIP(LowerCL1)||", "||STRIP(UpperCL1)||')';
	MODEL=2;
run;

proc print data=Log_ageNONCOV2adjusted3g1;
	where SCAN(upcase(EFFECT),1)='FIRSTDOSE';
run;

/*** output excel file for results ***/
libname results xlsx "&path\SAS output\&textadd\NON COVID PLUS FULL COHORT\NON COVID PLUS FULL COHORT.xlsx";

/* Export first dataset */
data results.sheet1;
   set work.logNONCOV2;
	where SCAN(upcase(EFFECT),1)='FIRSTDOSE';
run;

/* Export third dataset */
```

```
data results.sheet3;
   set work.Log_ageNONCOV2adjusted3g1;
        where SCAN(upcase(EFFECT),1)='FIRSTDOSE';
run;

libname results clear;

/*** 1.a. SUIC or HOMICIDE  with categorical age ***/

ods pdf file="&path\SAS output\&textadd\SUICIDE or HOMICIDE FULL COHORT\
&textadd Logistic SUICHOM &todaysdate..pdf";
ods output ODDSRATIOS=log_SUICHOMcrude;
proc logistic data=&dataset;
        title "&textadd SUICIDE OR HOMICIDE deaths" ;
        class firstdose (REF=last)/param=reference;
        model suicidehomicide1yrD1(ref='0')=firstdose/ parmlabel;
run;
ODS OUTPUT CLOSE;
ods pdf close;
title;

proc print;
run;

data log_SUICHOMcrude1;
        set log_SUICHOMcrude;
```

```
        ODDSRatio1=put(OddsRatioEst,8.3);
        LowerCL1=put(LowerCL,8.3);
        UpperCL1=put(UpperCL,8.3);
        OR=STRIP(ODDSRatio1)||" ("||STRIP(LowerCL1)||", "||STRIP(UpperCL1)||')';
        MODEL=1;
run;

proc print;
run;

ods pdf file="&path\SAS output\&textadd\SUICIDE OR HOMICIDE FULL COHORT\
&textadd Logistic SUICHOM adjusted model (cat age) &todaysdate..pdf";
ods output ODDSRATIOS=Log_SUICHOMadjusted;
proc logistic data=&dataset;
        TITLE 'SUICIDE OR HOMICIDE death adjusted (logistic regression)';
        class agenew (ref=first) firstdose (REF=last) sex2 (ref="M")
        race4 (ref='WHITE')  ethnicity3
        (ref='NOT HISPANIC') Vaccinationsite (ref="PUBLIX OR CVS OR
WALGREENS")/param=reference;
        model suicidehomicide1yrD1(ref='0')= firstdose agenew sex2 race4 ethnicity3
                        decile_sviscore1 decile_sviscore4
                       mortalityratepct Vaccinationsite/parmlabel;
                format agenew AGE10yGROUP.;
run;
ods output close;
```

```
ods pdf close;

title;

proc print;

run;

data Log_SUICHOMadjusted1;

	set Log_SUICHOMadjusted;

	ODDSRatio1=put(OddsRatioEst,8.3);

	LowerCL1=put(LowerCL,8.3);

	UpperCL1=put(UpperCL,8.3);

	OR=STRIP(ODDSRatio1)||" ("||STRIP(LowerCL1)||", "||STRIP(UpperCL1)||')';

	MODEL=2;

run;

proc print data=Log_SUICHOMadjusted1;

	where SCAN(upcase(EFFECT),1)='FIRSTDOSE';

run;

DATA LOGSUICHOM;

	SET lOG_SUICHOMCRUDE1 Log_SUICHOMadjusted1;

RUN;

PROC PRINT data= LOGSUICHOM;

	where SCAN(upcase(EFFECT),1)='FIRSTDOSE';

RUN;
```

```
proc sort data=&dataset;
	by agenew;
run;

ods pdf file="&path\SAS output\&textadd\SUICIDE OR HOMICIDE FULL COHORT\
&textadd logistic SUICHOM adjusted model (cat age) by Age (3 Age groups)
&todaysdate..pdf";
ods output ODDSRATIOS=Log_ageSUICHOMadjusted3g;
proc logistic data=&dataset;
	title 'SUICIDE OR HOMICIDE deaths adjusted by Age';
	class agenew (ref=first) FIRSTDOSE (REF=LAST) sex2 (ref="M")
	race4 (ref='WHITE')  ethnicity3
	(ref='NOT HISPANIC') Vaccinationsite (ref="PUBLIX OR CVS OR
WALGREENS")/param=reference;
	model suicidehomicide1yrD1(ref='0')= FIRSTDOSE agenew sex2 race4 ethnicity3
				decile_sviscore1 decile_sviscore4
				mortalityratepct Vaccinationsite/parmlabel;
	by agenew;
	format agenew age3groups.;
run;
ods output close;
ods pdf close;
title;

proc print data=Log_ageSUICHOMadjusted3g;
run;
```

```
data Log_ageSUICHOMadjusted3g1;
	set Log_ageSUICHOMadjusted3g;
	ODDSRatio1=put(OddsRatioEst,8.3);
	LowerCL1=put(LowerCL,8.3);
	UpperCL1=put(UpperCL,8.3);
	OR=STRIP(ODDSRatio1)||" ("||STRIP(LowerCL1)||", "||STRIP(UpperCL1)||')';
	MODEL=2;
run;

proc print data=Log_ageSUICHOMadjusted3g1;
	where SCAN(upcase(EFFECT),1)='FIRSTDOSE';
run;

/*** output excel file for results ***/
libname results xlsx "&path\SAS output\&textadd\SUICIDE OR HOMICIDE FULL COHORT\
SUICIDE OR HOMICIDE FULL COHORT.xlsx";

/* Export first dataset */
data results.sheet1;
   set work.logSUICHOM;
	where SCAN(upcase(EFFECT),1)='FIRSTDOSE';
run;

/* Export third dataset */
data results.sheet3;
```

```
   set work.Log_ageSUICHOMadjusted3g1;
         where SCAN(upcase(EFFECT),1)='FIRSTDOSE';
run;

libname results clear;

/*** 1.a. COVID +  with categorical age ***/

ods pdf file="&path\SAS output\&textadd\COVID PLUS FULL COHORT\
&textadd Logistic COVID + &todaysdate..pdf";
ods output ODDSRATIOS=log_COV2crude;
proc logistic data=&dataset;
         title "&textadd COVID + deaths" ;
         class firstdose (REF=last)/param=reference;
         model COVIDDEATHPLUS1YRD1(ref='0')=firstdose/ parmlabel;
run;
ODS OUTPUT CLOSE;
ods pdf close;
title;

proc print;
run;

data Log_COV2crude1;
         set Log_COV2crude;
```

```
            ODDSRatio1=put(OddsRatioEst,8.3);
            LowerCL1=put(LowerCL,8.3);
            UpperCL1=put(UpperCL,8.3);
            OR=STRIP(ODDSRatio1)||" ("||STRIP(LowerCL1)||", "||STRIP(UpperCL1)||')';
            MODEL=1;
run;

proc print;
run;

ods pdf file="&path\SAS output\&textadd\COVID PLUS FULL COHORT\
&textadd Logistic COVID + adjusted model (cat age) &todaysdate..pdf";
ods output ODDSRATIOS=Log_COV2adjusted;
proc logistic data=&dataset;
            TITLE 'COVID + death adjusted (logistic regression)';
            class agenew (ref=first) firstdose (REF=last) sex2 (ref="M")
            race4 (ref='WHITE')  ethnicity3
            (ref='NOT HISPANIC') Vaccinationsite (ref="PUBLIX OR CVS OR
WALGREENS")/param=reference;
            model COVIDDEATHPLUS1YRD1(ref='0')= firstdose agenew sex2 race4 ethnicity3
                              decile_sviscore1 decile_sviscore4
                             mortalityratepct Vaccinationsite/parmlabel;
                  format agenew AGE10yGROUP.;
run;
ods output close;
```

```
ods pdf close;

title;

proc print;

run;

data Log_COV2adjusted1;

	set Log_COV2adjusted;

	ODDSRatio1=put(OddsRatioEst,8.3);

	LowerCL1=put(LowerCL,8.3);

	UpperCL1=put(UpperCL,8.3);

	OR=STRIP(ODDSRatio1)||" ("||STRIP(LowerCL1)||", "||STRIP(UpperCL1)||')';

	MODEL=2;

run;

proc print data=Log_COV2adjusted1;

	where SCAN(upcase(EFFECT),1)='VACCINEGROUP2';

run;

DATA LOGCOV2;

	SET lOG_COV2CRUDE1 lOG_COV2adjusted1;

RUN;

PROC PRINT data= LOGCOV2;

	where SCAN(upcase(EFFECT),1)='FIRSTDOSE';

RUN;
```

```
proc sort data=&dataset;
	by agenew;
run;

ods pdf file="&path\SAS output\&textadd\COVID PLUS FULL COHORT\
&textadd logistic COVID + adjusted model (cat age) by Age (3 Age groups) &todaysdate..pdf";
ods output ODDSRATIOS=Log_ageCOV2adjusted3g;
proc logistic data=&dataset;
	title 'COVID + deaths adjusted by Age';
	class agenew (ref=first) FIRSTDOSE (REF=LAST) sex2 (ref="M")
	race4 (ref='WHITE')  ethnicity3
	(ref='NOT HISPANIC') Vaccinationsite (ref="PUBLIX OR CVS OR WALGREENS")/param=reference;
	model COVIDDEATHPLUS1YRD1(ref='0')= FIRSTDOSE agenew sex2 race4 ethnicity3
			decile_sviscore1 decile_sviscore4
			mortalityratepct Vaccinationsite/parmlabel;
	by agenew;
	format agenew age3groups.;
run;
ods output close;
ods pdf close;
title;

proc print data=Log_ageCOV2adjusted3g;
run;
```

```
data Log_ageCOV2adjusted3g1;
	set Log_ageCOV2adjusted3g;
	ODDSRatio1=put(OddsRatioEst,8.3);
	LowerCL1=put(LowerCL,8.3);
	UpperCL1=put(UpperCL,8.3);
	OR=STRIP(ODDSRatio1)||" ("||STRIP(LowerCL1)||", "||STRIP(UpperCL1)||')';
	MODEL=2;
run;

proc print data=Log_ageCOV2adjusted3g1;
	where SCAN(upcase(EFFECT),1)='FIRSTDOSE';
run;

/*** output excel file for results ***/
libname results xlsx "&path\SAS output\&textadd\COVID PLUS FULL COHORT\COVID PLUS FULL COHORT.xlsx";

/* Export first dataset */
data results.sheet1;
   set work.logCOV2;
	where SCAN(upcase(EFFECT),1)='FIRSTDOSE';
run;

/* Export third dataset */
data results.sheet3;
```

```
   set work.Log_ageCOV2adjusted3g1;
        where SCAN(upcase(EFFECT),1)='FIRSTDOSE';
run;

libname results clear;

/*** 1.a. All deaths with categorical age ***/

ods pdf file="&path\SAS output\&textadd\All cause Full cohort\
&textadd Logistic All-cause &todaysdate..pdf";
ods output ODDSRATIOS=log_allcrude;
proc logistic data=dose1matcheddatafinal;
        title "&textadd All-cause deaths" ;
        class firstdose (REF=last)/param=reference;
        model DEATH1YRd1(ref='0')=firstdose/ parmlabel;
run;
ODS OUTPUT CLOSE;
ods pdf close;
title;

proc print;
run;

data Log_allcrude1;
        set Log_allcrude;
        ODDSRatio1=put(OddsRatioEst,8.3);
```

```
            LowerCL1=put(LowerCL,8.3);

            UpperCL1=put(UpperCL,8.3);

            OR=STRIP(ODDSRatio1)||" ("||STRIP(LowerCL1)||", "||STRIP(UpperCL1)||')';

            MODEL=1;

run;

proc print;

run;

ods pdf file="&path\SAS output\&textadd\All cause Full cohort\

&textadd Logistic All-cause adjusted model (cat age) &todaysdate..pdf";

ods output ODDSRATIOS=Log_alladjusted;

proc logistic data=&dataset;

            TITLE 'All-cause death adjusted (logistic regression)';

            class agenew (ref=first) firstdose (REF=last) sex2 (ref="M")

            race4 (ref='WHITE')  ethnicity3

            (ref='NOT HISPANIC') Vaccinationsite (ref="PUBLIX OR CVS OR
WALGREENS")/param=reference;

            model DEATH1YRd1(ref='0')= firstdose agenew sex2 race4 ethnicity3

                              decile_sviscore1 decile_sviscore4

                             mortalityratepct Vaccinationsite/parmlabel;

                    format agenew AGE10yGROUP.;

run;

ods output close;

ods pdf close;
```

```
title;

proc print;
run;

data Log_alladjusted1;
	set log_alladjusted;
	ODDSRatio1=put(OddsRatioEst,8.3);
	LowerCL1=put(LowerCL,8.3);
	UpperCL1=put(UpperCL,8.3);
	OR=STRIP(ODDSRatio1)||" ("||STRIP(LowerCL1)||", "||STRIP(UpperCL1)||')';
	MODEL=2;
run;

proc print data=Log_alladjusted1;
	where SCAN(upcase(EFFECT),1)='VACCINEGROUP2';
run;

DATA LOGALL;
	SET lOG_ALLCRUDE1 lOG_alladjusted1;
RUN;

PROC PRINT data= logall;
	where SCAN(upcase(EFFECT),1)='FIRSTDOSE';
RUN;
```

```
proc sort data=&dataset;
	by agenew;
run;

ods pdf file="&path\SAS output\&textadd\All cause Full cohort\
&textadd logistic All-cause adjusted model (cat age) by Age (3 Age groups)
&todaysdate..pdf";
ods output ODDSRATIOS=Log_agealladjusted3g;
proc logistic data=&dataset;
	title 'All-cause deaths adjusted by Age';
	class agenew (ref=first) FIRSTDOSE (REF=LAST) sex2 (ref="M")
	race4 (ref='WHITE')  ethnicity3
	(ref='NOT HISPANIC') Vaccinationsite (ref="PUBLIX OR CVS OR
WALGREENS")/param=reference;
	model DEATH1YRd1(ref='0')= FIRSTDOSE agenew sex2 race4 ethnicity3
				 decile_sviscore1 decile_sviscore4
				mortalityratepct Vaccinationsite/parmlabel;
	by agenew;
	format agenew age3groups.;
run;
ods output close;
ods pdf close;
title;

proc print data=Log_agealladjusted3g;
run;
```

```
data Log_agealladjusted3g1;
	set Log_agealladjusted3g;
	ODDSRatio1=put(OddsRatioEst,8.3);
	LowerCL1=put(LowerCL,8.3);
	UpperCL1=put(UpperCL,8.3);
	OR=STRIP(ODDSRatio1)||" ("||STRIP(LowerCL1)||", "||STRIP(UpperCL1)||')';
	MODEL=2;
run;

proc print data=Log_agealladjusted3g1;
	where SCAN(upcase(EFFECT),1)='FIRSTDOSE';
run;

/*** output excel file for results ***/
libname results xlsx "&path\SAS output\&textadd\All cause full cohort\allcause full
cohort.xlsx";

/* Export first dataset */
data results.sheet1;
  set work.logall;
	where SCAN(upcase(EFFECT),1)='FIRSTDOSE';
run;

/* Export third dataset */
data results.sheet3;
  set work.log_agealladjusted3g1;
```

```
	where SCAN(upcase(EFFECT),1)='FIRSTDOSE';
run;

libname results clear;

/*** 1.a. CVD deaths with categorical age ***/

ods pdf file="&path\SAS output\&textadd\CVD DEATHS FULL COHORT\
&textadd Logistic CVD DEATHS &todaysdate..pdf";
ods output ODDSRATIOS=log_cvdcrude;
proc logistic data=&dataset;
	title "&textadd CVD deaths" ;
	class firstdose (REF=last)/param=reference;
	model CVD1YRD1(ref='0')=firstdose/ parmlabel;
run;
ODS OUTPUT CLOSE;
ods pdf close;
title;

proc print;
run;

data log_cvdcrude1;
	set log_cvdcrude;
	ODDSRatio1=put(OddsRatioEst,8.3);
	LowerCL1=put(LowerCL,8.3);
```

```
    UpperCL1=put(UpperCL,8.3);
    OR=STRIP(ODDSRatio1)||" ("||STRIP(LowerCL1)||", "||STRIP(UpperCL1)||')';
    MODEL=1;
run;

proc print;
run;

ods pdf file="&path\SAS output\&textadd\CVD DEATHS FULL COHORT\
&textadd Logistic CVD Deaths adjusted model (cat age) &todaysdate..pdf";
ods output ODDSRATIOS=Log_cvdadjusted;
proc logistic data=&dataset;
    TITLE 'CVD Death adjusted (logistic regression)';
    class agenew (ref=first) firstdose (REF=last) sex2 (ref="M")
    race4 (ref='WHITE')  ethnicity3
    (ref='NOT HISPANIC') Vaccinationsite (ref="PUBLIX OR CVS OR
WALGREENS")/param=reference;
    model CVD1YRD1(ref='0')= firstdose agenew sex2 race4 ethnicity3
                     decile_sviscore1 decile_sviscore4
                    mortalityratepct Vaccinationsite/parmlabel;
            format agenew AGE10yGROUP.;
run;
ods output close;
ods pdf close;
title;
```

```
proc print;
run;

data Log_cvdadjusted1;
	set Log_cvdadjusted;
	ODDSRatio1=put(OddsRatioEst,8.3);
	LowerCL1=put(LowerCL,8.3);
	UpperCL1=put(UpperCL,8.3);
	OR=STRIP(ODDSRatio1)||" ("||STRIP(LowerCL1)||", "||STRIP(UpperCL1)||')';
	MODEL=2;
run;

proc print data=Log_cvdadjusted1;
	where SCAN(upcase(EFFECT),1)='FIRSTDOSE';
run;

DATA LOGCVD;
	SET log_cvdcrude1 Log_cvdadjusted1;
RUN;

PROC PRINT data= LOGCVD;
	where SCAN(upcase(EFFECT),1)='FIRSTDOSE';
RUN;

proc sort data=&dataset;
```

```
        by agenew;
run;

ods pdf file="&path\SAS output\&textadd\CVD DEATHS FULL COHORT\
&textadd logistic CVD DEATHS adjusted model (cat age) by Age (3 Age groups)
&todaysdate..pdf";
ods output ODDSRATIOS=Log_agecvdadjusted3g;
proc logistic data=&dataset;
        title 'CVD deaths adjusted by Age';
        class agenew (ref=first) FIRSTDOSE (REF=LAST) sex2 (ref="M")
        race4 (ref='WHITE')  ethnicity3
        (ref='NOT HISPANIC') Vaccinationsite (ref="PUBLIX OR CVS OR
WALGREENS")/param=reference;
        model CVD1YRD1(ref='0')= FIRSTDOSE agenew sex2 race4 ethnicity3
                         decile_sviscore1 decile_sviscore4
                        mortalityratepct Vaccinationsite/parmlabel;
        by agenew;
        format agenew age3groups.;
run;
ods output close;
ods pdf close;
title;

proc print data=Log_agecvdadjusted3g;
run;

data Log_agecvdadjusted3g1;
```

```
    set Log_agecvdadjusted3g;

    ODDSRatio1=put(OddsRatioEst,8.3);

    LowerCL1=put(LowerCL,8.3);

    UpperCL1=put(UpperCL,8.3);

    OR=STRIP(ODDSRatio1)||" ("||STRIP(LowerCL1)||", "||STRIP(UpperCL1)||')';

    MODEL=2;

run;

proc print data=Log_agecvdadjusted3g1;

    where SCAN(upcase(EFFECT),1)='FIRSTDOSE';

run;

/*** output excel file for results ***/

libname results xlsx "&path\SAS output\&textadd\CVD DEATHS FULL COHORT\CVD DEATHS FULL COHORT.xlsx";

/* Export first dataset */

data results.sheet1;

   set work.logcvd;

    where SCAN(upcase(EFFECT),1)='FIRSTDOSE';

run;

/* Export third dataset */

data results.sheet3;

   set work.log_ageCVDadjusted3g1;

    where SCAN(upcase(EFFECT),1)='FIRSTDOSE';
```

```
run;

libname results clear;

/*** COVID DEATHS ***/

/*** 1.a. COVID deaths with categorical age ***/

ods pdf file="&path\SAS output\&textadd\COVID DEATHS FULL COHORT\
&textadd Logistic COVID DEATHS &todaysdate..pdf";
ods output ODDSRATIOS=log_COVIDcrude;
proc logistic data=&dataset;
	title "&textadd COVID deaths" ;
	class firstdose (REF=last)/param=reference;
	model COVIDDEATH1YRD1(ref='0')=firstdose/ parmlabel;
run;
ODS OUTPUT CLOSE;
ods pdf close;
title;

proc print;
run;

data log_COVIDcrude1;
	set log_COVIDcrude;
```

```
	ODDSRatio1=put(OddsRatioEst,8.3);
	LowerCL1=put(LowerCL,8.3);
	UpperCL1=put(UpperCL,8.3);
	OR=STRIP(ODDSRatio1)||" ("||STRIP(LowerCL1)||", "||STRIP(UpperCL1)||')';
	MODEL=1;
run;

proc print;
run;

ods pdf file="&path\SAS output\&textadd\COVID DEATHS FULL COHORT\
&textadd Logistic COVID Deaths adjusted model (cat age) &todaysdate..pdf";
ods output ODDSRATIOS=Log_COVIDadjusted;
proc logistic data=&dataset;
	TITLE 'COVID Death adjusted (logistic regression)';
	class agenew (ref=first) firstdose (REF=last) sex2 (ref="M")
	race4 (ref='WHITE')  ethnicity3
	(ref='NOT HISPANIC') Vaccinationsite (ref="PUBLIX OR CVS OR
WALGREENS")/param=reference;
	model COVIDDEATH1YRD1(ref='0')= firstdose agenew sex2 race4 ethnicity3
					decile_sviscore1 decile_sviscore4
				mortalityratepct Vaccinationsite/parmlabel;
		format agenew AGE10yGROUP.;
run;
ods output close;
```

```
ods pdf close;
title;

proc print;
run;

data Log_COVIDadjusted1;
	set Log_COVIDadjusted;
	ODDSRatio1=put(OddsRatioEst,8.3);
	LowerCL1=put(LowerCL,8.3);
	UpperCL1=put(UpperCL,8.3);
	OR=STRIP(ODDSRatio1)||" ("||STRIP(LowerCL1)||", "||STRIP(UpperCL1)||')';
	MODEL=2;
run;

proc print data=Log_COVIDadjusted1;
	where SCAN(upcase(EFFECT),1)='FIRSTDOSE';
run;

DATA LOGcovid;
	SET log_covidcrude1 Log_covidadjusted1;
RUN;

PROC PRINT data= LOGcovid;
	where SCAN(upcase(EFFECT),1)='FIRSTDOSE';
RUN;
```

```
proc sort data=&dataset;
	by agenew;
run;

ods pdf file="&path\SAS output\&textadd\COVID DEATHS FULL COHORT\
&textadd logistic COVID DEATHS adjusted model (cat age) by Age (3 Age groups)
&todaysdate..pdf";
ods output ODDSRATIOS=Log_ageCOVIDadjusted3g;
proc logistic data=&dataset;
	title 'COVID deaths adjusted by Age';
	class agenew (ref=first) FIRSTDOSE (REF=LAST) sex2 (ref="M")
	race4 (ref='WHITE')  ethnicity3
	(ref='NOT HISPANIC') Vaccinationsite (ref="PUBLIX OR CVS OR
WALGREENS")/param=reference;
	model COVIDDEATH1YRD1(ref='0')= FIRSTDOSE agenew sex2 race4 ethnicity3
				decile_sviscore1 decile_sviscore4
				mortalityratepct Vaccinationsite/parmlabel;
	by agenew;
	format agenew age3groups.;
run;
ods output close;
ods pdf close;
title;

proc print data=Log_ageCOVIDadjusted3g;
run;
```

```
data Log_ageCOVIDadjusted3g1;
	set Log_ageCOVIDadjusted3g;
	ODDSRatio1=put(OddsRatioEst,8.3);
	LowerCL1=put(LowerCL,8.3);
	UpperCL1=put(UpperCL,8.3);
	OR=STRIP(ODDSRatio1)||" ("||STRIP(LowerCL1)||", "||STRIP(UpperCL1)||')';
	MODEL=2;
run;

proc print data=Log_ageCOVIDadjusted3g1;
	where SCAN(upcase(EFFECT),1)='FIRSTDOSE';
run;

/*** output excel file for results ***/
libname results xlsx "&path\SAS output\&textadd\COVID DEATHS FULL COHORT\COVID DEATHS FULL COHORT.xlsx";

/* Export first dataset */
data results.sheet1;
   set work.logCOVID;
	where SCAN(upcase(EFFECT),1)='FIRSTDOSE';
run;

/* Export third dataset */
data results.sheet3;
```

```
   set work.log_ageCOVIDadjusted3g1;
         where SCAN(upcase(EFFECT),1)='FIRSTDOSE';
run;

libname results clear;

/*** NON COVID DEATHS ***/

ods pdf file="&path\SAS output\&textadd\NON COVID DEATHS FULL COHORT\
&textadd Logistic NON COVID DEATHS &todaysdate..pdf";
ods output ODDSRATIOS=log_NONCOVIDcrude;
proc logistic data=&dataset;
         title "&textadd NON COVID deaths" ;
         class firstdose (REF=last)/param=reference;
         model NONCOVIDDEATH1YRD1(ref='0')=firstdose/ parmlabel;
run;
ODS OUTPUT CLOSE;
ods pdf close;
title;

proc print;
run;
```

```
data log_nonCOVIDcrude1;
	set log_NONCOVIDcrude;
	ODDSRatio1=put(OddsRatioEst,8.3);
	LowerCL1=put(LowerCL,8.3);
	UpperCL1=put(UpperCL,8.3);
	OR=STRIP(ODDSRatio1)||" ("||STRIP(LowerCL1)||", "||STRIP(UpperCL1)||')';
	MODEL=1;
run;

proc print;
run;

ods pdf file="&path\SAS output\&textadd\NON COVID DEATHS FULL COHORT\
&textadd Logistic COVID Deaths adjusted model (cat age) &todaysdate..pdf";
ods output ODDSRATIOS=Log_NONCOVIDadjusted;
proc logistic data=&dataset;
	TITLE 'NON COVID Death adjusted (logistic regression)';
	class agenew (ref=first) firstdose (REF=last) sex2 (ref="M")
	race4 (ref='WHITE')  ethnicity3
	(ref='NOT HISPANIC') Vaccinationsite (ref="PUBLIX OR CVS OR
WALGREENS")/param=reference;
	model NONCOVIDDEATH1YRD1(ref='0')= firstdose agenew sex2 race4 ethnicity3
				decile_sviscore1 decile_sviscore4
				mortalityratepct Vaccinationsite/parmlabel;
		format agenew AGE10yGROUP.;
```

```
run;

ods output close;

ods pdf close;

title;

proc print;

run;

data Log_NONCOVIDadjusted1;

	set Log_NONCOVIDadjusted;

	ODDSRatio1=put(OddsRatioEst,8.3);

	LowerCL1=put(LowerCL,8.3);

	UpperCL1=put(UpperCL,8.3);

	OR=STRIP(ODDSRatio1)||" ("||STRIP(LowerCL1)||", "||STRIP(UpperCL1)||')';

	MODEL=2;

run;

proc print data=Log_NONCOVIDadjusted1;

	where SCAN(upcase(EFFECT),1)='FIRSTDOSE';

run;

DATA LOGNONcovid;

	SET log_NONcovidcrude1 Log_NONcovidadjusted1;

RUN;

PROC PRINT data= LOGNONcovid;
```

```
	where SCAN(upcase(EFFECT),1)='FIRSTDOSE';
RUN;

proc sort data=&dataset;
	by agenew;
run;

ods pdf file="&path\SAS output\&textadd\NON COVID DEATHS FULL COHORT\
&textadd logistic Non COVID DEATHS by Age (3 Age groups) &todaysdate..pdf";
ods output ODDSRATIOS=Log_ageNONCOVIDadjusted3g;
proc logistic data=&dataset;
	title 'NON COVID deaths adjusted by Age';
	class agenew (ref=first) FIRSTDOSE (REF=LAST) sex2 (ref="M")
	race4 (ref='WHITE')  ethnicity3
	(ref='NOT HISPANIC') Vaccinationsite (ref="PUBLIX OR CVS OR
WALGREENS")/param=reference;
	model NONCOVIDDEATH1YRD1(ref='0')= FIRSTDOSE agenew sex2 race4 ethnicity3
				decile_sviscore1 decile_sviscore4
				mortalityratepct Vaccinationsite/parmlabel;
	by agenew;
	format agenew age3groups.;
run;
ods output close;
ods pdf close;
title;
```

```
proc print data=Log_ageNONCOVIDadjusted3g;
run;

data Log_ageNONCOVIDadjusted3g1;
	set Log_ageNONCOVIDadjusted3g;
	ODDSRatio1=put(OddsRatioEst,8.3);
	LowerCL1=put(LowerCL,8.3);
	UpperCL1=put(UpperCL,8.3);
	OR=STRIP(ODDSRatio1)||" ("||STRIP(LowerCL1)||", "||STRIP(UpperCL1)||')';
	MODEL=2;
run;

proc print data=Log_ageNONCOVIDadjusted3g1;
	where SCAN(upcase(EFFECT),1)='FIRSTDOSE';
run;

/*** output excel file for results ***/
libname results xlsx "&path\SAS output\&textadd\NON COVID DEATHS FULL COHORT\NON COVID DEATHS FULL COHORT.xlsx";

/* Export first dataset */
data results.sheet1;
   set work.logNONCOVID;
	where SCAN(upcase(EFFECT),1)='FIRSTDOSE';
run;
```

```
/* Export third dataset */
data results.sheet3;
   set work.log_ageNONCOVIDadjusted3g1;
         where SCAN(upcase(EFFECT),1)='FIRSTDOSE';
run;

libname results clear;
```

```
libname store 'C:\Users\MansuriF\OneDrive - Florida Department of
Health\Documents\SAS Datasets';

run;

LIBNAME Deaths ODBC NOPROMPT="driver={ODBC Driver 17 for SQL Server};

Server=CHARTSDATA.DOH.AD.STATE.FL.US;

Database=Vitalstat;

Trusted_Connection=Yes"  schema=dbo stringdates=NO ;

run;

LIBNAME COVVAC ODBC NOPROMPT="Driver={ODBC Driver 17 for SQL Server};

        Server=merlindbreport.doh.ad.state.fl.us;

        Database=CovidVaccine;Trusted_Connection=Yes" schema=dbo stringdates=Yes;

run;

LIBNAME COVID ODBC NOPROMPT="Driver={ODBC Driver 17 for SQL Server};

        Server=merlindbreport.doh.ad.state.fl.us;

        Database=MerlinCOVID;Trusted_Connection=Yes" schema=dbo stringdates=Yes;

run;

options validvarname=any;

%include "H:\EPI\Surveillance Section\Fahad Mansuri\SURVIVAL ANALYSIS 2024\SAS
CODE\formats for analysis.sas";
```

```
%include "H:\EPI\Surveillance Section\Fahad Mansuri\SURVIVAL ANALYSIS 2024\SAS CODE\relrisk9.sas";

proc means data=store.dose1matcheddata nmiss;
	var sviscore:;
run;

proc stdize data=store.dose1matcheddata reponly method=mean missing=mean out=dose1matcheddata;
	var sviscore:;
run;

proc means data=dose1matcheddata nmiss;
	var sviscore:;
run;

/*** Descriptives for matched data vs all data ***/

proc rank data=dose1matcheddata out=dose1matcheddata groups=10;
	var sviscore1 sviscore2 sviscore3 sviscore4 sviscore SESPOV
	SESUNEMP SESINCOME	SESNOHIGH	HTMUS		HTMOBILEHOME
	HTNOVEHICLE		HTCROWDING		HTGQ;
	ranks decile_sviscore1 decile_sviscore2 decile_sviscore3 decile_sviscore4 decile_sviscore
	decile_SESPOV		decile_SESUNEMP decile_SESINCOME	decile_SESNOHIGH
	decile_HTMUS		decile_HTMOBILEHOME
	decile_HTNOVEHICLE		decile_HTCROWDING		decile_HTGQ;
```

```
run;

proc contents data=store.dose1matcheddata order=varnum;
run;

proc sort data=dose1matcheddata nodupkey out=test;
	by DWHPatient;
run;

LIBNAME COVID ODBC NOPROMPT="Driver={ODBC Driver 17 for SQL Server};
	Server=merlindbreport.doh.ad.state.fl.us;
	Database=MerlinCOVID;Trusted_Connection=Yes" schema=dbo stringdates=no;
run;

proc contents data=covid.cases order=varnum;
run;

proc contents data=COVVAC.COVIDPatientIdentity order=varnum;
run;

data covidcases;
	set covid.cases;
	keep 'Case ID'n 'Profile ID'n 'shots id'n 'Publish case'n Jurisdiction
	'Earliest lab event date'n 'Date became case'n 'Dx status'n;
	rename 'shots id'n =shotsid2 'Publish case'n =Publishcase
	'Earliest lab event date'n=ELD
```

```
    'Date became case'n =casedate
    'Dx status'n=dxstatus 'Profile ID'n=profileid 'Case ID'n=caseid;
    where upcase('Publish case'n)="YES" and  "01Mar2020"d le 'Earliest lab event date'n le "31dec2022"D
    and      Jurisdiction="FL resident";
run;

proc sql;
    select count(distinct profileid)
    from covidcases
    where shotsid2 ne " ";
quit;

proc sql;
    select count (profileid)
    from covidcases
    where shotsid2 ne " ";
quit;

proc sql;
    select count(distinct caseid)
    from covidcases        where shotsid2 ne " ";
quit;

proc sql;
    select count(caseid)
```

```
        from covidcases     where shotsid2 ne " ";
quit;

proc sql;
        select count (distinct shotsid2)
                from covidcases     where shotsid2 ne " ";
quit;

proc sql;
        select count (shotsid2)
                from covidcases     where shotsid2 ne " ";
quit;

proc sort data=work.covidcases;
        by shotsid2;
run;

proc transpose data=covidcases out=covidcases2 name=ELD prefix=ELD;
        by shotsid2;
        var eld;
        where shotsid2 ne " ";
run;

proc freq data=covidcases;
        table Jurisdiction dxstatus;
run;
```

```
proc contents data=store.dose1matcheddata;
run;

proc datasets lib=work;
run;

proc sort data=dose1matcheddata;
	by shotsid2;
run;

proc sort data=covidcases2;
	by shotsid2;
run;

proc sql;
	create table dose1matchedatawithcovid as
	select a.*,b.*
	from dose1matcheddata as a
	left join covidcases2 as b
	on a.CurrentStateIMMId=b.shotsid2;
quit;

options validvarname=any;
```

```
proc contents data=store.covidvaccination5 order=varnum;
run;

data dose1matcheddatafinal;
	set dose1matchedatawithcovid (keep= ageatdose1 agegroupnew sex2 race4
ethnicity3 firstdose agenew
		decile_sviscore1 decile_sviscore4 mortalityratepct Vaccinationsite
		vaccinedose2 seconddose DWHPatient CurrentStateIMMId
VaccinationDate1
		DWHPatient firstdose seconddose agenew sex2 race4 ethnicity3
Vaccinationsite
		sviscore: VaccinationDate: monthdose1 timebetweendose timeokay
mortalityratepct
		matchedpairs icd_code geoidnew vaccinedose2 daystodeath daystodeathd1
death1yrd1 death1yr
		'vs dod'n nursinghomevaccinationfinal HOMELESSFLAG
		TRANSAX1-TRANSAX15 eld:);
	where SEX2 ne "U" and
	Vaccinationsite not in ("CORRECTIONAL FACILITY","NURSING HOME")
	and nursinghomevaccinationfinal ne 1 AND HOMELESSFLAG=0;
	if seconddose=" " then noseconddose=1;
	else noseconddose=0;
	format agenew age10ygroup.;
	mortalityratepct=round(mortalityratepct,1);
	IF FIRSTDOSE="1. PFIZER" THEN EXPOS=1;
	ELSE EXPOS=0;
```

```
if death1yrd1=1 and substr(icd_code,1,2) in ("I0","I1","I2","I3","I4","I5","I6") then CVD1YRD1=1;
else if death1yrD1=1 and substr(icd_code,1,3) in ("I71","I72","I73","I74","I75","I76","I77") then CVD1YRD1=1;
else CVD1YRD1=0;
if SUBSTR(icd_code,1,3) IN ("X40","X41","X42","X43","X44",
"X60","X61","X62","X62","X64","X85","Y10","Y11","Y12","Y13","Y14")
and death1yrD1=1 then overdose1yrD1=1;
else overdose1yrD1=0;
if SUBSTR(icd_code,1,3) in ("X85","X86","X87","X88","X89","X90","X91","X92"
        ,"X93","X94","X95","X96","X97","X98","X99","Y00","Y01","Y02","Y03"
        ,"Y04","Y05","Y06","Y07","Y08","Y09")
and DEATH1YRD1=1 then homicide1yrD1=1;
else if substr(icd_code,1,4)="Y871" and death1yrD1=1 then homicide1yrD1=1;
else homicide1yrD1=0;
if SUBSTR (ICD_CODE,1,2) IN ("X6","X7") AND death1yrD1=1 THEN SUICIDE1YRD1=1;
ELSE IF SUBSTR (ICD_CODE,1,3) IN ("X80","X81","X82","X83","X84") AND DEATH1YRD1=1 THEN SUICIDE1YRD1=1;
ELSE IF substr(icd_code,1,4)="Y870" and death1yr=1 THEN SUICIDE1YR=1;
ELSE SUICIDE1YRD1=0;
if SUBSTR (ICD_CODE,1,2) IN ("V0","V1","V2","V3","V4","V5","V6","V7","V8","V9"
,"W0","W1","W2","W3","W4","W5","W6","W7","W8","W9","X0","X1","X2","X3","X4")
AND DEATH1YRD1=1 THEN UNINTENTIONAL1YRD1=1;
ELSE IF SUBSTR(ICD_CODE,1,3) IN ("X50","X51","X52","X53","X54","X55","X56","X57","X58","X59"
        ,"Y85","Y86") AND DEATH1YRD1=1 THEN UNINTENTIONAL1YRD1=1;
```

```
ELSE UNINTENTIONAL1YRD1=0;

if SUBSTR (ICD_CODE,1,2) IN ("V0","V1","V2","V3","V4","V5","V6","V7","V8","V9"
,"W2","W3","W4","W5","W6","W7","W8","W9","X0","X1","X2","X3","X4")
AND DEATH1YRD1=1 THEN NOFALLUNINTENTIONAL1YRD1=1;
ELSE IF SUBSTR(ICD_CODE,1,3) IN
("X50","X51","X52","X53","X54","X55","X56","X57","X58","X59"
                    ,"Y85","Y86") AND DEATH1YRD1=1 THEN
NOFALLUNINTENTIONAL1YRD1=1;
ELSE NOFALLUNINTENTIONAL1YRD1=0;

if homicide1yrD1=1 or UNINTENTIONAL1YRD1=1 or SUICIDE1YRD1=1 or
overdose1yrD1=1 then unnatural1yrD1=1;
else unnatural1yrD1=0;
if homicide1yrD1=1 or SUICIDE1YRD1=1 then suicidehomicide1yrD1=1;
else suicidehomicide1yrD1=0;
if homicide1yrD1=1 or NOFALLUNINTENTIONAL1YRD1=1 or SUICIDE1YRD1=1 or
overdose1yrD1=1
then NOFALLunnatural1yrD1=1;
else NOFALLunnatural1yrD1=0;

if SUBSTR(ICD_CODE,1,2) in ("V2","V3","V4","V5","V6","V7") and death1yrD1=1 then
mva1yrD1=1;
else if SUBSTR(ICD_CODE,1,3) in
("V02","V03","V04","V12","V13","V14","V83","V84","V85","V86")
and death1yrD1=1 then mva1yrD1=1;
if icd_code in
("V090","V092","V190","V191","V192","V194","V195","V196","V803","V804",
```

```
"V805","V810","V811","V820","V821","V870","V871","V872","V873","V874","V875",
"V876","V877","V878","V881","V882","V883","V884","V885","V886","V887","V888","V890"
,"V892")
AND DEATH1YRD1=1 THEN MVA1YRD1=1;
ELSE MVA1YRD1=0;
array test {*} transax: ; /*** Select all columns with ACME prefix ***/
count =0;
do j= 1 to 15;
    if test{j} ne " " then do;
    count + (test{j} in: ("U071"));
    end;
end;
if count>0 then COVIDDEATH=1;
else COVIDDEATH=0;
if COVIDDEATH=1 and DEATH1YRD1=1 then COVIDDEATH1YRD1=1;
ELSE COVIDDEATH1YRD1=0;
if COVIDDEATH=0 and DEATH1YRD1=1 then NONCOVIDDEATH1YRD1=1;
ELSE NONCOVIDDEATH1YRD1=0;
counteld=0;
array new ELD1-ELD6;
    do i =1 to dim(new);
    if new{i} ne . and 'vs dod'n ne . then do;
    counteld+(vaccinationdate1 le new{i} <= 'vs dod'n);
    end;
end;
if DEATH1YRD1=1 and (COVIDDEATH1YRD1=1 or counteld >0) then
COVIDDEATHPLUS1YRD1=1;
```

```
ELSE COVIDDEATHPLUS1YRD1=0;

if DEATH1YRD1=1 and (COVIDDEATHPLUS1YRD1=0) then
NONCOVIDDEATHPLUS1YRD1=1;
ELSE NONCOVIDDEATHPLUS1YRD1=0;

if counteld>0 then do;
    if eld6 ne . and 'vs dod'n >=eld6 then covinftodeathdys='vs dod'n-eld6;
    else if eld5 ne . and 'vs dod'n >=eld5 then covinftodeathdys='vs dod'n-eld5;
    else if eld4 ne . and 'vs dod'n >=eld4 then covinftodeathdys='vs dod'n-eld4;
    else if eld3 ne . and 'vs dod'n >=eld3 then covinftodeathdys='vs dod'n-eld3;
    else if eld2 ne . and 'vs dod'n >=eld2 then covinftodeathdys='vs dod'n-eld2;

    else if eld1 ne . and 'vs dod'n >=eld1 then covinftodeathdys='vs dod'n-eld1;
end;
if daystodeathd1=. then daystodeathd1=366;
countcovinf=0;
countcovinfbf20=0;

array covinf ELD1-ELD6;
    do k =1 to dim(covinf);
    if covinf{k} ne . then do;
    countcovinf+(covinf{k} le vaccinationdate1);
    countcovinfbf20+(covinf{k} le "20dec2020"d);

    end;
end;
```

```
    drop i j k;
    if countcovinf>0 then covidinf=1;
    else covidinf=0;
    if countcovinfbf20>0 then covinfbf20dec=1;
    else covinfbf20dec=0;
run;

proc freq data=work.dose1matcheddatafinal;
    table countcovinf:;
run;

proc means data=work.dose1matcheddatafinal;
    var daystodeath daystodeathd1;
run;

proc freq data=work.dose1matcheddatafinal;
    table counteld*death1yrd1;
    table covinftodeathdys;
run;

proc print data=work.dose1matcheddatafinal (obs= 100);
    where COVIDDEATHPLUS1YRD1=1 and covinftodeathdys<0;
    var DWHPatient VaccinationDate: eld: counteld 'vs dod'n covinftodeathdys;
run;
```

```
proc freq data=dose1matcheddatafinal;
	tABLE daystodeath;
RUN;

proc freq data=dose1matcheddatafinal;
	tABLE death1yrd1 CVD: covid: NONCOVID: suicidehomicide1yrD1;
RUN;

/** On matched cohort **/

DATA MATCHEDCOHORTnocovinf;
	SET dose1matcheddatafinal;
	WHERE matchedpairs=1 and countcovinf=0;
RUN;

/*** Risk of All-cause, CVD, COVID, and Non-COVID Deaths			***/

/*** MATCHED COHORT			***/

%let dataset=MATCHEDCOHORTnocovinf;

%let todaysdate=%sysfunc(today(),date9.);

%put &todaysdate;
```

```
%let textadd=Matched cohort Risk of death Dose 1 no COVINF;

%let path=H:\EPI\Surveillance Section\Fahad Mansuri\SURVIVAL ANALYSIS 2024;

proc contents data=work.MATCHEDCOHORT;

run;

proc freq data=work.dose1matcheddatafinal;

        table firstdose agenew firstdose agenew sex2 race4 ethnicity3

                        decile_sviscore1 decile_sviscore4

                        mortalityratepct Vaccinationsite/ missing;

run;

/*** 1.a. SUIC or HOMICIDE  with categorical age ***/

ods pdf file="&path\SAS output\&textadd\SUICIDE or HOMICIDE\

&textadd Logistic SUICHOM &todaysdate..pdf";

ods output ODDSRATIOS=log_SUICHOMcrude;

proc logistic data=&dataset;

        title "&textadd SUICIDE OR HOMICIDE deaths" ;

        class firstdose (REF=last)/param=reference;

        model suicidehomicide1yrD1(ref='0')=firstdose/ parmlabel;

run;

ODS OUTPUT CLOSE;
```

```
ods pdf close;

title;

proc print;

run;

data log_SUICHOMcrude1;

	set log_SUICHOMcrude;

	ODDSRatio1=put(OddsRatioEst,8.3);

	LowerCL1=put(LowerCL,8.3);

	UpperCL1=put(UpperCL,8.3);

	OR=STRIP(ODDSRatio1)||" ("||STRIP(LowerCL1)||", "||STRIP(UpperCL1)||')';

	MODEL=1;

run;

proc print;

run;

ods pdf file="&path\SAS output\&textadd\SUICIDE OR HOMICIDE\

&textadd Logistic SUICHOM adjusted model (cat age) &todaysdate..pdf";

ods output ODDSRATIOS=Log_SUICHOMadjusted;

proc logistic data=&dataset;

	TITLE 'SUICIDE OR HOMICIDE death adjusted (logistic regression)';

	class agenew (ref=first) firstdose (REF=last) sex2 (ref="M")

	race4 (ref='WHITE')  ethnicity3
```

```
	(ref='NOT HISPANIC') Vaccinationsite (ref="PUBLIX OR CVS OR
WALGREENS")/param=reference;
	model suicidehomicide1yrD1(ref='0')= firstdose agenew sex2 race4 ethnicity3
				decile_sviscore1 decile_sviscore4
				mortalityratepct Vaccinationsite/parmlabel;
		format agenew AGE10yGROUP.;
run;
ods output close;
ods pdf close;
title;

proc print;
run;

data Log_SUICHOMadjusted1;
	set Log_SUICHOMadjusted;
	ODDSRatio1=put(OddsRatioEst,8.3);
	LowerCL1=put(LowerCL,8.3);
	UpperCL1=put(UpperCL,8.3);
	OR=STRIP(ODDSRatio1)||" ("||STRIP(LowerCL1)||", "||STRIP(UpperCL1)||')';
	MODEL=2;
run;

proc print data=Log_SUICHOMadjusted1;
	where SCAN(upcase(EFFECT),1)='FIRSTDOSE';
run;
```

```
DATA LOGSUICHOM;
	SET lOG_SUICHOMCRUDE1 Log_SUICHOMadjusted1;
RUN;

PROC PRINT data= LOGSUICHOM;
	where SCAN(upcase(EFFECT),1)='FIRSTDOSE';
RUN;

proc sort data=&dataset;
	by agenew;
run;

ods pdf file="&path\SAS output\&textadd\SUICIDE OR HOMICIDE\
&textadd logistic SUICHOM adjusted model (cat age) by Age (3 Age groups)
&todaysdate..pdf";
ods output ODDSRATIOS=Log_ageSUICHOMadjusted3g;
proc logistic data=&dataset;
	title 'SUICIDE OR HOMICIDE deaths adjusted by Age';
	class agenew (ref=first) FIRSTDOSE (REF=LAST) sex2 (ref="M")
	race4 (ref='WHITE')  ethnicity3
	(ref='NOT HISPANIC') Vaccinationsite (ref="PUBLIX OR CVS OR
WALGREENS")/param=reference;
	model suicidehomicide1yrD1(ref='0')= FIRSTDOSE agenew sex2 race4 ethnicity3
				decile_sviscore1 decile_sviscore4
				mortalityratepct Vaccinationsite/parmlabel;
	by agenew;
```

```
    format agenew age3groups.;
run;
ods output close;
ods pdf close;
title;

proc print data=Log_ageSUICHOMadjusted3g;
run;

data Log_ageSUICHOMadjusted3g1;
    set Log_ageSUICHOMadjusted3g;
    ODDSRatio1=put(OddsRatioEst,8.3);
    LowerCL1=put(LowerCL,8.3);
    UpperCL1=put(UpperCL,8.3);
    OR=STRIP(ODDSRatio1)||" ("||STRIP(LowerCL1)||", "||STRIP(UpperCL1)||')';
    MODEL=2;
run;

proc print data=Log_ageSUICHOMadjusted3g1;
    where SCAN(upcase(EFFECT),1)='FIRSTDOSE';
run;

/*** output excel file for results ***/
libname results xlsx "&path\SAS output\&textadd\SUICIDE OR HOMICIDE\SUICIDE OR HOMICIDE.xlsx";
```

```
/* Export first dataset */

data results.sheet1;

   set work.logSUICHOM;

            where SCAN(upcase(EFFECT),1)='FIRSTDOSE';

run;

/* Export third dataset */

data results.sheet3;

   set work.Log_ageSUICHOMadjusted3g1;

            where SCAN(upcase(EFFECT),1)='FIRSTDOSE';

run;

libname results clear;

/*** 1.a. NON COVID +  with categorical age ***/

ods pdf file="&path\SAS output\&textadd\NON COVID PLUS\

&textadd Logistic COVID + &todaysdate..pdf";

ods output ODDSRATIOS=log_NONCOV2crude;

proc logistic data=&dataset;

            title "&textadd NON COVID + deaths" ;

            class firstdose (REF=last)/param=reference;

            model NONCOVIDDEATHPLUS1YRD1(ref='0')=firstdose/ parmlabel;

run;

ODS OUTPUT CLOSE;

ods pdf close;
```

```
title;

proc print;
run;

data Log_NONCOV2crude1;
	set Log_NONCOV2crude;
	ODDSRatio1=put(OddsRatioEst,8.3);
	LowerCL1=put(LowerCL,8.3);
	UpperCL1=put(UpperCL,8.3);
	OR=STRIP(ODDSRatio1)||" ("||STRIP(LowerCL1)||", "||STRIP(UpperCL1)||')';
	MODEL=1;
run;

proc print;
run;

ods pdf file="&path\SAS output\&textadd\NON COVID PLUS\
&textadd Logistic NON COVID + adjusted model (cat age) &todaysdate..pdf";
ods output ODDSRATIOS=Log_NONCOV2adjusted;
proc logistic data=&dataset;
	TITLE 'NON COVID + death adjusted (logistic regression)';
	class agenew (ref=first) firstdose (REF=last) sex2 (ref="M")
	race4 (ref='WHITE')  ethnicity3
```

```
	(ref='NOT HISPANIC') Vaccinationsite (ref="PUBLIX OR CVS OR WALGREENS")/param=reference;
	model NONCOVIDDEATHPLUS1YRD1(ref='0')= firstdose agenew sex2 race4 ethnicity3
				decile_sviscore1 decile_sviscore4
				mortalityratepct Vaccinationsite/parmlabel;
			format agenew AGE10yGROUP.;
run;
ods output close;
ods pdf close;
title;

proc print;
run;

data Log_NONCOV2adjusted1;
	set Log_NONCOV2adjusted;
	ODDSRatio1=put(OddsRatioEst,8.3);
	LowerCL1=put(LowerCL,8.3);
	UpperCL1=put(UpperCL,8.3);
	OR=STRIP(ODDSRatio1)||" ("||STRIP(LowerCL1)||", "||STRIP(UpperCL1)||')';
	MODEL=2;
run;

proc print data=Log_NONCOV2adjusted1;
	where SCAN(upcase(EFFECT),1)='VACCINEGROUP2';
run;
```

```
DATA LOGNONCOV2;
	SET lOG_NONCOV2CRUDE1 lOG_NONCOV2adjusted1;
RUN;

PROC PRINT data= LOGNONCOV2;
	where SCAN(upcase(EFFECT),1)='FIRSTDOSE';
RUN;

proc sort data=&dataset;
	by agenew;
run;

ods pdf file="&path\SAS output\&textadd\NON COVID PLUS\
&textadd logistic NON COVID + adjusted model (cat age) by Age (3 Age groups)
&todaysdate..pdf";
ods output ODDSRATIOS=Log_ageNONCOV2adjusted3g;
proc logistic data=&dataset;
	title 'NON COVID + deaths adjusted by Age';
	class agenew (ref=first) FIRSTDOSE (REF=LAST) sex2 (ref="M")
	race4 (ref='WHITE')  ethnicity3
	(ref='NOT HISPANIC') Vaccinationsite (ref="PUBLIX OR CVS OR
WALGREENS")/param=reference;
	model NONCOVIDDEATHPLUS1YRD1(ref='0')= FIRSTDOSE agenew sex2 race4
ethnicity3
				decile_sviscore1 decile_sviscore4
				mortalityratepct Vaccinationsite/parmlabel;
```

```
            by agenew;
            format agenew age3groups.;
run;
ods output close;
ods pdf close;
title;

proc print data=Log_ageNONCOV2adjusted3g;
run;

data Log_ageNONCOV2adjusted3g1;
            set Log_ageNONCOV2adjusted3g;
            ODDSRatio1=put(OddsRatioEst,8.3);
            LowerCL1=put(LowerCL,8.3);
            UpperCL1=put(UpperCL,8.3);
            OR=STRIP(ODDSRatio1)||" ("||STRIP(LowerCL1)||", "||STRIP(UpperCL1)||')';
            MODEL=2;
run;

proc print data=Log_ageNONCOV2adjusted3g1;
            where SCAN(upcase(EFFECT),1)='FIRSTDOSE';
run;

/*** output excel file for results ***/
libname results xlsx "&path\SAS output\&textadd\NON COVID PLUS\NON COVID
PLUS.xlsx";
```

```
/* Export first dataset */
data results.sheet1;
   set work.logNONCOV2;
         where SCAN(upcase(EFFECT),1)='FIRSTDOSE';
run;

/* Export third dataset */
data results.sheet3;
   set work.Log_ageNONCOV2adjusted3g1;
         where SCAN(upcase(EFFECT),1)='FIRSTDOSE';
run;

libname results clear;

/*** 1.a. COVID +  with categorical age ***/

ods pdf file="&path\SAS output\&textadd\COVID PLUS\
&textadd Logistic COVID + &todaysdate..pdf";
ods output ODDSRATIOS=log_COV2crude;
proc logistic data=&dataset;
         title "&textadd COVID + deaths" ;
         class firstdose (REF=last)/param=reference;
         model COVIDDEATHPLUS1YRD1(ref='0')=firstdose/ parmlabel;
run;
```

```
ODS OUTPUT CLOSE;
ods pdf close;
title;

proc print;
run;

data Log_COV2crude1;
    set Log_COV2crude;
    ODDSRatio1=put(OddsRatioEst,8.3);
    LowerCL1=put(LowerCL,8.3);
    UpperCL1=put(UpperCL,8.3);
    OR=STRIP(ODDSRatio1)||" ("||STRIP(LowerCL1)||", "||STRIP(UpperCL1)||')';
    MODEL=1;
run;

proc print;
run;

ods pdf file="&path\SAS output\&textadd\COVID PLUS\
&textadd Logistic COVID + adjusted model (cat age) &todaysdate..pdf";
ods output ODDSRATIOS=Log_COV2adjusted;
proc logistic data=&dataset;
    TITLE 'COVID + death adjusted (logistic regression)';
    class agenew (ref=first) firstdose (REF=last) sex2 (ref="M")
```

```
        race4 (ref='WHITE')  ethnicity3
        (ref='NOT HISPANIC') Vaccinationsite (ref="PUBLIX OR CVS OR
WALGREENS")/param=reference;
        model COVIDDEATHPLUS1YRD1(ref='0')= firstdose agenew sex2 race4 ethnicity3
                        decile_sviscore1 decile_sviscore4
                        mortalityratepct Vaccinationsite/parmlabel;
                format agenew AGE10yGROUP.;
run;
ods output close;
ods pdf close;
title;

proc print;
run;

data Log_COV2adjusted1;
        set Log_COV2adjusted;
        ODDSRatio1=put(OddsRatioEst,8.3);
        LowerCL1=put(LowerCL,8.3);
        UpperCL1=put(UpperCL,8.3);
        OR=STRIP(ODDSRatio1)||" ("||STRIP(LowerCL1)||", "||STRIP(UpperCL1)||')';
        MODEL=2;
run;

proc print data=Log_COV2adjusted1;
        where SCAN(upcase(EFFECT),1)='VACCINEGROUP2';
```

```
run;

DATA LOGCOV2;
	SET lOG_COV2CRUDE1 lOG_COV2adjusted1;
RUN;

PROC PRINT data= LOGCOV2;
	where SCAN(upcase(EFFECT),1)='FIRSTDOSE';
RUN;

proc sort data=&dataset;
	by agenew;
run;

ods pdf file="&path\SAS output\&textadd\COVID PLUS\
&textadd logistic COVID + adjusted model (cat age) by Age (3 Age groups)
&todaysdate..pdf";
ods output ODDSRATIOS=Log_ageCOV2adjusted3g;
proc logistic data=&dataset;
	title 'COVID + deaths adjusted by Age';
	class agenew (ref=first) FIRSTDOSE (REF=LAST) sex2 (ref="M")
	race4 (ref='WHITE')  ethnicity3
	(ref='NOT HISPANIC') Vaccinationsite (ref="PUBLIX OR CVS OR
WALGREENS")/param=reference;
	model COVIDDEATHPLUS1YRD1(ref='0')= FIRSTDOSE agenew sex2 race4 ethnicity3
				decile_sviscore1 decile_sviscore4
				mortalityratepct Vaccinationsite/parmlabel;
```

```
	by agenew;
	format agenew age3groups.;
run;
ods output close;
ods pdf close;
title;

proc print data=Log_ageCOV2adjusted3g;
run;

data Log_ageCOV2adjusted3g1;
	set Log_ageCOV2adjusted3g;
	ODDSRatio1=put(OddsRatioEst,8.3);
	LowerCL1=put(LowerCL,8.3);
	UpperCL1=put(UpperCL,8.3);
	OR=STRIP(ODDSRatio1)||" ("||STRIP(LowerCL1)||", "||STRIP(UpperCL1)||')';
	MODEL=2;
run;

proc print data=Log_ageCOV2adjusted3g1;
	where SCAN(upcase(EFFECT),1)='FIRSTDOSE';
run;

/*** output excel file for results ***/
libname results xlsx "&path\SAS output\&textadd\COVID PLUS\COVID PLUS.xlsx";
```

```
/* Export first dataset */
data results.sheet1;
   set work.logCOV2;
            where SCAN(upcase(EFFECT),1)='FIRSTDOSE';
run;

/* Export third dataset */
data results.sheet3;
   set work.Log_ageCOV2adjusted3g1;
            where SCAN(upcase(EFFECT),1)='FIRSTDOSE';
run;

libname results clear;

/*** 1.a. All deaths with categorical age ***/

ods pdf file="&path\SAS output\&textadd\All cause\
&textadd Logistic All-cause &todaysdate..pdf";
ods output ODDSRATIOS=log_allcrude;
proc logistic data=&dataset;
            title "&textadd All-cause deaths" ;
            class firstdose (REF=last)/param=reference;
            model DEATH1YRd1(ref='0')=firstdose/ parmlabel;
run;
ODS OUTPUT CLOSE;
ods pdf close;
```

```
title;

proc print;
run;

data Log_allcrude1;
	set Log_allcrude;
	ODDSRatio1=put(OddsRatioEst,8.3);
	LowerCL1=put(LowerCL,8.3);
	UpperCL1=put(UpperCL,8.3);
	OR=STRIP(ODDSRatio1)||" ("||STRIP(LowerCL1)||", "||STRIP(UpperCL1)||')';
	MODEL=1;
run;

proc print;
run;

ods pdf file="&path\SAS output\&textadd\All cause\
&textadd Logistic All-cause adjusted model (cat age) &todaysdate..pdf";
ods output ODDSRATIOS=Log_alladjusted;
proc logistic data=&dataset;
	TITLE 'All-cause death adjusted (logistic regression)';
	class agenew (ref=first) firstdose (REF=last) sex2 (ref="M")
	race4 (ref='WHITE')  ethnicity3
```

```
            (ref='NOT HISPANIC') Vaccinationsite (ref="PUBLIX OR CVS OR
WALGREENS")/param=reference;
            model DEATH1YRd1(ref='0')= firstdose agenew sex2 race4 ethnicity3
                                 decile_sviscore1 decile_sviscore4
                                mortalityratepct Vaccinationsite/parmlabel;
                      format agenew AGE10yGROUP.;
run;
ods output close;
ods pdf close;
title;

proc print;
run;

data Log_alladjusted1;
            set log_alladjusted;
            ODDSRatio1=put(OddsRatioEst,8.3);
            LowerCL1=put(LowerCL,8.3);
            UpperCL1=put(UpperCL,8.3);
            OR=STRIP(ODDSRatio1)||" ("||STRIP(LowerCL1)||", "||STRIP(UpperCL1)||')';
            MODEL=2;
run;

proc print data=Log_alladjusted1;
            where SCAN(upcase(EFFECT),1)='VACCINEGROUP2';
run;
```

```
DATA LOGALL;
	SET lOG_ALLCRUDE1 lOG_alladjusted1;
RUN;

PROC PRINT data= logall;
	where SCAN(upcase(EFFECT),1)='FIRSTDOSE';
RUN;

proc sort data=&dataset;
	by agenew;
run;

ods pdf file="&path\SAS output\&textadd\All cause\
&textadd logistic All-cause adjusted model (cat age) by Age (3 Age groups)
&todaysdate..pdf";
ods output ODDSRATIOS=Log_agealladjusted3g;
proc logistic data=&dataset;
	title 'All-cause deaths adjusted by Age';
	class agenew (ref=first) FIRSTDOSE (REF=LAST) sex2 (ref="M")
	race4 (ref='WHITE')  ethnicity3
	(ref='NOT HISPANIC') Vaccinationsite (ref="PUBLIX OR CVS OR
WALGREENS")/param=reference;
	model DEATH1YRd1(ref='0')= FIRSTDOSE agenew sex2 race4 ethnicity3
				decile_sviscore1 decile_sviscore4
				mortalityratepct Vaccinationsite/parmlabel;
	by agenew;
```

```
    format agenew age3groups.;
run;
ods output close;
ods pdf close;
title;

proc print data=Log_agealladjusted3g;
run;

data Log_agealladjusted3g1;
    set Log_agealladjusted3g;
    ODDSRatio1=put(OddsRatioEst,8.3);
    LowerCL1=put(LowerCL,8.3);
    UpperCL1=put(UpperCL,8.3);
    OR=STRIP(ODDSRatio1)||" ("||STRIP(LowerCL1)||", "||STRIP(UpperCL1)||')';
    MODEL=2;
run;

proc print data=Log_agealladjusted3g1;
    where SCAN(upcase(EFFECT),1)='FIRSTDOSE';
run;

/*** output excel file for results ***/
libname results xlsx "&path\SAS output\&textadd\All cause\allcause.xlsx";

/* Export first dataset */
```

```
data results.sheet1;
   set work.logall;
          where SCAN(upcase(EFFECT),1)='FIRSTDOSE';
run;

/* Export third dataset */
data results.sheet3;
   set work.log_agealladjusted3g1;
          where SCAN(upcase(EFFECT),1)='FIRSTDOSE';
run;

libname results clear;

/*** 1.a. CVD deaths with categorical age ***/

ods pdf file="&path\SAS output\&textadd\CVD DEATHS\
&textadd Logistic CVD DEATHS &todaysdate..pdf";
ods output ODDSRATIOS=log_cvdcrude;
proc logistic data=&dataset;
          title "&textadd CVD deaths" ;
          class firstdose (REF=last)/param=reference;
          model CVD1YRD1(ref='0')=firstdose/ parmlabel;
run;
ODS OUTPUT CLOSE;
ods pdf close;
title;
```

```
proc print;
run;

data log_cvdcrude1;
	set log_cvdcrude;
	ODDSRatio1=put(OddsRatioEst,8.3);
	LowerCL1=put(LowerCL,8.3);
	UpperCL1=put(UpperCL,8.3);
	OR=STRIP(ODDSRatio1)||" ("||STRIP(LowerCL1)||", "||STRIP(UpperCL1)||')';
	MODEL=1;
run;

proc print;
run;

ods pdf file="&path\SAS output\&textadd\CVD DEATHS\
&textadd Logistic CVD Deaths adjusted model (cat age) &todaysdate..pdf";
ods output ODDSRATIOS=Log_cvdadjusted;
proc logistic data=&dataset;
	TITLE 'CVD Death adjusted (logistic regression)';
	class agenew (ref=first) firstdose (REF=last) sex2 (ref="M")
	race4 (ref='WHITE')  ethnicity3
	(ref='NOT HISPANIC') Vaccinationsite (ref="PUBLIX OR CVS OR
WALGREENS")/param=reference;
```

```
	model CVD1YRD1(ref='0')= firstdose agenew sex2 race4 ethnicity3
				decile_sviscore1 decile_sviscore4
				mortalityratepct Vaccinationsite/parmlabel;
		format agenew AGE10yGROUP.;
run;
ods output close;
ods pdf close;
title;

proc print;
run;

data Log_cvdadjusted1;
	set Log_cvdadjusted;
	ODDSRatio1=put(OddsRatioEst,8.3);
	LowerCL1=put(LowerCL,8.3);
	UpperCL1=put(UpperCL,8.3);
	OR=STRIP(ODDSRatio1)||" ("||STRIP(LowerCL1)||", "||STRIP(UpperCL1)||')';
	MODEL=2;
run;

proc print data=Log_cvdadjusted1;
	where SCAN(upcase(EFFECT),1)='FIRSTDOSE';
run;

DATA LOGCVD;
```

```
	SET log_cvdcrude1 Log_cvdadjusted1;
RUN;

PROC PRINT data= LOGCVD;
	where SCAN(upcase(EFFECT),1)='FIRSTDOSE';
RUN;

proc sort data=&dataset;
	by agenew;
run;

ods pdf file="&path\SAS output\&textadd\CVD DEATHS\
&textadd logistic CVD DEATHS adjusted model (cat age) by Age (3 Age groups)
&todaysdate..pdf";
ods output ODDSRATIOS=Log_agecvdadjusted3g;
proc logistic data=&dataset;
	title 'CVD deaths adjusted by Age';
	class agenew (ref=first) FIRSTDOSE (REF=LAST) sex2 (ref="M")
	race4 (ref='WHITE')  ethnicity3
	(ref='NOT HISPANIC') Vaccinationsite (ref="PUBLIX OR CVS OR
WALGREENS")/param=reference;
	model CVD1YRD1(ref='0')= FIRSTDOSE agenew sex2 race4 ethnicity3
				decile_sviscore1 decile_sviscore4
				mortalityratepct Vaccinationsite/parmlabel;
	by agenew;
	format agenew age3groups.;
run;
```

```
ods output close;

ods pdf close;

title;

proc print data=Log_agecvdadjusted3g;

run;

data Log_agecvdadjusted3g1;

	set Log_agecvdadjusted3g;

	ODDSRatio1=put(OddsRatioEst,8.3);

	LowerCL1=put(LowerCL,8.3);

	UpperCL1=put(UpperCL,8.3);

	OR=STRIP(ODDSRatio1)||" ("||STRIP(LowerCL1)||", "||STRIP(UpperCL1)||')';

	MODEL=2;

run;

proc print data=Log_agecvdadjusted3g1;

	where SCAN(upcase(EFFECT),1)='FIRSTDOSE';

run;

/*** output excel file for results ***/

libname results xlsx "&path\SAS output\&textadd\CVD DEATHS\CVD DEATHS.xlsx";

/* Export first dataset */

data results.sheet1;

   set work.logcvd;
```

```
            where SCAN(upcase(EFFECT),1)='FIRSTDOSE';
run;

/* Export third dataset */
data results.sheet3;
   set work.log_ageCVDadjusted3g1;
            where SCAN(upcase(EFFECT),1)='FIRSTDOSE';
run;

libname results clear;

/*** COVID DEATHS ***/

/*** 1.a. COVID deaths with categorical age ***/

ods pdf file="&path\SAS output\&textadd\COVID DEATHS\
&textadd Logistic COVID DEATHS &todaysdate..pdf";
ods output ODDSRATIOS=log_COVIDcrude;
proc logistic data=&dataset;
            title "&textadd COVID deaths" ;
            class firstdose (REF=last)/param=reference;
            model COVIDDEATH1YRD1(ref='0')=firstdose/ parmlabel;
run;
ODS OUTPUT CLOSE;
ods pdf close;
```

```
title;

proc print;
run;

data log_COVIDcrude1;
	set log_COVIDcrude;
	ODDSRatio1=put(OddsRatioEst,8.3);
	LowerCL1=put(LowerCL,8.3);
	UpperCL1=put(UpperCL,8.3);
	OR=STRIP(ODDSRatio1)||" ("||STRIP(LowerCL1)||", "||STRIP(UpperCL1)||')';
	MODEL=1;
run;

proc print;
run;

ods pdf file="&path\SAS output\&textadd\COVID DEATHS\
&textadd Logistic COVID Deaths adjusted model (cat age) &todaysdate..pdf";
ods output ODDSRATIOS=Log_COVIDadjusted;
proc logistic data=&dataset;
	TITLE 'COVID Death adjusted (logistic regression)';
	class agenew (ref=first) firstdose (REF=last) sex2 (ref="M")
	race4 (ref='WHITE')  ethnicity3
```

```
        (ref='NOT HISPANIC') Vaccinationsite (ref="PUBLIX OR CVS OR
WALGREENS")/param=reference;

        model COVIDDEATH1YRD1(ref='0')= firstdose agenew sex2 race4 ethnicity3

                        decile_sviscore1 decile_sviscore4

                        mortalityratepct Vaccinationsite/parmlabel;

                format agenew AGE10yGROUP.;

run;

ods output close;

ods pdf close;

title;

proc print;

run;

data Log_COVIDadjusted1;

        set Log_COVIDadjusted;

        ODDSRatio1=put(OddsRatioEst,8.3);

        LowerCL1=put(LowerCL,8.3);

        UpperCL1=put(UpperCL,8.3);

        OR=STRIP(ODDSRatio1)||" ("||STRIP(LowerCL1)||", "||STRIP(UpperCL1)||')';

        MODEL=2;

run;

proc print data=Log_COVIDadjusted1;

        where SCAN(upcase(EFFECT),1)='FIRSTDOSE';

run;
```

```
DATA LOGcovid;
	SET log_covidcrude1 Log_covidadjusted1;
RUN;

PROC PRINT data= LOGcovid;
	where SCAN(upcase(EFFECT),1)='FIRSTDOSE';
RUN;

proc sort data=&dataset;
	by agenew;
run;

ods pdf file="&path\SAS output\&textadd\COVID DEATHS\
&textadd logistic COVID DEATHS adjusted model (cat age) by Age (3 Age groups)
&todaysdate..pdf";
ods output ODDSRATIOS=Log_ageCOVIDadjusted3g;
proc logistic data=&dataset;
	title 'COVID deaths adjusted by Age';
	class agenew (ref=first) FIRSTDOSE (REF=LAST) sex2 (ref="M")
	race4 (ref='WHITE')  ethnicity3
	(ref='NOT HISPANIC') Vaccinationsite (ref="PUBLIX OR CVS OR
WALGREENS")/param=reference;
	model COVIDDEATH1YRD1(ref='0')= FIRSTDOSE agenew sex2 race4 ethnicity3
			decile_sviscore1 decile_sviscore4
			mortalityratepct Vaccinationsite/parmlabel;
	by agenew;
```

```
    format agenew age3groups.;
run;
ods output close;
ods pdf close;
title;

proc print data=Log_ageCOVIDadjusted3g;
run;

data Log_ageCOVIDadjusted3g1;
    set Log_ageCOVIDadjusted3g;
    ODDSRatio1=put(OddsRatioEst,8.3);
    LowerCL1=put(LowerCL,8.3);
    UpperCL1=put(UpperCL,8.3);
    OR=STRIP(ODDSRatio1)||" ("||STRIP(LowerCL1)||", "||STRIP(UpperCL1)||')';
    MODEL=2;
run;

proc print data=Log_ageCOVIDadjusted3g1;
    where SCAN(upcase(EFFECT),1)='FIRSTDOSE';
run;

/*** output excel file for results ***/
libname results xlsx "&path\SAS output\&textadd\COVID DEATHS\COVID DEATHS.xlsx";

/* Export first dataset */
```

```
data results.sheet1;
   set work.logCOVID;
            where SCAN(upcase(EFFECT),1)='FIRSTDOSE';
run;

/* Export third dataset */
data results.sheet3;
   set work.log_ageCOVIDadjusted3g1;
            where SCAN(upcase(EFFECT),1)='FIRSTDOSE';
run;

libname results clear;

/*** NON COVID DEATHS ***/

ods pdf file="&path\SAS output\&textadd\NON COVID DEATHS\
&textadd Logistic NON COVID DEATHS &todaysdate..pdf";
ods output ODDSRATIOS=log_NONCOVIDcrude;
proc logistic data=&dataset;
            title "&textadd NON COVID deaths" ;
            class firstdose (REF=last)/param=reference;
            model NONCOVIDDEATH1YRD1(ref='0')=firstdose/ parmlabel;
run;
```

```
ODS OUTPUT CLOSE;

ods pdf close;

title;

proc print;

run;

data log_nonCOVIDcrude1;

	set log_NONCOVIDcrude;

	ODDSRatio1=put(OddsRatioEst,8.3);

	LowerCL1=put(LowerCL,8.3);

	UpperCL1=put(UpperCL,8.3);

	OR=STRIP(ODDSRatio1)||" ("||STRIP(LowerCL1)||", "||STRIP(UpperCL1)||')';

	MODEL=1;

run;

proc print;

run;

ods pdf file="&path\SAS output\&textadd\NON COVID DEATHS\

&textadd Logistic COVID Deaths adjusted model (cat age) &todaysdate..pdf";

ods output ODDSRATIOS=Log_NONCOVIDadjusted;

proc logistic data=&dataset;

	TITLE 'NON COVID Death adjusted (logistic regression)';

	class agenew (ref=first) firstdose (REF=last) sex2 (ref="M")
```

```
    race4 (ref='WHITE')  ethnicity3
    (ref='NOT HISPANIC') Vaccinationsite (ref="PUBLIX OR CVS OR
WALGREENS")/param=reference;
    model NONCOVIDDEATH1YRD1(ref='0')= firstdose agenew sex2 race4 ethnicity3
                decile_sviscore1 decile_sviscore4
               mortalityratepct Vaccinationsite/parmlabel;
        format agenew AGE10yGROUP.;
run;
ods output close;
ods pdf close;
title;

proc print;
run;

data Log_NONCOVIDadjusted1;
    set Log_NONCOVIDadjusted;
    ODDSRatio1=put(OddsRatioEst,8.3);
    LowerCL1=put(LowerCL,8.3);
    UpperCL1=put(UpperCL,8.3);
    OR=STRIP(ODDSRatio1)||" ("||STRIP(LowerCL1)||", "||STRIP(UpperCL1)||')';
    MODEL=2;
run;

proc print data=Log_NONCOVIDadjusted1;
    where SCAN(upcase(EFFECT),1)='FIRSTDOSE';
```

```
run;

DATA LOGNONcovid;
	SET log_NONcovidcrude1 Log_NONcovidadjusted1;
RUN;

PROC PRINT data= LOGNONcovid;
	where SCAN(upcase(EFFECT),1)='FIRSTDOSE';
RUN;

proc sort data=&dataset;
	by agenew;
run;

ods pdf file="&path\SAS output\&textadd\NON COVID DEATHS\
&textadd logistic Non COVID DEATHS adjusted model (cat age) by Age (3 Age groups) &todaysdate..pdf";
ods output ODDSRATIOS=Log_ageNONCOVIDadjusted3g;
proc logistic data=&dataset;
	title 'NON COVID deaths adjusted by Age';
	class agenew (ref=first) FIRSTDOSE (REF=LAST) sex2 (ref="M")
	race4 (ref='WHITE')  ethnicity3
	(ref='NOT HISPANIC') Vaccinationsite (ref="PUBLIX OR CVS OR WALGREENS")/param=reference;
	model NONCOVIDDEATH1YRD1(ref='0')= FIRSTDOSE agenew sex2 race4 ethnicity3
			decile_sviscore1 decile_sviscore4
			mortalityratepct Vaccinationsite/parmlabel;
```

```
	by agenew;
	format agenew age3groups.;
run;
ods output close;
ods pdf close;
title;

proc print data=Log_ageNONCOVIDadjusted3g;
run;

data Log_ageNONCOVIDadjusted3g1;
	set Log_ageNONCOVIDadjusted3g;
	ODDSRatio1=put(OddsRatioEst,8.3);
	LowerCL1=put(LowerCL,8.3);
	UpperCL1=put(UpperCL,8.3);
	OR=STRIP(ODDSRatio1)||" ("||STRIP(LowerCL1)||", "||STRIP(UpperCL1)||')';
	MODEL=2;
run;

proc print data=Log_ageNONCOVIDadjusted3g1;
	where SCAN(upcase(EFFECT),1)='FIRSTDOSE';
run;

/*** output excel file for results ***/
libname results xlsx "&path\SAS output\&textadd\NON COVID DEATHS\NON COVID DEATHS.xlsx";
```

```
/* Export first dataset */
data results.sheet1;
   set work.logNONCOVID;
         where SCAN(upcase(EFFECT),1)='FIRSTDOSE';
run;

/* Export third dataset */
data results.sheet3;
   set work.log_ageNONCOVIDadjusted3g1;
         where SCAN(upcase(EFFECT),1)='FIRSTDOSE';
run;

libname results clear;

/** On matched cohort **/

DATA MATCHEDCOHORT;
         SET dose1matcheddatafinal;
         WHERE matchedpairs=1;
RUN;

/*** Risk of All-cause, CVD, COVID, and Non-COVID Deaths                ***/
```

```
/*** MATCHED COHORT                    ***/

%let dataset=MATCHEDCOHORT;

%let todaysdate=%sysfunc(today(),date9.);

%put &todaysdate;

%let textadd=Matched cohort Risk of death Dose 1 adj COVINF;
%let path=H:\EPI\Surveillance Section\Fahad Mansuri\SURVIVAL ANALYSIS 2024;

proc contents data=work.MATCHEDCOHORT;
run;

/*** 1.a. SUIC or HOMICIDE  with categorical age ***/

ods pdf file="&path\SAS output\&textadd\SUICIDE or HOMICIDE\
&textadd Logistic SUICHOM &todaysdate..pdf";
ods output ODDSRATIOS=log_SUICHOMcrude;
proc logistic data=&dataset;
	title "&textadd SUICIDE OR HOMICIDE deaths" ;
	class firstdose (REF=last)/param=reference;
	model suicidehomicide1yrD1(ref='0')=firstdose/ parmlabel;
run;
ODS OUTPUT CLOSE;
```

```
ods pdf close;
title;

proc print;
run;

data log_SUICHOMcrude1;
	set log_SUICHOMcrude;
	ODDSRatio1=put(OddsRatioEst,8.3);
	LowerCL1=put(LowerCL,8.3);
	UpperCL1=put(UpperCL,8.3);
	OR=STRIP(ODDSRatio1)||" ("||STRIP(LowerCL1)||", "||STRIP(UpperCL1)||')';
	MODEL=1;
run;

proc print;
run;

ods pdf file="&path\SAS output\&textadd\SUICIDE OR HOMICIDE\
&textadd Logistic SUICHOM adjusted model (cat age) &todaysdate..pdf";
ods output ODDSRATIOS=Log_SUICHOMadjusted;
proc logistic data=&dataset;
	TITLE 'SUICIDE OR HOMICIDE death adjusted (logistic regression)';
	class agenew (ref=first) firstdose (REF=last) sex2 (ref="M")
	race4 (ref='WHITE')  ethnicity3
```

```
	(ref='NOT HISPANIC') Vaccinationsite (ref="PUBLIX OR CVS OR
WALGREENS")/param=reference;
	model suicidehomicide1yrD1(ref='0')= firstdose agenew sex2 race4 ethnicity3
			decile_sviscore1 decile_sviscore4
			mortalityratepct Vaccinationsite covidinf/parmlabel;
		format agenew AGE10yGROUP.;
run;
ods output close;
ods pdf close;
title;

proc print;
run;

data Log_SUICHOMadjusted1;
	set Log_SUICHOMadjusted;
	ODDSRatio1=put(OddsRatioEst,8.3);
	LowerCL1=put(LowerCL,8.3);
	UpperCL1=put(UpperCL,8.3);
	OR=STRIP(ODDSRatio1)||" ("||STRIP(LowerCL1)||", "||STRIP(UpperCL1)||')';
	MODEL=2;
run;

proc print data=Log_SUICHOMadjusted1;
	where SCAN(upcase(EFFECT),1)='FIRSTDOSE';
run;
```

```
DATA LOGSUICHOM;

	SET lOG_SUICHOMCRUDE1 Log_SUICHOMadjusted1;

RUN;

PROC PRINT data= LOGSUICHOM;

	where SCAN(upcase(EFFECT),1)='FIRSTDOSE';

RUN;

proc sort data=&dataset;

	by agenew;

run;

ods pdf file="&path\SAS output\&textadd\SUICIDE OR HOMICIDE\

&textadd logistic SUICHOM adjusted model (cat age) by Age (3 Age groups)
&todaysdate..pdf";

ods output ODDSRATIOS=Log_ageSUICHOMadjusted3g;

proc logistic data=&dataset;

	title 'SUICIDE OR HOMICIDE deaths adjusted by Age';

	class agenew (ref=first) FIRSTDOSE (REF=LAST) sex2 (ref="M")

	race4 (ref='WHITE')  ethnicity3

	(ref='NOT HISPANIC') Vaccinationsite (ref="PUBLIX OR CVS OR
WALGREENS")/param=reference;

	model suicidehomicide1yrD1(ref='0')= FIRSTDOSE agenew sex2 race4 ethnicity3

				decile_sviscore1 decile_sviscore4

				mortalityratepct Vaccinationsite covidinf/parmlabel;

	by agenew;
```

```
    format agenew age3groups.;
run;
ods output close;
ods pdf close;
title;

proc print data=Log_ageSUICHOMadjusted3g;
run;

data Log_ageSUICHOMadjusted3g1;
    set Log_ageSUICHOMadjusted3g;
    ODDSRatio1=put(OddsRatioEst,8.3);
    LowerCL1=put(LowerCL,8.3);
    UpperCL1=put(UpperCL,8.3);
    OR=STRIP(ODDSRatio1)||" ("||STRIP(LowerCL1)||", "||STRIP(UpperCL1)||')';
    MODEL=2;
run;

proc print data=Log_ageSUICHOMadjusted3g1;
    where SCAN(upcase(EFFECT),1)='FIRSTDOSE';
run;

/*** output excel file for results ***/
libname results xlsx "&path\SAS output\&textadd\SUICIDE OR HOMICIDE\SUICIDE OR
HOMICIDE.xlsx";
```

```
/* Export first dataset */
data results.sheet1;
   set work.logSUICHOM;
        where SCAN(upcase(EFFECT),1)='FIRSTDOSE';
run;

/* Export third dataset */
data results.sheet3;
   set work.Log_ageSUICHOMadjusted3g1;
        where SCAN(upcase(EFFECT),1)='FIRSTDOSE';
run;

libname results clear;

/*** 1.a. NON COVID +  with categorical age ***/

ods pdf file="&path\SAS output\&textadd\NON COVID PLUS\
&textadd Logistic COVID + &todaysdate..pdf";
ods output ODDSRATIOS=log_NONCOV2crude;
proc logistic data=&dataset;
        title "&textadd NON COVID + deaths" ;
        class firstdose (REF=last)/param=reference;
        model NONCOVIDDEATHPLUS1YRD1(ref='0')=firstdose/ parmlabel;
run;
ODS OUTPUT CLOSE;
ods pdf close;
```

```
title;

proc print;
run;

data Log_NONCOV2crude1;
	set Log_NONCOV2crude;
	ODDSRatio1=put(OddsRatioEst,8.3);
	LowerCL1=put(LowerCL,8.3);
	UpperCL1=put(UpperCL,8.3);
	OR=STRIP(ODDSRatio1)||" ("||STRIP(LowerCL1)||", "||STRIP(UpperCL1)||')';
	MODEL=1;
run;

proc print;
run;

ods pdf file="&path\SAS output\&textadd\NON COVID PLUS\
&textadd Logistic NON COVID + adjusted model (cat age) &todaysdate..pdf";
ods output ODDSRATIOS=Log_NONCOV2adjusted;
proc logistic data=&dataset;
	TITLE 'NON COVID + death adjusted (logistic regression)';
	class agenew (ref=first) firstdose (REF=last) sex2 (ref="M")
	race4 (ref='WHITE')  ethnicity3
```

```
	(ref='NOT HISPANIC') Vaccinationsite (ref="PUBLIX OR CVS OR WALGREENS")/param=reference;
	model NONCOVIDDEATHPLUS1YRD1(ref='0')= firstdose agenew sex2 race4 ethnicity3
				decile_sviscore1 decile_sviscore4
				mortalityratepct Vaccinationsite covidinf/parmlabel;
		format agenew AGE10yGROUP.;
run;
ods output close;
ods pdf close;
title;

proc print;
run;

data Log_NONCOV2adjusted1;
	set Log_NONCOV2adjusted;
	ODDSRatio1=put(OddsRatioEst,8.3);
	LowerCL1=put(LowerCL,8.3);
	UpperCL1=put(UpperCL,8.3);
	OR=STRIP(ODDSRatio1)||" ("||STRIP(LowerCL1)||", "||STRIP(UpperCL1)||')';
	MODEL=2;
run;

proc print data=Log_NONCOV2adjusted1;
	where SCAN(upcase(EFFECT),1)='VACCINEGROUP2';
run;
```

```
DATA LOGNONCOV2;
	SET lOG_NONCOV2CRUDE1 lOG_NONCOV2adjusted1;
RUN;

PROC PRINT data= LOGNONCOV2;
	where SCAN(upcase(EFFECT),1)='FIRSTDOSE';
RUN;

proc sort data=&dataset;
	by agenew;
run;

ods pdf file="&path\SAS output\&textadd\NON COVID PLUS\
&textadd logistic NON COVID + adjusted model (cat age) by Age (3 Age groups)
&todaysdate..pdf";
ods output ODDSRATIOS=Log_ageNONCOV2adjusted3g;
proc logistic data=&dataset;
	title 'NON COVID + deaths adjusted by Age';
	class agenew (ref=first) FIRSTDOSE (REF=LAST) sex2 (ref="M")
	race4 (ref='WHITE')  ethnicity3
	(ref='NOT HISPANIC') Vaccinationsite (ref="PUBLIX OR CVS OR
WALGREENS")/param=reference;
	model NONCOVIDDEATHPLUS1YRD1(ref='0')= FIRSTDOSE agenew sex2 race4
ethnicity3
				decile_sviscore1 decile_sviscore4
				mortalityratepct Vaccinationsite covidinf/parmlabel;
```

```
        by agenew;
        format agenew age3groups.;
run;
ods output close;
ods pdf close;
title;

proc print data=Log_ageNONCOV2adjusted3g;
run;

data Log_ageNONCOV2adjusted3g1;
        set Log_ageNONCOV2adjusted3g;
        ODDSRatio1=put(OddsRatioEst,8.3);
        LowerCL1=put(LowerCL,8.3);
        UpperCL1=put(UpperCL,8.3);
        OR=STRIP(ODDSRatio1)||" ("||STRIP(LowerCL1)||", "||STRIP(UpperCL1)||')';
        MODEL=2;
run;

proc print data=Log_ageNONCOV2adjusted3g1;
        where SCAN(upcase(EFFECT),1)='FIRSTDOSE';
run;

/*** output excel file for results ***/
libname results xlsx "&path\SAS output\&textadd\NON COVID PLUS\NON COVID
PLUS.xlsx";
```

```
/* Export first dataset */
data results.sheet1;
   set work.logNONCOV2;
         where SCAN(upcase(EFFECT),1)='FIRSTDOSE';
run;

/* Export third dataset */
data results.sheet3;
   set work.Log_ageNONCOV2adjusted3g1;
         where SCAN(upcase(EFFECT),1)='FIRSTDOSE';
run;

libname results clear;

/*** 1.a. COVID +  with categorical age ***/

ods pdf file="&path\SAS output\&textadd\COVID PLUS\
&textadd Logistic COVID + &todaysdate..pdf";
ods output ODDSRATIOS=log_COV2crude;
proc logistic data=&dataset;
         title "&textadd COVID + deaths" ;
         class firstdose (REF=last)/param=reference;
         model COVIDDEATHPLUS1YRD1(ref='0')=firstdose/ parmlabel;
run;
```

```
ODS OUTPUT CLOSE;

ods pdf close;

title;

proc print;

run;

data Log_COV2crude1;

        set Log_COV2crude;

        ODDSRatio1=put(OddsRatioEst,8.3);

        LowerCL1=put(LowerCL,8.3);

        UpperCL1=put(UpperCL,8.3);

        OR=STRIP(ODDSRatio1)||" ("||STRIP(LowerCL1)||", "||STRIP(UpperCL1)||')';

        MODEL=1;

run;

proc print;

run;

ods pdf file="&path\SAS output\&textadd\COVID PLUS\

&textadd Logistic COVID + adjusted model (cat age) &todaysdate..pdf";

ods output ODDSRATIOS=Log_COV2adjusted;

proc logistic data=&dataset;

        TITLE 'COVID + death adjusted (logistic regression)';

        class agenew (ref=first) firstdose (REF=last) sex2 (ref="M")
```

```
            race4 (ref='WHITE')  ethnicity3
            (ref='NOT HISPANIC') Vaccinationsite (ref="PUBLIX OR CVS OR
WALGREENS")/param=reference;
            model COVIDDEATHPLUS1YRD1(ref='0')= firstdose agenew sex2 race4 ethnicity3
                              decile_sviscore1 decile_sviscore4
                             mortalityratepct Vaccinationsite covidinf/parmlabel;
                  format agenew AGE10yGROUP.;
run;
ods output close;
ods pdf close;
title;

proc print;
run;

data Log_COV2adjusted1;
            set Log_COV2adjusted;
            ODDSRatio1=put(OddsRatioEst,8.3);
            LowerCL1=put(LowerCL,8.3);
            UpperCL1=put(UpperCL,8.3);
            OR=STRIP(ODDSRatio1)||" ("||STRIP(LowerCL1)||", "||STRIP(UpperCL1)||')';
            MODEL=2;
run;

proc print data=Log_COV2adjusted1;
            where SCAN(upcase(EFFECT),1)='VACCINEGROUP2';
```

```
run;

DATA LOGCOV2;
	SET lOG_COV2CRUDE1 lOG_COV2adjusted1;
RUN;

PROC PRINT data= LOGCOV2;
	where SCAN(upcase(EFFECT),1)='FIRSTDOSE';
RUN;

proc sort data=&dataset;
	by agenew;
run;

ods pdf file="&path\SAS output\&textadd\COVID PLUS\
&textadd logistic COVID + adjusted model (cat age) by Age (3 Age groups)
&todaysdate..pdf";
ods output ODDSRATIOS=Log_ageCOV2adjusted3g;
proc logistic data=&dataset;
	title 'COVID + deaths adjusted by Age';
	class agenew (ref=first) FIRSTDOSE (REF=LAST) sex2 (ref="M")
	race4 (ref='WHITE')  ethnicity3
	(ref='NOT HISPANIC') Vaccinationsite (ref="PUBLIX OR CVS OR
WALGREENS")/param=reference;
	model COVIDDEATHPLUS1YRD1(ref='0')= FIRSTDOSE agenew sex2 race4 ethnicity3
			decile_sviscore1 decile_sviscore4
			mortalityratepct Vaccinationsite covidinf/parmlabel;
```

```
        by agenew;
        format agenew age3groups.;
run;
ods output close;
ods pdf close;
title;

proc print data=Log_ageCOV2adjusted3g;
run;

data Log_ageCOV2adjusted3g1;
        set Log_ageCOV2adjusted3g;
        ODDSRatio1=put(OddsRatioEst,8.3);
        LowerCL1=put(LowerCL,8.3);
        UpperCL1=put(UpperCL,8.3);
        OR=STRIP(ODDSRatio1)||" ("||STRIP(LowerCL1)||", "||STRIP(UpperCL1)||')';
        MODEL=2;
run;

proc print data=Log_ageCOV2adjusted3g1;
        where SCAN(upcase(EFFECT),1)='FIRSTDOSE';
run;

/*** output excel file for results ***/
libname results xlsx "&path\SAS output\&textadd\COVID PLUS\COVID PLUS.xlsx";
```

```
/* Export first dataset */
data results.sheet1;
   set work.logCOV2;
          where SCAN(upcase(EFFECT),1)='FIRSTDOSE';
run;

/* Export third dataset */
data results.sheet3;
   set work.Log_ageCOV2adjusted3g1;
          where SCAN(upcase(EFFECT),1)='FIRSTDOSE';
run;

libname results clear;

/*** 1.a. All deaths with categorical age ***/

ods pdf file="&path\SAS output\&textadd\All cause\
&textadd Logistic All-cause &todaysdate..pdf";
ods output ODDSRATIOS=log_allcrude;
proc logistic data=&dataset;
          title "&textadd All-cause deaths" ;
          class firstdose (REF=last)/param=reference;
          model DEATH1YRd1(ref='0')=firstdose/ parmlabel;
run;
ODS OUTPUT CLOSE;
ods pdf close;
```

```
title;

proc print;
run;

data Log_allcrude1;
	set Log_allcrude;
	ODDSRatio1=put(OddsRatioEst,8.3);
	LowerCL1=put(LowerCL,8.3);
	UpperCL1=put(UpperCL,8.3);
	OR=STRIP(ODDSRatio1)||" ("||STRIP(LowerCL1)||", "||STRIP(UpperCL1)||')';
	MODEL=1;
run;

proc print;
run;

ods pdf file="&path\SAS output\&textadd\All cause\
&textadd Logistic All-cause adjusted model (cat age) &todaysdate..pdf";
ods output ODDSRATIOS=Log_alladjusted;
proc logistic data=&dataset;
	TITLE 'All-cause death adjusted (logistic regression)';
	class agenew (ref=first) firstdose (REF=last) sex2 (ref="M")
	race4 (ref='WHITE')  ethnicity3
```

```
        (ref='NOT HISPANIC') Vaccinationsite (ref="PUBLIX OR CVS OR
WALGREENS")/param=reference;
        model DEATH1YRd1(ref='0')= firstdose agenew sex2 race4 ethnicity3
                         decile_sviscore1 decile_sviscore4
                        mortalityratepct Vaccinationsite covidinf/parmlabel;
            format agenew AGE10yGROUP.;
run;
ods output close;
ods pdf close;
title;

proc print;
run;

data Log_alladjusted1;
        set log_alladjusted;
        ODDSRatio1=put(OddsRatioEst,8.3);
        LowerCL1=put(LowerCL,8.3);
        UpperCL1=put(UpperCL,8.3);
        OR=STRIP(ODDSRatio1)||" ("||STRIP(LowerCL1)||", "||STRIP(UpperCL1)||')';
        MODEL=2;
run;

proc print data=Log_alladjusted1;
        where SCAN(upcase(EFFECT),1)='VACCINEGROUP2';
run;
```

```
DATA LOGALL;
	SET lOG_ALLCRUDE1 lOG_alladjusted1;
RUN;

PROC PRINT data= logall;
	where SCAN(upcase(EFFECT),1)='FIRSTDOSE';
RUN;

proc sort data=&dataset;
	by agenew;
run;

ods pdf file="&path\SAS output\&textadd\All cause\
&textadd logistic All-cause adjusted model (cat age) by Age (3 Age groups)
&todaysdate..pdf";
ods output ODDSRATIOS=Log_agealladjusted3g;
proc logistic data=&dataset;
	title 'All-cause deaths adjusted by Age';
	class agenew (ref=first) FIRSTDOSE (REF=LAST) sex2 (ref="M")
	race4 (ref='WHITE')  ethnicity3
	(ref='NOT HISPANIC') Vaccinationsite (ref="PUBLIX OR CVS OR
WALGREENS")/param=reference;
	model DEATH1YRd1(ref='0')= FIRSTDOSE agenew sex2 race4 ethnicity3
				decile_sviscore1 decile_sviscore4
				mortalityratepct Vaccinationsite covidinf/parmlabel;
	by agenew;
```

```
	format agenew age3groups.;
run;
ods output close;
ods pdf close;
title;

proc print data=Log_agealladjusted3g;
run;

data Log_agealladjusted3g1;
	set Log_agealladjusted3g;
	ODDSRatio1=put(OddsRatioEst,8.3);
	LowerCL1=put(LowerCL,8.3);
	UpperCL1=put(UpperCL,8.3);
	OR=STRIP(ODDSRatio1)||" ("||STRIP(LowerCL1)||", "||STRIP(UpperCL1)||')';
	MODEL=2;
run;

proc print data=Log_agealladjusted3g1;
	where SCAN(upcase(EFFECT),1)='FIRSTDOSE';
run;

/*** output excel file for results ***/
libname results xlsx "&path\SAS output\&textadd\All cause\allcause.xlsx";

/* Export first dataset */
```

```
data results.sheet1;
   set work.logall;
          where SCAN(upcase(EFFECT),1)='FIRSTDOSE';
run;

/* Export third dataset */
data results.sheet3;
   set work.log_agealladjusted3g1;
          where SCAN(upcase(EFFECT),1)='FIRSTDOSE';
run;

libname results clear;

/*** 1.a. CVD deaths with categorical age ***/

ods pdf file="&path\SAS output\&textadd\CVD DEATHS\
&textadd Logistic CVD DEATHS &todaysdate..pdf";
ods output ODDSRATIOS=log_cvdcrude;
proc logistic data=&dataset;
          title "&textadd CVD deaths" ;
          class firstdose (REF=last)/param=reference;
          model CVD1YRD1(ref='0')=firstdose/ parmlabel;
run;
ODS OUTPUT CLOSE;
ods pdf close;
title;
```

```
proc print;

run;

data log_cvdcrude1;

	set log_cvdcrude;

	ODDSRatio1=put(OddsRatioEst,8.3);

	LowerCL1=put(LowerCL,8.3);

	UpperCL1=put(UpperCL,8.3);

	OR=STRIP(ODDSRatio1)||" ("||STRIP(LowerCL1)||", "||STRIP(UpperCL1)||')';

	MODEL=1;

run;

proc print;

run;

ods pdf file="&path\SAS output\&textadd\CVD DEATHS\

&textadd Logistic CVD Deaths adjusted model (cat age) &todaysdate..pdf";

ods output ODDSRATIOS=Log_cvdadjusted;

proc logistic data=&dataset;

	TITLE 'CVD Death adjusted (logistic regression)';

	class agenew (ref=first) firstdose (REF=last) sex2 (ref="M")

	race4 (ref='WHITE')  ethnicity3

	(ref='NOT HISPANIC') Vaccinationsite (ref="PUBLIX OR CVS OR
WALGREENS")/param=reference;
```

```
	model CVD1YRD1(ref='0')= firstdose agenew sex2 race4 ethnicity3
				decile_sviscore1 decile_sviscore4
				mortalityratepct Vaccinationsite covidinf/parmlabel;
		format agenew AGE10yGROUP.;
run;
ods output close;
ods pdf close;
title;

proc print;
run;

data Log_cvdadjusted1;
	set Log_cvdadjusted;
	ODDSRatio1=put(OddsRatioEst,8.3);
	LowerCL1=put(LowerCL,8.3);
	UpperCL1=put(UpperCL,8.3);
	OR=STRIP(ODDSRatio1)||" ("||STRIP(LowerCL1)||", "||STRIP(UpperCL1)||')';
	MODEL=2;
run;

proc print data=Log_cvdadjusted1;
	where SCAN(upcase(EFFECT),1)='FIRSTDOSE';
run;

DATA LOGCVD;
```

```
    SET log_cvdcrude1 Log_cvdadjusted1;
RUN;

PROC PRINT data= LOGCVD;
    where SCAN(upcase(EFFECT),1)='FIRSTDOSE';
RUN;

proc sort data=&dataset;
    by agenew;
run;

ods pdf file="&path\SAS output\&textadd\CVD DEATHS\
&textadd logistic CVD DEATHS adjusted model (cat age) by Age (3 Age groups)
&todaysdate..pdf";
ods output ODDSRATIOS=Log_agecvdadjusted3g;
proc logistic data=&dataset;
    title 'CVD deaths adjusted by Age';
    class agenew (ref=first) FIRSTDOSE (REF=LAST) sex2 (ref="M")
    race4 (ref='WHITE')  ethnicity3
    (ref='NOT HISPANIC') Vaccinationsite (ref="PUBLIX OR CVS OR
WALGREENS")/param=reference;
    model CVD1YRD1(ref='0')= FIRSTDOSE agenew sex2 race4 ethnicity3
                decile_sviscore1 decile_sviscore4
               mortalityratepct Vaccinationsite covidinf/parmlabel;
    by agenew;
    format agenew age3groups.;
run;
```

```
ods output close;

ods pdf close;

title;

proc print data=Log_agecvdadjusted3g;

run;

data Log_agecvdadjusted3g1;

	set Log_agecvdadjusted3g;

	ODDSRatio1=put(OddsRatioEst,8.3);

	LowerCL1=put(LowerCL,8.3);

	UpperCL1=put(UpperCL,8.3);

	OR=STRIP(ODDSRatio1)||" ("||STRIP(LowerCL1)||", "||STRIP(UpperCL1)||')';

	MODEL=2;

run;

proc print data=Log_agecvdadjusted3g1;

	where SCAN(upcase(EFFECT),1)='FIRSTDOSE';

run;

/*** output excel file for results ***/

libname results xlsx "&path\SAS output\&textadd\CVD DEATHS\CVD DEATHS.xlsx";

/* Export first dataset */

data results.sheet1;

   set work.logcvd;
```

```
            where SCAN(upcase(EFFECT),1)='FIRSTDOSE';
run;

/* Export third dataset */
data results.sheet3;
   set work.log_ageCVDadjusted3g1;
            where SCAN(upcase(EFFECT),1)='FIRSTDOSE';
run;

libname results clear;

/*** COVID DEATHS ***/

/*** 1.a. COVID deaths with categorical age ***/

ods pdf file="&path\SAS output\&textadd\COVID DEATHS\
&textadd Logistic COVID DEATHS &todaysdate..pdf";
ods output ODDSRATIOS=log_COVIDcrude;
proc logistic data=&dataset;
            title "&textadd COVID deaths" ;
            class firstdose (REF=last)/param=reference;
            model COVIDDEATH1YRD1(ref='0')=firstdose/ parmlabel;
run;
ODS OUTPUT CLOSE;
ods pdf close;
```

```
title;

proc print;
run;

data log_COVIDcrude1;
	set log_COVIDcrude;
	ODDSRatio1=put(OddsRatioEst,8.3);
	LowerCL1=put(LowerCL,8.3);
	UpperCL1=put(UpperCL,8.3);
	OR=STRIP(ODDSRatio1)||" ("||STRIP(LowerCL1)||", "||STRIP(UpperCL1)||')';
	MODEL=1;
run;

proc print;
run;

ods pdf file="&path\SAS output\&textadd\COVID DEATHS\
&textadd Logistic COVID Deaths adjusted model (cat age) &todaysdate..pdf";
ods output ODDSRATIOS=Log_COVIDadjusted;
proc logistic data=&dataset;
	TITLE 'COVID Death adjusted (logistic regression)';
	class agenew (ref=first) firstdose (REF=last) sex2 (ref="M")
	race4 (ref='WHITE')  ethnicity3
```

```
        (ref='NOT HISPANIC') Vaccinationsite (ref="PUBLIX OR CVS OR
WALGREENS")/param=reference;
        model COVIDDEATH1YRD1(ref='0')= firstdose agenew sex2 race4 ethnicity3
                  decile_sviscore1 decile_sviscore4
                 mortalityratepct Vaccinationsite covidinf/parmlabel;
            format agenew AGE10yGROUP.;
run;
ods output close;
ods pdf close;
title;

proc print;
run;

data Log_COVIDadjusted1;
        set Log_COVIDadjusted;
        ODDSRatio1=put(OddsRatioEst,8.3);
        LowerCL1=put(LowerCL,8.3);
        UpperCL1=put(UpperCL,8.3);
        OR=STRIP(ODDSRatio1)||" ("||STRIP(LowerCL1)||", "||STRIP(UpperCL1)||')';
        MODEL=2;
run;

proc print data=Log_COVIDadjusted1;
        where SCAN(upcase(EFFECT),1)='FIRSTDOSE';
run;
```

```
DATA LOGcovid;
	SET log_covidcrude1 Log_covidadjusted1;
RUN;

PROC PRINT data= LOGcovid;
	where SCAN(upcase(EFFECT),1)='FIRSTDOSE';
RUN;

proc sort data=&dataset;
	by agenew;
run;

ods pdf file="&path\SAS output\&textadd\COVID DEATHS\
&textadd logistic COVID DEATHS adjusted model (cat age) by Age (3 Age groups)
&todaysdate..pdf";
ods output ODDSRATIOS=Log_ageCOVIDadjusted3g;
proc logistic data=&dataset;
	title 'COVID deaths adjusted by Age';
	class agenew (ref=first) FIRSTDOSE (REF=LAST) sex2 (ref="M")
	race4 (ref='WHITE')  ethnicity3
	(ref='NOT HISPANIC') Vaccinationsite (ref="PUBLIX OR CVS OR
WALGREENS")/param=reference;
	model COVIDDEATH1YRD1(ref='0')= FIRSTDOSE agenew sex2 race4 ethnicity3
			decile_sviscore1 decile_sviscore4
			mortalityratepct Vaccinationsite covidinf/parmlabel;
	by agenew;
```

```
    format agenew age3groups.;
run;
ods output close;
ods pdf close;
title;

proc print data=Log_ageCOVIDadjusted3g;
run;

data Log_ageCOVIDadjusted3g1;
    set Log_ageCOVIDadjusted3g;
    ODDSRatio1=put(OddsRatioEst,8.3);
    LowerCL1=put(LowerCL,8.3);
    UpperCL1=put(UpperCL,8.3);
    OR=STRIP(ODDSRatio1)||" ("||STRIP(LowerCL1)||", "||STRIP(UpperCL1)||')';
    MODEL=2;
run;

proc print data=Log_ageCOVIDadjusted3g1;
    where SCAN(upcase(EFFECT),1)='FIRSTDOSE';
run;

/*** output excel file for results ***/
libname results xlsx "&path\SAS output\&textadd\COVID DEATHS\COVID DEATHS.xlsx";

/* Export first dataset */
```

```
data results.sheet1;
   set work.logCOVID;
            where SCAN(upcase(EFFECT),1)='FIRSTDOSE';
run;

/* Export third dataset */
data results.sheet3;
   set work.log_ageCOVIDadjusted3g1;
            where SCAN(upcase(EFFECT),1)='FIRSTDOSE';
run;

libname results clear;

/*** NON COVID DEATHS ***/

ods pdf file="&path\SAS output\&textadd\NON COVID DEATHS\
&textadd Logistic NON COVID DEATHS &todaysdate..pdf";
ods output ODDSRATIOS=log_NONCOVIDcrude;
proc logistic data=&dataset;
            title "&textadd NON COVID deaths" ;
            class firstdose (REF=last)/param=reference;
            model NONCOVIDDEATH1YRD1(ref='0')=firstdose/ parmlabel;
run;
```

```
ODS OUTPUT CLOSE;
ods pdf close;
title;

proc print;
run;

data log_nonCOVIDcrude1;
	set log_NONCOVIDcrude;
	ODDSRatio1=put(OddsRatioEst,8.3);
	LowerCL1=put(LowerCL,8.3);
	UpperCL1=put(UpperCL,8.3);
	OR=STRIP(ODDSRatio1)||" ("||STRIP(LowerCL1)||", "||STRIP(UpperCL1)||')';
	MODEL=1;
run;

proc print;
run;

ods pdf file="&path\SAS output\&textadd\NON COVID DEATHS\
&textadd Logistic COVID Deaths adjusted model (cat age) &todaysdate..pdf";
ods output ODDSRATIOS=Log_NONCOVIDadjusted;
proc logistic data=&dataset;
	TITLE 'NON COVID Death adjusted (logistic regression)';
	class agenew (ref=first) firstdose (REF=last) sex2 (ref="M")
```

```
            race4 (ref='WHITE')  ethnicity3
            (ref='NOT HISPANIC') Vaccinationsite (ref="PUBLIX OR CVS OR
WALGREENS")/param=reference;
            model NONCOVIDDEATH1YRD1(ref='0')= firstdose agenew sex2 race4 ethnicity3
                         decile_sviscore1 decile_sviscore4
                        mortalityratepct Vaccinationsite covidinf/parmlabel;
                format agenew AGE10yGROUP.;
run;
ods output close;
ods pdf close;
title;

proc print;
run;

data Log_NONCOVIDadjusted1;
            set Log_NONCOVIDadjusted;
            ODDSRatio1=put(OddsRatioEst,8.3);
            LowerCL1=put(LowerCL,8.3);
            UpperCL1=put(UpperCL,8.3);
            OR=STRIP(ODDSRatio1)||" ("||STRIP(LowerCL1)||", "||STRIP(UpperCL1)||')';
            MODEL=2;
run;

proc print data=Log_NONCOVIDadjusted1;
            where SCAN(upcase(EFFECT),1)='FIRSTDOSE';
```

```
run;

DATA LOGNONcovid;
	SET log_NONcovidcrude1 Log_NONcovidadjusted1;
RUN;

PROC PRINT data= LOGNONcovid;
	where SCAN(upcase(EFFECT),1)='FIRSTDOSE';
RUN;

proc sort data=&dataset;
	by agenew;
run;

ods pdf file="&path\SAS output\&textadd\NON COVID DEATHS\
&textadd logistic Non COVID DEATHS adjusted model (cat age) by Age (3 Age groups)
&todaysdate..pdf";
ods output ODDSRATIOS=Log_ageNONCOVIDadjusted3g;
proc logistic data=&dataset;
	title 'NON COVID deaths adjusted by Age';
	class agenew (ref=first) FIRSTDOSE (REF=LAST) sex2 (ref="M")
	race4 (ref='WHITE')  ethnicity3
	(ref='NOT HISPANIC') Vaccinationsite (ref="PUBLIX OR CVS OR
WALGREENS")/param=reference;
	model NONCOVIDDEATH1YRD1(ref='0')= FIRSTDOSE agenew sex2 race4 ethnicity3
				decile_sviscore1 decile_sviscore4
				mortalityratepct Vaccinationsite covidinf/parmlabel;
```

```
        by agenew;
        format agenew age3groups.;
run;
ods output close;
ods pdf close;
title;

proc print data=Log_ageNONCOVIDadjusted3g;
run;

data Log_ageNONCOVIDadjusted3g1;
        set Log_ageNONCOVIDadjusted3g;
        ODDSRatio1=put(OddsRatioEst,8.3);
        LowerCL1=put(LowerCL,8.3);
        UpperCL1=put(UpperCL,8.3);
        OR=STRIP(ODDSRatio1)||" ("||STRIP(LowerCL1)||", "||STRIP(UpperCL1)||')';
        MODEL=2;
run;

proc print data=Log_ageNONCOVIDadjusted3g1;
        where SCAN(upcase(EFFECT),1)='FIRSTDOSE';
run;

/*** output excel file for results ***/
libname results xlsx "&path\SAS output\&textadd\NON COVID DEATHS\NON COVID
DEATHS.xlsx";
```

```
/* Export first dataset */
data results.sheet1;
   set work.logNONCOVID;
          where SCAN(upcase(EFFECT),1)='FIRSTDOSE';
run;

/* Export third dataset */
data results.sheet3;
   set work.log_ageNONCOVIDadjusted3g1;
          where SCAN(upcase(EFFECT),1)='FIRSTDOSE';
run;

libname results clear;

/*** Calculating discordant pairs for the sensitivity analysis ***/

proc sort data=store.dose1matcheddatawithid out=dose1matcheddatawithid;
          by matchx firstdose;
run;

proc freq data=dose1matcheddatawithid;
          table death1yrd1;
run;

proc transpose data=dose1matcheddatawithid out=test1;
```

```
	by matchx;
	var death1yrd1;
	id firstdose;
run;

proc freq data=test1;
	table '1. pfizer'n*'2. modern'n/crosslist;
run;

data test1;
  set test1;
  Diff= "1. PFIZER"n -"2. MODERNA"N;
run;

/** On matched cohort **/

DATA MATCHEDCOHORT;
	SET dose1matcheddatafinal;
	WHERE matchedpairs=1;
RUN;

proc freq data=work.matchedcohort;
	table firstdose*covinfbf20dec/crosslist;
run;
```

```
/*** Risk of All-cause, CVD, COVID, and Non-COVID Deaths                    ***/

/*** MATCHED COHORT                    ***/

%let dataset=MATCHEDCOHORT;

%let todaysdate=%sysfunc(today(),date9.);

%put &todaysdate;

%let textadd=Matched cohort Risk of death Dose 1;
%let path=H:\EPI\Surveillance Section\Fahad Mansuri\SURVIVAL ANALYSIS 2024;

proc contents data=work.MATCHEDCOHORT;
run;

/*** COVID infections before 20 DEC 2020            ***/

ods pdf file="&path\SAS output\&textadd\COVID INFECTIONS\
&textadd Logistic COVID INFECTIONS crude &todaysdate..pdf";
ods output oddsratios=Log_COVIDINFcrude;
proc logistic data=&dataset;
        title 'COVID infections crude model';
        class firstdose (REF=last)/param=reference;
```

```
	model covinfbf20dec(ref='0')=firstdose/ parmlabel;
run;
ods output close;
ods pdf close;
title;

proc print;
run;

data Log_COVIDINFcrude1;
	set Log_COVIDINFcrude;
	ODDSRatio1=put(OddsRatioEst,8.3);
	LowerCL1=put(LowerCL,8.3);
	UpperCL1=put(UpperCL,8.3);
	OR=STRIP(ODDSRatio1)||" ("||STRIP(LowerCL1)||", "||STRIP(UpperCL1)||')';
	MODEL=1;
run;

proc print;
run;

ods pdf file="&path\SAS output\&textadd\COVID INFECTIONS\
&textadd logistic COVID infections adjusted model (cat age) &todaysdate..pdf";
ods output oddsratios=log_COVIDINFadjusted;
proc logistic data=&dataset;
	title 'COVID infections adjusted model logistic';
```

```
	class agenew (ref=first) FIRSTDOSE (REF=LAST) sex2 (ref="M")
	race4 (ref='WHITE')  ethnicity3
	(ref='NOT HISPANIC') Vaccinationsite (ref="PUBLIX OR CVS OR WALGREENS")/param=reference;
	model covinfbf20dec(ref='0')= FIRSTDOSE agenew sex2 race4 ethnicity3
	                decile_sviscore1 decile_sviscore4
	               mortalityratepct Vaccinationsite covidinf/parmlabel;
	format agenew age10ygroup.;
run;
ods output close;
ods pdf close;

proc print;
run;

data log_COVIDINFadjusted1;
	set log_COVIDINFadjusted;
	ODDSRatio1=put(OddsRatioEst,8.3);
	LowerCL1=put(LowerCL,8.3);
	UpperCL1=put(UpperCL,8.3);
	OR=STRIP(ODDSRatio1)||" ("||STRIP(LowerCL1)||", "||STRIP(UpperCL1)||')';
	MODEL=2;
run;

proc print;
run;
```

```
DATA Log_COVIDinfALL;
	SET log_COVIDinfCRUDE1 log_COVIDinfadjusted1;
RUN;

PROC PRINT data=log_COVIDinfALL;
	where SCAN(upcase(EFFECT),1)='FIRSTDOSE';
RUN;

proc sort data=&dataset;
	by agenew;
run;

ods pdf file="&path\SAS output\&textadd\COVID INFECTIONS\
&textadd logistic COVID INFECTIONS by age &todaysdate..pdf";
ods output oddsratios=log_covidinfagealladjusted3g;
proc logistic data=&dataset;
	title 'COVID INFECTIONS adjusted model by Age (3 groups) logistic model';
	class agenew (ref=first) FIRSTDOSE (REF=LAST) sex2 (ref="M")
	race4 (ref='WHITE')  ethnicity3
	(ref='NOT HISPANIC') Vaccinationsite (ref="PUBLIX OR CVS OR
WALGREENS")/param=reference;
	model covinfbf20dec(ref='0')= FIRSTDOSE agenew sex2 race4 ethnicity3
				decile_sviscore1 decile_sviscore4
				mortalityratepct Vaccinationsite covidinf/parmlabel;
	by agenew;
```

```
	format agenew age3groups.;
run;
ods output close;
ods pdf close;

proc print data=log_covidinfagealladjusted3g;
run;

data log_covidinfagealladjusted3g1;
	set log_covidinfagealladjusted3g;
	ODDSRatio1=put(OddsRatioEst,8.3);
	LowerCL1=put(LowerCL,8.3);
	UpperCL1=put(UpperCL,8.3);
	OR=STRIP(ODDSRatio1)||" ("||STRIP(LowerCL1)||", "||STRIP(UpperCL1)||')';
	MODEL=2;
run;

proc print data=log_covidinfagealladjusted3g1;
	where SCAN(upcase(EFFECT),1)='FIRSTDOSE';
run;

/*** output excel file for results ***/
libname results xlsx "&path\SAS output\&textadd\COVID infections\COVID infections.xlsx";

/* Export first dataset */
data results.sheet1;
```

```
   set work.log_COVIDinfALL;
            where SCAN(upcase(EFFECT),1)='FIRSTDOSE';
run;

/* Export third dataset */
data results.sheet3;
   set work.log_covidinfagealladjusted3g1;
            where SCAN(upcase(EFFECT),1)='FIRSTDOSE';
run;

libname results clear;
```

### Matched cohort Risk of death Dose 1 All-cause deaths

#### The LOGISTIC Procedure

| Model Information | |
|---|---|
| Data Set | WORK.MATCHEDCOHORT |
| Response Variable | death1yrd1 |
| Number of Response Levels | 2 |
| Model | binary logit |
| Optimization Technique | Fisher's scoring |

| | |
|---|---|
| Number of Observations Read | 1962822 |
| Number of Observations Used | 1962822 |

| Response Profile | | |
|---|---|---|
| Ordered Value | death1yrd1 | Total Frequency |
| 1 | 0 | 1949613 |
| 2 | 1 | 13209 |

**Probability modeled is death1yrd1=1.**

| Class Level Information | | |
|---|---|---|
| Class | Value | Design Variables |
| firstdose | 1. PFIZER | 1 |
| | 2. MODERN | 0 |

| Model Convergence Status |
|---|
| Convergence criterion (GCONV=1E-8) satisfied. |

| Model Fit Statistics | | |
|---|---|---|
| Criterion | Intercept Only | Intercept and Covariates |
| AIC | 158453.67 | 158094.81 |
| SC | 158466.16 | 158119.79 |
| -2 Log L | 158451.67 | 158090.81 |

| Testing Global Null Hypothesis: BETA=0 | | | |
|---|---|---|---|
| Test | Chi-Square | DF | Pr > ChiSq |
| Likelihood Ratio | 360.8619 | 1 | <.0001 |
| Score | 359.2380 | 1 | <.0001 |
| Wald | 355.9300 | 1 | <.0001 |

### Matched cohort Risk of death Dose 1 All-cause deaths

#### The LOGISTIC Procedure

| Type 3 Analysis of Effects | | | |
|---|---|---|---|
| Effect | DF | Wald Chi-Square | Pr > ChiSq |
| firstdose | 1 | 355.9300 | <.0001 |

| Analysis of Maximum Likelihood Estimates | | | | | | | |
|---|---|---|---|---|---|---|---|
| Parameter | | DF | Estimate | Standard Error | Wald Chi-Square | Pr > ChiSq | Label |
| Intercept | | 1 | -5.1750 | 0.0135 | 146989.971 | <.0001 | Intercept: death1yrd1=1 |
| firstdose | 1. PFIZER | 1 | 0.3339 | 0.0177 | 355.9300 | <.0001 | firstdose 1. PFIZER |

| Odds Ratio Estimates | | | |
|---|---|---|---|
| Effect | Point Estimate | 95% Wald Confidence Limits | |
| firstdose 1. PFIZER vs 2. MODERN | 1.396 | 1.349 | 1.446 |

| Association of Predicted Probabilities and Observed Responses | | | |
|---|---|---|---|
| Percent Concordant | 29.1 | Somers' D | 0.083 |
| Percent Discordant | 20.9 | Gamma | 0.165 |
| Percent Tied | 50.0 | Tau-a | 0.001 |
| Pairs | 25752438117 | c | 0.541 |

### All-cause death adjusted (logistic regression)

#### The LOGISTIC Procedure

| Model Information | |
|---|---|
| Data Set | WORK.MATCHEDCOHORT |
| Response Variable | death1yrd1 |
| Number of Response Levels | 2 |
| Model | binary logit |
| Optimization Technique | Fisher's scoring |

| | |
|---|---|
| Number of Observations Read | 1962822 |
| Number of Observations Used | 1962822 |

| Response Profile | | |
|---|---|---|
| Ordered Value | death1yrd1 | Total Frequency |
| 1 | 0 | 1949613 |
| 2 | 1 | 13209 |

**Probability modeled is death1yrd1=1.**

| Class Level Information | | | | | | | | | |
|---|---|---|---|---|---|---|---|---|---|
| Class | Value | Design Variables | | | | | | | |
| agenew | 18-39 | 0 | 0 | 0 | 0 | 0 | | | |
| | 40-49 | 1 | 0 | 0 | 0 | 0 | | | |
| | 50-59 | 0 | 1 | 0 | 0 | 0 | | | |
| | 60-69 | 0 | 0 | 1 | 0 | 0 | | | |
| | 70-79 | 0 | 0 | 0 | 1 | 0 | | | |
| | 80+ | 0 | 0 | 0 | 0 | 1 | | | |
| firstdose | 1. PFIZER | 1 | | | | | | | |
| | 2. MODERN | 0 | | | | | | | |
| sex2 | F | 1 | | | | | | | |
| | M | 0 | | | | | | | |
| RACE4 | BLACK | 1 | 0 | 0 | | | | | |
| | OTHERS | 0 | 1 | 0 | | | | | |
| | UNKNOWN | 0 | 0 | 1 | | | | | |
| | WHITE | 0 | 0 | 0 | | | | | |
| ETHNICITY3 | HISPANIC | 1 | 0 | | | | | | |
| | NOT HISPANIC | 0 | 0 | | | | | | |
| | UNKNOWN | 0 | 1 | | | | | | |
| Vaccinationsite | CHD OR DOH | 1 | 0 | 0 | 0 | 0 | 0 | 0 | 0 |
| | DOCTOR CLINIC | 0 | 1 | 0 | 0 | 0 | 0 | 0 | 0 |

#### The LOGISTIC Procedure

| Class Level Information | | | | | | | | | |
|---|---|---|---|---|---|---|---|---|---|
| Class | Value | Design Variables | | | | | | | |
| | FIRE RESQUE OR EMS | 0 | 0 | 1 | 0 | 0 | 0 | 0 | 0 |
| | FQHC OR RHC OR CHC | 0 | 0 | 0 | 1 | 0 | 0 | 0 | 0 |
| | HOSPITAL CLINIC | 0 | 0 | 0 | 0 | 1 | 0 | 0 | 0 |
| | HOSPITAL ER | 0 | 0 | 0 | 0 | 0 | 1 | 0 | 0 |
| | OTHER PHARMACIES | 0 | 0 | 0 | 0 | 0 | 0 | 1 | 0 |
| | OTHERS OR MISC | 0 | 0 | 0 | 0 | 0 | 0 | 0 | 1 |
| | PUBLIX OR CVS OR WALGREENS | 0 | 0 | 0 | 0 | 0 | 0 | 0 | 0 |

| Model Convergence Status |
|---|
| Convergence criterion (GCONV=1E-8) satisfied. |

| Model Fit Statistics | | |
|---|---|---|
| Criterion | Intercept Only | Intercept and Covariates |
| AIC | 158453.67 | 135246.14 |
| SC | 158466.16 | 135545.90 |
| -2 Log L | 158451.67 | 135198.14 |

| Testing Global Null Hypothesis: BETA=0 | | | |
|---|---|---|---|
| Test | Chi-Square | DF | Pr > ChiSq |
| Likelihood Ratio | 23253.5309 | 23 | <.0001 |
| Score | 35187.9700 | 23 | <.0001 |
| Wald | 19691.8371 | 23 | <.0001 |

| Type 3 Analysis of Effects | | | |
|---|---|---|---|
| Effect | DF | Wald Chi-Square | Pr > ChiSq |
| firstdose | 1 | 363.4066 | <.0001 |
| agenew | 5 | 6306.7829 | <.0001 |
| sex2 | 1 | 358.1795 | <.0001 |
| RACE4 | 3 | 197.8344 | <.0001 |
| ETHNICITY3 | 2 | 1561.5107 | <.0001 |
| decile_sviscore1 | 1 | 198.0893 | <.0001 |
| decile_sviscore4 | 1 | 5.8927 | 0.0152 |
| mortalityratepct | 1 | 674.1070 | <.0001 |
| Vaccinationsite | 8 | 1627.6409 | <.0001 |

#### The LOGISTIC Procedure

| Analysis of Maximum Likelihood Estimates | | | | | | | |
|---|---|---|---|---|---|---|---|
| Parameter | | DF | Estimate | Standard Error | Wald Chi-Square | Pr > ChiSq | Label |
| **Intercept** | | 1 | -7.7796 | 0.0638 | 14888.5538 | <.0001 | Intercept: death1yrd1=1 |
| **firstdose** | **1. PFIZER** | 1 | 0.3414 | 0.0179 | 363.4066 | <.0001 | firstdose 1. PFIZER |
| **agenew** | **40-49** | 1 | 0.9265 | 0.0786 | 138.7901 | <.0001 | agenew 40-49 |
| **agenew** | **50-59** | 1 | 1.6286 | 0.0662 | 604.6049 | <.0001 | agenew 50-59 |
| **agenew** | **60-69** | 1 | 2.4781 | 0.0625 | 1573.0270 | <.0001 | agenew 60-69 |
| **agenew** | **70-79** | 1 | 3.2906 | 0.0624 | 2778.1007 | <.0001 | agenew 70-79 |
| **agenew** | **80+** | 1 | 3.9598 | 0.0676 | 3434.3688 | <.0001 | agenew 80+ |
| **sex2** | **F** | 1 | -0.3427 | 0.0181 | 358.1795 | <.0001 | sex2 F |
| **RACE4** | **BLACK** | 1 | 0.0580 | 0.0417 | 1.9356 | 0.1641 | RACE4 BLACK |
| **RACE4** | **OTHERS** | 1 | -0.4688 | 0.0338 | 192.8868 | <.0001 | RACE4 OTHERS |
| **RACE4** | **UNKNOWN** | 1 | -0.0393 | 0.0509 | 0.5955 | 0.4403 | RACE4 UNKNOWN |
| **ETHNICITY3** | **HISPANIC** | 1 | -0.5045 | 0.0337 | 224.6398 | <.0001 | ETHNICITY3 HISPANIC |
| **ETHNICITY3** | **UNKNOWN** | 1 | 0.9557 | 0.0281 | 1156.6424 | <.0001 | ETHNICITY3 UNKNOWN |
| **decile_sviscore1** | | 1 | 0.0552 | 0.00392 | 198.0893 | <.0001 | Rank for Variable SVISCORE1 |
| **decile_sviscore4** | | 1 | 0.00895 | 0.00369 | 5.8927 | 0.0152 | Rank for Variable SVISCORE4 |
| **mortalityratepct** | | 1 | 0.0786 | 0.00303 | 674.1070 | <.0001 | |
| **Vaccinationsite** | **CHD OR DOH** | 1 | -0.3398 | 0.0262 | 167.8760 | <.0001 | Vaccinationsite CHD OR DOH |
| **Vaccinationsite** | **DOCTOR CLINIC** | 1 | 0.1414 | 0.0240 | 34.7449 | <.0001 | Vaccinationsite DOCTOR CLINIC |
| **Vaccinationsite** | **FIRE RESQUE OR EMS** | 1 | -0.3208 | 0.0520 | 38.0547 | <.0001 | Vaccinationsite FIRE RESQUE OR EMS |
| **Vaccinationsite** | **FQHC OR RHC OR CHC** | 1 | -0.3684 | 0.0832 | 19.6124 | <.0001 | Vaccinationsite FQHC OR RHC OR CHC |
| **Vaccinationsite** | **HOSPITAL CLINIC** | 1 | 0.0161 | 0.0368 | 0.1911 | 0.6620 | Vaccinationsite HOSPITAL CLINIC |
| **Vaccinationsite** | **HOSPITAL ER** | 1 | -0.1277 | 0.1690 | 0.5704 | 0.4501 | Vaccinationsite HOSPITAL ER |
| **Vaccinationsite** | **OTHER PHARMACIES** | 1 | 0.3700 | 0.0515 | 51.5622 | <.0001 | Vaccinationsite OTHER PHARMACIES |
| **Vaccinationsite** | **OTHERS OR MISC** | 1 | -1.5381 | 0.0445 | 1196.7365 | <.0001 | Vaccinationsite OTHERS OR MISC |

| Odds Ratio Estimates | | | |
|---|---|---|---|
| Effect | Point Estimate | 95% Wald Confidence Limits | |
| **firstdose 1. PFIZER vs 2. MODERN** | 1.407 | 1.358 | 1.457 |
| **agenew 40-49 vs 18-39** | 2.526 | 2.165 | 2.947 |
| **agenew 50-59 vs 18-39** | 5.097 | 4.476 | 5.803 |
| **agenew 60-69 vs 18-39** | 11.919 | 10.545 | 13.472 |
| **agenew 70-79 vs 18-39** | 26.860 | 23.766 | 30.356 |
| **agenew 80+ vs 18-39** | 52.448 | 45.942 | 59.875 |
| **sex2 F vs M** | 0.710 | 0.685 | 0.736 |

#### The LOGISTIC Procedure

| Odds Ratio Estimates | | | |
|---|---|---|---|
| Effect | Point Estimate | 95% Wald Confidence Limits | |
| RACE4 BLACK vs WHITE | 1.060 | 0.977 | 1.150 |
| RACE4 OTHERS vs WHITE | 0.626 | 0.586 | 0.669 |
| RACE4 UNKNOWN vs WHITE | 0.961 | 0.870 | 1.062 |
| ETHNICITY3 HISPANIC vs NOT HISPANIC | 0.604 | 0.565 | 0.645 |
| ETHNICITY3 UNKNOWN vs NOT HISPANIC | 2.601 | 2.461 | 2.748 |
| decile_sviscore1 | 1.057 | 1.049 | 1.065 |
| decile_sviscore4 | 1.009 | 1.002 | 1.016 |
| mortalityratepct | 1.082 | 1.075 | 1.088 |
| Vaccinationsite CHD OR DOH vs PUBLIX OR CVS OR WALGREENS | 0.712 | 0.676 | 0.749 |
| Vaccinationsite DOCTOR CLINIC vs PUBLIX OR CVS OR WALGREENS | 1.152 | 1.099 | 1.207 |
| Vaccinationsite FIRE RESQUE OR EMS vs PUBLIX OR CVS OR WALGREENS | 0.726 | 0.655 | 0.803 |
| Vaccinationsite FQHC OR RHC OR CHC vs PUBLIX OR CVS OR WALGREENS | 0.692 | 0.588 | 0.814 |
| Vaccinationsite HOSPITAL CLINIC vs PUBLIX OR CVS OR WALGREENS | 1.016 | 0.945 | 1.092 |
| Vaccinationsite HOSPITAL ER vs PUBLIX OR CVS OR WALGREENS | 0.880 | 0.632 | 1.226 |
| Vaccinationsite OTHER PHARMACIES vs PUBLIX OR CVS OR WALGREENS | 1.448 | 1.309 | 1.602 |
| Vaccinationsite OTHERS OR MISC vs PUBLIX OR CVS OR WALGREENS | 0.215 | 0.197 | 0.234 |

| Association of Predicted Probabilities and Observed Responses | | | |
|---|---|---|---|
| Percent Concordant | 83.4 | Somers' D | 0.667 |
| Percent Discordant | 16.6 | Gamma | 0.667 |
| Percent Tied | 0.0 | Tau-a | 0.009 |
| Pairs | 25752438117 | c | 0.834 |

### All-cause deaths adjusted by Age

#### The LOGISTIC Procedure

##### agenew=18-39 years

| Model Information | |
|---|---|
| Data Set | WORK.MATCHEDCOHORT |
| Response Variable | death1yrd1 |
| Number of Response Levels | 2 |
| Model | binary logit |
| Optimization Technique | Fisher's scoring |

| | |
|---|---|
| Number of Observations Read | 492484 |
| Number of Observations Used | 492484 |

| Response Profile | | |
|---|---|---|
| Ordered Value | death1yrd1 | Total Frequency |
| 1 | 0 | 492195 |
| 2 | 1 | 289 |

**Probability modeled is death1yrd1=1.**

| Class Level Information | | | | | | | | | |
|---|---|---|---|---|---|---|---|---|---|
| Class | Value | Design Variables | | | | | | | |
| agenew | 18-39 years | | | | | | | | |
| firstdose | 1. PFIZER | 1 | | | | | | | |
| | 2. MODERN | 0 | | | | | | | |
| sex2 | F | 1 | | | | | | | |
| | M | 0 | | | | | | | |
| RACE4 | BLACK | 1 | 0 | 0 | | | | | |
| | OTHERS | 0 | 1 | 0 | | | | | |
| | UNKNOWN | 0 | 0 | 1 | | | | | |
| | WHITE | 0 | 0 | 0 | | | | | |
| ETHNICITY3 | HISPANIC | 1 | 0 | | | | | | |
| | NOT HISPANIC | 0 | 0 | | | | | | |
| | UNKNOWN | 0 | 1 | | | | | | |
| Vaccinationsite | CHD OR DOH | 1 | 0 | 0 | 0 | 0 | 0 | 0 | 0 |
| | DOCTOR CLINIC | 0 | 1 | 0 | 0 | 0 | 0 | 0 | 0 |
| | FIRE RESQUE OR EMS | 0 | 0 | 1 | 0 | 0 | 0 | 0 | 0 |
| | FQHC OR RHC OR CHC | 0 | 0 | 0 | 1 | 0 | 0 | 0 | 0 |
| | HOSPITAL CLINIC | 0 | 0 | 0 | 0 | 1 | 0 | 0 | 0 |
| | HOSPITAL ER | 0 | 0 | 0 | 0 | 0 | 1 | 0 | 0 |

### All-cause deaths adjusted by Age

#### The LOGISTIC Procedure

**agenew=18-39 years**

| Class Level Information | | | | | | | | | |
|---|---|---|---|---|---|---|---|---|---|
| Class | Value | Design Variables | | | | | | | |
| | OTHER PHARMACIES | 0 | 0 | 0 | 0 | 0 | 0 | 1 | 0 |
| | OTHERS OR MISC | 0 | 0 | 0 | 0 | 0 | 0 | 0 | 1 |
| | PUBLIX OR CVS OR WALGREENS | 0 | 0 | 0 | 0 | 0 | 0 | 0 | 0 |

| Model Convergence Status |
|---|
| Convergence criterion (GCONV=1E-8) satisfied. |

| Model Fit Statistics | | |
|---|---|---|
| Criterion | Intercept Only | Intercept and Covariates |
| AIC | 4880.607 | 4761.795 |
| SC | 4891.715 | 4972.832 |
| -2 Log L | 4878.607 | 4723.795 |

| Testing Global Null Hypothesis: BETA=0 | | | |
|---|---|---|---|
| Test | Chi-Square | DF | Pr > ChiSq |
| Likelihood Ratio | 154.8122 | 18 | <.0001 |
| Score | 154.5957 | 18 | <.0001 |
| Wald | 145.4467 | 18 | <.0001 |

| Type 3 Analysis of Effects | | | |
|---|---|---|---|
| Effect | DF | Wald Chi-Square | Pr > ChiSq |
| firstdose | 1 | 6.9472 | 0.0084 |
| agenew | 0 | . | . |
| sex2 | 1 | 43.4752 | <.0001 |
| RACE4 | 3 | 5.7979 | 0.1219 |
| ETHNICITY3 | 2 | 46.2124 | <.0001 |
| decile_sviscore1 | 1 | 21.4342 | <.0001 |
| decile_sviscore4 | 1 | 0.4039 | 0.5251 |
| mortalityratepct | 1 | 0.0009 | 0.9761 |
| Vaccinationsite | 8 | 18.3553 | 0.0187 |

#### The LOGISTIC Procedure

**agenew=18-39 years**

| Analysis of Maximum Likelihood Estimates | | | | | | | |
|---|---|---|---|---|---|---|---|
| **Parameter** | | **DF** | **Estimate** | **Standard Error** | **Wald Chi-Square** | **Pr > ChiSq** | **Label** |
| **Intercept** | | 1 | -7.6616 | 0.1650 | 2155.1249 | <.0001 | Intercept: death1yrd1=1 |
| **firstdose** | **1. PFIZER** | 1 | 0.3141 | 0.1192 | 6.9472 | 0.0084 | firstdose 1. PFIZER |
| **sex2** | **F** | 1 | -0.8174 | 0.1240 | 43.4752 | <.0001 | sex2 F |
| **RACE4** | **BLACK** | 1 | 0.1468 | 0.1928 | 0.5797 | 0.4464 | RACE4 BLACK |
| **RACE4** | **OTHERS** | 1 | -0.1100 | 0.1684 | 0.4267 | 0.5136 | RACE4 OTHERS |
| **RACE4** | **UNKNOWN** | 1 | -0.7413 | 0.3426 | 4.6821 | 0.0305 | RACE4 UNKNOWN |
| **ETHNICITY3** | **HISPANIC** | 1 | -0.7700 | 0.1757 | 19.1998 | <.0001 | ETHNICITY3 HISPANIC |
| **ETHNICITY3** | **UNKNOWN** | 1 | 0.5684 | 0.1506 | 14.2489 | 0.0002 | ETHNICITY3 UNKNOWN |
| **decile_sviscore1** | | 1 | 0.1213 | 0.0262 | 21.4342 | <.0001 | Rank for Variable SVISCORE1 |
| **decile_sviscore4** | | 1 | -0.0157 | 0.0247 | 0.4039 | 0.5251 | Rank for Variable SVISCORE4 |
| **mortalityratepct** | | 1 | -10.8783 | 363.7 | 0.0009 | 0.9761 | |
| **Vaccinationsite** | **CHD OR DOH** | 1 | -0.3996 | 0.3621 | 1.2175 | 0.2698 | Vaccinationsite CHD OR DOH |
| **Vaccinationsite** | **DOCTOR CLINIC** | 1 | 0.2924 | 0.1675 | 3.0460 | 0.0809 | Vaccinationsite DOCTOR CLINIC |
| **Vaccinationsite** | **FIRE RESQUE OR EMS** | 1 | -10.6951 | 419.2 | 0.0007 | 0.9796 | Vaccinationsite FIRE RESQUE OR EMS |
| **Vaccinationsite** | **FQHC OR RHC OR CHC** | 1 | 0.0764 | 0.5863 | 0.0170 | 0.8964 | Vaccinationsite FQHC OR RHC OR CHC |
| **Vaccinationsite** | **HOSPITAL CLINIC** | 1 | -0.4960 | 0.4544 | 1.1913 | 0.2751 | Vaccinationsite HOSPITAL CLINIC |
| **Vaccinationsite** | **HOSPITAL ER** | 1 | 0.5741 | 1.0041 | 0.3269 | 0.5675 | Vaccinationsite HOSPITAL ER |
| **Vaccinationsite** | **OTHER PHARMACIES** | 1 | 0.3778 | 0.2242 | 2.8390 | 0.0920 | Vaccinationsite OTHER PHARMACIES |
| **Vaccinationsite** | **OTHERS OR MISC** | 1 | -1.5064 | 0.5064 | 8.8505 | 0.0029 | Vaccinationsite OTHERS OR MISC |

| Odds Ratio Estimates | | | |
|---|---|---|---|
| **Effect** | **Point Estimate** | **95% Wald Confidence Limits** | |
| **firstdose 1. PFIZER vs 2. MODERN** | 1.369 | 1.084 | 1.729 |
| **sex2 F vs M** | 0.442 | 0.346 | 0.563 |
| **RACE4 BLACK vs WHITE** | 1.158 | 0.794 | 1.690 |
| **RACE4 OTHERS vs WHITE** | 0.896 | 0.644 | 1.246 |
| **RACE4 UNKNOWN vs WHITE** | 0.477 | 0.243 | 0.933 |
| **ETHNICITY3 HISPANIC vs NOT HISPANIC** | 0.463 | 0.328 | 0.653 |
| **ETHNICITY3 UNKNOWN vs NOT HISPANIC** | 1.765 | 1.314 | 2.372 |
| **decile_sviscore1** | 1.129 | 1.072 | 1.189 |
| **decile_sviscore4** | 0.984 | 0.938 | 1.033 |
| **mortalityratepct** | <0.001 | <0.001 | >999.999 |
| **Vaccinationsite CHD OR DOH vs PUBLIX OR CVS OR WALGREENS** | 0.671 | 0.330 | 1.364 |

#### The LOGISTIC Procedure

**agenew=18-39 years**

| Odds Ratio Estimates | | | |
|---|---|---|---|
| Effect | Point Estimate | 95% Wald Confidence Limits | |
| Vaccinationsite DOCTOR CLINIC vs PUBLIX OR CVS OR WALGREENS | 1.340 | 0.965 | 1.860 |
| Vaccinationsite FIRE RESQUE OR EMS vs PUBLIX OR CVS OR WALGREENS | <0.001 | <0.001 | >999.999 |
| Vaccinationsite FQHC OR RHC OR CHC vs PUBLIX OR CVS OR WALGREENS | 1.079 | 0.342 | 3.406 |
| Vaccinationsite HOSPITAL CLINIC vs PUBLIX OR CVS OR WALGREENS | 0.609 | 0.250 | 1.484 |
| Vaccinationsite HOSPITAL ER vs PUBLIX OR CVS OR WALGREENS | 1.776 | 0.248 | 12.707 |
| Vaccinationsite OTHER PHARMACIES vs PUBLIX OR CVS OR WALGREENS | 1.459 | 0.940 | 2.264 |
| Vaccinationsite OTHERS OR MISC vs PUBLIX OR CVS OR WALGREENS | 0.222 | 0.082 | 0.598 |

| Association of Predicted Probabilities and Observed Responses | | | |
|---|---|---|---|
| Percent Concordant | 70.8 | Somers' D | 0.417 |
| Percent Discordant | 29.1 | Gamma | 0.417 |
| Percent Tied | 0.0 | Tau-a | 0.000 |
| Pairs | 142244355 | c | 0.709 |

#### The LOGISTIC Procedure

**agenew=40-59 years**

| Model Information | |
|---|---|
| **Data Set** | WORK.MATCHEDCOHORT |
| **Response Variable** | death1yrd1 |
| **Number of Response Levels** | 2 |
| **Model** | binary logit |
| **Optimization Technique** | Fisher's scoring |

| | |
|---|---|
| **Number of Observations Read** | 634102 |
| **Number of Observations Used** | 634102 |

| Response Profile | | |
|---|---|---|
| **Ordered Value** | **death1yrd1** | **Total Frequency** |
| **1** | 0 | 632619 |
| **2** | 1 | 1483 |

**Probability modeled is death1yrd1=1.**

| Class Level Information | | | | | | | | | |
|---|---|---|---|---|---|---|---|---|---|
| **Class** | **Value** | **Design Variables** | | | | | | | |
| **agenew** | **40-59 years** | | | | | | | | |
| **firstdose** | **1. PFIZER** | 1 | | | | | | | |
| | **2. MODERN** | 0 | | | | | | | |
| **sex2** | **F** | 1 | | | | | | | |
| | **M** | 0 | | | | | | | |
| **RACE4** | **BLACK** | 1 | 0 | 0 | | | | | |
| | **OTHERS** | 0 | 1 | 0 | | | | | |
| | **UNKNOWN** | 0 | 0 | 1 | | | | | |
| | **WHITE** | 0 | 0 | 0 | | | | | |
| **ETHNICITY3** | **HISPANIC** | 1 | 0 | | | | | | |
| | **NOT HISPANIC** | 0 | 0 | | | | | | |
| | **UNKNOWN** | 0 | 1 | | | | | | |
| **Vaccinationsite** | **CHD OR DOH** | 1 | 0 | 0 | 0 | 0 | 0 | 0 | 0 |
| | **DOCTOR CLINIC** | 0 | 1 | 0 | 0 | 0 | 0 | 0 | 0 |
| | **FIRE RESQUE OR EMS** | 0 | 0 | 1 | 0 | 0 | 0 | 0 | 0 |
| | **FQHC OR RHC OR CHC** | 0 | 0 | 0 | 1 | 0 | 0 | 0 | 0 |
| | **HOSPITAL CLINIC** | 0 | 0 | 0 | 0 | 1 | 0 | 0 | 0 |
| | **HOSPITAL ER** | 0 | 0 | 0 | 0 | 0 | 1 | 0 | 0 |

### All-cause deaths adjusted by Age

#### The LOGISTIC Procedure

**agenew=40-59 years**

| Class Level Information | | | | | | | | | |
|---|---|---|---|---|---|---|---|---|---|
| Class | Value | Design Variables | | | | | | | |
| | OTHER PHARMACIES | 0 | 0 | 0 | 0 | 0 | 0 | 1 | 0 |
| | OTHERS OR MISC | 0 | 0 | 0 | 0 | 0 | 0 | 0 | 1 |
| | PUBLIX OR CVS OR WALGREENS | 0 | 0 | 0 | 0 | 0 | 0 | 0 | 0 |

| Model Convergence Status |
|---|
| Convergence criterion (GCONV=1E-8) satisfied. |

| Model Fit Statistics | | |
|---|---|---|
| Criterion | Intercept Only | Intercept and Covariates |
| AIC | 20932.980 | 20140.620 |
| SC | 20944.340 | 20356.459 |
| -2 Log L | 20930.980 | 20102.620 |

| Testing Global Null Hypothesis: BETA=0 | | | |
|---|---|---|---|
| Test | Chi-Square | DF | Pr > ChiSq |
| Likelihood Ratio | 828.3604 | 18 | <.0001 |
| Score | 846.7057 | 18 | <.0001 |
| Wald | 799.1272 | 18 | <.0001 |

| Type 3 Analysis of Effects | | | |
|---|---|---|---|
| Effect | DF | Wald Chi-Square | Pr > ChiSq |
| firstdose | 1 | 7.8474 | 0.0051 |
| agenew | 0 | . | . |
| sex2 | 1 | 28.7600 | <.0001 |
| RACE4 | 3 | 16.7869 | 0.0008 |
| ETHNICITY3 | 2 | 160.7530 | <.0001 |
| decile_sviscore1 | 1 | 78.5136 | <.0001 |
| decile_sviscore4 | 1 | 1.6553 | 0.1982 |
| mortalityratepct | 1 | 170.5752 | <.0001 |
| Vaccinationsite | 8 | 142.7416 | <.0001 |

#### The LOGISTIC Procedure

**agenew=40-59 years**

| Analysis of Maximum Likelihood Estimates | | | | | | | |
|---|---|---|---|---|---|---|---|
| **Parameter** | | **DF** | **Estimate** | **Standard Error** | **Wald Chi-Square** | **Pr > ChiSq** | **Label** |
| **Intercept** | | 1 | -6.8886 | 0.0829 | 6904.0623 | <.0001 | Intercept: death1yrd1=1 |
| **firstdose** | **1. PFIZER** | 1 | 0.1462 | 0.0522 | 7.8474 | 0.0051 | firstdose 1. PFIZER |
| **sex2** | **F** | 1 | -0.3005 | 0.0560 | 28.7600 | <.0001 | sex2 F |
| **RACE4** | **BLACK** | 1 | 0.0876 | 0.0859 | 1.0402 | 0.3078 | RACE4 BLACK |
| **RACE4** | **OTHERS** | 1 | -0.3856 | 0.1036 | 13.8439 | 0.0002 | RACE4 OTHERS |
| **RACE4** | **UNKNOWN** | 1 | -0.2054 | 0.1390 | 2.1815 | 0.1397 | RACE4 UNKNOWN |
| **ETHNICITY3** | **HISPANIC** | 1 | -0.6826 | 0.0881 | 60.0248 | <.0001 | ETHNICITY3 HISPANIC |
| **ETHNICITY3** | **UNKNOWN** | 1 | 0.6340 | 0.0792 | 64.0514 | <.0001 | ETHNICITY3 UNKNOWN |
| **decile_sviscore1** | | 1 | 0.1045 | 0.0118 | 78.5136 | <.0001 | Rank for Variable SVISCORE1 |
| **decile_sviscore4** | | 1 | 0.0143 | 0.0111 | 1.6553 | 0.1982 | Rank for Variable SVISCORE4 |
| **mortalityratepct** | | 1 | 0.7220 | 0.0553 | 170.5752 | <.0001 | |
| **Vaccinationsite** | **CHD OR DOH** | 1 | -0.0785 | 0.1495 | 0.2759 | 0.5994 | Vaccinationsite CHD OR DOH |
| **Vaccinationsite** | **DOCTOR CLINIC** | 1 | 0.2384 | 0.0690 | 11.9270 | 0.0006 | Vaccinationsite DOCTOR CLINIC |
| **Vaccinationsite** | **FIRE RESQUE OR EMS** | 1 | -0.4482 | 0.7089 | 0.3998 | 0.5272 | Vaccinationsite FIRE RESQUE OR EMS |
| **Vaccinationsite** | **FQHC OR RHC OR CHC** | 1 | 0.2844 | 0.1949 | 2.1278 | 0.1446 | Vaccinationsite FQHC OR RHC OR CHC |
| **Vaccinationsite** | **HOSPITAL CLINIC** | 1 | 0.7070 | 0.0972 | 52.9553 | <.0001 | Vaccinationsite HOSPITAL CLINIC |
| **Vaccinationsite** | **HOSPITAL ER** | 1 | 0.0339 | 0.5021 | 0.0046 | 0.9462 | Vaccinationsite HOSPITAL ER |
| **Vaccinationsite** | **OTHER PHARMACIES** | 1 | 0.4626 | 0.1000 | 21.4191 | <.0001 | Vaccinationsite OTHER PHARMACIES |
| **Vaccinationsite** | **OTHERS OR MISC** | 1 | -1.1394 | 0.1564 | 53.0816 | <.0001 | Vaccinationsite OTHERS OR MISC |

| Odds Ratio Estimates | | | |
|---|---|---|---|
| **Effect** | **Point Estimate** | **95% Wald Confidence Limits** | |
| **firstdose 1. PFIZER vs 2. MODERN** | 1.157 | 1.045 | 1.282 |
| **sex2 F vs M** | 0.740 | 0.663 | 0.826 |
| **RACE4 BLACK vs WHITE** | 1.092 | 0.922 | 1.292 |
| **RACE4 OTHERS vs WHITE** | 0.680 | 0.555 | 0.833 |
| **RACE4 UNKNOWN vs WHITE** | 0.814 | 0.620 | 1.069 |
| **ETHNICITY3 HISPANIC vs NOT HISPANIC** | 0.505 | 0.425 | 0.601 |
| **ETHNICITY3 UNKNOWN vs NOT HISPANIC** | 1.885 | 1.614 | 2.202 |
| **decile_sviscore1** | 1.110 | 1.085 | 1.136 |
| **decile_sviscore4** | 1.014 | 0.993 | 1.037 |
| **mortalityratepct** | 2.058 | 1.847 | 2.294 |
| **Vaccinationsite CHD OR DOH vs PUBLIX OR CVS OR WALGREENS** | 0.924 | 0.690 | 1.239 |

#### The LOGISTIC Procedure

**agenew=40-59 years**

| Odds Ratio Estimates | | | |
|---|---|---|---|
| Effect | Point Estimate | 95% Wald Confidence Limits | |
| Vaccinationsite DOCTOR CLINIC vs PUBLIX OR CVS OR WALGREENS | 1.269 | 1.109 | 1.453 |
| Vaccinationsite FIRE RESQUE OR EMS vs PUBLIX OR CVS OR WALGREENS | 0.639 | 0.159 | 2.563 |
| Vaccinationsite FQHC OR RHC OR CHC vs PUBLIX OR CVS OR WALGREENS | 1.329 | 0.907 | 1.947 |
| Vaccinationsite HOSPITAL CLINIC vs PUBLIX OR CVS OR WALGREENS | 2.028 | 1.676 | 2.453 |
| Vaccinationsite HOSPITAL ER vs PUBLIX OR CVS OR WALGREENS | 1.034 | 0.387 | 2.768 |
| Vaccinationsite OTHER PHARMACIES vs PUBLIX OR CVS OR WALGREENS | 1.588 | 1.306 | 1.932 |
| Vaccinationsite OTHERS OR MISC vs PUBLIX OR CVS OR WALGREENS | 0.320 | 0.236 | 0.435 |

| Association of Predicted Probabilities and Observed Responses | | | |
|---|---|---|---|
| Percent Concordant | 70.9 | Somers' D | 0.417 |
| Percent Discordant | 29.1 | Gamma | 0.417 |
| Percent Tied | 0.0 | Tau-a | 0.002 |
| Pairs | 938173977 | c | 0.709 |

### All-cause deaths adjusted by Age

#### The LOGISTIC Procedure

**agenew=60+ years**

| Model Information | |
|---|---|
| **Data Set** | WORK.MATCHEDCOHORT |
| **Response Variable** | death1yrd1 |
| **Number of Response Levels** | 2 |
| **Model** | binary logit |
| **Optimization Technique** | Fisher's scoring |

| | |
|---|---|
| **Number of Observations Read** | 836236 |
| **Number of Observations Used** | 836236 |

| Response Profile | | |
|---|---|---|
| **Ordered Value** | **death1yrd1** | **Total Frequency** |
| **1** | 0 | 824799 |
| **2** | 1 | 11437 |

**Probability modeled is death1yrd1=1.**

| Class Level Information | | | | | | | | | |
|---|---|---|---|---|---|---|---|---|---|
| **Class** | **Value** | **Design Variables** | | | | | | | |
| **agenew** | **60+ years** | | | | | | | | |
| **firstdose** | **1. PFIZER** | 1 | | | | | | | |
| | **2. MODERN** | 0 | | | | | | | |
| **sex2** | **F** | 1 | | | | | | | |
| | **M** | 0 | | | | | | | |
| **RACE4** | **BLACK** | 1 | 0 | 0 | | | | | |
| | **OTHERS** | 0 | 1 | 0 | | | | | |
| | **UNKNOWN** | 0 | 0 | 1 | | | | | |
| | **WHITE** | 0 | 0 | 0 | | | | | |
| **ETHNICITY3** | **HISPANIC** | 1 | 0 | | | | | | |
| | **NOT HISPANIC** | 0 | 0 | | | | | | |
| | **UNKNOWN** | 0 | 1 | | | | | | |
| **Vaccinationsite** | **CHD OR DOH** | 1 | 0 | 0 | 0 | 0 | 0 | 0 | 0 |
| | **DOCTOR CLINIC** | 0 | 1 | 0 | 0 | 0 | 0 | 0 | 0 |
| | **FIRE RESQUE OR EMS** | 0 | 0 | 1 | 0 | 0 | 0 | 0 | 0 |
| | **FQHC OR RHC OR CHC** | 0 | 0 | 0 | 1 | 0 | 0 | 0 | 0 |
| | **HOSPITAL CLINIC** | 0 | 0 | 0 | 0 | 1 | 0 | 0 | 0 |
| | **HOSPITAL ER** | 0 | 0 | 0 | 0 | 0 | 1 | 0 | 0 |

#### The LOGISTIC Procedure

**agenew=60+ years**

| Class Level Information | | | | | | | | | |
|---|---|---|---|---|---|---|---|---|---|
| Class | Value | Design Variables | | | | | | | |
| | OTHER PHARMACIES | 0 | 0 | 0 | 0 | 0 | 0 | 1 | 0 |
| | OTHERS OR MISC | 0 | 0 | 0 | 0 | 0 | 0 | 0 | 1 |
| | PUBLIX OR CVS OR WALGREENS | 0 | 0 | 0 | 0 | 0 | 0 | 0 | 0 |

| Model Convergence Status |
|---|
| Convergence criterion (GCONV=1E-8) satisfied. |

| Model Fit Statistics | | |
|---|---|---|
| Criterion | Intercept Only | Intercept and Covariates |
| AIC | 120895.38 | 112103.19 |
| SC | 120907.01 | 112324.29 |
| -2 Log L | 120893.38 | 112065.19 |

| Testing Global Null Hypothesis: BETA=0 | | | |
|---|---|---|---|
| Test | Chi-Square | DF | Pr > ChiSq |
| Likelihood Ratio | 8828.1872 | 18 | <.0001 |
| Score | 13432.5994 | 18 | <.0001 |
| Wald | 9428.8131 | 18 | <.0001 |

| Type 3 Analysis of Effects | | | |
|---|---|---|---|
| Effect | DF | Wald Chi-Square | Pr > ChiSq |
| firstdose | 1 | 363.8920 | <.0001 |
| agenew | 0 | . | . |
| sex2 | 1 | 124.8730 | <.0001 |
| RACE4 | 3 | 181.5484 | <.0001 |
| ETHNICITY3 | 2 | 1501.7581 | <.0001 |
| decile_sviscore1 | 1 | 68.0886 | <.0001 |
| decile_sviscore4 | 1 | 8.8109 | 0.0030 |
| mortalityratepct | 1 | 6052.9853 | <.0001 |
| Vaccinationsite | 8 | 1308.8278 | <.0001 |

#### The LOGISTIC Procedure

**agenew=60+ years**

| Analysis of Maximum Likelihood Estimates | | | | | | | |
|---|---|---|---|---|---|---|---|
| **Parameter** | | **DF** | **Estimate** | **Standard Error** | **Wald Chi-Square** | **Pr > ChiSq** | **Label** |
| **Intercept** | | 1 | -4.9819 | 0.0295 | 28483.6525 | <.0001 | Intercept: death1yrd1=1 |
| **firstdose** | **1. PFIZER** | 1 | 0.3695 | 0.0194 | 363.8920 | <.0001 | firstdose 1. PFIZER |
| **sex2** | **F** | 1 | -0.2156 | 0.0193 | 124.8730 | <.0001 | sex2 F |
| **RACE4** | **BLACK** | 1 | -0.1418 | 0.0493 | 8.2931 | 0.0040 | RACE4 BLACK |
| **RACE4** | **OTHERS** | 1 | -0.5079 | 0.0385 | 174.3057 | <.0001 | RACE4 OTHERS |
| **RACE4** | **UNKNOWN** | 1 | 0.0300 | 0.0556 | 0.2899 | 0.5903 | RACE4 UNKNOWN |
| **ETHNICITY3** | **HISPANIC** | 1 | -0.5262 | 0.0387 | 184.5162 | <.0001 | ETHNICITY3 HISPANIC |
| **ETHNICITY3** | **UNKNOWN** | 1 | 1.0529 | 0.0306 | 1184.9031 | <.0001 | ETHNICITY3 UNKNOWN |
| **decile_sviscore1** | | 1 | 0.0349 | 0.00423 | 68.0886 | <.0001 | Rank for Variable SVISCORE1 |
| **decile_sviscore4** | | 1 | 0.0118 | 0.00398 | 8.8109 | 0.0030 | Rank for Variable SVISCORE4 |
| **mortalityratepct** | | 1 | 0.1666 | 0.00214 | 6052.9853 | <.0001 | |
| **Vaccinationsite** | **CHD OR DOH** | 1 | -0.2009 | 0.0267 | 56.6128 | <.0001 | Vaccinationsite CHD OR DOH |
| **Vaccinationsite** | **DOCTOR CLINIC** | 1 | 0.2314 | 0.0258 | 80.2837 | <.0001 | Vaccinationsite DOCTOR CLINIC |
| **Vaccinationsite** | **FIRE RESQUE OR EMS** | 1 | -0.0649 | 0.0525 | 1.5237 | 0.2171 | Vaccinationsite FIRE RESQUE OR EMS |
| **Vaccinationsite** | **FQHC OR RHC OR CHC** | 1 | -0.3583 | 0.0929 | 14.8729 | 0.0001 | Vaccinationsite FQHC OR RHC OR CHC |
| **Vaccinationsite** | **HOSPITAL CLINIC** | 1 | 0.00396 | 0.0403 | 0.0097 | 0.9217 | Vaccinationsite HOSPITAL CLINIC |
| **Vaccinationsite** | **HOSPITAL ER** | 1 | -0.0874 | 0.1820 | 0.2307 | 0.6310 | Vaccinationsite HOSPITAL ER |
| **Vaccinationsite** | **OTHER PHARMACIES** | 1 | 0.2655 | 0.0626 | 18.0047 | <.0001 | Vaccinationsite OTHER PHARMACIES |
| **Vaccinationsite** | **OTHERS OR MISC** | 1 | -1.4491 | 0.0467 | 963.2897 | <.0001 | Vaccinationsite OTHERS OR MISC |

| Odds Ratio Estimates | | | |
|---|---|---|---|
| **Effect** | **Point Estimate** | **95% Wald Confidence Limits** | |
| **firstdose 1. PFIZER vs 2. MODERN** | 1.447 | 1.393 | 1.503 |
| **sex2 F vs M** | 0.806 | 0.776 | 0.837 |
| **RACE4 BLACK vs WHITE** | 0.868 | 0.788 | 0.956 |
| **RACE4 OTHERS vs WHITE** | 0.602 | 0.558 | 0.649 |
| **RACE4 UNKNOWN vs WHITE** | 1.030 | 0.924 | 1.149 |
| **ETHNICITY3 HISPANIC vs NOT HISPANIC** | 0.591 | 0.548 | 0.637 |
| **ETHNICITY3 UNKNOWN vs NOT HISPANIC** | 2.866 | 2.699 | 3.043 |
| **decile_sviscore1** | 1.035 | 1.027 | 1.044 |
| **decile_sviscore4** | 1.012 | 1.004 | 1.020 |
| **mortalityratepct** | 1.181 | 1.176 | 1.186 |
| **Vaccinationsite CHD OR DOH vs PUBLIX OR CVS OR WALGREENS** | 0.818 | 0.776 | 0.862 |

#### The LOGISTIC Procedure

**agenew=60+ years**

| Odds Ratio Estimates | | | |
|---|---|---|---|
| Effect | Point Estimate | 95% Wald Confidence Limits | |
| Vaccinationsite DOCTOR CLINIC vs PUBLIX OR CVS OR WALGREENS | 1.260 | 1.198 | 1.326 |
| Vaccinationsite FIRE RESQUE OR EMS vs PUBLIX OR CVS OR WALGREENS | 0.937 | 0.845 | 1.039 |
| Vaccinationsite FQHC OR RHC OR CHC vs PUBLIX OR CVS OR WALGREENS | 0.699 | 0.582 | 0.838 |
| Vaccinationsite HOSPITAL CLINIC vs PUBLIX OR CVS OR WALGREENS | 1.004 | 0.928 | 1.086 |
| Vaccinationsite HOSPITAL ER vs PUBLIX OR CVS OR WALGREENS | 0.916 | 0.641 | 1.309 |
| Vaccinationsite OTHER PHARMACIES vs PUBLIX OR CVS OR WALGREENS | 1.304 | 1.154 | 1.474 |
| Vaccinationsite OTHERS OR MISC vs PUBLIX OR CVS OR WALGREENS | 0.235 | 0.214 | 0.257 |

| Association of Predicted Probabilities and Observed Responses | | | |
|---|---|---|---|
| Percent Concordant | 73.4 | Somers' D | 0.469 |
| Percent Discordant | 26.6 | Gamma | 0.469 |
| Percent Tied | 0.0 | Tau-a | 0.013 |
| Pairs | 9433226163 | c | 0.734 |

#### The LOGISTIC Procedure

| Model Information | |
|---|---|
| **Data Set** | WORK.MATCHEDCOHORT |
| **Response Variable** | CVD1YRD1 |
| **Number of Response Levels** | 2 |
| **Model** | binary logit |
| **Optimization Technique** | Fisher's scoring |

| | |
|---|---|
| **Number of Observations Read** | 1962822 |
| **Number of Observations Used** | 1962822 |

| Response Profile | | |
|---|---|---|
| **Ordered Value** | **CVD1YRD1** | **Total Frequency** |
| **1** | 0 | 1959183 |
| **2** | 1 | 3639 |

**Probability modeled is CVD1YRD1=1.**

| Class Level Information | | |
|---|---|---|
| **Class** | **Value** | **Design Variables** |
| **firstdose** | **1. PFIZER** | 1 |
| | **2. MODERN** | 0 |

| Model Convergence Status |
|---|
| Convergence criterion (GCONV=1E-8) satisfied. |

| Model Fit Statistics | | |
|---|---|---|
| **Criterion** | **Intercept Only** | **Intercept and Covariates** |
| **AIC** | 53054.996 | 52884.243 |
| **SC** | 53067.486 | 52909.222 |
| **-2 Log L** | 53052.996 | 52880.243 |

| Testing Global Null Hypothesis: BETA=0 | | | |
|---|---|---|---|
| **Test** | **Chi-Square** | **DF** | **Pr > ChiSq** |
| **Likelihood Ratio** | 172.7532 | 1 | <.0001 |
| **Score** | 171.3870 | 1 | <.0001 |
| **Wald** | 168.6455 | 1 | <.0001 |

#### The LOGISTIC Procedure

| Type 3 Analysis of Effects | | | |
|---|---|---|---|
| Effect | DF | Wald Chi-Square | Pr > ChiSq |
| firstdose | 1 | 168.6455 | <.0001 |

| Analysis of Maximum Likelihood Estimates | | | | | | | |
|---|---|---|---|---|---|---|---|
| Parameter | | DF | Estimate | Standard Error | Wald Chi-Square | Pr > ChiSq | Label |
| Intercept | | 1 | -6.5334 | 0.0265 | 60737.6346 | <.0001 | Intercept: CVD1YRD1=1 |
| firstdose | 1. PFIZER | 1 | 0.4414 | 0.0340 | 168.6455 | <.0001 | firstdose 1. PFIZER |

| Odds Ratio Estimates | | | |
|---|---|---|---|
| Effect | Point Estimate | 95% Wald Confidence Limits | |
| firstdose 1. PFIZER vs 2. MODERN | 1.555 | 1.455 | 1.662 |

| Association of Predicted Probabilities and Observed Responses | | | |
|---|---|---|---|
| Percent Concordant | 30.4 | Somers' D | 0.109 |
| Percent Discordant | 19.6 | Gamma | 0.217 |
| Percent Tied | 50.0 | Tau-a | 0.000 |
| Pairs | 7129466937 | c | 0.554 |

### CVD Death adjusted (logistic regression)

#### The LOGISTIC Procedure

| Model Information | |
|---|---|
| Data Set | WORK.MATCHEDCOHORT |
| Response Variable | CVD1YRD1 |
| Number of Response Levels | 2 |
| Model | binary logit |
| Optimization Technique | Fisher's scoring |

| | |
|---|---|
| Number of Observations Read | 1962822 |
| Number of Observations Used | 1962822 |

| Response Profile | | |
|---|---|---|
| Ordered Value | CVD1YRD1 | Total Frequency |
| 1 | 0 | 1959183 |
| 2 | 1 | 3639 |

**Probability modeled is CVD1YRD1=1.**

| Class Level Information | | | | | | | | | |
|---|---|---|---|---|---|---|---|---|---|
| Class | Value | Design Variables | | | | | | | |
| agenew | 18-39 | 0 | 0 | 0 | 0 | 0 | | | |
| | 40-49 | 1 | 0 | 0 | 0 | 0 | | | |
| | 50-59 | 0 | 1 | 0 | 0 | 0 | | | |
| | 60-69 | 0 | 0 | 1 | 0 | 0 | | | |
| | 70-79 | 0 | 0 | 0 | 1 | 0 | | | |
| | 80+ | 0 | 0 | 0 | 0 | 1 | | | |
| firstdose | 1. PFIZER | 1 | | | | | | | |
| | 2. MODERN | 0 | | | | | | | |
| sex2 | F | 1 | | | | | | | |
| | M | 0 | | | | | | | |
| RACE4 | BLACK | 1 | 0 | 0 | | | | | |
| | OTHERS | 0 | 1 | 0 | | | | | |
| | UNKNOWN | 0 | 0 | 1 | | | | | |
| | WHITE | 0 | 0 | 0 | | | | | |
| ETHNICITY3 | HISPANIC | 1 | 0 | | | | | | |
| | NOT HISPANIC | 0 | 0 | | | | | | |
| | UNKNOWN | 0 | 1 | | | | | | |
| Vaccinationsite | CHD OR DOH | 1 | 0 | 0 | 0 | 0 | 0 | 0 | 0 |
| | DOCTOR CLINIC | 0 | 1 | 0 | 0 | 0 | 0 | 0 | 0 |

#### The LOGISTIC Procedure

| Class Level Information | | | | | | | | | |
|---|---|---|---|---|---|---|---|---|---|
| Class | Value | Design Variables | | | | | | | |
| | FIRE RESQUE OR EMS | 0 | 0 | 1 | 0 | 0 | 0 | 0 | 0 |
| | FQHC OR RHC OR CHC | 0 | 0 | 0 | 1 | 0 | 0 | 0 | 0 |
| | HOSPITAL CLINIC | 0 | 0 | 0 | 0 | 1 | 0 | 0 | 0 |
| | HOSPITAL ER | 0 | 0 | 0 | 0 | 0 | 1 | 0 | 0 |
| | OTHER PHARMACIES | 0 | 0 | 0 | 0 | 0 | 0 | 1 | 0 |
| | OTHERS OR MISC | 0 | 0 | 0 | 0 | 0 | 0 | 0 | 1 |
| | PUBLIX OR CVS OR WALGREENS | 0 | 0 | 0 | 0 | 0 | 0 | 0 | 0 |

| Model Convergence Status |
|---|
| Convergence criterion (GCONV=1E-8) satisfied. |

| Model Fit Statistics | | |
|---|---|---|
| Criterion | Intercept Only | Intercept and Covariates |
| AIC | 53054.996 | 44667.351 |
| SC | 53067.486 | 44967.108 |
| -2 Log L | 53052.996 | 44619.351 |

| Testing Global Null Hypothesis: BETA=0 | | | |
|---|---|---|---|
| Test | Chi-Square | DF | Pr > ChiSq |
| Likelihood Ratio | 8433.6450 | 23 | <.0001 |
| Score | 15433.7488 | 23 | <.0001 |
| Wald | 6917.0518 | 23 | <.0001 |

| Type 3 Analysis of Effects | | | |
|---|---|---|---|
| Effect | DF | Wald Chi-Square | Pr > ChiSq |
| firstdose | 1 | 170.1471 | <.0001 |
| agenew | 5 | 2028.1076 | <.0001 |
| sex2 | 1 | 132.2635 | <.0001 |
| RACE4 | 3 | 72.4480 | <.0001 |
| ETHNICITY3 | 2 | 437.1980 | <.0001 |
| decile_sviscore1 | 1 | 82.2229 | <.0001 |
| decile_sviscore4 | 1 | 0.2936 | 0.5879 |
| mortalityratepct | 1 | 403.6260 | <.0001 |
| Vaccinationsite | 8 | 485.9043 | <.0001 |

#### The LOGISTIC Procedure

| Analysis of Maximum Likelihood Estimates | | | | | | | |
|---|---|---|---|---|---|---|---|
| **Parameter** | | **DF** | **Estimate** | **Standard Error** | **Wald Chi-Square** | **Pr > ChiSq** | **Label** |
| **Intercept** | | 1 | -10.2862 | 0.2053 | 2509.4051 | <.0001 | Intercept: CVD1YRD1=1 |
| **firstdose** | **1. PFIZER** | 1 | 0.4459 | 0.0342 | 170.1471 | <.0001 | firstdose 1. PFIZER |
| **agenew** | **40-49** | 1 | 1.8250 | 0.2298 | 63.0749 | <.0001 | agenew 40-49 |
| **agenew** | **50-59** | 1 | 2.5446 | 0.2102 | 146.4958 | <.0001 | agenew 50-59 |
| **agenew** | **60-69** | 1 | 3.4981 | 0.2043 | 293.2648 | <.0001 | agenew 60-69 |
| **agenew** | **70-79** | 1 | 4.3919 | 0.2037 | 464.9682 | <.0001 | agenew 70-79 |
| **agenew** | **80+** | 1 | 5.2474 | 0.2071 | 642.2345 | <.0001 | agenew 80+ |
| **sex2** | **F** | 1 | -0.3940 | 0.0343 | 132.2635 | <.0001 | sex2 F |
| **RACE4** | **BLACK** | 1 | 0.1260 | 0.0794 | 2.5194 | 0.1125 | RACE4 BLACK |
| **RACE4** | **OTHERS** | 1 | -0.5534 | 0.0675 | 67.2449 | <.0001 | RACE4 OTHERS |
| **RACE4** | **UNKNOWN** | 1 | 0.0453 | 0.0923 | 0.2406 | 0.6238 | RACE4 UNKNOWN |
| **ETHNICITY3** | **HISPANIC** | 1 | -0.4949 | 0.0657 | 56.6871 | <.0001 | ETHNICITY3 HISPANIC |
| **ETHNICITY3** | **UNKNOWN** | 1 | 0.9676 | 0.0529 | 334.7993 | <.0001 | ETHNICITY3 UNKNOWN |
| **decile_sviscore1** | | 1 | 0.0670 | 0.00739 | 82.2229 | <.0001 | Rank for Variable SVISCORE1 |
| **decile_sviscore4** | | 1 | 0.00376 | 0.00694 | 0.2936 | 0.5879 | Rank for Variable SVISCORE4 |
| **mortalityratepct** | | 1 | 0.0889 | 0.00442 | 403.6260 | <.0001 | |
| **Vaccinationsite** | **CHD OR DOH** | 1 | -0.3313 | 0.0479 | 47.9193 | <.0001 | Vaccinationsite CHD OR DOH |
| **Vaccinationsite** | **DOCTOR CLINIC** | 1 | 0.0659 | 0.0455 | 2.0931 | 0.1480 | Vaccinationsite DOCTOR CLINIC |
| **Vaccinationsite** | **FIRE RESQUE OR EMS** | 1 | -0.3611 | 0.0945 | 14.6150 | 0.0001 | Vaccinationsite FIRE RESQUE OR EMS |
| **Vaccinationsite** | **FQHC OR RHC OR CHC** | 1 | -0.7315 | 0.1798 | 16.5446 | <.0001 | Vaccinationsite FQHC OR RHC OR CHC |
| **Vaccinationsite** | **HOSPITAL CLINIC** | 1 | -0.2943 | 0.0764 | 14.8337 | 0.0001 | Vaccinationsite HOSPITAL CLINIC |
| **Vaccinationsite** | **HOSPITAL ER** | 1 | 0.1558 | 0.2804 | 0.3088 | 0.5784 | Vaccinationsite HOSPITAL ER |
| **Vaccinationsite** | **OTHER PHARMACIES** | 1 | 0.3405 | 0.1008 | 11.3996 | 0.0007 | Vaccinationsite OTHER PHARMACIES |
| **Vaccinationsite** | **OTHERS OR MISC** | 1 | -1.6645 | 0.0849 | 383.9588 | <.0001 | Vaccinationsite OTHERS OR MISC |

| Odds Ratio Estimates | | | |
|---|---|---|---|
| **Effect** | **Point Estimate** | **95% Wald Confidence Limits** | |
| **firstdose 1. PFIZER vs 2. MODERN** | 1.562 | 1.461 | 1.670 |
| **agenew 40-49 vs 18-39** | 6.202 | 3.953 | 9.731 |
| **agenew 50-59 vs 18-39** | 12.738 | 8.436 | 19.233 |
| **agenew 60-69 vs 18-39** | 33.052 | 22.147 | 49.325 |
| **agenew 70-79 vs 18-39** | 80.795 | 54.202 | 120.436 |
| **agenew 80+ vs 18-39** | 190.072 | 126.668 | 285.212 |
| **sex2 F vs M** | 0.674 | 0.631 | 0.721 |

#### The LOGISTIC Procedure

| Odds Ratio Estimates | | | |
|---|---|---|---|
| Effect | Point Estimate | 95% Wald Confidence Limits | |
| RACE4 BLACK vs WHITE | 1.134 | 0.971 | 1.325 |
| RACE4 OTHERS vs WHITE | 0.575 | 0.504 | 0.656 |
| RACE4 UNKNOWN vs WHITE | 1.046 | 0.873 | 1.254 |
| ETHNICITY3 HISPANIC vs NOT HISPANIC | 0.610 | 0.536 | 0.693 |
| ETHNICITY3 UNKNOWN vs NOT HISPANIC | 2.631 | 2.372 | 2.919 |
| decile_sviscore1 | 1.069 | 1.054 | 1.085 |
| decile_sviscore4 | 1.004 | 0.990 | 1.018 |
| mortalityratepct | 1.093 | 1.084 | 1.102 |
| Vaccinationsite CHD OR DOH vs PUBLIX OR CVS OR WALGREENS | 0.718 | 0.654 | 0.789 |
| Vaccinationsite DOCTOR CLINIC vs PUBLIX OR CVS OR WALGREENS | 1.068 | 0.977 | 1.168 |
| Vaccinationsite FIRE RESQUE OR EMS vs PUBLIX OR CVS OR WALGREENS | 0.697 | 0.579 | 0.839 |
| Vaccinationsite FQHC OR RHC OR CHC vs PUBLIX OR CVS OR WALGREENS | 0.481 | 0.338 | 0.685 |
| Vaccinationsite HOSPITAL CLINIC vs PUBLIX OR CVS OR WALGREENS | 0.745 | 0.641 | 0.865 |
| Vaccinationsite HOSPITAL ER vs PUBLIX OR CVS OR WALGREENS | 1.169 | 0.675 | 2.024 |
| Vaccinationsite OTHER PHARMACIES vs PUBLIX OR CVS OR WALGREENS | 1.406 | 1.154 | 1.713 |
| Vaccinationsite OTHERS OR MISC vs PUBLIX OR CVS OR WALGREENS | 0.189 | 0.160 | 0.224 |

| Association of Predicted Probabilities and Observed Responses | | | |
|---|---|---|---|
| Percent Concordant | 86.8 | Somers' D | 0.737 |
| Percent Discordant | 13.2 | Gamma | 0.737 |
| Percent Tied | 0.0 | Tau-a | 0.003 |
| Pairs | 7129466937 | c | 0.868 |

### CVD deaths adjusted by Age

#### The LOGISTIC Procedure

**agenew=18-39 years**

| Model Information | |
|---|---|
| **Data Set** | WORK.MATCHEDCOHORT |
| **Response Variable** | CVD1YRD1 |
| **Number of Response Levels** | 2 |
| **Model** | binary logit |
| **Optimization Technique** | Fisher's scoring |

| | |
|---|---|
| **Number of Observations Read** | 492484 |
| **Number of Observations Used** | 492484 |

| Response Profile | | |
|---|---|---|
| **Ordered Value** | **CVD1YRD1** | **Total Frequency** |
| **1** | 0 | 492459 |
| **2** | 1 | 25 |

**Probability modeled is CVD1YRD1=1.**

| Class Level Information | | | | | | | | | |
|---|---|---|---|---|---|---|---|---|---|
| **Class** | **Value** | **Design Variables** | | | | | | | |
| **agenew** | **18-39 years** | | | | | | | | |
| **firstdose** | **1. PFIZER** | 1 | | | | | | | |
| | **2. MODERN** | 0 | | | | | | | |
| **sex2** | **F** | 1 | | | | | | | |
| | **M** | 0 | | | | | | | |
| **RACE4** | **BLACK** | 1 | 0 | 0 | | | | | |
| | **OTHERS** | 0 | 1 | 0 | | | | | |
| | **UNKNOWN** | 0 | 0 | 1 | | | | | |
| | **WHITE** | 0 | 0 | 0 | | | | | |
| **ETHNICITY3** | **HISPANIC** | 1 | 0 | | | | | | |
| | **NOT HISPANIC** | 0 | 0 | | | | | | |
| | **UNKNOWN** | 0 | 1 | | | | | | |
| **Vaccinationsite** | **CHD OR DOH** | 1 | 0 | 0 | 0 | 0 | 0 | 0 | 0 |
| | **DOCTOR CLINIC** | 0 | 1 | 0 | 0 | 0 | 0 | 0 | 0 |
| | **FIRE RESQUE OR EMS** | 0 | 0 | 1 | 0 | 0 | 0 | 0 | 0 |
| | **FQHC OR RHC OR CHC** | 0 | 0 | 0 | 1 | 0 | 0 | 0 | 0 |
| | **HOSPITAL CLINIC** | 0 | 0 | 0 | 0 | 1 | 0 | 0 | 0 |
| | **HOSPITAL ER** | 0 | 0 | 0 | 0 | 0 | 1 | 0 | 0 |

### CVD deaths adjusted by Age

#### The LOGISTIC Procedure

**agenew=18-39 years**

| Class Level Information | | | | | | | | | |
|---|---|---|---|---|---|---|---|---|---|
| **Class** | **Value** | **Design Variables** | | | | | | | |
| | **OTHER PHARMACIES** | 0 | 0 | 0 | 0 | 0 | 0 | 1 | 0 |
| | **OTHERS OR MISC** | 0 | 0 | 0 | 0 | 0 | 0 | 0 | 1 |
| | **PUBLIX OR CVS OR WALGREENS** | 0 | 0 | 0 | 0 | 0 | 0 | 0 | 0 |

| Model Convergence Status |
|---|
| Quasi-complete separation of data points detected. |

**Warning:** The maximum likelihood estimate may not exist.

**Warning:** The LOGISTIC procedure continues in spite of the above warning. Results shown are based on the last maximum likelihood iteration. Validity of the model fit is questionable.

| Model Fit Statistics | | |
|---|---|---|
| **Criterion** | **Intercept Only** | **Intercept and Covariates** |
| **AIC** | 546.416 | 550.731 |
| **SC** | 557.523 | 761.769 |
| **-2 Log L** | 544.416 | 512.731 |

| Testing Global Null Hypothesis: BETA=0 | | | |
|---|---|---|---|
| **Test** | **Chi-Square** | **DF** | **Pr > ChiSq** |
| **Likelihood Ratio** | 31.6843 | 18 | 0.0240 |
| **Score** | 32.6035 | 18 | 0.0186 |
| **Wald** | 25.1067 | 18 | 0.1220 |

| Type 3 Analysis of Effects | | | |
|---|---|---|---|
| **Effect** | **DF** | **Wald Chi-Square** | **Pr > ChiSq** |
| **firstdose** | 1 | 0.9871 | 0.3204 |
| **agenew** | 0 | . | . |
| **sex2** | 1 | 2.5248 | 0.1121 |
| **RACE4** | 3 | 1.9846 | 0.5756 |
| **ETHNICITY3** | 2 | 3.0713 | 0.2153 |
| **decile_sviscore1** | 1 | 8.0093 | 0.0047 |
| **decile_sviscore4** | 1 | 0.0345 | 0.8526 |
| **mortalityratepct** | 1 | 0.0000 | 0.9957 |
| **Vaccinationsite** | 8 | 2.8695 | 0.9423 |

#### The LOGISTIC Procedure

**Warning:** The validity of the model fit is questionable.

**agenew=18-39 years**

| Analysis of Maximum Likelihood Estimates | | | | | | | |
|---|---|---|---|---|---|---|---|
| Parameter | | DF | Estimate | Standard Error | Wald Chi-Square | Pr > ChiSq | Label |
| Intercept | | 1 | -11.0096 | 0.6581 | 279.8918 | <.0001 | Intercept: CVD1YRD1=1 |
| firstdose | 1. PFIZER | 1 | 0.4056 | 0.4083 | 0.9871 | 0.3204 | firstdose 1. PFIZER |
| sex2 | F | 1 | -0.6520 | 0.4103 | 2.5248 | 0.1121 | sex2 F |
| RACE4 | BLACK | 1 | 0.6691 | 0.5191 | 1.6614 | 0.1974 | RACE4 BLACK |
| RACE4 | OTHERS | 1 | -0.1959 | 0.6431 | 0.0928 | 0.7607 | RACE4 OTHERS |
| RACE4 | UNKNOWN | 1 | -0.0430 | 1.1229 | 0.0015 | 0.9695 | RACE4 UNKNOWN |
| ETHNICITY3 | HISPANIC | 1 | -1.1104 | 0.6594 | 2.8356 | 0.0922 | ETHNICITY3 HISPANIC |
| ETHNICITY3 | UNKNOWN | 1 | 0.0625 | 0.5669 | 0.0121 | 0.9123 | ETHNICITY3 UNKNOWN |
| decile_sviscore1 | | 1 | 0.2765 | 0.0977 | 8.0093 | 0.0047 | Rank for Variable SVISCORE1 |
| decile_sviscore4 | | 1 | -0.0158 | 0.0850 | 0.0345 | 0.8526 | Rank for Variable SVISCORE4 |
| mortalityratepct | | 1 | -12.4365 | 2317.7 | 0.0000 | 0.9957 | |
| Vaccinationsite | CHD OR DOH | 1 | 0.9776 | 0.6332 | 2.3836 | 0.1226 | Vaccinationsite CHD OR DOH |
| Vaccinationsite | DOCTOR CLINIC | 1 | 0.00920 | 0.6265 | 0.0002 | 0.9883 | Vaccinationsite DOCTOR CLINIC |
| Vaccinationsite | FIRE RESQUE OR EMS | 1 | -12.3029 | 3095.3 | 0.0000 | 0.9968 | Vaccinationsite FIRE RESQUE OR EMS |
| Vaccinationsite | FQHC OR RHC OR CHC | 1 | -12.2473 | 792.8 | 0.0002 | 0.9877 | Vaccinationsite FQHC OR RHC OR CHC |
| Vaccinationsite | HOSPITAL CLINIC | 1 | -12.2050 | 548.2 | 0.0005 | 0.9822 | Vaccinationsite HOSPITAL CLINIC |
| Vaccinationsite | HOSPITAL ER | 1 | -12.4653 | 2349.5 | 0.0000 | 0.9958 | Vaccinationsite HOSPITAL ER |
| Vaccinationsite | OTHER PHARMACIES | 1 | -0.5587 | 1.0346 | 0.2916 | 0.5892 | Vaccinationsite OTHER PHARMACIES |
| Vaccinationsite | OTHERS OR MISC | 1 | -12.5873 | 438.3 | 0.0008 | 0.9771 | Vaccinationsite OTHERS OR MISC |

| Odds Ratio Estimates | | | |
|---|---|---|---|
| Effect | Point Estimate | 95% Wald Confidence Limits | |
| firstdose 1. PFIZER vs 2. MODERN | 1.500 | 0.674 | 3.340 |
| sex2 F vs M | 0.521 | 0.233 | 1.164 |
| RACE4 BLACK vs WHITE | 1.953 | 0.706 | 5.401 |
| RACE4 OTHERS vs WHITE | 0.822 | 0.233 | 2.899 |
| RACE4 UNKNOWN vs WHITE | 0.958 | 0.106 | 8.653 |
| ETHNICITY3 HISPANIC vs NOT HISPANIC | 0.329 | 0.090 | 1.200 |
| ETHNICITY3 UNKNOWN vs NOT HISPANIC | 1.064 | 0.350 | 3.234 |
| decile_sviscore1 | 1.319 | 1.089 | 1.597 |
| decile_sviscore4 | 0.984 | 0.833 | 1.163 |
| mortalityratepct | <0.001 | <0.001 | >999.999 |
| Vaccinationsite CHD OR DOH vs PUBLIX OR CVS OR WALGREENS | 2.658 | 0.768 | 9.195 |

#### The LOGISTIC Procedure

**Warning:** The validity of the model fit is questionable.

**agenew=18-39 years**

| Odds Ratio Estimates | | | |
|---|---|---|---|
| Effect | Point Estimate | 95% Wald Confidence Limits | |
| Vaccinationsite DOCTOR CLINIC vs PUBLIX OR CVS OR WALGREENS | 1.009 | 0.296 | 3.446 |
| Vaccinationsite FIRE RESQUE OR EMS vs PUBLIX OR CVS OR WALGREENS | <0.001 | <0.001 | >999.999 |
| Vaccinationsite FQHC OR RHC OR CHC vs PUBLIX OR CVS OR WALGREENS | <0.001 | <0.001 | >999.999 |
| Vaccinationsite HOSPITAL CLINIC vs PUBLIX OR CVS OR WALGREENS | <0.001 | <0.001 | >999.999 |
| Vaccinationsite HOSPITAL ER vs PUBLIX OR CVS OR WALGREENS | <0.001 | <0.001 | >999.999 |
| Vaccinationsite OTHER PHARMACIES vs PUBLIX OR CVS OR WALGREENS | 0.572 | 0.075 | 4.345 |
| Vaccinationsite OTHERS OR MISC vs PUBLIX OR CVS OR WALGREENS | <0.001 | <0.001 | >999.999 |

| Association of Predicted Probabilities and Observed Responses | | | |
|---|---|---|---|
| Percent Concordant | 81.1 | Somers' D | 0.621 |
| Percent Discordant | 18.9 | Gamma | 0.622 |
| Percent Tied | 0.0 | Tau-a | 0.000 |
| Pairs | 12311475 | c | 0.811 |

#### The LOGISTIC Procedure

**agenew=40-59 years**

| Model Information | |
|---|---|
| Data Set | WORK.MATCHEDCOHORT |
| Response Variable | CVD1YRD1 |
| Number of Response Levels | 2 |
| Model | binary logit |
| Optimization Technique | Fisher's scoring |

| | |
|---|---|
| Number of Observations Read | 634102 |
| Number of Observations Used | 634102 |

| Response Profile | | |
|---|---|---|
| Ordered Value | CVD1YRD1 | Total Frequency |
| 1 | 0 | 633784 |
| 2 | 1 | 318 |

**Probability modeled is CVD1YRD1=1.**

| Class Level Information | | | | | | | | | |
|---|---|---|---|---|---|---|---|---|---|
| Class | Value | Design Variables | | | | | | | |
| agenew | 40-59 years | | | | | | | | |
| firstdose | 1. PFIZER | 1 | | | | | | | |
| | 2. MODERN | 0 | | | | | | | |
| sex2 | F | 1 | | | | | | | |
| | M | 0 | | | | | | | |
| RACE4 | BLACK | 1 | 0 | 0 | | | | | |
| | OTHERS | 0 | 1 | 0 | | | | | |
| | UNKNOWN | 0 | 0 | 1 | | | | | |
| | WHITE | 0 | 0 | 0 | | | | | |
| ETHNICITY3 | HISPANIC | 1 | 0 | | | | | | |
| | NOT HISPANIC | 0 | 0 | | | | | | |
| | UNKNOWN | 0 | 1 | | | | | | |
| Vaccinationsite | CHD OR DOH | 1 | 0 | 0 | 0 | 0 | 0 | 0 | 0 |
| | DOCTOR CLINIC | 0 | 1 | 0 | 0 | 0 | 0 | 0 | 0 |
| | FIRE RESQUE OR EMS | 0 | 0 | 1 | 0 | 0 | 0 | 0 | 0 |
| | FQHC OR RHC OR CHC | 0 | 0 | 0 | 1 | 0 | 0 | 0 | 0 |
| | HOSPITAL CLINIC | 0 | 0 | 0 | 0 | 1 | 0 | 0 | 0 |
| | HOSPITAL ER | 0 | 0 | 0 | 0 | 0 | 1 | 0 | 0 |

#### The LOGISTIC Procedure

**agenew=40-59 years**

| Class Level Information | | | | | | | | | |
|---|---|---|---|---|---|---|---|---|---|
| Class | Value | Design Variables | | | | | | | |
| | OTHER PHARMACIES | 0 | 0 | 0 | 0 | 0 | 0 | 1 | 0 |
| | OTHERS OR MISC | 0 | 0 | 0 | 0 | 0 | 0 | 0 | 1 |
| | PUBLIX OR CVS OR WALGREENS | 0 | 0 | 0 | 0 | 0 | 0 | 0 | 0 |

| Model Convergence Status |
|---|
| Convergence criterion (GCONV=1E-8) satisfied. |

| Model Fit Statistics | | |
|---|---|---|
| Criterion | Intercept Only | Intercept and Covariates |
| AIC | 5470.114 | 5253.419 |
| SC | 5481.474 | 5469.258 |
| -2 Log L | 5468.114 | 5215.419 |

| Testing Global Null Hypothesis: BETA=0 | | | |
|---|---|---|---|
| Test | Chi-Square | DF | Pr > ChiSq |
| Likelihood Ratio | 252.6950 | 18 | <.0001 |
| Score | 260.9674 | 18 | <.0001 |
| Wald | 243.4141 | 18 | <.0001 |

| Type 3 Analysis of Effects | | | |
|---|---|---|---|
| Effect | DF | Wald Chi-Square | Pr > ChiSq |
| firstdose | 1 | 1.5505 | 0.2131 |
| agenew | 0 | . | . |
| sex2 | 1 | 26.4633 | <.0001 |
| RACE4 | 3 | 2.4541 | 0.4836 |
| ETHNICITY3 | 2 | 38.1191 | <.0001 |
| decile_sviscore1 | 1 | 36.5211 | <.0001 |
| decile_sviscore4 | 1 | 0.1020 | 0.7495 |
| mortalityratepct | 1 | 43.9136 | <.0001 |
| Vaccinationsite | 8 | 21.8787 | 0.0051 |

#### The LOGISTIC Procedure

**agenew=40-59 years**

| Analysis of Maximum Likelihood Estimates | | | | | | | |
|---|---|---|---|---|---|---|---|
| Parameter | | DF | Estimate | Standard Error | Wald Chi-Square | Pr > ChiSq | Label |
| Intercept | | 1 | -8.5175 | 0.1813 | 2206.7256 | <.0001 | Intercept: CVD1YRD1=1 |
| firstdose | 1. PFIZER | 1 | 0.1401 | 0.1125 | 1.5505 | 0.2131 | firstdose 1. PFIZER |
| sex2 | F | 1 | -0.6479 | 0.1259 | 26.4633 | <.0001 | sex2 F |
| RACE4 | BLACK | 1 | 0.1399 | 0.1779 | 0.6187 | 0.4315 | RACE4 BLACK |
| RACE4 | OTHERS | 1 | -0.2064 | 0.2127 | 0.9419 | 0.3318 | RACE4 OTHERS |
| RACE4 | UNKNOWN | 1 | -0.2812 | 0.3051 | 0.8496 | 0.3567 | RACE4 UNKNOWN |
| ETHNICITY3 | HISPANIC | 1 | -0.6973 | 0.1908 | 13.3615 | 0.0003 | ETHNICITY3 HISPANIC |
| ETHNICITY3 | UNKNOWN | 1 | 0.6638 | 0.1668 | 15.8467 | <.0001 | ETHNICITY3 UNKNOWN |
| decile_sviscore1 | | 1 | 0.1564 | 0.0259 | 36.5211 | <.0001 | Rank for Variable SVISCORE1 |
| decile_sviscore4 | | 1 | 0.00767 | 0.0240 | 0.1020 | 0.7495 | Rank for Variable SVISCORE4 |
| mortalityratepct | | 1 | 0.7767 | 0.1172 | 43.9136 | <.0001 | |
| Vaccinationsite | CHD OR DOH | 1 | 0.2910 | 0.2626 | 1.2279 | 0.2678 | Vaccinationsite CHD OR DOH |
| Vaccinationsite | DOCTOR CLINIC | 1 | -0.0203 | 0.1562 | 0.0169 | 0.8965 | Vaccinationsite DOCTOR CLINIC |
| Vaccinationsite | FIRE RESQUE OR EMS | 1 | -10.5368 | 228.7 | 0.0021 | 0.9633 | Vaccinationsite FIRE RESQUE OR EMS |
| Vaccinationsite | FQHC OR RHC OR CHC | 1 | -0.0389 | 0.4580 | 0.0072 | 0.9323 | Vaccinationsite FQHC OR RHC OR CHC |
| Vaccinationsite | HOSPITAL CLINIC | 1 | 0.1984 | 0.2621 | 0.5726 | 0.4492 | Vaccinationsite HOSPITAL CLINIC |
| Vaccinationsite | HOSPITAL ER | 1 | 0.1957 | 1.0035 | 0.0380 | 0.8454 | Vaccinationsite HOSPITAL ER |
| Vaccinationsite | OTHER PHARMACIES | 1 | 0.4379 | 0.2064 | 4.4991 | 0.0339 | Vaccinationsite OTHER PHARMACIES |
| Vaccinationsite | OTHERS OR MISC | 1 | -1.2967 | 0.3444 | 14.1773 | 0.0002 | Vaccinationsite OTHERS OR MISC |

| Odds Ratio Estimates | | | |
|---|---|---|---|
| Effect | Point Estimate | 95% Wald Confidence Limits | |
| firstdose 1. PFIZER vs 2. MODERN | 1.150 | 0.923 | 1.434 |
| sex2 F vs M | 0.523 | 0.409 | 0.670 |
| RACE4 BLACK vs WHITE | 1.150 | 0.812 | 1.630 |
| RACE4 OTHERS vs WHITE | 0.813 | 0.536 | 1.234 |
| RACE4 UNKNOWN vs WHITE | 0.755 | 0.415 | 1.373 |
| ETHNICITY3 HISPANIC vs NOT HISPANIC | 0.498 | 0.343 | 0.724 |
| ETHNICITY3 UNKNOWN vs NOT HISPANIC | 1.942 | 1.401 | 2.693 |
| decile_sviscore1 | 1.169 | 1.111 | 1.230 |
| decile_sviscore4 | 1.008 | 0.961 | 1.056 |
| mortalityratepct | 2.174 | 1.728 | 2.736 |
| Vaccinationsite CHD OR DOH vs PUBLIX OR CVS OR WALGREENS | 1.338 | 0.800 | 2.238 |

#### The LOGISTIC Procedure

**agenew=40-59 years**

| Odds Ratio Estimates | | | |
|---|---|---|---|
| Effect | Point Estimate | 95% Wald Confidence Limits | |
| Vaccinationsite DOCTOR CLINIC vs PUBLIX OR CVS OR WALGREENS | 0.980 | 0.721 | 1.331 |
| Vaccinationsite FIRE RESQUE OR EMS vs PUBLIX OR CVS OR WALGREENS | <0.001 | <0.001 | >999.999 |
| Vaccinationsite FQHC OR RHC OR CHC vs PUBLIX OR CVS OR WALGREENS | 0.962 | 0.392 | 2.360 |
| Vaccinationsite HOSPITAL CLINIC vs PUBLIX OR CVS OR WALGREENS | 1.219 | 0.729 | 2.038 |
| Vaccinationsite HOSPITAL ER vs PUBLIX OR CVS OR WALGREENS | 1.216 | 0.170 | 8.694 |
| Vaccinationsite OTHER PHARMACIES vs PUBLIX OR CVS OR WALGREENS | 1.549 | 1.034 | 2.322 |
| Vaccinationsite OTHERS OR MISC vs PUBLIX OR CVS OR WALGREENS | 0.273 | 0.139 | 0.537 |

| Association of Predicted Probabilities and Observed Responses | | | |
|---|---|---|---|
| Percent Concordant | 74.3 | Somers' D | 0.486 |
| Percent Discordant | 25.7 | Gamma | 0.487 |
| Percent Tied | 0.0 | Tau-a | 0.000 |
| Pairs | 201543312 | c | 0.743 |

### CVD deaths adjusted by Age

#### The LOGISTIC Procedure

**agenew=60+ years**

| Model Information | |
|---|---|
| **Data Set** | WORK.MATCHEDCOHORT |
| **Response Variable** | CVD1YRD1 |
| **Number of Response Levels** | 2 |
| **Model** | binary logit |
| **Optimization Technique** | Fisher's scoring |

| | |
|---|---|
| **Number of Observations Read** | 836236 |
| **Number of Observations Used** | 836236 |

| Response Profile | | |
|---|---|---|
| **Ordered Value** | **CVD1YRD1** | **Total Frequency** |
| **1** | 0 | 832940 |
| **2** | 1 | 3296 |

**Probability modeled is CVD1YRD1=1.**

| Class Level Information | | | | | | | | | |
|---|---|---|---|---|---|---|---|---|---|
| **Class** | **Value** | **Design Variables** | | | | | | | |
| **agenew** | **60+ years** | | | | | | | | |
| **firstdose** | **1. PFIZER** | 1 | | | | | | | |
| | **2. MODERN** | 0 | | | | | | | |
| **sex2** | **F** | 1 | | | | | | | |
| | **M** | 0 | | | | | | | |
| **RACE4** | **BLACK** | 1 | 0 | 0 | | | | | |
| | **OTHERS** | 0 | 1 | 0 | | | | | |
| | **UNKNOWN** | 0 | 0 | 1 | | | | | |
| | **WHITE** | 0 | 0 | 0 | | | | | |
| **ETHNICITY3** | **HISPANIC** | 1 | 0 | | | | | | |
| | **NOT HISPANIC** | 0 | 0 | | | | | | |
| | **UNKNOWN** | 0 | 1 | | | | | | |
| **Vaccinationsite** | **CHD OR DOH** | 1 | 0 | 0 | 0 | 0 | 0 | 0 | 0 |
| | **DOCTOR CLINIC** | 0 | 1 | 0 | 0 | 0 | 0 | 0 | 0 |
| | **FIRE RESQUE OR EMS** | 0 | 0 | 1 | 0 | 0 | 0 | 0 | 0 |
| | **FQHC OR RHC OR CHC** | 0 | 0 | 0 | 1 | 0 | 0 | 0 | 0 |
| | **HOSPITAL CLINIC** | 0 | 0 | 0 | 0 | 1 | 0 | 0 | 0 |
| | **HOSPITAL ER** | 0 | 0 | 0 | 0 | 0 | 1 | 0 | 0 |

#### The LOGISTIC Procedure

**agenew=60+ years**

| Class Level Information | | | | | | | | | |
|---|---|---|---|---|---|---|---|---|---|
| Class | Value | Design Variables | | | | | | | |
| | OTHER PHARMACIES | 0 | 0 | 0 | 0 | 0 | 0 | 1 | 0 |
| | OTHERS OR MISC | 0 | 0 | 0 | 0 | 0 | 0 | 0 | 1 |
| | PUBLIX OR CVS OR WALGREENS | 0 | 0 | 0 | 0 | 0 | 0 | 0 | 0 |

| Model Convergence Status |
|---|
| Convergence criterion (GCONV=1E-8) satisfied. |

| Model Fit Statistics | | |
|---|---|---|
| Criterion | Intercept Only | Intercept and Covariates |
| AIC | 43075.631 | 39696.716 |
| SC | 43087.267 | 39917.813 |
| -2 Log L | 43073.631 | 39658.716 |

| Testing Global Null Hypothesis: BETA=0 | | | |
|---|---|---|---|
| Test | Chi-Square | DF | Pr > ChiSq |
| Likelihood Ratio | 3414.9143 | 18 | <.0001 |
| Score | 6235.3574 | 18 | <.0001 |
| Wald | 3899.9536 | 18 | <.0001 |

| Type 3 Analysis of Effects | | | |
|---|---|---|---|
| Effect | DF | Wald Chi-Square | Pr > ChiSq |
| firstdose | 1 | 176.6634 | <.0001 |
| agenew | 0 | . | . |
| sex2 | 1 | 38.2573 | <.0001 |
| RACE4 | 3 | 115.7179 | <.0001 |
| ETHNICITY3 | 2 | 483.8196 | <.0001 |
| decile_sviscore1 | 1 | 30.7969 | <.0001 |
| decile_sviscore4 | 1 | 0.9114 | 0.3397 |
| mortalityratepct | 1 | 2863.6831 | <.0001 |
| Vaccinationsite | 8 | 394.6635 | <.0001 |

#### The LOGISTIC Procedure

**agenew=60+ years**

| Analysis of Maximum Likelihood Estimates | | | | | | | |
|---|---|---|---|---|---|---|---|
| **Parameter** | | **DF** | **Estimate** | **Standard Error** | **Wald Chi-Square** | **Pr > ChiSq** | **Label** |
| **Intercept** | | 1 | -6.2994 | 0.0544 | 13413.7106 | <.0001 | Intercept: CVD1YRD1=1 |
| **firstdose** | **1. PFIZER** | 1 | 0.4824 | 0.0363 | 176.6634 | <.0001 | firstdose 1. PFIZER |
| **sex2** | **F** | 1 | -0.2215 | 0.0358 | 38.2573 | <.0001 | sex2 F |
| **RACE4** | **BLACK** | 1 | -0.1286 | 0.0901 | 2.0355 | 0.1537 | RACE4 BLACK |
| **RACE4** | **OTHERS** | 1 | -0.8615 | 0.0816 | 111.4008 | <.0001 | RACE4 OTHERS |
| **RACE4** | **UNKNOWN** | 1 | 0.1052 | 0.0972 | 1.1732 | 0.2787 | RACE4 UNKNOWN |
| **ETHNICITY3** | **HISPANIC** | 1 | -0.6805 | 0.0773 | 77.5818 | <.0001 | ETHNICITY3 HISPANIC |
| **ETHNICITY3** | **UNKNOWN** | 1 | 1.0594 | 0.0558 | 360.8263 | <.0001 | ETHNICITY3 UNKNOWN |
| **decile_sviscore1** | | 1 | 0.0432 | 0.00779 | 30.7969 | <.0001 | Rank for Variable SVISCORE1 |
| **decile_sviscore4** | | 1 | 0.00701 | 0.00734 | 0.9114 | 0.3397 | Rank for Variable SVISCORE4 |
| **mortalityratepct** | | 1 | 0.1685 | 0.00315 | 2863.6831 | <.0001 | |
| **Vaccinationsite** | **CHD OR DOH** | 1 | -0.1676 | 0.0484 | 11.9725 | 0.0005 | Vaccinationsite CHD OR DOH |
| **Vaccinationsite** | **DOCTOR CLINIC** | 1 | 0.1893 | 0.0478 | 15.6856 | <.0001 | Vaccinationsite DOCTOR CLINIC |
| **Vaccinationsite** | **FIRE RESQUE OR EMS** | 1 | -0.0412 | 0.0957 | 0.1851 | 0.6670 | Vaccinationsite FIRE RESQUE OR EMS |
| **Vaccinationsite** | **FQHC OR RHC OR CHC** | 1 | -0.6522 | 0.1954 | 11.1347 | 0.0008 | Vaccinationsite FQHC OR RHC OR CHC |
| **Vaccinationsite** | **HOSPITAL CLINIC** | 1 | -0.3052 | 0.0832 | 13.4467 | 0.0002 | Vaccinationsite HOSPITAL CLINIC |
| **Vaccinationsite** | **HOSPITAL ER** | 1 | 0.2085 | 0.2915 | 0.5119 | 0.4743 | Vaccinationsite HOSPITAL ER |
| **Vaccinationsite** | **OTHER PHARMACIES** | 1 | 0.2176 | 0.1166 | 3.4848 | 0.0619 | Vaccinationsite OTHER PHARMACIES |
| **Vaccinationsite** | **OTHERS OR MISC** | 1 | -1.5294 | 0.0878 | 303.2832 | <.0001 | Vaccinationsite OTHERS OR MISC |

| Odds Ratio Estimates | | | |
|---|---|---|---|
| **Effect** | **Point Estimate** | **95% Wald Confidence Limits** | |
| **firstdose 1. PFIZER vs 2. MODERN** | 1.620 | 1.509 | 1.739 |
| **sex2 F vs M** | 0.801 | 0.747 | 0.860 |
| **RACE4 BLACK vs WHITE** | 0.879 | 0.737 | 1.049 |
| **RACE4 OTHERS vs WHITE** | 0.423 | 0.360 | 0.496 |
| **RACE4 UNKNOWN vs WHITE** | 1.111 | 0.918 | 1.344 |
| **ETHNICITY3 HISPANIC vs NOT HISPANIC** | 0.506 | 0.435 | 0.589 |
| **ETHNICITY3 UNKNOWN vs NOT HISPANIC** | 2.885 | 2.586 | 3.218 |
| **decile_sviscore1** | 1.044 | 1.028 | 1.060 |
| **decile_sviscore4** | 1.007 | 0.993 | 1.022 |
| **mortalityratepct** | 1.184 | 1.176 | 1.191 |
| **Vaccinationsite CHD OR DOH vs PUBLIX OR CVS OR WALGREENS** | 0.846 | 0.769 | 0.930 |

#### The LOGISTIC Procedure

**agenew=60+ years**

| Odds Ratio Estimates | | | |
|---|---|---|---|
| Effect | Point Estimate | 95% Wald Confidence Limits | |
| Vaccinationsite DOCTOR CLINIC vs PUBLIX OR CVS OR WALGREENS | 1.208 | 1.100 | 1.327 |
| Vaccinationsite FIRE RESQUE OR EMS vs PUBLIX OR CVS OR WALGREENS | 0.960 | 0.796 | 1.158 |
| Vaccinationsite FQHC OR RHC OR CHC vs PUBLIX OR CVS OR WALGREENS | 0.521 | 0.355 | 0.764 |
| Vaccinationsite HOSPITAL CLINIC vs PUBLIX OR CVS OR WALGREENS | 0.737 | 0.626 | 0.868 |
| Vaccinationsite HOSPITAL ER vs PUBLIX OR CVS OR WALGREENS | 1.232 | 0.696 | 2.181 |
| Vaccinationsite OTHER PHARMACIES vs PUBLIX OR CVS OR WALGREENS | 1.243 | 0.989 | 1.562 |
| Vaccinationsite OTHERS OR MISC vs PUBLIX OR CVS OR WALGREENS | 0.217 | 0.182 | 0.257 |

| Association of Predicted Probabilities and Observed Responses | | | |
|---|---|---|---|
| Percent Concordant | 76.4 | Somers' D | 0.529 |
| Percent Discordant | 23.5 | Gamma | 0.529 |
| Percent Tied | 0.0 | Tau-a | 0.004 |
| Pairs | 2745370240 | c | 0.765 |

#### The LOGISTIC Procedure

| Model Information | |
|---|---|
| Data Set | WORK.MATCHEDCOHORT |
| Response Variable | COVIDDEATH1YRD1 |
| Number of Response Levels | 2 |
| Model | binary logit |
| Optimization Technique | Fisher's scoring |

| | |
|---|---|
| Number of Observations Read | 1962822 |
| Number of Observations Used | 1962822 |

| Response Profile | | |
|---|---|---|
| Ordered Value | COVIDDEATH1YRD1 | Total Frequency |
| 1 | 0 | 1961901 |
| 2 | 1 | 921 |

**Probability modeled is COVIDDEATH1YRD1=1.**

| Class Level Information | | |
|---|---|---|
| Class | Value | Design Variables |
| firstdose | 1. PFIZER | 1 |
| | 2. MODERN | 0 |

| Model Convergence Status |
|---|
| Convergence criterion (GCONV=1E-8) satisfied. |

| Model Fit Statistics | | |
|---|---|---|
| Criterion | Intercept Only | Intercept and Covariates |
| AIC | 15961.455 | 15873.757 |
| SC | 15973.945 | 15898.737 |
| -2 Log L | 15959.455 | 15869.757 |

| Testing Global Null Hypothesis: BETA=0 | | | |
|---|---|---|---|
| Test | Chi-Square | DF | Pr > ChiSq |
| Likelihood Ratio | 89.6979 | 1 | <.0001 |
| Score | 88.2336 | 1 | <.0001 |
| Wald | 85.2847 | 1 | <.0001 |

#### The LOGISTIC Procedure

| Type 3 Analysis of Effects | | | |
|---|---|---|---|
| Effect | DF | Wald Chi-Square | Pr > ChiSq |
| firstdose | 1 | 85.2847 | <.0001 |

| Analysis of Maximum Likelihood Estimates | | | | | | | |
|---|---|---|---|---|---|---|---|
| Parameter | | DF | Estimate | Standard Error | Wald Chi-Square | Pr > ChiSq | Label |
| Intercept | | 1 | -8.0344 | 0.0561 | 20520.6300 | <.0001 | Intercept: COVIDDEATH1YRD1=1 |
| firstdose | 1. PFIZER | 1 | 0.6402 | 0.0693 | 85.2847 | <.0001 | firstdose 1. PFIZER |

| Odds Ratio Estimates | | | |
|---|---|---|---|
| Effect | Point Estimate | 95% Wald Confidence Limits | |
| firstdose 1. PFIZER vs 2. MODERN | 1.897 | 1.656 | 2.173 |

| Association of Predicted Probabilities and Observed Responses | | | |
|---|---|---|---|
| Percent Concordant | 32.7 | Somers' D | 0.155 |
| Percent Discordant | 17.3 | Gamma | 0.310 |
| Percent Tied | 50.0 | Tau-a | 0.000 |
| Pairs | 1806910821 | c | 0.577 |

### COVID Death adjusted (logistic regression)

#### The LOGISTIC Procedure

| Model Information | |
|---|---|
| **Data Set** | WORK.MATCHEDCOHORT |
| **Response Variable** | COVIDDEATH1YRD1 |
| **Number of Response Levels** | 2 |
| **Model** | binary logit |
| **Optimization Technique** | Fisher's scoring |

| | |
|---|---|
| **Number of Observations Read** | 1962822 |
| **Number of Observations Used** | 1962822 |

| Response Profile | | |
|---|---|---|
| **Ordered Value** | **COVIDDEATH1YRD1** | **Total Frequency** |
| **1** | 0 | 1961901 |
| **2** | 1 | 921 |

**Probability modeled is COVIDDEATH1YRD1=1.**

| Class Level Information | | | | | | | | | |
|---|---|---|---|---|---|---|---|---|---|
| **Class** | **Value** | **Design Variables** | | | | | | | |
| **agenew** | **18-39** | 0 | 0 | 0 | 0 | 0 | | | |
| | **40-49** | 1 | 0 | 0 | 0 | 0 | | | |
| | **50-59** | 0 | 1 | 0 | 0 | 0 | | | |
| | **60-69** | 0 | 0 | 1 | 0 | 0 | | | |
| | **70-79** | 0 | 0 | 0 | 1 | 0 | | | |
| | **80+** | 0 | 0 | 0 | 0 | 1 | | | |
| **firstdose** | **1. PFIZER** | 1 | | | | | | | |
| | **2. MODERN** | 0 | | | | | | | |
| **sex2** | **F** | 1 | | | | | | | |
| | **M** | 0 | | | | | | | |
| **RACE4** | **BLACK** | 1 | 0 | 0 | | | | | |
| | **OTHERS** | 0 | 1 | 0 | | | | | |
| | **UNKNOWN** | 0 | 0 | 1 | | | | | |
| | **WHITE** | 0 | 0 | 0 | | | | | |
| **ETHNICITY3** | **HISPANIC** | 1 | 0 | | | | | | |
| | **NOT HISPANIC** | 0 | 0 | | | | | | |
| | **UNKNOWN** | 0 | 1 | | | | | | |
| **Vaccinationsite** | **CHD OR DOH** | 1 | 0 | 0 | 0 | 0 | 0 | 0 | 0 |
| | **DOCTOR CLINIC** | 0 | 1 | 0 | 0 | 0 | 0 | 0 | 0 |

#### The LOGISTIC Procedure

| Class Level Information | | | | | | | | | |
|---|---|---|---|---|---|---|---|---|---|
| **Class** | **Value** | **Design Variables** | | | | | | | |
| | **FIRE RESQUE OR EMS** | 0 | 0 | 1 | 0 | 0 | 0 | 0 | 0 |
| | **FQHC OR RHC OR CHC** | 0 | 0 | 0 | 1 | 0 | 0 | 0 | 0 |
| | **HOSPITAL CLINIC** | 0 | 0 | 0 | 0 | 1 | 0 | 0 | 0 |
| | **HOSPITAL ER** | 0 | 0 | 0 | 0 | 0 | 1 | 0 | 0 |
| | **OTHER PHARMACIES** | 0 | 0 | 0 | 0 | 0 | 0 | 1 | 0 |
| | **OTHERS OR MISC** | 0 | 0 | 0 | 0 | 0 | 0 | 0 | 1 |
| | **PUBLIX OR CVS OR WALGREENS** | 0 | 0 | 0 | 0 | 0 | 0 | 0 | 0 |

| Model Convergence Status |
|---|
| Convergence criterion (GCONV=1E-8) satisfied. |

| Model Fit Statistics | | |
|---|---|---|
| **Criterion** | **Intercept Only** | **Intercept and Covariates** |
| **AIC** | 15961.455 | 14175.371 |
| **SC** | 15973.945 | 14475.128 |
| **-2 Log L** | 15959.455 | 14127.371 |

| Testing Global Null Hypothesis: BETA=0 | | | |
|---|---|---|---|
| **Test** | **Chi-Square** | **DF** | **Pr > ChiSq** |
| **Likelihood Ratio** | 1832.0841 | 23 | <.0001 |
| **Score** | 2670.4284 | 23 | <.0001 |
| **Wald** | 1548.9418 | 23 | <.0001 |

| Type 3 Analysis of Effects | | | |
|---|---|---|---|
| **Effect** | **DF** | **Wald Chi-Square** | **Pr > ChiSq** |
| **firstdose** | 1 | 85.3869 | <.0001 |
| **agenew** | 5 | 573.6126 | <.0001 |
| **sex2** | 1 | 38.8829 | <.0001 |
| **RACE4** | 3 | 12.5862 | 0.0056 |
| **ETHNICITY3** | 2 | 105.3860 | <.0001 |
| **decile_sviscore1** | 1 | 40.3162 | <.0001 |
| **decile_sviscore4** | 1 | 0.0648 | 0.7990 |
| **mortalityratepct** | 1 | 37.2033 | <.0001 |
| **Vaccinationsite** | 8 | 149.6582 | <.0001 |

#### The LOGISTIC Procedure

| Analysis of Maximum Likelihood Estimates | | | | | | | |
|---|---|---|---|---|---|---|---|
| **Parameter** | | **DF** | **Estimate** | **Standard Error** | **Wald Chi-Square** | **Pr > ChiSq** | **Label** |
| **Intercept** | | 1 | -11.2899 | 0.2936 | 1478.8827 | <.0001 | Intercept: COVIDDEATH1YRD1=1 |
| **firstdose** | **1. PFIZER** | 1 | 0.6411 | 0.0694 | 85.3869 | <.0001 | firstdose 1. PFIZER |
| **agenew** | **40-49** | 1 | 1.5307 | 0.3322 | 21.2329 | <.0001 | agenew 40-49 |
| **agenew** | **50-59** | 1 | 2.0112 | 0.3028 | 44.1061 | <.0001 | agenew 50-59 |
| **agenew** | **60-69** | 1 | 3.0519 | 0.2883 | 112.0460 | <.0001 | agenew 60-69 |
| **agenew** | **70-79** | 1 | 3.9924 | 0.2875 | 192.8018 | <.0001 | agenew 70-79 |
| **agenew** | **80+** | 1 | 4.7240 | 0.2995 | 248.8159 | <.0001 | agenew 80+ |
| **sex2** | **F** | 1 | -0.4226 | 0.0678 | 38.8829 | <.0001 | sex2 F |
| **RACE4** | **BLACK** | 1 | 0.3346 | 0.1405 | 5.6723 | 0.0172 | RACE4 BLACK |
| **RACE4** | **OTHERS** | 1 | -0.2866 | 0.1171 | 5.9857 | 0.0144 | RACE4 OTHERS |
| **RACE4** | **UNKNOWN** | 1 | 0.0155 | 0.1804 | 0.0073 | 0.9317 | RACE4 UNKNOWN |
| **ETHNICITY3** | **HISPANIC** | 1 | -0.1279 | 0.1120 | 1.3053 | 0.2532 | ETHNICITY3 HISPANIC |
| **ETHNICITY3** | **UNKNOWN** | 1 | 1.0096 | 0.1025 | 97.0883 | <.0001 | ETHNICITY3 UNKNOWN |
| **decile_sviscore1** | | 1 | 0.0936 | 0.0147 | 40.3162 | <.0001 | Rank for Variable SVISCORE1 |
| **decile_sviscore4** | | 1 | -0.00352 | 0.0138 | 0.0648 | 0.7990 | Rank for Variable SVISCORE4 |
| **mortalityratepct** | | 1 | 0.0592 | 0.00970 | 37.2033 | <.0001 | |
| **Vaccinationsite** | **CHD OR DOH** | 1 | -0.5598 | 0.1014 | 30.4703 | <.0001 | Vaccinationsite CHD OR DOH |
| **Vaccinationsite** | **DOCTOR CLINIC** | 1 | -0.0177 | 0.0889 | 0.0398 | 0.8419 | Vaccinationsite DOCTOR CLINIC |
| **Vaccinationsite** | **FIRE RESQUE OR EMS** | 1 | -0.6566 | 0.2042 | 10.3337 | 0.0013 | Vaccinationsite FIRE RESQUE OR EMS |
| **Vaccinationsite** | **FQHC OR RHC OR CHC** | 1 | -0.5727 | 0.3213 | 3.1758 | 0.0747 | Vaccinationsite FQHC OR RHC OR CHC |
| **Vaccinationsite** | **HOSPITAL CLINIC** | 1 | -0.1514 | 0.1397 | 1.1755 | 0.2783 | Vaccinationsite HOSPITAL CLINIC |
| **Vaccinationsite** | **HOSPITAL ER** | 1 | -10.6830 | 122.1 | 0.0077 | 0.9303 | Vaccinationsite HOSPITAL ER |
| **Vaccinationsite** | **OTHER PHARMACIES** | 1 | 0.5249 | 0.1694 | 9.6048 | 0.0019 | Vaccinationsite OTHER PHARMACIES |
| **Vaccinationsite** | **OTHERS OR MISC** | 1 | -1.7046 | 0.1698 | 100.7953 | <.0001 | Vaccinationsite OTHERS OR MISC |

| Odds Ratio Estimates | | | |
|---|---|---|---|
| **Effect** | **Point Estimate** | **95% Wald Confidence Limits** | |
| **firstdose 1. PFIZER vs 2. MODERN** | 1.899 | 1.657 | 2.175 |
| **agenew 40-49 vs 18-39** | 4.621 | 2.410 | 8.862 |
| **agenew 50-59 vs 18-39** | 7.473 | 4.128 | 13.528 |
| **agenew 60-69 vs 18-39** | 21.155 | 12.023 | 37.225 |
| **agenew 70-79 vs 18-39** | 54.185 | 30.841 | 95.196 |
| **agenew 80+ vs 18-39** | 112.620 | 62.617 | 202.552 |
| **sex2 F vs M** | 0.655 | 0.574 | 0.748 |

#### The LOGISTIC Procedure

| Odds Ratio Estimates | | | |
|---|---|---|---|
| Effect | Point Estimate | 95% Wald Confidence Limits | |
| RACE4 BLACK vs WHITE | 1.397 | 1.061 | 1.840 |
| RACE4 OTHERS vs WHITE | 0.751 | 0.597 | 0.945 |
| RACE4 UNKNOWN vs WHITE | 1.016 | 0.713 | 1.446 |
| ETHNICITY3 HISPANIC vs NOT HISPANIC | 0.880 | 0.706 | 1.096 |
| ETHNICITY3 UNKNOWN vs NOT HISPANIC | 2.745 | 2.245 | 3.355 |
| decile_sviscore1 | 1.098 | 1.067 | 1.130 |
| decile_sviscore4 | 0.996 | 0.970 | 1.024 |
| mortalityratepct | 1.061 | 1.041 | 1.081 |
| Vaccinationsite CHD OR DOH vs PUBLIX OR CVS OR WALGREENS | 0.571 | 0.468 | 0.697 |
| Vaccinationsite DOCTOR CLINIC vs PUBLIX OR CVS OR WALGREENS | 0.982 | 0.825 | 1.169 |
| Vaccinationsite FIRE RESQUE OR EMS vs PUBLIX OR CVS OR WALGREENS | 0.519 | 0.348 | 0.774 |
| Vaccinationsite FQHC OR RHC OR CHC vs PUBLIX OR CVS OR WALGREENS | 0.564 | 0.300 | 1.059 |
| Vaccinationsite HOSPITAL CLINIC vs PUBLIX OR CVS OR WALGREENS | 0.859 | 0.654 | 1.130 |
| Vaccinationsite HOSPITAL ER vs PUBLIX OR CVS OR WALGREENS | <0.001 | <0.001 | >999.999 |
| Vaccinationsite OTHER PHARMACIES vs PUBLIX OR CVS OR WALGREENS | 1.690 | 1.213 | 2.356 |
| Vaccinationsite OTHERS OR MISC vs PUBLIX OR CVS OR WALGREENS | 0.182 | 0.130 | 0.254 |

| Association of Predicted Probabilities and Observed Responses | | | |
|---|---|---|---|
| Percent Concordant | 84.8 | Somers' D | 0.697 |
| Percent Discordant | 15.1 | Gamma | 0.697 |
| Percent Tied | 0.0 | Tau-a | 0.001 |
| Pairs | 1806910821 | c | 0.849 |

### COVID deaths adjusted by Age

#### The LOGISTIC Procedure

**agenew=18-39 years**

| Model Information | |
|---|---|
| **Data Set** | WORK.MATCHEDCOHORT |
| **Response Variable** | COVIDDEATH1YRD1 |
| **Number of Response Levels** | 2 |
| **Model** | binary logit |
| **Optimization Technique** | Fisher's scoring |

| | |
|---|---|
| **Number of Observations Read** | 492484 |
| **Number of Observations Used** | 492484 |

| Response Profile | | |
|---|---|---|
| **Ordered Value** | **COVIDDEATH1YRD1** | **Total Frequency** |
| **1** | 0 | 492471 |
| **2** | 1 | 13 |

**Probability modeled is COVIDDEATH1YRD1=1.**

| Class Level Information | | | | | | | | | |
|---|---|---|---|---|---|---|---|---|---|
| **Class** | **Value** | **Design Variables** | | | | | | | |
| **agenew** | **18-39 years** | | | | | | | | |
| **firstdose** | **1. PFIZER** | 1 | | | | | | | |
| | **2. MODERN** | 0 | | | | | | | |
| **sex2** | **F** | 1 | | | | | | | |
| | **M** | 0 | | | | | | | |
| **RACE4** | **BLACK** | 1 | 0 | 0 | | | | | |
| | **OTHERS** | 0 | 1 | 0 | | | | | |
| | **UNKNOWN** | 0 | 0 | 1 | | | | | |
| | **WHITE** | 0 | 0 | 0 | | | | | |
| **ETHNICITY3** | **HISPANIC** | 1 | 0 | | | | | | |
| | **NOT HISPANIC** | 0 | 0 | | | | | | |
| | **UNKNOWN** | 0 | 1 | | | | | | |
| **Vaccinationsite** | **CHD OR DOH** | 1 | 0 | 0 | 0 | 0 | 0 | 0 | 0 |
| | **DOCTOR CLINIC** | 0 | 1 | 0 | 0 | 0 | 0 | 0 | 0 |
| | **FIRE RESQUE OR EMS** | 0 | 0 | 1 | 0 | 0 | 0 | 0 | 0 |
| | **FQHC OR RHC OR CHC** | 0 | 0 | 0 | 1 | 0 | 0 | 0 | 0 |
| | **HOSPITAL CLINIC** | 0 | 0 | 0 | 0 | 1 | 0 | 0 | 0 |
| | **HOSPITAL ER** | 0 | 0 | 0 | 0 | 0 | 1 | 0 | 0 |

#### The LOGISTIC Procedure

**agenew=18-39 years**

| Class Level Information | | | | | | | | | |
|---|---|---|---|---|---|---|---|---|---|
| Class | Value | Design Variables | | | | | | | |
| | OTHER PHARMACIES | 0 | 0 | 0 | 0 | 0 | 0 | 1 | 0 |
| | OTHERS OR MISC | 0 | 0 | 0 | 0 | 0 | 0 | 0 | 1 |
| | PUBLIX OR CVS OR WALGREENS | 0 | 0 | 0 | 0 | 0 | 0 | 0 | 0 |

| Model Convergence Status |
|---|
| Quasi-complete separation of data points detected. |

**Warning:** The maximum likelihood estimate may not exist.

**Warning:** The LOGISTIC procedure continues in spite of the above warning. Results shown are based on the last maximum likelihood iteration. Validity of the model fit is questionable.

| Model Fit Statistics | | |
|---|---|---|
| Criterion | Intercept Only | Intercept and Covariates |
| AIC | 302.099 | 303.220 |
| SC | 313.206 | 514.257 |
| -2 Log L | 300.099 | 265.220 |

| Testing Global Null Hypothesis: BETA=0 | | | |
|---|---|---|---|
| Test | Chi-Square | DF | Pr > ChiSq |
| Likelihood Ratio | 34.8783 | 18 | 0.0098 |
| Score | 41.3398 | 18 | 0.0014 |
| Wald | 25.2591 | 18 | 0.1179 |

| Type 3 Analysis of Effects | | | |
|---|---|---|---|
| Effect | DF | Wald Chi-Square | Pr > ChiSq |
| firstdose | 1 | 0.6776 | 0.4104 |
| agenew | 0 | . | . |
| sex2 | 1 | 0.1110 | 0.7390 |
| RACE4 | 3 | 0.4189 | 0.9363 |
| ETHNICITY3 | 2 | 2.0374 | 0.3611 |
| decile_sviscore1 | 1 | 7.1833 | 0.0074 |
| decile_sviscore4 | 1 | 0.1910 | 0.6621 |
| mortalityratepct | 1 | 0.0001 | 0.9939 |
| Vaccinationsite | 8 | 11.9276 | 0.1545 |

#### The LOGISTIC Procedure

**Warning:** The validity of the model fit is questionable.

**agenew=18-39 years**

| Analysis of Maximum Likelihood Estimates | | | | | | | |
|---|---|---|---|---|---|---|---|
| Parameter | | DF | Estimate | Standard Error | Wald Chi-Square | Pr > ChiSq | Label |
| Intercept | | 1 | -13.4179 | 1.1506 | 135.9837 | <.0001 | Intercept: COVIDDEATH1YRD1=1 |
| firstdose | 1. PFIZER | 1 | 0.4693 | 0.5701 | 0.6776 | 0.4104 | firstdose 1. PFIZER |
| sex2 | F | 1 | 0.1908 | 0.5727 | 0.1110 | 0.7390 | sex2 F |
| RACE4 | BLACK | 1 | 0.1637 | 0.7368 | 0.0494 | 0.8242 | RACE4 BLACK |
| RACE4 | OTHERS | 1 | -0.6100 | 1.0589 | 0.3318 | 0.5646 | RACE4 OTHERS |
| RACE4 | UNKNOWN | 1 | -10.6824 | 324.2 | 0.0011 | 0.9737 | RACE4 UNKNOWN |
| ETHNICITY3 | HISPANIC | 1 | -0.9881 | 0.7278 | 1.8430 | 0.1746 | ETHNICITY3 HISPANIC |
| ETHNICITY3 | UNKNOWN | 1 | -0.7764 | 1.0839 | 0.5131 | 0.4738 | ETHNICITY3 UNKNOWN |
| decile_sviscore1 | | 1 | 0.4196 | 0.1566 | 7.1833 | 0.0074 | Rank for Variable SVISCORE1 |
| decile_sviscore4 | | 1 | -0.0523 | 0.1198 | 0.1910 | 0.6621 | Rank for Variable SVISCORE4 |
| mortalityratepct | | 1 | -10.6657 | 1396.5 | 0.0001 | 0.9939 | |
| Vaccinationsite | CHD OR DOH | 1 | -10.8708 | 429.5 | 0.0006 | 0.9798 | Vaccinationsite CHD OR DOH |
| Vaccinationsite | DOCTOR CLINIC | 1 | 2.1755 | 0.6510 | 11.1678 | 0.0008 | Vaccinationsite DOCTOR CLINIC |
| Vaccinationsite | FIRE RESQUE OR EMS | 1 | -10.6600 | 2755.4 | 0.0000 | 0.9969 | Vaccinationsite FIRE RESQUE OR EMS |
| Vaccinationsite | FQHC OR RHC OR CHC | 1 | -10.9144 | 772.5 | 0.0002 | 0.9887 | Vaccinationsite FQHC OR RHC OR CHC |
| Vaccinationsite | HOSPITAL CLINIC | 1 | -10.5251 | 471.5 | 0.0005 | 0.9822 | Vaccinationsite HOSPITAL CLINIC |
| Vaccinationsite | HOSPITAL ER | 1 | -10.7713 | 1887.9 | 0.0000 | 0.9954 | Vaccinationsite HOSPITAL ER |
| Vaccinationsite | OTHER PHARMACIES | 1 | 1.8905 | 0.7756 | 5.9415 | 0.0148 | Vaccinationsite OTHER PHARMACIES |
| Vaccinationsite | OTHERS OR MISC | 1 | -10.3679 | 361.5 | 0.0008 | 0.9771 | Vaccinationsite OTHERS OR MISC |

| Odds Ratio Estimates | | | |
|---|---|---|---|
| Effect | Point Estimate | 95% Wald Confidence Limits | |
| firstdose 1. PFIZER vs 2. MODERN | 1.599 | 0.523 | 4.888 |
| sex2 F vs M | 1.210 | 0.394 | 3.719 |
| RACE4 BLACK vs WHITE | 1.178 | 0.278 | 4.992 |
| RACE4 OTHERS vs WHITE | 0.543 | 0.068 | 4.329 |
| RACE4 UNKNOWN vs WHITE | <0.001 | <0.001 | >999.999 |
| ETHNICITY3 HISPANIC vs NOT HISPANIC | 0.372 | 0.089 | 1.550 |
| ETHNICITY3 UNKNOWN vs NOT HISPANIC | 0.460 | 0.055 | 3.850 |
| decile_sviscore1 | 1.521 | 1.119 | 2.068 |
| decile_sviscore4 | 0.949 | 0.750 | 1.200 |
| mortalityratepct | <0.001 | <0.001 | >999.999 |
| Vaccinationsite CHD OR DOH vs PUBLIX OR CVS OR WALGREENS | <0.001 | <0.001 | >999.999 |

#### The LOGISTIC Procedure

**Warning:** The validity of the model fit is questionable.

**agenew=18-39 years**

| Odds Ratio Estimates | | | |
|---|---|---|---|
| Effect | Point Estimate | 95% Wald Confidence Limits | |
| Vaccinationsite DOCTOR CLINIC vs PUBLIX OR CVS OR WALGREENS | 8.806 | 2.459 | 31.544 |
| Vaccinationsite FIRE RESQUE OR EMS vs PUBLIX OR CVS OR WALGREENS | <0.001 | <0.001 | >999.999 |
| Vaccinationsite FQHC OR RHC OR CHC vs PUBLIX OR CVS OR WALGREENS | <0.001 | <0.001 | >999.999 |
| Vaccinationsite HOSPITAL CLINIC vs PUBLIX OR CVS OR WALGREENS | <0.001 | <0.001 | >999.999 |
| Vaccinationsite HOSPITAL ER vs PUBLIX OR CVS OR WALGREENS | <0.001 | <0.001 | >999.999 |
| Vaccinationsite OTHER PHARMACIES vs PUBLIX OR CVS OR WALGREENS | 6.622 | 1.448 | 30.282 |
| Vaccinationsite OTHERS OR MISC vs PUBLIX OR CVS OR WALGREENS | <0.001 | <0.001 | >999.999 |

| Association of Predicted Probabilities and Observed Responses | | | |
|---|---|---|---|
| Percent Concordant | 86.5 | Somers' D | 0.730 |
| Percent Discordant | 13.5 | Gamma | 0.730 |
| Percent Tied | 0.0 | Tau-a | 0.000 |
| Pairs | 6402123 | c | 0.865 |

### COVID deaths adjusted by Age

#### The LOGISTIC Procedure

##### agenew=40-59 years

| Model Information | |
|---|---|
| **Data Set** | WORK.MATCHEDCOHORT |
| **Response Variable** | COVIDDEATH1YRD1 |
| **Number of Response Levels** | 2 |
| **Model** | binary logit |
| **Optimization Technique** | Fisher's scoring |

| | |
|---|---|
| **Number of Observations Read** | 634102 |
| **Number of Observations Used** | 634102 |

| Response Profile | | |
|---|---|---|
| **Ordered Value** | **COVIDDEATH1YRD1** | **Total Frequency** |
| **1** | 0 | 634003 |
| **2** | 1 | 99 |

**Probability modeled is COVIDDEATH1YRD1=1.**

| Class Level Information | | | | | | | | | |
|---|---|---|---|---|---|---|---|---|---|
| **Class** | **Value** | **Design Variables** | | | | | | | |
| **agenew** | **40-59 years** | | | | | | | | |
| **firstdose** | **1. PFIZER** | 1 | | | | | | | |
| | **2. MODERN** | 0 | | | | | | | |
| **sex2** | **F** | 1 | | | | | | | |
| | **M** | 0 | | | | | | | |
| **RACE4** | **BLACK** | 1 | 0 | 0 | | | | | |
| | **OTHERS** | 0 | 1 | 0 | | | | | |
| | **UNKNOWN** | 0 | 0 | 1 | | | | | |
| | **WHITE** | 0 | 0 | 0 | | | | | |
| **ETHNICITY3** | **HISPANIC** | 1 | 0 | | | | | | |
| | **NOT HISPANIC** | 0 | 0 | | | | | | |
| | **UNKNOWN** | 0 | 1 | | | | | | |
| **Vaccinationsite** | **CHD OR DOH** | 1 | 0 | 0 | 0 | 0 | 0 | 0 | 0 |
| | **DOCTOR CLINIC** | 0 | 1 | 0 | 0 | 0 | 0 | 0 | 0 |
| | **FIRE RESQUE OR EMS** | 0 | 0 | 1 | 0 | 0 | 0 | 0 | 0 |
| | **FQHC OR RHC OR CHC** | 0 | 0 | 0 | 1 | 0 | 0 | 0 | 0 |
| | **HOSPITAL CLINIC** | 0 | 0 | 0 | 0 | 1 | 0 | 0 | 0 |
| | **HOSPITAL ER** | 0 | 0 | 0 | 0 | 0 | 1 | 0 | 0 |

### COVID deaths adjusted by Age

#### The LOGISTIC Procedure

##### agenew=40-59 years

| Class Level Information | | | | | | | | | |
|---|---|---|---|---|---|---|---|---|---|
| Class | Value | Design Variables | | | | | | | |
| | OTHER PHARMACIES | 0 | 0 | 0 | 0 | 0 | 0 | 1 | 0 |
| | OTHERS OR MISC | 0 | 0 | 0 | 0 | 0 | 0 | 0 | 1 |
| | PUBLIX OR CVS OR WALGREENS | 0 | 0 | 0 | 0 | 0 | 0 | 0 | 0 |

| Model Convergence Status |
|---|
| Convergence criterion (GCONV=1E-8) satisfied. |

| Model Fit Statistics | | |
|---|---|---|
| Criterion | Intercept Only | Intercept and Covariates |
| AIC | 1935.424 | 1903.994 |
| SC | 1946.784 | 2119.833 |
| -2 Log L | 1933.424 | 1865.994 |

| Testing Global Null Hypothesis: BETA=0 | | | |
|---|---|---|---|
| Test | Chi-Square | DF | Pr > ChiSq |
| Likelihood Ratio | 67.4300 | 18 | <.0001 |
| Score | 72.8124 | 18 | <.0001 |
| Wald | 67.6293 | 18 | <.0001 |

| Type 3 Analysis of Effects | | | |
|---|---|---|---|
| Effect | DF | Wald Chi-Square | Pr > ChiSq |
| firstdose | 1 | 0.8221 | 0.3646 |
| agenew | 0 | . | . |
| sex2 | 1 | 0.8419 | 0.3589 |
| RACE4 | 3 | 5.4271 | 0.1431 |
| ETHNICITY3 | 2 | 10.2523 | 0.0059 |
| decile_sviscore1 | 1 | 6.5478 | 0.0105 |
| decile_sviscore4 | 1 | 2.6850 | 0.1013 |
| mortalityratepct | 1 | 5.7478 | 0.0165 |
| Vaccinationsite | 8 | 15.4556 | 0.0509 |

#### The LOGISTIC Procedure

**agenew=40-59 years**

| Analysis of Maximum Likelihood Estimates | | | | | | | |
|---|---|---|---|---|---|---|---|
| **Parameter** | | **DF** | **Estimate** | **Standard Error** | **Wald Chi-Square** | **Pr > ChiSq** | **Label** |
| **Intercept** | | 1 | -10.1238 | 0.3401 | 886.1324 | <.0001 | Intercept: COVIDDEATH1YRD1=1 |
| **firstdose** | **1. PFIZER** | 1 | 0.1830 | 0.2019 | 0.8221 | 0.3646 | firstdose 1. PFIZER |
| **sex2** | **F** | 1 | -0.1957 | 0.2133 | 0.8419 | 0.3589 | sex2 F |
| **RACE4** | **BLACK** | 1 | 0.5814 | 0.2960 | 3.8578 | 0.0495 | RACE4 BLACK |
| **RACE4** | **OTHERS** | 1 | -0.2640 | 0.3504 | 0.5678 | 0.4511 | RACE4 OTHERS |
| **RACE4** | **UNKNOWN** | 1 | -0.3943 | 0.5650 | 0.4870 | 0.4853 | RACE4 UNKNOWN |
| **ETHNICITY3** | **HISPANIC** | 1 | 0.0879 | 0.2850 | 0.0950 | 0.7579 | ETHNICITY3 HISPANIC |
| **ETHNICITY3** | **UNKNOWN** | 1 | 0.9381 | 0.3000 | 9.7812 | 0.0018 | ETHNICITY3 UNKNOWN |
| **decile_sviscore1** | | 1 | 0.1204 | 0.0470 | 6.5478 | 0.0105 | Rank for Variable SVISCORE1 |
| **decile_sviscore4** | | 1 | 0.0725 | 0.0442 | 2.6850 | 0.1013 | Rank for Variable SVISCORE4 |
| **mortalityratepct** | | 1 | 0.5050 | 0.2106 | 5.7478 | 0.0165 | |
| **Vaccinationsite** | **CHD OR DOH** | 1 | 0.2226 | 0.5216 | 0.1821 | 0.6695 | Vaccinationsite CHD OR DOH |
| **Vaccinationsite** | **DOCTOR CLINIC** | 1 | -0.2563 | 0.2967 | 0.7462 | 0.3877 | Vaccinationsite DOCTOR CLINIC |
| **Vaccinationsite** | **FIRE RESQUE OR EMS** | 1 | -11.6370 | 674.6 | 0.0003 | 0.9862 | Vaccinationsite FIRE RESQUE OR EMS |
| **Vaccinationsite** | **FQHC OR RHC OR CHC** | 1 | 0.3699 | 0.6039 | 0.3750 | 0.5403 | Vaccinationsite FQHC OR RHC OR CHC |
| **Vaccinationsite** | **HOSPITAL CLINIC** | 1 | 0.8399 | 0.3636 | 5.3352 | 0.0209 | Vaccinationsite HOSPITAL CLINIC |
| **Vaccinationsite** | **HOSPITAL ER** | 1 | -11.3876 | 607.5 | 0.0004 | 0.9850 | Vaccinationsite HOSPITAL ER |
| **Vaccinationsite** | **OTHER PHARMACIES** | 1 | 0.6334 | 0.3386 | 3.5004 | 0.0614 | Vaccinationsite OTHER PHARMACIES |
| **Vaccinationsite** | **OTHERS OR MISC** | 1 | -1.5020 | 0.7250 | 4.2921 | 0.0383 | Vaccinationsite OTHERS OR MISC |

| Odds Ratio Estimates | | | |
|---|---|---|---|
| **Effect** | **Point Estimate** | **95% Wald Confidence Limits** | |
| **firstdose 1. PFIZER vs 2. MODERN** | 1.201 | 0.808 | 1.784 |
| **sex2 F vs M** | 0.822 | 0.541 | 1.249 |
| **RACE4 BLACK vs WHITE** | 1.789 | 1.001 | 3.195 |
| **RACE4 OTHERS vs WHITE** | 0.768 | 0.386 | 1.526 |
| **RACE4 UNKNOWN vs WHITE** | 0.674 | 0.223 | 2.040 |
| **ETHNICITY3 HISPANIC vs NOT HISPANIC** | 1.092 | 0.625 | 1.909 |
| **ETHNICITY3 UNKNOWN vs NOT HISPANIC** | 2.555 | 1.419 | 4.600 |
| **decile_sviscore1** | 1.128 | 1.029 | 1.237 |
| **decile_sviscore4** | 1.075 | 0.986 | 1.173 |
| **mortalityratepct** | 1.657 | 1.097 | 2.504 |
| **Vaccinationsite CHD OR DOH vs PUBLIX OR CVS OR WALGREENS** | 1.249 | 0.449 | 3.473 |

#### The LOGISTIC Procedure

**agenew=40-59 years**

| Odds Ratio Estimates | | | |
|---|---|---|---|
| Effect | Point Estimate | 95% Wald Confidence Limits | |
| Vaccinationsite DOCTOR CLINIC vs PUBLIX OR CVS OR WALGREENS | 0.774 | 0.433 | 1.384 |
| Vaccinationsite FIRE RESQUE OR EMS vs PUBLIX OR CVS OR WALGREENS | <0.001 | <0.001 | >999.999 |
| Vaccinationsite FQHC OR RHC OR CHC vs PUBLIX OR CVS OR WALGREENS | 1.448 | 0.443 | 4.728 |
| Vaccinationsite HOSPITAL CLINIC vs PUBLIX OR CVS OR WALGREENS | 2.316 | 1.136 | 4.724 |
| Vaccinationsite HOSPITAL ER vs PUBLIX OR CVS OR WALGREENS | <0.001 | <0.001 | >999.999 |
| Vaccinationsite OTHER PHARMACIES vs PUBLIX OR CVS OR WALGREENS | 1.884 | 0.970 | 3.658 |
| Vaccinationsite OTHERS OR MISC vs PUBLIX OR CVS OR WALGREENS | 0.223 | 0.054 | 0.922 |

| Association of Predicted Probabilities and Observed Responses | | | |
|---|---|---|---|
| Percent Concordant | 72.8 | Somers' D | 0.456 |
| Percent Discordant | 27.2 | Gamma | 0.456 |
| Percent Tied | 0.0 | Tau-a | 0.000 |
| Pairs | 62766297 | c | 0.728 |

### The LOGISTIC Procedure

**agenew=60+ years**

| Model Information | |
|---|---|
| Data Set | WORK.MATCHEDCOHORT |
| Response Variable | COVIDDEATH1YRD1 |
| Number of Response Levels | 2 |
| Model | binary logit |
| Optimization Technique | Fisher's scoring |

| | |
|---|---|
| Number of Observations Read | 836236 |
| Number of Observations Used | 836236 |

| Response Profile | | |
|---|---|---|
| Ordered Value | COVIDDEATH1YRD1 | Total Frequency |
| 1 | 0 | 835427 |
| 2 | 1 | 809 |

**Probability modeled is COVIDDEATH1YRD1=1.**

| Class Level Information | | | | | | | | | |
|---|---|---|---|---|---|---|---|---|---|
| Class | Value | Design Variables | | | | | | | |
| agenew | 60+ years | | | | | | | | |
| firstdose | 1. PFIZER | 1 | | | | | | | |
| | 2. MODERN | 0 | | | | | | | |
| sex2 | F | 1 | | | | | | | |
| | M | 0 | | | | | | | |
| RACE4 | BLACK | 1 | 0 | 0 | | | | | |
| | OTHERS | 0 | 1 | 0 | | | | | |
| | UNKNOWN | 0 | 0 | 1 | | | | | |
| | WHITE | 0 | 0 | 0 | | | | | |
| ETHNICITY3 | HISPANIC | 1 | 0 | | | | | | |
| | NOT HISPANIC | 0 | 0 | | | | | | |
| | UNKNOWN | 0 | 1 | | | | | | |
| Vaccinationsite | CHD OR DOH | 1 | 0 | 0 | 0 | 0 | 0 | 0 | 0 |
| | DOCTOR CLINIC | 0 | 1 | 0 | 0 | 0 | 0 | 0 | 0 |
| | FIRE RESQUE OR EMS | 0 | 0 | 1 | 0 | 0 | 0 | 0 | 0 |
| | FQHC OR RHC OR CHC | 0 | 0 | 0 | 1 | 0 | 0 | 0 | 0 |
| | HOSPITAL CLINIC | 0 | 0 | 0 | 0 | 1 | 0 | 0 | 0 |
| | HOSPITAL ER | 0 | 0 | 0 | 0 | 0 | 1 | 0 | 0 |

#### The LOGISTIC Procedure

**agenew=60+ years**

| Class Level Information | | | | | | | | | |
|---|---|---|---|---|---|---|---|---|---|
| Class | Value | Design Variables | | | | | | | |
| | OTHER PHARMACIES | 0 | 0 | 0 | 0 | 0 | 0 | 1 | 0 |
| | OTHERS OR MISC | 0 | 0 | 0 | 0 | 0 | 0 | 0 | 1 |
| | PUBLIX OR CVS OR WALGREENS | 0 | 0 | 0 | 0 | 0 | 0 | 0 | 0 |

| Model Convergence Status |
|---|
| Convergence criterion (GCONV=1E-8) satisfied. |

| Model Fit Statistics | | |
|---|---|---|
| Criterion | Intercept Only | Intercept and Covariates |
| AIC | 12849.540 | 12188.589 |
| SC | 12861.177 | 12409.685 |
| -2 Log L | 12847.540 | 12150.589 |

| Testing Global Null Hypothesis: BETA=0 | | | |
|---|---|---|---|
| Test | Chi-Square | DF | Pr > ChiSq |
| Likelihood Ratio | 696.9516 | 18 | <.0001 |
| Score | 1077.7590 | 18 | <.0001 |
| Wald | 954.4806 | 18 | <.0001 |

| Type 3 Analysis of Effects | | | |
|---|---|---|---|
| Effect | DF | Wald Chi-Square | Pr > ChiSq |
| firstdose | 1 | 88.9220 | <.0001 |
| agenew | 0 | . | . |
| sex2 | 1 | 20.1160 | <.0001 |
| RACE4 | 3 | 13.5834 | 0.0035 |
| ETHNICITY3 | 2 | 117.8493 | <.0001 |
| decile_sviscore1 | 1 | 21.3682 | <.0001 |
| decile_sviscore4 | 1 | 0.2937 | 0.5879 |
| mortalityratepct | 1 | 537.0439 | <.0001 |
| Vaccinationsite | 8 | 107.9892 | <.0001 |

#### The LOGISTIC Procedure

**agenew=60+ years**

| Analysis of Maximum Likelihood Estimates | | | | | | | |
|---|---|---|---|---|---|---|---|
| Parameter | | DF | Estimate | Standard Error | Wald Chi-Square | Pr > ChiSq | Label |
| Intercept | | 1 | -7.6800 | 0.1085 | 5011.5460 | <.0001 | Intercept: COVIDDEATH1YRD1=1 |
| firstdose | 1. PFIZER | 1 | 0.7063 | 0.0749 | 88.9220 | <.0001 | firstdose 1. PFIZER |
| sex2 | F | 1 | -0.3229 | 0.0720 | 20.1160 | <.0001 | sex2 F |
| RACE4 | BLACK | 1 | 0.0352 | 0.1657 | 0.0450 | 0.8320 | RACE4 BLACK |
| RACE4 | OTHERS | 1 | -0.4865 | 0.1358 | 12.8297 | 0.0003 | RACE4 OTHERS |
| RACE4 | UNKNOWN | 1 | 0.0960 | 0.1910 | 0.2527 | 0.6152 | RACE4 UNKNOWN |
| ETHNICITY3 | HISPANIC | 1 | -0.3248 | 0.1333 | 5.9335 | 0.0149 | ETHNICITY3 HISPANIC |
| ETHNICITY3 | UNKNOWN | 1 | 1.1062 | 0.1093 | 102.3501 | <.0001 | ETHNICITY3 UNKNOWN |
| decile_sviscore1 | | 1 | 0.0724 | 0.0157 | 21.3682 | <.0001 | Rank for Variable SVISCORE1 |
| decile_sviscore4 | | 1 | -0.00802 | 0.0148 | 0.2937 | 0.5879 | Rank for Variable SVISCORE4 |
| mortalityratepct | | 1 | 0.1262 | 0.00545 | 537.0439 | <.0001 | |
| Vaccinationsite | CHD OR DOH | 1 | -0.3999 | 0.1024 | 15.2459 | <.0001 | Vaccinationsite CHD OR DOH |
| Vaccinationsite | DOCTOR CLINIC | 1 | 0.0839 | 0.0941 | 0.7950 | 0.3726 | Vaccinationsite DOCTOR CLINIC |
| Vaccinationsite | FIRE RESQUE OR EMS | 1 | -0.3119 | 0.2045 | 2.3272 | 0.1271 | Vaccinationsite FIRE RESQUE OR EMS |
| Vaccinationsite | FQHC OR RHC OR CHC | 1 | -0.6625 | 0.3827 | 2.9974 | 0.0834 | Vaccinationsite FQHC OR RHC OR CHC |
| Vaccinationsite | HOSPITAL CLINIC | 1 | -0.2104 | 0.1528 | 1.8949 | 0.1686 | Vaccinationsite HOSPITAL CLINIC |
| Vaccinationsite | HOSPITAL ER | 1 | -11.4614 | 192.3 | 0.0036 | 0.9525 | Vaccinationsite HOSPITAL ER |
| Vaccinationsite | OTHER PHARMACIES | 1 | 0.3265 | 0.2065 | 2.4993 | 0.1139 | Vaccinationsite OTHER PHARMACIES |
| Vaccinationsite | OTHERS OR MISC | 1 | -1.5645 | 0.1744 | 80.4856 | <.0001 | Vaccinationsite OTHERS OR MISC |

| Odds Ratio Estimates | | | |
|---|---|---|---|
| Effect | Point Estimate | 95% Wald Confidence Limits | |
| firstdose 1. PFIZER vs 2. MODERN | 2.027 | 1.750 | 2.347 |
| sex2 F vs M | 0.724 | 0.629 | 0.834 |
| RACE4 BLACK vs WHITE | 1.036 | 0.749 | 1.433 |
| RACE4 OTHERS vs WHITE | 0.615 | 0.471 | 0.802 |
| RACE4 UNKNOWN vs WHITE | 1.101 | 0.757 | 1.601 |
| ETHNICITY3 HISPANIC vs NOT HISPANIC | 0.723 | 0.556 | 0.939 |
| ETHNICITY3 UNKNOWN vs NOT HISPANIC | 3.023 | 2.440 | 3.745 |
| decile_sviscore1 | 1.075 | 1.043 | 1.109 |
| decile_sviscore4 | 0.992 | 0.964 | 1.021 |
| mortalityratepct | 1.135 | 1.123 | 1.147 |
| Vaccinationsite CHD OR DOH vs PUBLIX OR CVS OR WALGREENS | 0.670 | 0.548 | 0.819 |

#### The LOGISTIC Procedure

**agenew=60+ years**

| Odds Ratio Estimates | | | |
|---|---|---|---|
| Effect | Point Estimate | 95% Wald Confidence Limits | |
| Vaccinationsite DOCTOR CLINIC vs PUBLIX OR CVS OR WALGREENS | 1.087 | 0.904 | 1.308 |
| Vaccinationsite FIRE RESQUE OR EMS vs PUBLIX OR CVS OR WALGREENS | 0.732 | 0.490 | 1.093 |
| Vaccinationsite FQHC OR RHC OR CHC vs PUBLIX OR CVS OR WALGREENS | 0.516 | 0.244 | 1.091 |
| Vaccinationsite HOSPITAL CLINIC vs PUBLIX OR CVS OR WALGREENS | 0.810 | 0.601 | 1.093 |
| Vaccinationsite HOSPITAL ER vs PUBLIX OR CVS OR WALGREENS | <0.001 | <0.001 | >999.999 |
| Vaccinationsite OTHER PHARMACIES vs PUBLIX OR CVS OR WALGREENS | 1.386 | 0.925 | 2.078 |
| Vaccinationsite OTHERS OR MISC vs PUBLIX OR CVS OR WALGREENS | 0.209 | 0.149 | 0.294 |

| Association of Predicted Probabilities and Observed Responses | | | |
|---|---|---|---|
| Percent Concordant | 74.4 | Somers' D | 0.489 |
| Percent Discordant | 25.6 | Gamma | 0.489 |
| Percent Tied | 0.0 | Tau-a | 0.001 |
| Pairs | 675860443 | c | 0.744 |

#### The LOGISTIC Procedure

| Model Information | |
|---|---|
| Data Set | WORK.MATCHEDCOHORT |
| Response Variable | NONCOVIDDEATH1YRD1 |
| Number of Response Levels | 2 |
| Model | binary logit |
| Optimization Technique | Fisher's scoring |

| | |
|---|---|
| Number of Observations Read | 1962822 |
| Number of Observations Used | 1962822 |

| Response Profile | | |
|---|---|---|
| Ordered Value | NONCOVIDDEATH1YRD1 | Total Frequency |
| 1 | 0 | 1950534 |
| 2 | 1 | 12288 |

**Probability modeled is NONCOVIDDEATH1YRD1=1.**

| Class Level Information | | | | | | | | | |
|---|---|---|---|---|---|---|---|---|---|
| Class | Value | Design Variables | | | | | | | |
| agenew | 18-39 | 0 | 0 | 0 | 0 | 0 | | | |
| | 40-49 | 1 | 0 | 0 | 0 | 0 | | | |
| | 50-59 | 0 | 1 | 0 | 0 | 0 | | | |
| | 60-69 | 0 | 0 | 1 | 0 | 0 | | | |
| | 70-79 | 0 | 0 | 0 | 1 | 0 | | | |
| | 80+ | 0 | 0 | 0 | 0 | 1 | | | |
| firstdose | 1. PFIZER | 1 | | | | | | | |
| | 2. MODERN | 0 | | | | | | | |
| sex2 | F | 1 | | | | | | | |
| | M | 0 | | | | | | | |
| RACE4 | BLACK | 1 | 0 | 0 | | | | | |
| | OTHERS | 0 | 1 | 0 | | | | | |
| | UNKNOWN | 0 | 0 | 1 | | | | | |
| | WHITE | 0 | 0 | 0 | | | | | |
| ETHNICITY3 | HISPANIC | 1 | 0 | | | | | | |
| | NOT HISPANIC | 0 | 0 | | | | | | |
| | UNKNOWN | 0 | 1 | | | | | | |
| Vaccinationsite | CHD OR DOH | 1 | 0 | 0 | 0 | 0 | 0 | 0 | 0 |
| | DOCTOR CLINIC | 0 | 1 | 0 | 0 | 0 | 0 | 0 | 0 |

#### The LOGISTIC Procedure

| Class Level Information | | | | | | | | | |
|---|---|---|---|---|---|---|---|---|---|
| **Class** | **Value** | **Design Variables** | | | | | | | |
| | **FIRE RESQUE OR EMS** | 0 | 0 | 1 | 0 | 0 | 0 | 0 | 0 |
| | **FQHC OR RHC OR CHC** | 0 | 0 | 0 | 1 | 0 | 0 | 0 | 0 |
| | **HOSPITAL CLINIC** | 0 | 0 | 0 | 0 | 1 | 0 | 0 | 0 |
| | **HOSPITAL ER** | 0 | 0 | 0 | 0 | 0 | 1 | 0 | 0 |
| | **OTHER PHARMACIES** | 0 | 0 | 0 | 0 | 0 | 0 | 1 | 0 |
| | **OTHERS OR MISC** | 0 | 0 | 0 | 0 | 0 | 0 | 0 | 1 |
| | **PUBLIX OR CVS OR WALGREENS** | 0 | 0 | 0 | 0 | 0 | 0 | 0 | 0 |

| Model Convergence Status |
|---|
| Convergence criterion (GCONV=1E-8) satisfied. |

| Model Fit Statistics | | |
|---|---|---|
| **Criterion** | **Intercept Only** | **Intercept and Covariates** |
| **AIC** | 149187.62 | 127780.32 |
| **SC** | 149200.11 | 128080.08 |
| **-2 Log L** | 149185.62 | 127732.32 |

| Testing Global Null Hypothesis: BETA=0 | | | |
|---|---|---|---|
| **Test** | **Chi-Square** | **DF** | **Pr > ChiSq** |
| **Likelihood Ratio** | 21453.3041 | 23 | <.0001 |
| **Score** | 32555.0907 | 23 | <.0001 |
| **Wald** | 18243.1772 | 23 | <.0001 |

| Type 3 Analysis of Effects | | | |
|---|---|---|---|
| **Effect** | **DF** | **Wald Chi-Square** | **Pr > ChiSq** |
| **firstdose** | 1 | 294.3961 | <.0001 |
| **agenew** | 5 | 5793.8478 | <.0001 |
| **sex2** | 1 | 320.8286 | <.0001 |
| **RACE4** | 3 | 191.6315 | <.0001 |
| **ETHNICITY3** | 2 | 1459.7989 | <.0001 |
| **decile_sviscore1** | 1 | 164.6786 | <.0001 |
| **decile_sviscore4** | 1 | 6.6767 | 0.0098 |
| **mortalityratepct** | 1 | 643.3779 | <.0001 |
| **Vaccinationsite** | 8 | 1481.5742 | <.0001 |

#### The LOGISTIC Procedure

| Analysis of Maximum Likelihood Estimates | | | | | | | |
|---|---|---|---|---|---|---|---|
| Parameter | | DF | Estimate | Standard Error | Wald Chi-Square | Pr > ChiSq | Label |
| **Intercept** | | 1 | -7.7933 | 0.0653 | 14236.0323 | <.0001 | Intercept: NONCOVIDDEATH1YRD1=1 |
| **firstdose** | **1. PFIZER** | 1 | 0.3176 | 0.0185 | 294.3961 | <.0001 | firstdose 1. PFIZER |
| **agenew** | **40-49** | 1 | 0.8865 | 0.0812 | 119.2811 | <.0001 | agenew 40-49 |
| **agenew** | **50-59** | 1 | 1.6055 | 0.0679 | 559.0582 | <.0001 | agenew 50-59 |
| **agenew** | **60-69** | 1 | 2.4402 | 0.0640 | 1452.6033 | <.0001 | agenew 60-69 |
| **agenew** | **70-79** | 1 | 3.2421 | 0.0640 | 2568.7062 | <.0001 | agenew 70-79 |
| **agenew** | **80+** | 1 | 3.9074 | 0.0693 | 3181.6847 | <.0001 | agenew 80+ |
| **sex2** | **F** | 1 | -0.3358 | 0.0187 | 320.8286 | <.0001 | sex2 F |
| **RACE4** | **BLACK** | 1 | 0.0337 | 0.0436 | 0.5981 | 0.4393 | RACE4 BLACK |
| **RACE4** | **OTHERS** | 1 | -0.4839 | 0.0352 | 189.1327 | <.0001 | RACE4 OTHERS |
| **RACE4** | **UNKNOWN** | 1 | -0.0441 | 0.0529 | 0.6953 | 0.4044 | RACE4 UNKNOWN |
| **ETHNICITY3** | **HISPANIC** | 1 | -0.5373 | 0.0353 | 232.1126 | <.0001 | ETHNICITY3 HISPANIC |
| **ETHNICITY3** | **UNKNOWN** | 1 | 0.9462 | 0.0291 | 1056.6473 | <.0001 | ETHNICITY3 UNKNOWN |
| **decile_sviscore1** | | 1 | 0.0521 | 0.00406 | 164.6786 | <.0001 | Rank for Variable SVISCORE1 |
| **decile_sviscore4** | | 1 | 0.00986 | 0.00382 | 6.6767 | 0.0098 | Rank for Variable SVISCORE4 |
| **mortalityratepct** | | 1 | 0.0789 | 0.00311 | 643.3779 | <.0001 | |
| **Vaccinationsite** | **CHD OR DOH** | 1 | -0.3214 | 0.0271 | 140.5360 | <.0001 | Vaccinationsite CHD OR DOH |
| **Vaccinationsite** | **DOCTOR CLINIC** | 1 | 0.1533 | 0.0249 | 38.0701 | <.0001 | Vaccinationsite DOCTOR CLINIC |
| **Vaccinationsite** | **FIRE RESQUE OR EMS** | 1 | -0.2933 | 0.0537 | 29.8636 | <.0001 | Vaccinationsite FIRE RESQUE OR EMS |
| **Vaccinationsite** | **FQHC OR RHC OR CHC** | 1 | -0.3508 | 0.0860 | 16.6536 | <.0001 | Vaccinationsite FQHC OR RHC OR CHC |
| **Vaccinationsite** | **HOSPITAL CLINIC** | 1 | 0.0289 | 0.0381 | 0.5736 | 0.4488 | Vaccinationsite HOSPITAL CLINIC |
| **Vaccinationsite** | **HOSPITAL ER** | 1 | -0.0516 | 0.1691 | 0.0932 | 0.7602 | Vaccinationsite HOSPITAL ER |
| **Vaccinationsite** | **OTHER PHARMACIES** | 1 | 0.3533 | 0.0540 | 42.8357 | <.0001 | Vaccinationsite OTHER PHARMACIES |
| **Vaccinationsite** | **OTHERS OR MISC** | 1 | -1.5188 | 0.0460 | 1089.8668 | <.0001 | Vaccinationsite OTHERS OR MISC |

| Odds Ratio Estimates | | | |
|---|---|---|---|
| Effect | Point Estimate | 95% Wald Confidence Limits | |
| **firstdose 1. PFIZER vs 2. MODERN** | 1.374 | 1.325 | 1.425 |
| **agenew 40-49 vs 18-39** | 2.427 | 2.070 | 2.845 |
| **agenew 50-59 vs 18-39** | 4.980 | 4.360 | 5.689 |
| **agenew 60-69 vs 18-39** | 11.475 | 10.122 | 13.010 |
| **agenew 70-79 vs 18-39** | 25.587 | 22.572 | 29.005 |
| **agenew 80+ vs 18-39** | 49.768 | 43.449 | 57.005 |
| **sex2 F vs M** | 0.715 | 0.689 | 0.742 |

#### The LOGISTIC Procedure

| Odds Ratio Estimates | | | |
|---|---|---|---|
| Effect | Point Estimate | 95% Wald Confidence Limits | |
| RACE4 BLACK vs WHITE | 1.034 | 0.950 | 1.127 |
| RACE4 OTHERS vs WHITE | 0.616 | 0.575 | 0.660 |
| RACE4 UNKNOWN vs WHITE | 0.957 | 0.863 | 1.061 |
| ETHNICITY3 HISPANIC vs NOT HISPANIC | 0.584 | 0.545 | 0.626 |
| ETHNICITY3 UNKNOWN vs NOT HISPANIC | 2.576 | 2.433 | 2.727 |
| decile_sviscore1 | 1.053 | 1.045 | 1.062 |
| decile_sviscore4 | 1.010 | 1.002 | 1.017 |
| mortalityratepct | 1.082 | 1.075 | 1.089 |
| Vaccinationsite CHD OR DOH vs PUBLIX OR CVS OR WALGREENS | 0.725 | 0.688 | 0.765 |
| Vaccinationsite DOCTOR CLINIC vs PUBLIX OR CVS OR WALGREENS | 1.166 | 1.110 | 1.224 |
| Vaccinationsite FIRE RESQUE OR EMS vs PUBLIX OR CVS OR WALGREENS | 0.746 | 0.671 | 0.829 |
| Vaccinationsite FQHC OR RHC OR CHC vs PUBLIX OR CVS OR WALGREENS | 0.704 | 0.595 | 0.833 |
| Vaccinationsite HOSPITAL CLINIC vs PUBLIX OR CVS OR WALGREENS | 1.029 | 0.955 | 1.109 |
| Vaccinationsite HOSPITAL ER vs PUBLIX OR CVS OR WALGREENS | 0.950 | 0.682 | 1.323 |
| Vaccinationsite OTHER PHARMACIES vs PUBLIX OR CVS OR WALGREENS | 1.424 | 1.281 | 1.583 |
| Vaccinationsite OTHERS OR MISC vs PUBLIX OR CVS OR WALGREENS | 0.219 | 0.200 | 0.240 |

| Association of Predicted Probabilities and Observed Responses | | | |
|---|---|---|---|
| Percent Concordant | 83.3 | Somers' D | 0.666 |
| Percent Discordant | 16.7 | Gamma | 0.666 |
| Percent Tied | 0.0 | Tau-a | 0.008 |
| Pairs | 23968161792 | c | 0.833 |

### Matched cohort Risk of death Dose 1 NON COVID deaths

#### The LOGISTIC Procedure

| Model Information | |
|---|---|
| Data Set | WORK.MATCHEDCOHORT |
| Response Variable | NONCOVIDDEATH1YRD1 |
| Number of Response Levels | 2 |
| Model | binary logit |
| Optimization Technique | Fisher's scoring |

| | |
|---|---|
| Number of Observations Read | 1962822 |
| Number of Observations Used | 1962822 |

| Response Profile | | |
|---|---|---|
| Ordered Value | NONCOVIDDEATH1YRD1 | Total Frequency |
| 1 | 0 | 1950534 |
| 2 | 1 | 12288 |

**Probability modeled is NONCOVIDDEATH1YRD1=1.**

| Class Level Information | | |
|---|---|---|
| Class | Value | Design Variables |
| firstdose | 1. PFIZER | 1 |
| | 2. MODERN | 0 |

| Model Convergence Status |
|---|
| Convergence criterion (GCONV=1E-8) satisfied. |

| Model Fit Statistics | | |
|---|---|---|
| Criterion | Intercept Only | Intercept and Covariates |
| AIC | 149187.62 | 148897.18 |
| SC | 149200.11 | 148922.16 |
| -2 Log L | 149185.62 | 148893.18 |

| Testing Global Null Hypothesis: BETA=0 | | | |
|---|---|---|---|
| Test | Chi-Square | DF | Pr > ChiSq |
| Likelihood Ratio | 292.4400 | 1 | <.0001 |
| Score | 291.2927 | 1 | <.0001 |
| Wald | 288.9611 | 1 | <.0001 |

### Matched cohort Risk of death Dose 1 NON COVID deaths

#### The LOGISTIC Procedure

| Type 3 Analysis of Effects | | | |
|---|---|---|---|
| Effect | DF | Wald Chi-Square | Pr > ChiSq |
| firstdose | 1 | 288.9611 | <.0001 |

| Analysis of Maximum Likelihood Estimates | | | | | | | |
|---|---|---|---|---|---|---|---|
| Parameter | | DF | Estimate | Standard Error | Wald Chi-Square | Pr > ChiSq | Label |
| Intercept | | 1 | -5.2347 | 0.0139 | 141777.155 | <.0001 | Intercept: NONCOVIDDEATH1YRD1=1 |
| firstdose | 1. PFIZER | 1 | 0.3113 | 0.0183 | 288.9611 | <.0001 | firstdose 1. PFIZER |

| Odds Ratio Estimates | | | |
|---|---|---|---|
| Effect | Point Estimate | 95% Wald Confidence Limits | |
| firstdose 1. PFIZER vs 2. MODERN | 1.365 | 1.317 | 1.415 |

| Association of Predicted Probabilities and Observed Responses | | | |
|---|---|---|---|
| Percent Concordant | 28.9 | Somers' D | 0.077 |
| Percent Discordant | 21.1 | Gamma | 0.154 |
| Percent Tied | 50.0 | Tau-a | 0.001 |
| Pairs | 23968161792 | c | 0.539 |

### NON COVID deaths adjusted by Age

#### The LOGISTIC Procedure

##### agenew=18-39 years

| Model Information | |
|---|---|
| Data Set | WORK.MATCHEDCOHORT |
| Response Variable | NONCOVIDDEATH1YRD1 |
| Number of Response Levels | 2 |
| Model | binary logit |
| Optimization Technique | Fisher's scoring |

| | |
|---|---|
| Number of Observations Read | 492484 |
| Number of Observations Used | 492484 |

| Response Profile | | |
|---|---|---|
| Ordered Value | NONCOVIDDEATH1YRD1 | Total Frequency |
| 1 | 0 | 492208 |
| 2 | 1 | 276 |

**Probability modeled is NONCOVIDDEATH1YRD1=1.**

| Class Level Information | | | | | | | | | |
|---|---|---|---|---|---|---|---|---|---|
| Class | Value | Design Variables | | | | | | | |
| agenew | 18-39 years | | | | | | | | |
| firstdose | 1. PFIZER | 1 | | | | | | | |
| | 2. MODERN | 0 | | | | | | | |
| sex2 | F | 1 | | | | | | | |
| | M | 0 | | | | | | | |
| RACE4 | BLACK | 1 | 0 | 0 | | | | | |
| | OTHERS | 0 | 1 | 0 | | | | | |
| | UNKNOWN | 0 | 0 | 1 | | | | | |
| | WHITE | 0 | 0 | 0 | | | | | |
| ETHNICITY3 | HISPANIC | 1 | 0 | | | | | | |
| | NOT HISPANIC | 0 | 0 | | | | | | |
| | UNKNOWN | 0 | 1 | | | | | | |
| Vaccinationsite | CHD OR DOH | 1 | 0 | 0 | 0 | 0 | 0 | 0 | 0 |
| | DOCTOR CLINIC | 0 | 1 | 0 | 0 | 0 | 0 | 0 | 0 |
| | FIRE RESQUE OR EMS | 0 | 0 | 1 | 0 | 0 | 0 | 0 | 0 |
| | FQHC OR RHC OR CHC | 0 | 0 | 0 | 1 | 0 | 0 | 0 | 0 |
| | HOSPITAL CLINIC | 0 | 0 | 0 | 0 | 1 | 0 | 0 | 0 |
| | HOSPITAL ER | 0 | 0 | 0 | 0 | 0 | 1 | 0 | 0 |

#### The LOGISTIC Procedure

**agenew=18-39 years**

| Class Level Information | | | | | | | | | |
|---|---|---|---|---|---|---|---|---|---|
| **Class** | **Value** | **Design Variables** | | | | | | | |
| | **OTHER PHARMACIES** | 0 | 0 | 0 | 0 | 0 | 0 | 1 | 0 |
| | **OTHERS OR MISC** | 0 | 0 | 0 | 0 | 0 | 0 | 0 | 1 |
| | **PUBLIX OR CVS OR WALGREENS** | 0 | 0 | 0 | 0 | 0 | 0 | 0 | 0 |

| Model Convergence Status |
|---|
| Convergence criterion (GCONV=1E-8) satisfied. |

| Model Fit Statistics | | |
|---|---|---|
| **Criterion** | **Intercept Only** | **Intercept and Covariates** |
| **AIC** | 4686.568 | 4575.568 |
| **SC** | 4697.675 | 4786.605 |
| **-2 Log L** | 4684.568 | 4537.568 |

| Testing Global Null Hypothesis: BETA=0 | | | |
|---|---|---|---|
| **Test** | **Chi-Square** | **DF** | **Pr > ChiSq** |
| **Likelihood Ratio** | 146.9997 | 18 | <.0001 |
| **Score** | 147.3218 | 18 | <.0001 |
| **Wald** | 137.8613 | 18 | <.0001 |

| Type 3 Analysis of Effects | | | |
|---|---|---|---|
| **Effect** | **DF** | **Wald Chi-Square** | **Pr > ChiSq** |
| **firstdose** | 1 | 6.3397 | 0.0118 |
| **agenew** | 0 | . | . |
| **sex2** | 1 | 46.1992 | <.0001 |
| **RACE4** | 3 | 5.1974 | 0.1579 |
| **ETHNICITY3** | 2 | 46.7232 | <.0001 |
| **decile_sviscore1** | 1 | 17.0615 | <.0001 |
| **decile_sviscore4** | 1 | 0.3104 | 0.5774 |
| **mortalityratepct** | 1 | 0.0008 | 0.9769 |
| **Vaccinationsite** | 8 | 14.0070 | 0.0816 |

#### The LOGISTIC Procedure

**agenew=18-39 years**

| Analysis of Maximum Likelihood Estimates | | | | | | | |
|---|---|---|---|---|---|---|---|
| Parameter | | DF | Estimate | Standard Error | Wald Chi-Square | Pr > ChiSq | Label |
| Intercept | | 1 | -7.6244 | 0.1670 | 2083.4915 | <.0001 | Intercept: NONCOVIDDEATH1YRD1=1 |
| firstdose | 1. PFIZER | 1 | 0.3068 | 0.1219 | 6.3397 | 0.0118 | firstdose 1. PFIZER |
| sex2 | F | 1 | -0.8697 | 0.1280 | 46.1992 | <.0001 | sex2 F |
| RACE4 | BLACK | 1 | 0.1257 | 0.2006 | 0.3926 | 0.5309 | RACE4 BLACK |
| RACE4 | OTHERS | 1 | -0.0955 | 0.1709 | 0.3123 | 0.5763 | RACE4 OTHERS |
| RACE4 | UNKNOWN | 1 | -0.7218 | 0.3434 | 4.4197 | 0.0355 | RACE4 UNKNOWN |
| ETHNICITY3 | HISPANIC | 1 | -0.7657 | 0.1812 | 17.8484 | <.0001 | ETHNICITY3 HISPANIC |
| ETHNICITY3 | UNKNOWN | 1 | 0.6074 | 0.1527 | 15.8136 | <.0001 | ETHNICITY3 UNKNOWN |
| decile_sviscore1 | | 1 | 0.1104 | 0.0267 | 17.0615 | <.0001 | Rank for Variable SVISCORE1 |
| decile_sviscore4 | | 1 | -0.0141 | 0.0253 | 0.3104 | 0.5774 | Rank for Variable SVISCORE4 |
| mortalityratepct | | 1 | -10.8617 | 375.3 | 0.0008 | 0.9769 | |
| Vaccinationsite | CHD OR DOH | 1 | -0.3718 | 0.3624 | 1.0529 | 0.3048 | Vaccinationsite CHD OR DOH |
| Vaccinationsite | DOCTOR CLINIC | 1 | 0.1662 | 0.1783 | 0.8696 | 0.3511 | Vaccinationsite DOCTOR CLINIC |
| Vaccinationsite | FIRE RESQUE OR EMS | 1 | -10.7108 | 428.4 | 0.0006 | 0.9801 | Vaccinationsite FIRE RESQUE OR EMS |
| Vaccinationsite | FQHC OR RHC OR CHC | 1 | 0.1225 | 0.5867 | 0.0436 | 0.8347 | Vaccinationsite FQHC OR RHC OR CHC |
| Vaccinationsite | HOSPITAL CLINIC | 1 | -0.4731 | 0.4546 | 1.0829 | 0.2980 | Vaccinationsite HOSPITAL CLINIC |
| Vaccinationsite | HOSPITAL ER | 1 | 0.6003 | 1.0042 | 0.3574 | 0.5500 | Vaccinationsite HOSPITAL ER |
| Vaccinationsite | OTHER PHARMACIES | 1 | 0.2764 | 0.2388 | 1.3393 | 0.2472 | Vaccinationsite OTHER PHARMACIES |
| Vaccinationsite | OTHERS OR MISC | 1 | -1.4956 | 0.5066 | 8.7176 | 0.0032 | Vaccinationsite OTHERS OR MISC |

| Odds Ratio Estimates | | | |
|---|---|---|---|
| Effect | Point Estimate | 95% Wald Confidence Limits | |
| firstdose 1. PFIZER vs 2. MODERN | 1.359 | 1.070 | 1.726 |
| sex2 F vs M | 0.419 | 0.326 | 0.539 |
| RACE4 BLACK vs WHITE | 1.134 | 0.765 | 1.680 |
| RACE4 OTHERS vs WHITE | 0.909 | 0.650 | 1.271 |
| RACE4 UNKNOWN vs WHITE | 0.486 | 0.248 | 0.952 |
| ETHNICITY3 HISPANIC vs NOT HISPANIC | 0.465 | 0.326 | 0.663 |
| ETHNICITY3 UNKNOWN vs NOT HISPANIC | 1.836 | 1.361 | 2.476 |
| decile_sviscore1 | 1.117 | 1.060 | 1.177 |
| decile_sviscore4 | 0.986 | 0.938 | 1.036 |
| mortalityratepct | <0.001 | <0.001 | >999.999 |
| Vaccinationsite CHD OR DOH vs PUBLIX OR CVS OR WALGREENS | 0.689 | 0.339 | 1.403 |

#### The LOGISTIC Procedure

**agenew=18-39 years**

| Odds Ratio Estimates | | | |
|---|---|---|---|
| Effect | Point Estimate | 95% Wald Confidence Limits | |
| Vaccinationsite DOCTOR CLINIC vs PUBLIX OR CVS OR WALGREENS | 1.181 | 0.833 | 1.675 |
| Vaccinationsite FIRE RESQUE OR EMS vs PUBLIX OR CVS OR WALGREENS | <0.001 | <0.001 | >999.999 |
| Vaccinationsite FQHC OR RHC OR CHC vs PUBLIX OR CVS OR WALGREENS | 1.130 | 0.358 | 3.569 |
| Vaccinationsite HOSPITAL CLINIC vs PUBLIX OR CVS OR WALGREENS | 0.623 | 0.256 | 1.519 |
| Vaccinationsite HOSPITAL ER vs PUBLIX OR CVS OR WALGREENS | 1.823 | 0.255 | 13.046 |
| Vaccinationsite OTHER PHARMACIES vs PUBLIX OR CVS OR WALGREENS | 1.318 | 0.826 | 2.105 |
| Vaccinationsite OTHERS OR MISC vs PUBLIX OR CVS OR WALGREENS | 0.224 | 0.083 | 0.605 |

| Association of Predicted Probabilities and Observed Responses | | | |
|---|---|---|---|
| Percent Concordant | 70.7 | Somers' D | 0.415 |
| Percent Discordant | 29.2 | Gamma | 0.415 |
| Percent Tied | 0.0 | Tau-a | 0.000 |
| Pairs | 135849408 | c | 0.708 |

### The LOGISTIC Procedure

**agenew=40-59 years**

| Model Information | |
|---|---|
| **Data Set** | WORK.MATCHEDCOHORT |
| **Response Variable** | NONCOVIDDEATH1YRD1 |
| **Number of Response Levels** | 2 |
| **Model** | binary logit |
| **Optimization Technique** | Fisher's scoring |

| | |
|---|---|
| **Number of Observations Read** | 634102 |
| **Number of Observations Used** | 634102 |

| Response Profile | | |
|---|---|---|
| **Ordered Value** | **NONCOVIDDEATH1YRD1** | **Total Frequency** |
| **1** | 0 | 632718 |
| **2** | 1 | 1384 |

**Probability modeled is NONCOVIDDEATH1YRD1=1.**

| Class Level Information | | | | | | | | | |
|---|---|---|---|---|---|---|---|---|---|
| **Class** | **Value** | **Design Variables** | | | | | | | |
| **agenew** | **40-59 years** | | | | | | | | |
| **firstdose** | **1. PFIZER** | 1 | | | | | | | |
| | **2. MODERN** | 0 | | | | | | | |
| **sex2** | **F** | 1 | | | | | | | |
| | **M** | 0 | | | | | | | |
| **RACE4** | **BLACK** | 1 | 0 | 0 | | | | | |
| | **OTHERS** | 0 | 1 | 0 | | | | | |
| | **UNKNOWN** | 0 | 0 | 1 | | | | | |
| | **WHITE** | 0 | 0 | 0 | | | | | |
| **ETHNICITY3** | **HISPANIC** | 1 | 0 | | | | | | |
| | **NOT HISPANIC** | 0 | 0 | | | | | | |
| | **UNKNOWN** | 0 | 1 | | | | | | |
| **Vaccinationsite** | **CHD OR DOH** | 1 | 0 | 0 | 0 | 0 | 0 | 0 | 0 |
| | **DOCTOR CLINIC** | 0 | 1 | 0 | 0 | 0 | 0 | 0 | 0 |
| | **FIRE RESQUE OR EMS** | 0 | 0 | 1 | 0 | 0 | 0 | 0 | 0 |
| | **FQHC OR RHC OR CHC** | 0 | 0 | 0 | 1 | 0 | 0 | 0 | 0 |
| | **HOSPITAL CLINIC** | 0 | 0 | 0 | 0 | 1 | 0 | 0 | 0 |
| | **HOSPITAL ER** | 0 | 0 | 0 | 0 | 0 | 1 | 0 | 0 |

### The LOGISTIC Procedure

**agenew=40-59 years**

| Class Level Information | | | | | | | | | |
|---|---|---|---|---|---|---|---|---|---|
| Class | Value | Design Variables | | | | | | | |
| | OTHER PHARMACIES | 0 | 0 | 0 | 0 | 0 | 0 | 1 | 0 |
| | OTHERS OR MISC | 0 | 0 | 0 | 0 | 0 | 0 | 0 | 1 |
| | PUBLIX OR CVS OR WALGREENS | 0 | 0 | 0 | 0 | 0 | 0 | 0 | 0 |

| Model Convergence Status |
|---|
| Convergence criterion (GCONV=1E-8) satisfied. |

| Model Fit Statistics | | |
|---|---|---|
| Criterion | Intercept Only | Intercept and Covariates |
| AIC | 19727.155 | 18977.170 |
| SC | 19738.515 | 19193.009 |
| -2 Log L | 19725.155 | 18939.170 |

| Testing Global Null Hypothesis: BETA=0 | | | |
|---|---|---|---|
| Test | Chi-Square | DF | Pr > ChiSq |
| Likelihood Ratio | 785.9852 | 18 | <.0001 |
| Score | 798.4404 | 18 | <.0001 |
| Wald | 750.7531 | 18 | <.0001 |

| Type 3 Analysis of Effects | | | |
|---|---|---|---|
| Effect | DF | Wald Chi-Square | Pr > ChiSq |
| firstdose | 1 | 7.0570 | 0.0079 |
| agenew | 0 | . | . |
| sex2 | 1 | 28.1087 | <.0001 |
| RACE4 | 3 | 14.8718 | 0.0019 |
| ETHNICITY3 | 2 | 155.4374 | <.0001 |
| decile_sviscore1 | 1 | 72.0482 | <.0001 |
| decile_sviscore4 | 1 | 0.8021 | 0.3705 |
| mortalityratepct | 1 | 165.3420 | <.0001 |
| Vaccinationsite | 8 | 132.0224 | <.0001 |

#### The LOGISTIC Procedure

**agenew=40-59 years**

| Analysis of Maximum Likelihood Estimates | | | | | | | |
|---|---|---|---|---|---|---|---|
| Parameter | | DF | Estimate | Standard Error | Wald Chi-Square | Pr > ChiSq | Label |
| Intercept | | 1 | -6.9263 | 0.0856 | 6552.8927 | <.0001 | Intercept: NONCOVIDDEATH1YRD1=1 |
| firstdose | 1. PFIZER | 1 | 0.1434 | 0.0540 | 7.0570 | 0.0079 | firstdose 1. PFIZER |
| sex2 | F | 1 | -0.3079 | 0.0581 | 28.1087 | <.0001 | sex2 F |
| RACE4 | BLACK | 1 | 0.0478 | 0.0899 | 0.2826 | 0.5950 | RACE4 BLACK |
| RACE4 | OTHERS | 1 | -0.3964 | 0.1085 | 13.3403 | 0.0003 | RACE4 OTHERS |
| RACE4 | UNKNOWN | 1 | -0.1914 | 0.1434 | 1.7813 | 0.1820 | RACE4 UNKNOWN |
| ETHNICITY3 | HISPANIC | 1 | -0.7535 | 0.0932 | 65.3554 | <.0001 | ETHNICITY3 HISPANIC |
| ETHNICITY3 | UNKNOWN | 1 | 0.6129 | 0.0821 | 55.7130 | <.0001 | ETHNICITY3 UNKNOWN |
| decile_sviscore1 | | 1 | 0.1035 | 0.0122 | 72.0482 | <.0001 | Rank for Variable SVISCORE1 |
| decile_sviscore4 | | 1 | 0.0103 | 0.0115 | 0.8021 | 0.3705 | Rank for Variable SVISCORE4 |
| mortalityratepct | | 1 | 0.7372 | 0.0573 | 165.3420 | <.0001 | |
| Vaccinationsite | CHD OR DOH | 1 | -0.1010 | 0.1560 | 0.4194 | 0.5172 | Vaccinationsite CHD OR DOH |
| Vaccinationsite | DOCTOR CLINIC | 1 | 0.2715 | 0.0710 | 14.6280 | 0.0001 | Vaccinationsite DOCTOR CLINIC |
| Vaccinationsite | FIRE RESQUE OR EMS | 1 | -0.3660 | 0.7090 | 0.2665 | 0.6057 | Vaccinationsite FIRE RESQUE OR EMS |
| Vaccinationsite | FQHC OR RHC OR CHC | 1 | 0.2703 | 0.2060 | 1.7219 | 0.1894 | Vaccinationsite FQHC OR RHC OR CHC |
| Vaccinationsite | HOSPITAL CLINIC | 1 | 0.6977 | 0.1008 | 47.9197 | <.0001 | Vaccinationsite HOSPITAL CLINIC |
| Vaccinationsite | HOSPITAL ER | 1 | 0.0985 | 0.5022 | 0.0385 | 0.8445 | Vaccinationsite HOSPITAL ER |
| Vaccinationsite | OTHER PHARMACIES | 1 | 0.4456 | 0.1047 | 18.1277 | <.0001 | Vaccinationsite OTHER PHARMACIES |
| Vaccinationsite | OTHERS OR MISC | 1 | -1.1157 | 0.1602 | 48.5097 | <.0001 | Vaccinationsite OTHERS OR MISC |

| Odds Ratio Estimates | | | |
|---|---|---|---|
| Effect | Point Estimate | 95% Wald Confidence Limits | |
| firstdose 1. PFIZER vs 2. MODERN | 1.154 | 1.038 | 1.283 |
| sex2 F vs M | 0.735 | 0.656 | 0.824 |
| RACE4 BLACK vs WHITE | 1.049 | 0.880 | 1.251 |
| RACE4 OTHERS vs WHITE | 0.673 | 0.544 | 0.832 |
| RACE4 UNKNOWN vs WHITE | 0.826 | 0.623 | 1.094 |
| ETHNICITY3 HISPANIC vs NOT HISPANIC | 0.471 | 0.392 | 0.565 |
| ETHNICITY3 UNKNOWN vs NOT HISPANIC | 1.846 | 1.571 | 2.168 |
| decile_sviscore1 | 1.109 | 1.083 | 1.136 |
| decile_sviscore4 | 1.010 | 0.988 | 1.033 |
| mortalityratepct | 2.090 | 1.868 | 2.339 |
| Vaccinationsite CHD OR DOH vs PUBLIX OR CVS OR WALGREENS | 0.904 | 0.666 | 1.227 |

### The LOGISTIC Procedure

**agenew=40-59 years**

| Odds Ratio Estimates | | | |
|---|---|---|---|
| Effect | Point Estimate | 95% Wald Confidence Limits | |
| Vaccinationsite DOCTOR CLINIC vs PUBLIX OR CVS OR WALGREENS | 1.312 | 1.142 | 1.508 |
| Vaccinationsite FIRE RESQUE OR EMS vs PUBLIX OR CVS OR WALGREENS | 0.693 | 0.173 | 2.783 |
| Vaccinationsite FQHC OR RHC OR CHC vs PUBLIX OR CVS OR WALGREENS | 1.310 | 0.875 | 1.962 |
| Vaccinationsite HOSPITAL CLINIC vs PUBLIX OR CVS OR WALGREENS | 2.009 | 1.649 | 2.448 |
| Vaccinationsite HOSPITAL ER vs PUBLIX OR CVS OR WALGREENS | 1.104 | 0.412 | 2.953 |
| Vaccinationsite OTHER PHARMACIES vs PUBLIX OR CVS OR WALGREENS | 1.561 | 1.272 | 1.917 |
| Vaccinationsite OTHERS OR MISC vs PUBLIX OR CVS OR WALGREENS | 0.328 | 0.239 | 0.449 |

| Association of Predicted Probabilities and Observed Responses | | | |
|---|---|---|---|
| Percent Concordant | 71.1 | Somers' D | 0.422 |
| Percent Discordant | 28.9 | Gamma | 0.422 |
| Percent Tied | 0.0 | Tau-a | 0.002 |
| Pairs | 875681712 | c | 0.711 |

### NON COVID deaths adjusted by Age

#### The LOGISTIC Procedure

**agenew=60+ years**

| Model Information | |
|---|---|
| **Data Set** | WORK.MATCHEDCOHORT |
| **Response Variable** | NONCOVIDDEATH1YRD1 |
| **Number of Response Levels** | 2 |
| **Model** | binary logit |
| **Optimization Technique** | Fisher's scoring |

| | |
|---|---|
| **Number of Observations Read** | 836236 |
| **Number of Observations Used** | 836236 |

| Response Profile | | |
|---|---|---|
| **Ordered Value** | **NONCOVIDDEATH1YRD1** | **Total Frequency** |
| **1** | 0 | 825608 |
| **2** | 1 | 10628 |

**Probability modeled is NONCOVIDDEATH1YRD1=1.**

| Class Level Information | | | | | | | | | |
|---|---|---|---|---|---|---|---|---|---|
| **Class** | **Value** | **Design Variables** | | | | | | | |
| **agenew** | **60+ years** | | | | | | | | |
| **firstdose** | **1. PFIZER** | 1 | | | | | | | |
| | **2. MODERN** | 0 | | | | | | | |
| **sex2** | **F** | 1 | | | | | | | |
| | **M** | 0 | | | | | | | |
| **RACE4** | **BLACK** | 1 | 0 | 0 | | | | | |
| | **OTHERS** | 0 | 1 | 0 | | | | | |
| | **UNKNOWN** | 0 | 0 | 1 | | | | | |
| | **WHITE** | 0 | 0 | 0 | | | | | |
| **ETHNICITY3** | **HISPANIC** | 1 | 0 | | | | | | |
| | **NOT HISPANIC** | 0 | 0 | | | | | | |
| | **UNKNOWN** | 0 | 1 | | | | | | |
| **Vaccinationsite** | **CHD OR DOH** | 1 | 0 | 0 | 0 | 0 | 0 | 0 | 0 |
| | **DOCTOR CLINIC** | 0 | 1 | 0 | 0 | 0 | 0 | 0 | 0 |
| | **FIRE RESQUE OR EMS** | 0 | 0 | 1 | 0 | 0 | 0 | 0 | 0 |
| | **FQHC OR RHC OR CHC** | 0 | 0 | 0 | 1 | 0 | 0 | 0 | 0 |
| | **HOSPITAL CLINIC** | 0 | 0 | 0 | 0 | 1 | 0 | 0 | 0 |
| | **HOSPITAL ER** | 0 | 0 | 0 | 0 | 0 | 1 | 0 | 0 |

#### The LOGISTIC Procedure

##### agenew=60+ years

| Class Level Information | | | | | | | | | |
|---|---|---|---|---|---|---|---|---|---|
| Class | Value | Design Variables | | | | | | | |
| | OTHER PHARMACIES | 0 | 0 | 0 | 0 | 0 | 0 | 1 | 0 |
| | OTHERS OR MISC | 0 | 0 | 0 | 0 | 0 | 0 | 0 | 1 |
| | PUBLIX OR CVS OR WALGREENS | 0 | 0 | 0 | 0 | 0 | 0 | 0 | 0 |

| Model Convergence Status |
|---|
| Convergence criterion (GCONV=1E-8) satisfied. |

| Model Fit Statistics | | |
|---|---|---|
| Criterion | Intercept Only | Intercept and Covariates |
| AIC | 113913.69 | 105853.08 |
| SC | 113925.33 | 106074.18 |
| -2 Log L | 113911.69 | 105815.08 |

| Testing Global Null Hypothesis: BETA=0 | | | |
|---|---|---|---|
| Test | Chi-Square | DF | Pr > ChiSq |
| Likelihood Ratio | 8096.6101 | 18 | <.0001 |
| Score | 12388.7778 | 18 | <.0001 |
| Wald | 8665.3529 | 18 | <.0001 |

| Type 3 Analysis of Effects | | | |
|---|---|---|---|
| Effect | DF | Wald Chi-Square | Pr > ChiSq |
| firstdose | 1 | 292.4983 | <.0001 |
| agenew | 0 | . | . |
| sex2 | 1 | 105.3796 | <.0001 |
| RACE4 | 3 | 184.7028 | <.0001 |
| ETHNICITY3 | 2 | 1398.3486 | <.0001 |
| decile_sviscore1 | 1 | 52.1052 | <.0001 |
| decile_sviscore4 | 1 | 10.5809 | 0.0011 |
| mortalityratepct | 1 | 5599.7442 | <.0001 |
| Vaccinationsite | 8 | 1194.6947 | <.0001 |

#### The LOGISTIC Procedure

**agenew=60+ years**

| Analysis of Maximum Likelihood Estimates | | | | | | | |
|---|---|---|---|---|---|---|---|
| Parameter | | DF | Estimate | Standard Error | Wald Chi-Square | Pr > ChiSq | Label |
| Intercept | | 1 | -5.0358 | 0.0305 | 27208.8013 | <.0001 | Intercept: NONCOVIDDEATH1YRD1=1 |
| firstdose | 1. PFIZER | 1 | 0.3425 | 0.0200 | 292.4983 | <.0001 | firstdose 1. PFIZER |
| sex2 | F | 1 | -0.2053 | 0.0200 | 105.3796 | <.0001 | sex2 F |
| RACE4 | BLACK | 1 | -0.1620 | 0.0515 | 9.9033 | 0.0016 | RACE4 BLACK |
| RACE4 | OTHERS | 1 | -0.5351 | 0.0403 | 176.5361 | <.0001 | RACE4 OTHERS |
| RACE4 | UNKNOWN | 1 | 0.0230 | 0.0578 | 0.1576 | 0.6914 | RACE4 UNKNOWN |
| ETHNICITY3 | HISPANIC | 1 | -0.5588 | 0.0407 | 188.9102 | <.0001 | ETHNICITY3 HISPANIC |
| ETHNICITY3 | UNKNOWN | 1 | 1.0442 | 0.0317 | 1084.2737 | <.0001 | ETHNICITY3 UNKNOWN |
| decile_sviscore1 | | 1 | 0.0316 | 0.00438 | 52.1052 | <.0001 | Rank for Variable SVISCORE1 |
| decile_sviscore4 | | 1 | 0.0134 | 0.00412 | 10.5809 | 0.0011 | Rank for Variable SVISCORE4 |
| mortalityratepct | | 1 | 0.1646 | 0.00220 | 5599.7442 | <.0001 | |
| Vaccinationsite | CHD OR DOH | 1 | -0.1807 | 0.0276 | 42.8401 | <.0001 | Vaccinationsite CHD OR DOH |
| Vaccinationsite | DOCTOR CLINIC | 1 | 0.2430 | 0.0268 | 82.2645 | <.0001 | Vaccinationsite DOCTOR CLINIC |
| Vaccinationsite | FIRE RESQUE OR EMS | 1 | -0.0398 | 0.0543 | 0.5368 | 0.4638 | Vaccinationsite FIRE RESQUE OR EMS |
| Vaccinationsite | FQHC OR RHC OR CHC | 1 | -0.3304 | 0.0956 | 11.9375 | 0.0006 | Vaccinationsite FQHC OR RHC OR CHC |
| Vaccinationsite | HOSPITAL CLINIC | 1 | 0.0168 | 0.0417 | 0.1618 | 0.6875 | Vaccinationsite HOSPITAL CLINIC |
| Vaccinationsite | HOSPITAL ER | 1 | -0.0107 | 0.1820 | 0.0034 | 0.9532 | Vaccinationsite HOSPITAL ER |
| Vaccinationsite | OTHER PHARMACIES | 1 | 0.2551 | 0.0655 | 15.1873 | <.0001 | Vaccinationsite OTHER PHARMACIES |
| Vaccinationsite | OTHERS OR MISC | 1 | -1.4287 | 0.0484 | 872.7402 | <.0001 | Vaccinationsite OTHERS OR MISC |

| Odds Ratio Estimates | | | |
|---|---|---|---|
| Effect | Point Estimate | 95% Wald Confidence Limits | |
| firstdose 1. PFIZER vs 2. MODERN | 1.408 | 1.354 | 1.465 |
| sex2 F vs M | 0.814 | 0.783 | 0.847 |
| RACE4 BLACK vs WHITE | 0.850 | 0.769 | 0.941 |
| RACE4 OTHERS vs WHITE | 0.586 | 0.541 | 0.634 |
| RACE4 UNKNOWN vs WHITE | 1.023 | 0.914 | 1.146 |
| ETHNICITY3 HISPANIC vs NOT HISPANIC | 0.572 | 0.528 | 0.619 |
| ETHNICITY3 UNKNOWN vs NOT HISPANIC | 2.841 | 2.670 | 3.023 |
| decile_sviscore1 | 1.032 | 1.023 | 1.041 |
| decile_sviscore4 | 1.013 | 1.005 | 1.022 |
| mortalityratepct | 1.179 | 1.174 | 1.184 |
| Vaccinationsite CHD OR DOH vs PUBLIX OR CVS OR WALGREENS | 0.835 | 0.791 | 0.881 |

### NON COVID deaths adjusted by Age

#### The LOGISTIC Procedure

##### agenew=60+ years

| Odds Ratio Estimates | | | |
|---|---|---|---|
| Effect | Point Estimate | 95% Wald Confidence Limits | |
| Vaccinationsite DOCTOR CLINIC vs PUBLIX OR CVS OR WALGREENS | 1.275 | 1.210 | 1.344 |
| Vaccinationsite FIRE RESQUE OR EMS vs PUBLIX OR CVS OR WALGREENS | 0.961 | 0.864 | 1.069 |
| Vaccinationsite FQHC OR RHC OR CHC vs PUBLIX OR CVS OR WALGREENS | 0.719 | 0.596 | 0.867 |
| Vaccinationsite HOSPITAL CLINIC vs PUBLIX OR CVS OR WALGREENS | 1.017 | 0.937 | 1.104 |
| Vaccinationsite HOSPITAL ER vs PUBLIX OR CVS OR WALGREENS | 0.989 | 0.692 | 1.414 |
| Vaccinationsite OTHER PHARMACIES vs PUBLIX OR CVS OR WALGREENS | 1.291 | 1.135 | 1.467 |
| Vaccinationsite OTHERS OR MISC vs PUBLIX OR CVS OR WALGREENS | 0.240 | 0.218 | 0.263 |

| Association of Predicted Probabilities and Observed Responses | | | |
|---|---|---|---|
| Percent Concordant | 73.4 | Somers' D | 0.467 |
| Percent Discordant | 26.6 | Gamma | 0.467 |
| Percent Tied | 0.0 | Tau-a | 0.012 |
| Pairs | 8774561824 | c | 0.734 |

#### The LOGISTIC Procedure

| Model Information | |
|---|---|
| **Data Set** | WORK.MATCHEDCOHORT |
| **Response Variable** | COVIDDEATHPLUS1YRD1 |
| **Number of Response Levels** | 2 |
| **Model** | binary logit |
| **Optimization Technique** | Fisher's scoring |

| | |
|---|---|
| **Number of Observations Read** | 1962822 |
| **Number of Observations Used** | 1962822 |

| Response Profile | | |
|---|---|---|
| **Ordered Value** | **COVIDDEATHPLUS1YRD1** | **Total Frequency** |
| **1** | 0 | 1961185 |
| **2** | 1 | 1637 |

**Probability modeled is COVIDDEATHPLUS1YRD1=1.**

| Class Level Information | | |
|---|---|---|
| **Class** | **Value** | **Design Variables** |
| **firstdose** | **1. PFIZER** | 1 |
| | **2. MODERN** | 0 |

| Model Convergence Status |
|---|
| Convergence criterion (GCONV=1E-8) satisfied. |

| Model Fit Statistics | | |
|---|---|---|
| **Criterion** | **Intercept Only** | **Intercept and Covariates** |
| **AIC** | 26484.915 | 26335.989 |
| **SC** | 26497.405 | 26360.969 |
| **-2 Log L** | 26482.915 | 26331.989 |

| Testing Global Null Hypothesis: BETA=0 | | | |
|---|---|---|---|
| **Test** | **Chi-Square** | **DF** | **Pr > ChiSq** |
| **Likelihood Ratio** | 150.9261 | 1 | <.0001 |
| **Score** | 148.5961 | 1 | <.0001 |
| **Wald** | 143.9058 | 1 | <.0001 |

#### The LOGISTIC Procedure

| Type 3 Analysis of Effects | | | |
|---|---|---|---|
| Effect | DF | Wald Chi-Square | Pr > ChiSq |
| firstdose | 1 | 143.9058 | <.0001 |

| Analysis of Maximum Likelihood Estimates | | | | | | | |
|---|---|---|---|---|---|---|---|
| Parameter | | DF | Estimate | Standard Error | Wald Chi-Square | Pr > ChiSq | Label |
| Intercept | | 1 | -7.4470 | 0.0418 | 31703.6174 | <.0001 | Intercept: COVIDDEATHPLUS1YRD1=1 |
| firstdose | 1. PFIZER | 1 | 0.6221 | 0.0519 | 143.9058 | <.0001 | firstdose 1. PFIZER |

| Odds Ratio Estimates | | | |
|---|---|---|---|
| Effect | Point Estimate | 95% Wald Confidence Limits | |
| firstdose 1. PFIZER vs 2. MODERN | 1.863 | 1.683 | 2.062 |

| Association of Predicted Probabilities and Observed Responses | | | |
|---|---|---|---|
| Percent Concordant | 32.5 | Somers' D | 0.151 |
| Percent Discordant | 17.5 | Gamma | 0.301 |
| Percent Tied | 50.0 | Tau-a | 0.000 |
| Pairs | 3210459845 | c | 0.575 |

#### The LOGISTIC Procedure

| Model Information | |
|---|---|
| **Data Set** | WORK.MATCHEDCOHORT |
| **Response Variable** | COVIDDEATHPLUS1YRD1 |
| **Number of Response Levels** | 2 |
| **Model** | binary logit |
| **Optimization Technique** | Fisher's scoring |

| | |
|---|---|
| **Number of Observations Read** | 1962822 |
| **Number of Observations Used** | 1962822 |

| Response Profile | | |
|---|---|---|
| **Ordered Value** | **COVIDDEATHPLUS1YRD1** | **Total Frequency** |
| **1** | 0 | 1961185 |
| **2** | 1 | 1637 |

**Probability modeled is COVIDDEATHPLUS1YRD1=1.**

| Class Level Information | | | | | | | | | |
|---|---|---|---|---|---|---|---|---|---|
| **Class** | **Value** | **Design Variables** | | | | | | | |
| **agenew** | **18-39** | 0 | 0 | 0 | 0 | 0 | | | |
| | **40-49** | 1 | 0 | 0 | 0 | 0 | | | |
| | **50-59** | 0 | 1 | 0 | 0 | 0 | | | |
| | **60-69** | 0 | 0 | 1 | 0 | 0 | | | |
| | **70-79** | 0 | 0 | 0 | 1 | 0 | | | |
| | **80+** | 0 | 0 | 0 | 0 | 1 | | | |
| **firstdose** | **1. PFIZER** | 1 | | | | | | | |
| | **2. MODERN** | 0 | | | | | | | |
| **sex2** | **F** | 1 | | | | | | | |
| | **M** | 0 | | | | | | | |
| **RACE4** | **BLACK** | 1 | 0 | 0 | | | | | |
| | **OTHERS** | 0 | 1 | 0 | | | | | |
| | **UNKNOWN** | 0 | 0 | 1 | | | | | |
| | **WHITE** | 0 | 0 | 0 | | | | | |
| **ETHNICITY3** | **HISPANIC** | 1 | 0 | | | | | | |
| | **NOT HISPANIC** | 0 | 0 | | | | | | |
| | **UNKNOWN** | 0 | 1 | | | | | | |
| **Vaccinationsite** | **CHD OR DOH** | 1 | 0 | 0 | 0 | 0 | 0 | 0 | 0 |
| | **DOCTOR CLINIC** | 0 | 1 | 0 | 0 | 0 | 0 | 0 | 0 |

#### The LOGISTIC Procedure

| Class Level Information | | | | | | | | | |
|---|---|---|---|---|---|---|---|---|---|
| **Class** | **Value** | **Design Variables** | | | | | | | |
| | **FIRE RESQUE OR EMS** | 0 | 0 | 1 | 0 | 0 | 0 | 0 | 0 |
| | **FQHC OR RHC OR CHC** | 0 | 0 | 0 | 1 | 0 | 0 | 0 | 0 |
| | **HOSPITAL CLINIC** | 0 | 0 | 0 | 0 | 1 | 0 | 0 | 0 |
| | **HOSPITAL ER** | 0 | 0 | 0 | 0 | 0 | 1 | 0 | 0 |
| | **OTHER PHARMACIES** | 0 | 0 | 0 | 0 | 0 | 0 | 1 | 0 |
| | **OTHERS OR MISC** | 0 | 0 | 0 | 0 | 0 | 0 | 0 | 1 |
| | **PUBLIX OR CVS OR WALGREENS** | 0 | 0 | 0 | 0 | 0 | 0 | 0 | 0 |

| Model Convergence Status |
|---|
| Convergence criterion (GCONV=1E-8) satisfied. |

| Model Fit Statistics | | |
|---|---|---|
| **Criterion** | **Intercept Only** | **Intercept and Covariates** |
| **AIC** | 26484.915 | 23418.429 |
| **SC** | 26497.405 | 23718.187 |
| **-2 Log L** | 26482.915 | 23370.429 |

| Testing Global Null Hypothesis: BETA=0 | | | |
|---|---|---|---|
| **Test** | **Chi-Square** | **DF** | **Pr > ChiSq** |
| **Likelihood Ratio** | 3112.4857 | 23 | <.0001 |
| **Score** | 4639.1003 | 23 | <.0001 |
| **Wald** | 2784.5880 | 23 | <.0001 |

| Type 3 Analysis of Effects | | | |
|---|---|---|---|
| **Effect** | **DF** | **Wald Chi-Square** | **Pr > ChiSq** |
| **firstdose** | 1 | 144.2727 | <.0001 |
| **agenew** | 5 | 999.2663 | <.0001 |
| **sex2** | 1 | 49.2529 | <.0001 |
| **RACE4** | 3 | 25.1301 | <.0001 |
| **ETHNICITY3** | 2 | 155.5961 | <.0001 |
| **decile_sviscore1** | 1 | 61.7374 | <.0001 |
| **decile_sviscore4** | 1 | 2.0506 | 0.1521 |
| **mortalityratepct** | 1 | 70.0149 | <.0001 |
| **Vaccinationsite** | 8 | 271.8083 | <.0001 |

#### The LOGISTIC Procedure

| Analysis of Maximum Likelihood Estimates | | | | | | | |
|---|---|---|---|---|---|---|---|
| Parameter | | DF | Estimate | Standard Error | Wald Chi-Square | Pr > ChiSq | Label |
| Intercept | | 1 | -10.3007 | 0.1838 | 3140.5926 | <.0001 | Intercept: COVIDDEATHPLUS1YRD1=1 |
| firstdose | 1. PFIZER | 1 | 0.6239 | 0.0519 | 144.2727 | <.0001 | firstdose 1. PFIZER |
| agenew | 40-49 | 1 | 1.2705 | 0.2116 | 36.0681 | <.0001 | agenew 40-49 |
| agenew | 50-59 | 1 | 1.6979 | 0.1898 | 79.9911 | <.0001 | agenew 50-59 |
| agenew | 60-69 | 1 | 2.5507 | 0.1796 | 201.7113 | <.0001 | agenew 60-69 |
| agenew | 70-79 | 1 | 3.5310 | 0.1785 | 391.3353 | <.0001 | agenew 70-79 |
| agenew | 80+ | 1 | 4.2766 | 0.1895 | 509.2397 | <.0001 | agenew 80+ |
| sex2 | F | 1 | -0.3561 | 0.0507 | 49.2529 | <.0001 | sex2 F |
| RACE4 | BLACK | 1 | 0.4064 | 0.1010 | 16.1968 | <.0001 | RACE4 BLACK |
| RACE4 | OTHERS | 1 | -0.2259 | 0.0870 | 6.7463 | 0.0094 | RACE4 OTHERS |
| RACE4 | UNKNOWN | 1 | 0.0806 | 0.1362 | 0.3497 | 0.5543 | RACE4 UNKNOWN |
| ETHNICITY3 | HISPANIC | 1 | -0.3090 | 0.0877 | 12.4000 | 0.0004 | ETHNICITY3 HISPANIC |
| ETHNICITY3 | UNKNOWN | 1 | 0.8866 | 0.0789 | 126.3975 | <.0001 | ETHNICITY3 UNKNOWN |
| decile_sviscore1 | | 1 | 0.0868 | 0.0110 | 61.7374 | <.0001 | Rank for Variable SVISCORE1 |
| decile_sviscore4 | | 1 | 0.0148 | 0.0104 | 2.0506 | 0.1521 | Rank for Variable SVISCORE4 |
| mortalityratepct | | 1 | 0.0623 | 0.00744 | 70.0149 | <.0001 | |
| Vaccinationsite | CHD OR DOH | 1 | -0.6129 | 0.0765 | 64.1828 | <.0001 | Vaccinationsite CHD OR DOH |
| Vaccinationsite | DOCTOR CLINIC | 1 | -0.0166 | 0.0658 | 0.0635 | 0.8010 | Vaccinationsite DOCTOR CLINIC |
| Vaccinationsite | FIRE RESQUE OR EMS | 1 | -0.6159 | 0.1491 | 17.0546 | <.0001 | Vaccinationsite FIRE RESQUE OR EMS |
| Vaccinationsite | FQHC OR RHC OR CHC | 1 | -0.7082 | 0.2537 | 7.7918 | 0.0052 | Vaccinationsite FQHC OR RHC OR CHC |
| Vaccinationsite | HOSPITAL CLINIC | 1 | -0.4209 | 0.1167 | 13.0003 | 0.0003 | Vaccinationsite HOSPITAL CLINIC |
| Vaccinationsite | HOSPITAL ER | 1 | -10.8465 | 97.5728 | 0.0124 | 0.9115 | Vaccinationsite HOSPITAL ER |
| Vaccinationsite | OTHER PHARMACIES | 1 | 0.4138 | 0.1294 | 10.2276 | 0.0014 | Vaccinationsite OTHER PHARMACIES |
| Vaccinationsite | OTHERS OR MISC | 1 | -1.7545 | 0.1297 | 182.8812 | <.0001 | Vaccinationsite OTHERS OR MISC |

| Odds Ratio Estimates | | | |
|---|---|---|---|
| Effect | Point Estimate | 95% Wald Confidence Limits | |
| firstdose 1. PFIZER vs 2. MODERN | 1.866 | 1.685 | 2.066 |
| agenew 40-49 vs 18-39 | 3.563 | 2.353 | 5.393 |
| agenew 50-59 vs 18-39 | 5.463 | 3.765 | 7.925 |
| agenew 60-69 vs 18-39 | 12.816 | 9.013 | 18.224 |
| agenew 70-79 vs 18-39 | 34.157 | 24.074 | 48.464 |
| agenew 80+ vs 18-39 | 71.992 | 49.656 | 104.375 |
| sex2 F vs M | 0.700 | 0.634 | 0.774 |

#### The LOGISTIC Procedure

| Odds Ratio Estimates | | | |
|---|---|---|---|
| Effect | Point Estimate | 95% Wald Confidence Limits | |
| RACE4 BLACK vs WHITE | 1.501 | 1.232 | 1.830 |
| RACE4 OTHERS vs WHITE | 0.798 | 0.673 | 0.946 |
| RACE4 UNKNOWN vs WHITE | 1.084 | 0.830 | 1.416 |
| ETHNICITY3 HISPANIC vs NOT HISPANIC | 0.734 | 0.618 | 0.872 |
| ETHNICITY3 UNKNOWN vs NOT HISPANIC | 2.427 | 2.079 | 2.832 |
| decile_sviscore1 | 1.091 | 1.067 | 1.115 |
| decile_sviscore4 | 1.015 | 0.995 | 1.036 |
| mortalityratepct | 1.064 | 1.049 | 1.080 |
| Vaccinationsite CHD OR DOH vs PUBLIX OR CVS OR WALGREENS | 0.542 | 0.466 | 0.629 |
| Vaccinationsite DOCTOR CLINIC vs PUBLIX OR CVS OR WALGREENS | 0.984 | 0.865 | 1.119 |
| Vaccinationsite FIRE RESQUE OR EMS vs PUBLIX OR CVS OR WALGREENS | 0.540 | 0.403 | 0.724 |
| Vaccinationsite FQHC OR RHC OR CHC vs PUBLIX OR CVS OR WALGREENS | 0.493 | 0.300 | 0.810 |
| Vaccinationsite HOSPITAL CLINIC vs PUBLIX OR CVS OR WALGREENS | 0.656 | 0.522 | 0.825 |
| Vaccinationsite HOSPITAL ER vs PUBLIX OR CVS OR WALGREENS | <0.001 | <0.001 | >999.999 |
| Vaccinationsite OTHER PHARMACIES vs PUBLIX OR CVS OR WALGREENS | 1.513 | 1.174 | 1.949 |
| Vaccinationsite OTHERS OR MISC vs PUBLIX OR CVS OR WALGREENS | 0.173 | 0.134 | 0.223 |

| Association of Predicted Probabilities and Observed Responses | | | |
|---|---|---|---|
| Percent Concordant | 84.1 | Somers' D | 0.682 |
| Percent Discordant | 15.9 | Gamma | 0.682 |
| Percent Tied | 0.0 | Tau-a | 0.001 |
| Pairs | 3210459845 | c | 0.841 |

### COVID + deaths adjusted by Age

#### The LOGISTIC Procedure

**agenew=18-39 years**

| Model Information | |
|---|---|
| **Data Set** | WORK.MATCHEDCOHORT |
| **Response Variable** | COVIDDEATHPLUS1YRD1 |
| **Number of Response Levels** | 2 |
| **Model** | binary logit |
| **Optimization Technique** | Fisher's scoring |

| | |
|---|---|
| **Number of Observations Read** | 492484 |
| **Number of Observations Used** | 492484 |

| Response Profile | | |
|---|---|---|
| **Ordered Value** | **COVIDDEATHPLUS1YRD1** | **Total Frequency** |
| **1** | 0 | 492449 |
| **2** | 1 | 35 |

**Probability modeled is COVIDDEATHPLUS1YRD1=1.**

| Class Level Information | | | | | | | | | |
|---|---|---|---|---|---|---|---|---|---|
| **Class** | **Value** | **Design Variables** | | | | | | | |
| **agenew** | **18-39 years** | | | | | | | | |
| **firstdose** | **1. PFIZER** | 1 | | | | | | | |
| | **2. MODERN** | 0 | | | | | | | |
| **sex2** | **F** | 1 | | | | | | | |
| | **M** | 0 | | | | | | | |
| **RACE4** | **BLACK** | 1 | 0 | 0 | | | | | |
| | **OTHERS** | 0 | 1 | 0 | | | | | |
| | **UNKNOWN** | 0 | 0 | 1 | | | | | |
| | **WHITE** | 0 | 0 | 0 | | | | | |
| **ETHNICITY3** | **HISPANIC** | 1 | 0 | | | | | | |
| | **NOT HISPANIC** | 0 | 0 | | | | | | |
| | **UNKNOWN** | 0 | 1 | | | | | | |
| **Vaccinationsite** | **CHD OR DOH** | 1 | 0 | 0 | 0 | 0 | 0 | 0 | 0 |
| | **DOCTOR CLINIC** | 0 | 1 | 0 | 0 | 0 | 0 | 0 | 0 |
| | **FIRE RESQUE OR EMS** | 0 | 0 | 1 | 0 | 0 | 0 | 0 | 0 |
| | **FQHC OR RHC OR CHC** | 0 | 0 | 0 | 1 | 0 | 0 | 0 | 0 |
| | **HOSPITAL CLINIC** | 0 | 0 | 0 | 0 | 1 | 0 | 0 | 0 |
| | **HOSPITAL ER** | 0 | 0 | 0 | 0 | 0 | 1 | 0 | 0 |

#### The LOGISTIC Procedure

**agenew=18-39 years**

| Class Level Information | | | | | | | | | |
|---|---|---|---|---|---|---|---|---|---|
| Class | Value | Design Variables | | | | | | | |
| | OTHER PHARMACIES | 0 | 0 | 0 | 0 | 0 | 0 | 1 | 0 |
| | OTHERS OR MISC | 0 | 0 | 0 | 0 | 0 | 0 | 0 | 1 |
| | PUBLIX OR CVS OR WALGREENS | 0 | 0 | 0 | 0 | 0 | 0 | 0 | 0 |

| Model Convergence Status |
|---|
| Quasi-complete separation of data points detected. |

**Warning:** The maximum likelihood estimate may not exist.

**Warning:** The LOGISTIC procedure continues in spite of the above warning. Results shown are based on the last maximum likelihood iteration. Validity of the model fit is questionable.

| Model Fit Statistics | | |
|---|---|---|
| Criterion | Intercept Only | Intercept and Covariates |
| AIC | 740.628 | 736.312 |
| SC | 751.736 | 947.349 |
| -2 Log L | 738.628 | 698.312 |

| Testing Global Null Hypothesis: BETA=0 | | | |
|---|---|---|---|
| Test | Chi-Square | DF | Pr > ChiSq |
| Likelihood Ratio | 40.3168 | 18 | 0.0019 |
| Score | 38.0398 | 18 | 0.0038 |
| Wald | 28.4093 | 18 | 0.0561 |

| Type 3 Analysis of Effects | | | |
|---|---|---|---|
| Effect | DF | Wald Chi-Square | Pr > ChiSq |
| firstdose | 1 | 3.3340 | 0.0679 |
| agenew | 0 | . | . |
| sex2 | 1 | 0.3802 | 0.5375 |
| RACE4 | 3 | 0.9258 | 0.8192 |
| ETHNICITY3 | 2 | 6.2710 | 0.0435 |
| decile_sviscore1 | 1 | 12.2845 | 0.0005 |
| decile_sviscore4 | 1 | 2.1696 | 0.1408 |
| mortalityratepct | 1 | 0.0000 | 0.9956 |
| Vaccinationsite | 8 | 7.2383 | 0.5112 |

#### The LOGISTIC Procedure

**Warning:** The validity of the model fit is questionable.

**agenew=18-39 years**

| Analysis of Maximum Likelihood Estimates | | | | | | | |
|---|---|---|---|---|---|---|---|
| **Parameter** | | **DF** | **Estimate** | **Standard Error** | **Wald Chi-Square** | **Pr > ChiSq** | **Label** |
| **Intercept** | | 1 | -10.8857 | 0.5540 | 386.0768 | <.0001 | Intercept: COVIDDEATHPLUS1YRD1=1 |
| **firstdose** | **1. PFIZER** | 1 | 0.6503 | 0.3561 | 3.3340 | 0.0679 | firstdose 1. PFIZER |
| **sex2** | **F** | 1 | 0.2136 | 0.3463 | 0.3802 | 0.5375 | sex2 F |
| **RACE4** | **BLACK** | 1 | -0.2873 | 0.5644 | 0.2591 | 0.6107 | RACE4 BLACK |
| **RACE4** | **OTHERS** | 1 | -0.2100 | 0.4989 | 0.1772 | 0.6738 | RACE4 OTHERS |
| **RACE4** | **UNKNOWN** | 1 | -0.8537 | 1.0724 | 0.6338 | 0.4260 | RACE4 UNKNOWN |
| **ETHNICITY3** | **HISPANIC** | 1 | -0.9734 | 0.4879 | 3.9797 | 0.0461 | ETHNICITY3 HISPANIC |
| **ETHNICITY3** | **UNKNOWN** | 1 | 0.4240 | 0.4518 | 0.8810 | 0.3479 | ETHNICITY3 UNKNOWN |
| **decile_sviscore1** | | 1 | 0.2783 | 0.0794 | 12.2845 | 0.0005 | Rank for Variable SVISCORE1 |
| **decile_sviscore4** | | 1 | -0.1029 | 0.0698 | 2.1696 | 0.1408 | Rank for Variable SVISCORE4 |
| **mortalityratepct** | | 1 | -11.9850 | 2159.0 | 0.0000 | 0.9956 | |
| **Vaccinationsite** | **CHD OR DOH** | 1 | -12.6608 | 481.5 | 0.0007 | 0.9790 | Vaccinationsite CHD OR DOH |
| **Vaccinationsite** | **DOCTOR CLINIC** | 1 | 0.6740 | 0.4378 | 2.3702 | 0.1237 | Vaccinationsite DOCTOR CLINIC |
| **Vaccinationsite** | **FIRE RESQUE OR EMS** | 1 | -12.4638 | 3182.2 | 0.0000 | 0.9969 | Vaccinationsite FIRE RESQUE OR EMS |
| **Vaccinationsite** | **FQHC OR RHC OR CHC** | 1 | -12.4139 | 898.8 | 0.0002 | 0.9890 | Vaccinationsite FQHC OR RHC OR CHC |
| **Vaccinationsite** | **HOSPITAL CLINIC** | 1 | -12.4954 | 521.2 | 0.0006 | 0.9809 | Vaccinationsite HOSPITAL CLINIC |
| **Vaccinationsite** | **HOSPITAL ER** | 1 | -12.6168 | 2076.5 | 0.0000 | 0.9952 | Vaccinationsite HOSPITAL ER |
| **Vaccinationsite** | **OTHER PHARMACIES** | 1 | 1.1847 | 0.4746 | 6.2305 | 0.0126 | Vaccinationsite OTHER PHARMACIES |
| **Vaccinationsite** | **OTHERS OR MISC** | 1 | -12.6322 | 417.9 | 0.0009 | 0.9759 | Vaccinationsite OTHERS OR MISC |

| Odds Ratio Estimates | | | |
|---|---|---|---|
| **Effect** | **Point Estimate** | **95% Wald Confidence Limits** | |
| **firstdose 1. PFIZER vs 2. MODERN** | 1.916 | 0.953 | 3.851 |
| **sex2 F vs M** | 1.238 | 0.628 | 2.441 |
| **RACE4 BLACK vs WHITE** | 0.750 | 0.248 | 2.268 |
| **RACE4 OTHERS vs WHITE** | 0.811 | 0.305 | 2.155 |
| **RACE4 UNKNOWN vs WHITE** | 0.426 | 0.052 | 3.484 |
| **ETHNICITY3 HISPANIC vs NOT HISPANIC** | 0.378 | 0.145 | 0.983 |
| **ETHNICITY3 UNKNOWN vs NOT HISPANIC** | 1.528 | 0.630 | 3.704 |
| **decile_sviscore1** | 1.321 | 1.130 | 1.543 |
| **decile_sviscore4** | 0.902 | 0.787 | 1.035 |
| **mortalityratepct** | <0.001 | <0.001 | >999.999 |
| **Vaccinationsite CHD OR DOH vs PUBLIX OR CVS OR WALGREENS** | <0.001 | <0.001 | >999.999 |

#### The LOGISTIC Procedure

**Warning:** The validity of the model fit is questionable.

**agenew=18-39 years**

| Odds Ratio Estimates | | | |
|---|---|---|---|
| Effect | Point Estimate | 95% Wald Confidence Limits | |
| Vaccinationsite DOCTOR CLINIC vs PUBLIX OR CVS OR WALGREENS | 1.962 | 0.832 | 4.627 |
| Vaccinationsite FIRE RESQUE OR EMS vs PUBLIX OR CVS OR WALGREENS | <0.001 | <0.001 | >999.999 |
| Vaccinationsite FQHC OR RHC OR CHC vs PUBLIX OR CVS OR WALGREENS | <0.001 | <0.001 | >999.999 |
| Vaccinationsite HOSPITAL CLINIC vs PUBLIX OR CVS OR WALGREENS | <0.001 | <0.001 | >999.999 |
| Vaccinationsite HOSPITAL ER vs PUBLIX OR CVS OR WALGREENS | <0.001 | <0.001 | >999.999 |
| Vaccinationsite OTHER PHARMACIES vs PUBLIX OR CVS OR WALGREENS | 3.270 | 1.290 | 8.289 |
| Vaccinationsite OTHERS OR MISC vs PUBLIX OR CVS OR WALGREENS | <0.001 | <0.001 | >999.999 |

| Association of Predicted Probabilities and Observed Responses | | | |
|---|---|---|---|
| Percent Concordant | 76.8 | Somers' D | 0.536 |
| Percent Discordant | 23.2 | Gamma | 0.536 |
| Percent Tied | 0.0 | Tau-a | 0.000 |
| Pairs | 17235715 | c | 0.768 |

### COVID + deaths adjusted by Age

#### The LOGISTIC Procedure

**agenew=40-59 years**

| Model Information | |
|---|---|
| **Data Set** | WORK.MATCHEDCOHORT |
| **Response Variable** | COVIDDEATHPLUS1YRD1 |
| **Number of Response Levels** | 2 |
| **Model** | binary logit |
| **Optimization Technique** | Fisher's scoring |

| | |
|---|---|
| **Number of Observations Read** | 634102 |
| **Number of Observations Used** | 634102 |

| Response Profile | | |
|---|---|---|
| **Ordered Value** | **COVIDDEATHPLUS1YRD1** | **Total Frequency** |
| **1** | 0 | 633903 |
| **2** | 1 | 199 |

**Probability modeled is COVIDDEATHPLUS1YRD1=1.**

| Class Level Information | | | | | | | | | |
|---|---|---|---|---|---|---|---|---|---|
| **Class** | **Value** | **Design Variables** | | | | | | | |
| **agenew** | **40-59 years** | | | | | | | | |
| **firstdose** | **1. PFIZER** | 1 | | | | | | | |
| | **2. MODERN** | 0 | | | | | | | |
| **sex2** | **F** | 1 | | | | | | | |
| | **M** | 0 | | | | | | | |
| **RACE4** | **BLACK** | 1 | 0 | 0 | | | | | |
| | **OTHERS** | 0 | 1 | 0 | | | | | |
| | **UNKNOWN** | 0 | 0 | 1 | | | | | |
| | **WHITE** | 0 | 0 | 0 | | | | | |
| **ETHNICITY3** | **HISPANIC** | 1 | 0 | | | | | | |
| | **NOT HISPANIC** | 0 | 0 | | | | | | |
| | **UNKNOWN** | 0 | 1 | | | | | | |
| **Vaccinationsite** | **CHD OR DOH** | 1 | 0 | 0 | 0 | 0 | 0 | 0 | 0 |
| | **DOCTOR CLINIC** | 0 | 1 | 0 | 0 | 0 | 0 | 0 | 0 |
| | **FIRE RESQUE OR EMS** | 0 | 0 | 1 | 0 | 0 | 0 | 0 | 0 |
| | **FQHC OR RHC OR CHC** | 0 | 0 | 0 | 1 | 0 | 0 | 0 | 0 |
| | **HOSPITAL CLINIC** | 0 | 0 | 0 | 0 | 1 | 0 | 0 | 0 |
| | **HOSPITAL ER** | 0 | 0 | 0 | 0 | 0 | 1 | 0 | 0 |

#### The LOGISTIC Procedure

##### agenew=40-59 years

| Class Level Information | | | | | | | | | |
|---|---|---|---|---|---|---|---|---|---|
| **Class** | **Value** | **Design Variables** | | | | | | | |
| | **OTHER PHARMACIES** | 0 | 0 | 0 | 0 | 0 | 0 | 1 | 0 |
| | **OTHERS OR MISC** | 0 | 0 | 0 | 0 | 0 | 0 | 0 | 1 |
| | **PUBLIX OR CVS OR WALGREENS** | 0 | 0 | 0 | 0 | 0 | 0 | 0 | 0 |

| Model Convergence Status |
|---|
| Convergence criterion (GCONV=1E-8) satisfied. |

| Model Fit Statistics | | |
|---|---|---|
| **Criterion** | **Intercept Only** | **Intercept and Covariates** |
| **AIC** | 3610.468 | 3500.734 |
| **SC** | 3621.828 | 3716.573 |
| **-2 Log L** | 3608.468 | 3462.734 |

| Testing Global Null Hypothesis: BETA=0 | | | |
|---|---|---|---|
| **Test** | **Chi-Square** | **DF** | **Pr > ChiSq** |
| **Likelihood Ratio** | 145.7346 | 18 | <.0001 |
| **Score** | 163.9868 | 18 | <.0001 |
| **Wald** | 149.8314 | 18 | <.0001 |

| Type 3 Analysis of Effects | | | |
|---|---|---|---|
| **Effect** | **DF** | **Wald Chi-Square** | **Pr > ChiSq** |
| **firstdose** | 1 | 4.2195 | 0.0400 |
| **agenew** | 0 | . | . |
| **sex2** | 1 | 0.0003 | 0.9853 |
| **RACE4** | 3 | 13.6885 | 0.0034 |
| **ETHNICITY3** | 2 | 19.0803 | <.0001 |
| **decile_sviscore1** | 1 | 9.0146 | 0.0027 |
| **decile_sviscore4** | 1 | 9.0196 | 0.0027 |
| **mortalityratepct** | 1 | 16.4263 | <.0001 |
| **Vaccinationsite** | 8 | 21.5551 | 0.0058 |

#### The LOGISTIC Procedure

##### agenew=40-59 years

| Analysis of Maximum Likelihood Estimates | | | | | | | |
|---|---|---|---|---|---|---|---|
| Parameter | | DF | Estimate | Standard Error | Wald Chi-Square | Pr > ChiSq | Label |
| Intercept | | 1 | -9.4776 | 0.2413 | 1543.3079 | <.0001 | Intercept: COVIDDEATHPLUS1YRD1=1 |
| firstdose | 1. PFIZER | 1 | 0.2945 | 0.1434 | 4.2195 | 0.0400 | firstdose 1. PFIZER |
| sex2 | F | 1 | 0.00276 | 0.1501 | 0.0003 | 0.9853 | sex2 F |
| RACE4 | BLACK | 1 | 0.5901 | 0.1968 | 8.9899 | 0.0027 | RACE4 BLACK |
| RACE4 | OTHERS | 1 | -0.4720 | 0.2891 | 2.6659 | 0.1025 | RACE4 OTHERS |
| RACE4 | UNKNOWN | 1 | -0.3478 | 0.4019 | 0.7492 | 0.3867 | RACE4 UNKNOWN |
| ETHNICITY3 | HISPANIC | 1 | -0.5047 | 0.2288 | 4.8659 | 0.0274 | ETHNICITY3 HISPANIC |
| ETHNICITY3 | UNKNOWN | 1 | 0.6755 | 0.2160 | 9.7846 | 0.0018 | ETHNICITY3 UNKNOWN |
| decile_sviscore1 | | 1 | 0.0994 | 0.0331 | 9.0146 | 0.0027 | Rank for Variable SVISCORE1 |
| decile_sviscore4 | | 1 | 0.0943 | 0.0314 | 9.0196 | 0.0027 | Rank for Variable SVISCORE4 |
| mortalityratepct | | 1 | 0.5989 | 0.1478 | 16.4263 | <.0001 | |
| Vaccinationsite | CHD OR DOH | 1 | -0.1725 | 0.4212 | 0.1678 | 0.6821 | Vaccinationsite CHD OR DOH |
| Vaccinationsite | DOCTOR CLINIC | 1 | 0.0353 | 0.1937 | 0.0332 | 0.8555 | Vaccinationsite DOCTOR CLINIC |
| Vaccinationsite | FIRE RESQUE OR EMS | 1 | -11.4469 | 433.2 | 0.0007 | 0.9789 | Vaccinationsite FIRE RESQUE OR EMS |
| Vaccinationsite | FQHC OR RHC OR CHC | 1 | 0.0744 | 0.5153 | 0.0208 | 0.8852 | Vaccinationsite FQHC OR RHC OR CHC |
| Vaccinationsite | HOSPITAL CLINIC | 1 | 0.2733 | 0.3183 | 0.7370 | 0.3906 | Vaccinationsite HOSPITAL CLINIC |
| Vaccinationsite | HOSPITAL ER | 1 | -11.4158 | 416.4 | 0.0008 | 0.9781 | Vaccinationsite HOSPITAL ER |
| Vaccinationsite | OTHER PHARMACIES | 1 | 0.7742 | 0.2274 | 11.5941 | 0.0007 | Vaccinationsite OTHER PHARMACIES |
| Vaccinationsite | OTHERS OR MISC | 1 | -1.1184 | 0.4234 | 6.9771 | 0.0083 | Vaccinationsite OTHERS OR MISC |

| Odds Ratio Estimates | | | |
|---|---|---|---|
| Effect | Point Estimate | 95% Wald Confidence Limits | |
| firstdose 1. PFIZER vs 2. MODERN | 1.342 | 1.014 | 1.778 |
| sex2 F vs M | 1.003 | 0.747 | 1.346 |
| RACE4 BLACK vs WHITE | 1.804 | 1.227 | 2.653 |
| RACE4 OTHERS vs WHITE | 0.624 | 0.354 | 1.099 |
| RACE4 UNKNOWN vs WHITE | 0.706 | 0.321 | 1.552 |
| ETHNICITY3 HISPANIC vs NOT HISPANIC | 0.604 | 0.385 | 0.945 |
| ETHNICITY3 UNKNOWN vs NOT HISPANIC | 1.965 | 1.287 | 3.000 |
| decile_sviscore1 | 1.104 | 1.035 | 1.179 |
| decile_sviscore4 | 1.099 | 1.033 | 1.169 |
| mortalityratepct | 1.820 | 1.362 | 2.432 |
| Vaccinationsite CHD OR DOH vs PUBLIX OR CVS OR WALGREENS | 0.842 | 0.369 | 1.921 |

#### The LOGISTIC Procedure

**agenew=40-59 years**

| Odds Ratio Estimates | | | |
|---|---|---|---|
| Effect | Point Estimate | 95% Wald Confidence Limits | |
| Vaccinationsite DOCTOR CLINIC vs PUBLIX OR CVS OR WALGREENS | 1.036 | 0.709 | 1.514 |
| Vaccinationsite FIRE RESQUE OR EMS vs PUBLIX OR CVS OR WALGREENS | <0.001 | <0.001 | >999.999 |
| Vaccinationsite FQHC OR RHC OR CHC vs PUBLIX OR CVS OR WALGREENS | 1.077 | 0.392 | 2.958 |
| Vaccinationsite HOSPITAL CLINIC vs PUBLIX OR CVS OR WALGREENS | 1.314 | 0.704 | 2.453 |
| Vaccinationsite HOSPITAL ER vs PUBLIX OR CVS OR WALGREENS | <0.001 | <0.001 | >999.999 |
| Vaccinationsite OTHER PHARMACIES vs PUBLIX OR CVS OR WALGREENS | 2.169 | 1.389 | 3.387 |
| Vaccinationsite OTHERS OR MISC vs PUBLIX OR CVS OR WALGREENS | 0.327 | 0.143 | 0.749 |

| Association of Predicted Probabilities and Observed Responses | | | |
|---|---|---|---|
| Percent Concordant | 72.9 | Somers' D | 0.459 |
| Percent Discordant | 27.0 | Gamma | 0.459 |
| Percent Tied | 0.0 | Tau-a | 0.000 |
| Pairs | 126146697 | c | 0.729 |

### COVID + deaths adjusted by Age

#### The LOGISTIC Procedure

##### agenew=60+ years

| Model Information | |
|---|---|
| Data Set | WORK.MATCHEDCOHORT |
| Response Variable | COVIDDEATHPLUS1YRD1 |
| Number of Response Levels | 2 |
| Model | binary logit |
| Optimization Technique | Fisher's scoring |

| | |
|---|---|
| Number of Observations Read | 836236 |
| Number of Observations Used | 836236 |

| Response Profile | | |
|---|---|---|
| Ordered Value | COVIDDEATHPLUS1YRD1 | Total Frequency |
| 1 | 0 | 834833 |
| 2 | 1 | 1403 |

**Probability modeled is COVIDDEATHPLUS1YRD1=1.**

| Class Level Information | | | | | | | | | |
|---|---|---|---|---|---|---|---|---|---|
| Class | Value | Design Variables | | | | | | | |
| agenew | 60+ years | | | | | | | | |
| firstdose | 1. PFIZER | 1 | | | | | | | |
| | 2. MODERN | 0 | | | | | | | |
| sex2 | F | 1 | | | | | | | |
| | M | 0 | | | | | | | |
| RACE4 | BLACK | 1 | 0 | 0 | | | | | |
| | OTHERS | 0 | 1 | 0 | | | | | |
| | UNKNOWN | 0 | 0 | 1 | | | | | |
| | WHITE | 0 | 0 | 0 | | | | | |
| ETHNICITY3 | HISPANIC | 1 | 0 | | | | | | |
| | NOT HISPANIC | 0 | 0 | | | | | | |
| | UNKNOWN | 0 | 1 | | | | | | |
| Vaccinationsite | CHD OR DOH | 1 | 0 | 0 | 0 | 0 | 0 | 0 | 0 |
| | DOCTOR CLINIC | 0 | 1 | 0 | 0 | 0 | 0 | 0 | 0 |
| | FIRE RESQUE OR EMS | 0 | 0 | 1 | 0 | 0 | 0 | 0 | 0 |
| | FQHC OR RHC OR CHC | 0 | 0 | 0 | 1 | 0 | 0 | 0 | 0 |
| | HOSPITAL CLINIC | 0 | 0 | 0 | 0 | 1 | 0 | 0 | 0 |
| | HOSPITAL ER | 0 | 0 | 0 | 0 | 0 | 1 | 0 | 0 |

#### The LOGISTIC Procedure

**agenew=60+ years**

| Class Level Information | | | | | | | | | |
|---|---|---|---|---|---|---|---|---|---|
| Class | Value | Design Variables | | | | | | | |
| | OTHER PHARMACIES | 0 | 0 | 0 | 0 | 0 | 0 | 1 | 0 |
| | OTHERS OR MISC | 0 | 0 | 0 | 0 | 0 | 0 | 0 | 1 |
| | PUBLIX OR CVS OR WALGREENS | 0 | 0 | 0 | 0 | 0 | 0 | 0 | 0 |

| Model Convergence Status |
|---|
| Convergence criterion (GCONV=1E-8) satisfied. |

| Model Fit Statistics | | |
|---|---|---|
| Criterion | Intercept Only | Intercept and Covariates |
| AIC | 20736.821 | 19563.269 |
| SC | 20748.458 | 19784.366 |
| -2 Log L | 20734.821 | 19525.269 |

| Testing Global Null Hypothesis: BETA=0 | | | |
|---|---|---|---|
| Test | Chi-Square | DF | Pr > ChiSq |
| Likelihood Ratio | 1209.5520 | 18 | <.0001 |
| Score | 1892.2293 | 18 | <.0001 |
| Wald | 1574.6429 | 18 | <.0001 |

| Type 3 Analysis of Effects | | | |
|---|---|---|---|
| Effect | DF | Wald Chi-Square | Pr > ChiSq |
| firstdose | 1 | 142.2597 | <.0001 |
| agenew | 0 | . | . |
| sex2 | 1 | 24.7785 | <.0001 |
| RACE4 | 3 | 17.9683 | 0.0004 |
| ETHNICITY3 | 2 | 166.2213 | <.0001 |
| decile_sviscore1 | 1 | 30.8186 | <.0001 |
| decile_sviscore4 | 1 | 0.9671 | 0.3254 |
| mortalityratepct | 1 | 935.5849 | <.0001 |
| Vaccinationsite | 8 | 198.2299 | <.0001 |

#### The LOGISTIC Procedure

**agenew=60+ years**

| Analysis of Maximum Likelihood Estimates | | | | | | | |
|---|---|---|---|---|---|---|---|
| Parameter | | DF | Estimate | Standard Error | Wald Chi-Square | Pr > ChiSq | Label |
| Intercept | | 1 | -7.1620 | 0.0827 | 7497.0039 | <.0001 | Intercept: COVIDDEATHPLUS1YRD1=1 |
| firstdose | 1. PFIZER | 1 | 0.6763 | 0.0567 | 142.2597 | <.0001 | firstdose 1. PFIZER |
| sex2 | F | 1 | -0.2718 | 0.0546 | 24.7785 | <.0001 | sex2 F |
| RACE4 | BLACK | 1 | 0.1127 | 0.1217 | 0.8573 | 0.3545 | RACE4 BLACK |
| RACE4 | OTHERS | 1 | -0.3785 | 0.1002 | 14.2623 | 0.0002 | RACE4 OTHERS |
| RACE4 | UNKNOWN | 1 | 0.1873 | 0.1466 | 1.6326 | 0.2013 | RACE4 UNKNOWN |
| ETHNICITY3 | HISPANIC | 1 | -0.4268 | 0.1040 | 16.8569 | <.0001 | ETHNICITY3 HISPANIC |
| ETHNICITY3 | UNKNOWN | 1 | 0.9925 | 0.0859 | 133.6137 | <.0001 | ETHNICITY3 UNKNOWN |
| decile_sviscore1 | | 1 | 0.0661 | 0.0119 | 30.8186 | <.0001 | Rank for Variable SVISCORE1 |
| decile_sviscore4 | | 1 | 0.0110 | 0.0112 | 0.9671 | 0.3254 | Rank for Variable SVISCORE4 |
| mortalityratepct | | 1 | 0.1313 | 0.00429 | 935.5849 | <.0001 | |
| Vaccinationsite | CHD OR DOH | 1 | -0.4366 | 0.0775 | 31.7715 | <.0001 | Vaccinationsite CHD OR DOH |
| Vaccinationsite | DOCTOR CLINIC | 1 | 0.0775 | 0.0706 | 1.2051 | 0.2723 | Vaccinationsite DOCTOR CLINIC |
| Vaccinationsite | FIRE RESQUE OR EMS | 1 | -0.2766 | 0.1499 | 3.4035 | 0.0651 | Vaccinationsite FIRE RESQUE OR EMS |
| Vaccinationsite | FQHC OR RHC OR CHC | 1 | -0.7099 | 0.2923 | 5.8977 | 0.0152 | Vaccinationsite FQHC OR RHC OR CHC |
| Vaccinationsite | HOSPITAL CLINIC | 1 | -0.4766 | 0.1282 | 13.8191 | 0.0002 | Vaccinationsite HOSPITAL CLINIC |
| Vaccinationsite | HOSPITAL ER | 1 | -11.4861 | 145.2 | 0.0063 | 0.9370 | Vaccinationsite HOSPITAL ER |
| Vaccinationsite | OTHER PHARMACIES | 1 | 0.0833 | 0.1716 | 0.2354 | 0.6276 | Vaccinationsite OTHER PHARMACIES |
| Vaccinationsite | OTHERS OR MISC | 1 | -1.6519 | 0.1359 | 147.6576 | <.0001 | Vaccinationsite OTHERS OR MISC |

| Odds Ratio Estimates | | | |
|---|---|---|---|
| Effect | Point Estimate | 95% Wald Confidence Limits | |
| firstdose 1. PFIZER vs 2. MODERN | 1.967 | 1.760 | 2.198 |
| sex2 F vs M | 0.762 | 0.685 | 0.848 |
| RACE4 BLACK vs WHITE | 1.119 | 0.882 | 1.421 |
| RACE4 OTHERS vs WHITE | 0.685 | 0.563 | 0.834 |
| RACE4 UNKNOWN vs WHITE | 1.206 | 0.905 | 1.607 |
| ETHNICITY3 HISPANIC vs NOT HISPANIC | 0.653 | 0.532 | 0.800 |
| ETHNICITY3 UNKNOWN vs NOT HISPANIC | 2.698 | 2.280 | 3.192 |
| decile_sviscore1 | 1.068 | 1.044 | 1.094 |
| decile_sviscore4 | 1.011 | 0.989 | 1.034 |
| mortalityratepct | 1.140 | 1.131 | 1.150 |
| Vaccinationsite CHD OR DOH vs PUBLIX OR CVS OR WALGREENS | 0.646 | 0.555 | 0.752 |

#### The LOGISTIC Procedure

**agenew=60+ years**

| Odds Ratio Estimates | | | |
|---|---|---|---|
| Effect | Point Estimate | 95% Wald Confidence Limits | |
| Vaccinationsite DOCTOR CLINIC vs PUBLIX OR CVS OR WALGREENS | 1.081 | 0.941 | 1.241 |
| Vaccinationsite FIRE RESQUE OR EMS vs PUBLIX OR CVS OR WALGREENS | 0.758 | 0.565 | 1.017 |
| Vaccinationsite FQHC OR RHC OR CHC vs PUBLIX OR CVS OR WALGREENS | 0.492 | 0.277 | 0.872 |
| Vaccinationsite HOSPITAL CLINIC vs PUBLIX OR CVS OR WALGREENS | 0.621 | 0.483 | 0.798 |
| Vaccinationsite HOSPITAL ER vs PUBLIX OR CVS OR WALGREENS | <0.001 | <0.001 | >999.999 |
| Vaccinationsite OTHER PHARMACIES vs PUBLIX OR CVS OR WALGREENS | 1.087 | 0.776 | 1.521 |
| Vaccinationsite OTHERS OR MISC vs PUBLIX OR CVS OR WALGREENS | 0.192 | 0.147 | 0.250 |

| Association of Predicted Probabilities and Observed Responses | | | |
|---|---|---|---|
| Percent Concordant | 74.5 | Somers' D | 0.491 |
| Percent Discordant | 25.4 | Gamma | 0.491 |
| Percent Tied | 0.0 | Tau-a | 0.002 |
| Pairs | 1171270699 | c | 0.746 |

### Matched cohort Risk of death Dose 1 NON COVID + deaths

#### The LOGISTIC Procedure

| Model Information | |
|---|---|
| Data Set | WORK.MATCHEDCOHORT |
| Response Variable | NONCOVIDDEATHPLUS1YRD1 |
| Number of Response Levels | 2 |
| Model | binary logit |
| Optimization Technique | Fisher's scoring |

| | |
|---|---|
| Number of Observations Read | 1962822 |
| Number of Observations Used | 1962822 |

| Response Profile | | |
|---|---|---|
| Ordered Value | NONCOVIDDEATHPLUS1YRD1 | Total Frequency |
| 1 | 0 | 1951250 |
| 2 | 1 | 11572 |

**Probability modeled is NONCOVIDDEATHPLUS1YRD1=1.**

| Class Level Information | | |
|---|---|---|
| Class | Value | Design Variables |
| firstdose | 1. PFIZER | 1 |
| | 2. MODERN | 0 |

| Model Convergence Status |
|---|
| Convergence criterion (GCONV=1E-8) satisfied. |

| Model Fit Statistics | | |
|---|---|---|
| Criterion | Intercept Only | Intercept and Covariates |
| AIC | 141888.53 | 141644.90 |
| SC | 141901.02 | 141669.88 |
| -2 Log L | 141886.53 | 141640.90 |

| Testing Global Null Hypothesis: BETA=0 | | | |
|---|---|---|---|
| Test | Chi-Square | DF | Pr > ChiSq |
| Likelihood Ratio | 245.6216 | 1 | <.0001 |
| Score | 244.7617 | 1 | <.0001 |
| Wald | 243.0171 | 1 | <.0001 |

### Matched cohort Risk of death Dose 1 NON COVID + deaths

#### The LOGISTIC Procedure

| Type 3 Analysis of Effects | | | |
|---|---|---|---|
| **Effect** | **DF** | **Wald Chi-Square** | **Pr > ChiSq** |
| **firstdose** | 1 | 243.0171 | <.0001 |

| Analysis of Maximum Likelihood Estimates | | | | | | | |
|---|---|---|---|---|---|---|---|
| **Parameter** | | **DF** | **Estimate** | **Standard Error** | **Wald Chi-Square** | **Pr > ChiSq** | **Label** |
| **Intercept** | | 1 | -5.2851 | 0.0143 | 137493.042 | <.0001 | Intercept: NONCOVIDDEATHPLUS1YRD1=1 |
| **firstdose** | **1. PFIZER** | 1 | 0.2938 | 0.0188 | 243.0171 | <.0001 | firstdose 1. PFIZER |

| Odds Ratio Estimates | | | |
|---|---|---|---|
| **Effect** | **Point Estimate** | **95% Wald Confidence Limits** | |
| **firstdose 1. PFIZER vs 2. MODERN** | 1.341 | 1.293 | 1.392 |

| Association of Predicted Probabilities and Observed Responses | | | |
|---|---|---|---|
| **Percent Concordant** | 28.6 | **Somers' D** | 0.073 |
| **Percent Discordant** | 21.4 | **Gamma** | 0.146 |
| **Percent Tied** | 50.0 | **Tau-a** | 0.001 |
| **Pairs** | 22579865000 | **c** | 0.536 |

#### The LOGISTIC Procedure

| Model Information | |
|---|---|
| **Data Set** | WORK.MATCHEDCOHORT |
| **Response Variable** | NONCOVIDDEATHPLUS1YRD1 |
| **Number of Response Levels** | 2 |
| **Model** | binary logit |
| **Optimization Technique** | Fisher's scoring |

| | |
|---|---|
| **Number of Observations Read** | 1962822 |
| **Number of Observations Used** | 1962822 |

| Response Profile | | |
|---|---|---|
| **Ordered Value** | **NONCOVIDDEATHPLUS1YRD1** | **Total Frequency** |
| **1** | 0 | 1951250 |
| **2** | 1 | 11572 |

**Probability modeled is NONCOVIDDEATHPLUS1YRD1=1.**

| Class Level Information | | | | | | | | | |
|---|---|---|---|---|---|---|---|---|---|
| **Class** | **Value** | **Design Variables** | | | | | | | |
| **agenew** | **18-39** | 0 | 0 | 0 | 0 | 0 | | | |
| | **40-49** | 1 | 0 | 0 | 0 | 0 | | | |
| | **50-59** | 0 | 1 | 0 | 0 | 0 | | | |
| | **60-69** | 0 | 0 | 1 | 0 | 0 | | | |
| | **70-79** | 0 | 0 | 0 | 1 | 0 | | | |
| | **80+** | 0 | 0 | 0 | 0 | 1 | | | |
| **firstdose** | **1. PFIZER** | 1 | | | | | | | |
| | **2. MODERN** | 0 | | | | | | | |
| **sex2** | **F** | 1 | | | | | | | |
| | **M** | 0 | | | | | | | |
| **RACE4** | **BLACK** | 1 | 0 | 0 | | | | | |
| | **OTHERS** | 0 | 1 | 0 | | | | | |
| | **UNKNOWN** | 0 | 0 | 1 | | | | | |
| | **WHITE** | 0 | 0 | 0 | | | | | |
| **ETHNICITY3** | **HISPANIC** | 1 | 0 | | | | | | |
| | **NOT HISPANIC** | 0 | 0 | | | | | | |
| | **UNKNOWN** | 0 | 1 | | | | | | |
| **Vaccinationsite** | **CHD OR DOH** | 1 | 0 | 0 | 0 | 0 | 0 | 0 | 0 |
| | **DOCTOR CLINIC** | 0 | 1 | 0 | 0 | 0 | 0 | 0 | 0 |

#### The LOGISTIC Procedure

| Class Level Information | | | | | | | | | |
|---|---|---|---|---|---|---|---|---|---|
| **Class** | **Value** | **Design Variables** | | | | | | | |
| | **FIRE RESQUE OR EMS** | 0 | 0 | 1 | 0 | 0 | 0 | 0 | 0 |
| | **FQHC OR RHC OR CHC** | 0 | 0 | 0 | 1 | 0 | 0 | 0 | 0 |
| | **HOSPITAL CLINIC** | 0 | 0 | 0 | 0 | 1 | 0 | 0 | 0 |
| | **HOSPITAL ER** | 0 | 0 | 0 | 0 | 0 | 1 | 0 | 0 |
| | **OTHER PHARMACIES** | 0 | 0 | 0 | 0 | 0 | 0 | 1 | 0 |
| | **OTHERS OR MISC** | 0 | 0 | 0 | 0 | 0 | 0 | 0 | 1 |
| | **PUBLIX OR CVS OR WALGREENS** | 0 | 0 | 0 | 0 | 0 | 0 | 0 | 0 |

| Model Convergence Status |
|---|
| Convergence criterion (GCONV=1E-8) satisfied. |

| Model Fit Statistics | | |
|---|---|---|
| **Criterion** | **Intercept Only** | **Intercept and Covariates** |
| **AIC** | 141888.53 | 121723.02 |
| **SC** | 141901.02 | 122022.78 |
| **-2 Log L** | 141886.53 | 121675.02 |

| Testing Global Null Hypothesis: BETA=0 | | | |
|---|---|---|---|
| **Test** | **Chi-Square** | **DF** | **Pr > ChiSq** |
| **Likelihood Ratio** | 20211.5060 | 23 | <.0001 |
| **Score** | 30637.3977 | 23 | <.0001 |
| **Wald** | 17101.5501 | 23 | <.0001 |

| Type 3 Analysis of Effects | | | |
|---|---|---|---|
| **Effect** | **DF** | **Wald Chi-Square** | **Pr > ChiSq** |
| **firstdose** | 1 | 247.2064 | <.0001 |
| **agenew** | 5 | 5421.0513 | <.0001 |
| **sex2** | 1 | 309.0613 | <.0001 |
| **RACE4** | 3 | 193.2859 | <.0001 |
| **ETHNICITY3** | 2 | 1403.6186 | <.0001 |
| **decile_sviscore1** | 1 | 145.0864 | <.0001 |
| **decile_sviscore4** | 1 | 4.1666 | 0.0412 |
| **mortalityratepct** | 1 | 615.2576 | <.0001 |
| **Vaccinationsite** | 8 | 1373.9141 | <.0001 |

#### The LOGISTIC Procedure

| Analysis of Maximum Likelihood Estimates | | | | | | | |
|---|---|---|---|---|---|---|---|
| Parameter | | DF | Estimate | Standard Error | Wald Chi-Square | Pr > ChiSq | Label |
| Intercept | | 1 | -7.8480 | 0.0679 | 13348.3801 | <.0001 | Intercept: NONCOVIDDEATHPLUS1YRD1=1 |
| firstdose | 1. PFIZER | 1 | 0.2993 | 0.0190 | 247.2064 | <.0001 | firstdose 1. PFIZER |
| agenew | 40-49 | 1 | 0.8686 | 0.0849 | 104.6342 | <.0001 | agenew 40-49 |
| agenew | 50-59 | 1 | 1.6179 | 0.0707 | 524.3195 | <.0001 | agenew 50-59 |
| agenew | 60-69 | 1 | 2.4648 | 0.0666 | 1368.8286 | <.0001 | agenew 60-69 |
| agenew | 70-79 | 1 | 3.2521 | 0.0666 | 2385.7976 | <.0001 | agenew 70-79 |
| agenew | 80+ | 1 | 3.9116 | 0.0719 | 2956.9870 | <.0001 | agenew 80+ |
| sex2 | F | 1 | -0.3394 | 0.0193 | 309.0613 | <.0001 | sex2 F |
| RACE4 | BLACK | 1 | -0.00516 | 0.0457 | 0.0127 | 0.9101 | RACE4 BLACK |
| RACE4 | OTHERS | 1 | -0.5072 | 0.0365 | 192.8481 | <.0001 | RACE4 OTHERS |
| RACE4 | UNKNOWN | 1 | -0.0578 | 0.0545 | 1.1234 | 0.2892 | RACE4 UNKNOWN |
| ETHNICITY3 | HISPANIC | 1 | -0.5351 | 0.0364 | 216.3660 | <.0001 | ETHNICITY3 HISPANIC |
| ETHNICITY3 | UNKNOWN | 1 | 0.9564 | 0.0299 | 1024.6355 | <.0001 | ETHNICITY3 UNKNOWN |
| decile_sviscore1 | | 1 | 0.0503 | 0.00418 | 145.0864 | <.0001 | Rank for Variable SVISCORE1 |
| decile_sviscore4 | | 1 | 0.00802 | 0.00393 | 4.1666 | 0.0412 | Rank for Variable SVISCORE4 |
| mortalityratepct | | 1 | 0.0788 | 0.00318 | 615.2576 | <.0001 | |
| Vaccinationsite | CHD OR DOH | 1 | -0.2981 | 0.0279 | 114.5193 | <.0001 | Vaccinationsite CHD OR DOH |
| Vaccinationsite | DOCTOR CLINIC | 1 | 0.1642 | 0.0257 | 40.9655 | <.0001 | Vaccinationsite DOCTOR CLINIC |
| Vaccinationsite | FIRE RESQUE OR EMS | 1 | -0.2741 | 0.0553 | 24.5850 | <.0001 | Vaccinationsite FIRE RESQUE OR EMS |
| Vaccinationsite | FQHC OR RHC OR CHC | 1 | -0.3178 | 0.0878 | 13.0987 | 0.0003 | Vaccinationsite FQHC OR RHC OR CHC |
| Vaccinationsite | HOSPITAL CLINIC | 1 | 0.0729 | 0.0387 | 3.5458 | 0.0597 | Vaccinationsite HOSPITAL CLINIC |
| Vaccinationsite | HOSPITAL ER | 1 | 0.0172 | 0.1691 | 0.0104 | 0.9188 | Vaccinationsite HOSPITAL ER |
| Vaccinationsite | OTHER PHARMACIES | 1 | 0.3574 | 0.0560 | 40.7907 | <.0001 | Vaccinationsite OTHER PHARMACIES |
| Vaccinationsite | OTHERS OR MISC | 1 | -1.4954 | 0.0472 | 1002.4813 | <.0001 | Vaccinationsite OTHERS OR MISC |

| Odds Ratio Estimates | | | |
|---|---|---|---|
| Effect | Point Estimate | 95% Wald Confidence Limits | |
| firstdose 1. PFIZER vs 2. MODERN | 1.349 | 1.300 | 1.400 |
| agenew 40-49 vs 18-39 | 2.383 | 2.018 | 2.815 |
| agenew 50-59 vs 18-39 | 5.042 | 4.390 | 5.791 |
| agenew 60-69 vs 18-39 | 11.761 | 10.321 | 13.401 |
| agenew 70-79 vs 18-39 | 25.843 | 22.682 | 29.446 |
| agenew 80+ vs 18-39 | 49.977 | 43.406 | 57.544 |
| sex2 F vs M | 0.712 | 0.686 | 0.740 |

#### The LOGISTIC Procedure

| Odds Ratio Estimates | | | |
|---|---|---|---|
| Effect | Point Estimate | 95% Wald Confidence Limits | |
| RACE4 BLACK vs WHITE | 0.995 | 0.910 | 1.088 |
| RACE4 OTHERS vs WHITE | 0.602 | 0.561 | 0.647 |
| RACE4 UNKNOWN vs WHITE | 0.944 | 0.848 | 1.050 |
| ETHNICITY3 HISPANIC vs NOT HISPANIC | 0.586 | 0.545 | 0.629 |
| ETHNICITY3 UNKNOWN vs NOT HISPANIC | 2.602 | 2.454 | 2.759 |
| decile_sviscore1 | 1.052 | 1.043 | 1.060 |
| decile_sviscore4 | 1.008 | 1.000 | 1.016 |
| mortalityratepct | 1.082 | 1.075 | 1.089 |
| Vaccinationsite CHD OR DOH vs PUBLIX OR CVS OR WALGREENS | 0.742 | 0.703 | 0.784 |
| Vaccinationsite DOCTOR CLINIC vs PUBLIX OR CVS OR WALGREENS | 1.178 | 1.121 | 1.239 |
| Vaccinationsite FIRE RESQUE OR EMS vs PUBLIX OR CVS OR WALGREENS | 0.760 | 0.682 | 0.847 |
| Vaccinationsite FQHC OR RHC OR CHC vs PUBLIX OR CVS OR WALGREENS | 0.728 | 0.613 | 0.864 |
| Vaccinationsite HOSPITAL CLINIC vs PUBLIX OR CVS OR WALGREENS | 1.076 | 0.997 | 1.161 |
| Vaccinationsite HOSPITAL ER vs PUBLIX OR CVS OR WALGREENS | 1.017 | 0.730 | 1.417 |
| Vaccinationsite OTHER PHARMACIES vs PUBLIX OR CVS OR WALGREENS | 1.430 | 1.281 | 1.595 |
| Vaccinationsite OTHERS OR MISC vs PUBLIX OR CVS OR WALGREENS | 0.224 | 0.204 | 0.246 |

| Association of Predicted Probabilities and Observed Responses | | | |
|---|---|---|---|
| Percent Concordant | 83.3 | Somers' D | 0.666 |
| Percent Discordant | 16.7 | Gamma | 0.666 |
| Percent Tied | 0.0 | Tau-a | 0.008 |
| Pairs | 22579865000 | c | 0.833 |

### NON COVID + deaths adjusted by Age

#### The LOGISTIC Procedure

##### agenew=18-39 years

| Model Information | |
|---|---|
| **Data Set** | WORK.MATCHEDCOHORT |
| **Response Variable** | NONCOVIDDEATHPLUS1YRD1 |
| **Number of Response Levels** | 2 |
| **Model** | binary logit |
| **Optimization Technique** | Fisher's scoring |

| | |
|---|---|
| **Number of Observations Read** | 492484 |
| **Number of Observations Used** | 492484 |

| Response Profile | | |
|---|---|---|
| **Ordered Value** | **NONCOVIDDEATHPLUS1YRD1** | **Total Frequency** |
| **1** | 0 | 492230 |
| **2** | 1 | 254 |

**Probability modeled is NONCOVIDDEATHPLUS1YRD1=1.**

| Class Level Information | | | | | | | | | |
|---|---|---|---|---|---|---|---|---|---|
| **Class** | **Value** | **Design Variables** | | | | | | | |
| **agenew** | **18-39 years** | | | | | | | | |
| **firstdose** | **1. PFIZER** | 1 | | | | | | | |
| | **2. MODERN** | 0 | | | | | | | |
| **sex2** | **F** | 1 | | | | | | | |
| | **M** | 0 | | | | | | | |
| **RACE4** | **BLACK** | 1 | 0 | 0 | | | | | |
| | **OTHERS** | 0 | 1 | 0 | | | | | |
| | **UNKNOWN** | 0 | 0 | 1 | | | | | |
| | **WHITE** | 0 | 0 | 0 | | | | | |
| **ETHNICITY3** | **HISPANIC** | 1 | 0 | | | | | | |
| | **NOT HISPANIC** | 0 | 0 | | | | | | |
| | **UNKNOWN** | 0 | 1 | | | | | | |
| **Vaccinationsite** | **CHD OR DOH** | 1 | 0 | 0 | 0 | 0 | 0 | 0 | 0 |
| | **DOCTOR CLINIC** | 0 | 1 | 0 | 0 | 0 | 0 | 0 | 0 |
| | **FIRE RESQUE OR EMS** | 0 | 0 | 1 | 0 | 0 | 0 | 0 | 0 |
| | **FQHC OR RHC OR CHC** | 0 | 0 | 0 | 1 | 0 | 0 | 0 | 0 |
| | **HOSPITAL CLINIC** | 0 | 0 | 0 | 0 | 1 | 0 | 0 | 0 |
| | **HOSPITAL ER** | 0 | 0 | 0 | 0 | 0 | 1 | 0 | 0 |

#### The LOGISTIC Procedure

##### agenew=18-39 years

| Class Level Information | | | | | | | | | |
|---|---|---|---|---|---|---|---|---|---|
| Class | Value | Design Variables | | | | | | | |
| | OTHER PHARMACIES | 0 | 0 | 0 | 0 | 0 | 0 | 1 | 0 |
| | OTHERS OR MISC | 0 | 0 | 0 | 0 | 0 | 0 | 0 | 1 |
| | PUBLIX OR CVS OR WALGREENS | 0 | 0 | 0 | 0 | 0 | 0 | 0 | 0 |

| Model Convergence Status |
|---|
| Convergence criterion (GCONV=1E-8) satisfied. |

| Model Fit Statistics | | |
|---|---|---|
| Criterion | Intercept Only | Intercept and Covariates |
| AIC | 4355.370 | 4250.674 |
| SC | 4366.477 | 4461.711 |
| -2 Log L | 4353.370 | 4212.674 |

| Testing Global Null Hypothesis: BETA=0 | | | |
|---|---|---|---|
| Test | Chi-Square | DF | Pr > ChiSq |
| Likelihood Ratio | 140.6959 | 18 | <.0001 |
| Score | 141.3307 | 18 | <.0001 |
| Wald | 131.7265 | 18 | <.0001 |

| Type 3 Analysis of Effects | | | |
|---|---|---|---|
| Effect | DF | Wald Chi-Square | Pr > ChiSq |
| firstdose | 1 | 4.5217 | 0.0335 |
| agenew | 0 | . | . |
| sex2 | 1 | 51.4660 | <.0001 |
| RACE4 | 3 | 5.5630 | 0.1349 |
| ETHNICITY3 | 2 | 40.2342 | <.0001 |
| decile_sviscore1 | 1 | 13.0974 | 0.0003 |
| decile_sviscore4 | 1 | 0.0187 | 0.8912 |
| mortalityratepct | 1 | 0.0008 | 0.9777 |
| Vaccinationsite | 8 | 12.2992 | 0.1383 |

#### The LOGISTIC Procedure

**agenew=18-39 years**

| Analysis of Maximum Likelihood Estimates | | | | | | | |
|---|---|---|---|---|---|---|---|
| **Parameter** | | **DF** | **Estimate** | **Standard Error** | **Wald Chi-Square** | **Pr > ChiSq** | **Label** |
| **Intercept** | | 1 | -7.6729 | 0.1734 | 1958.9116 | <.0001 | Intercept: NONCOVIDDEATHPLUS1YRD1=1 |
| **firstdose** | **1. PFIZER** | 1 | 0.2694 | 0.1267 | 4.5217 | 0.0335 | firstdose 1. PFIZER |
| **sex2** | **F** | 1 | -0.9736 | 0.1357 | 51.4660 | <.0001 | sex2 F |
| **RACE4** | **BLACK** | 1 | 0.2066 | 0.2051 | 1.0146 | 0.3138 | RACE4 BLACK |
| **RACE4** | **OTHERS** | 1 | -0.0946 | 0.1790 | 0.2796 | 0.5969 | RACE4 OTHERS |
| **RACE4** | **UNKNOWN** | 1 | -0.7239 | 0.3615 | 4.0101 | 0.0452 | RACE4 UNKNOWN |
| **ETHNICITY3** | **HISPANIC** | 1 | -0.7421 | 0.1883 | 15.5317 | <.0001 | ETHNICITY3 HISPANIC |
| **ETHNICITY3** | **UNKNOWN** | 1 | 0.5857 | 0.1597 | 13.4472 | 0.0002 | ETHNICITY3 UNKNOWN |
| **decile_sviscore1** | | 1 | 0.1008 | 0.0278 | 13.0974 | 0.0003 | Rank for Variable SVISCORE1 |
| **decile_sviscore4** | | 1 | -0.00361 | 0.0264 | 0.0187 | 0.8912 | Rank for Variable SVISCORE4 |
| **mortalityratepct** | | 1 | -10.9205 | 390.2 | 0.0008 | 0.9777 | |
| **Vaccinationsite** | **CHD OR DOH** | 1 | -0.2784 | 0.3632 | 0.5874 | 0.4434 | Vaccinationsite CHD OR DOH |
| **Vaccinationsite** | **DOCTOR CLINIC** | 1 | 0.2333 | 0.1817 | 1.6477 | 0.1993 | Vaccinationsite DOCTOR CLINIC |
| **Vaccinationsite** | **FIRE RESQUE OR EMS** | 1 | -10.6949 | 442.5 | 0.0006 | 0.9807 | Vaccinationsite FIRE RESQUE OR EMS |
| **Vaccinationsite** | **FQHC OR RHC OR CHC** | 1 | 0.1944 | 0.5874 | 0.1095 | 0.7407 | Vaccinationsite FQHC OR RHC OR CHC |
| **Vaccinationsite** | **HOSPITAL CLINIC** | 1 | -0.3649 | 0.4553 | 0.6423 | 0.4229 | Vaccinationsite HOSPITAL CLINIC |
| **Vaccinationsite** | **HOSPITAL ER** | 1 | 0.7071 | 1.0046 | 0.4955 | 0.4815 | Vaccinationsite HOSPITAL ER |
| **Vaccinationsite** | **OTHER PHARMACIES** | 1 | 0.2004 | 0.2579 | 0.6040 | 0.4370 | Vaccinationsite OTHER PHARMACIES |
| **Vaccinationsite** | **OTHERS OR MISC** | 1 | -1.4027 | 0.5071 | 7.6509 | 0.0057 | Vaccinationsite OTHERS OR MISC |

| Odds Ratio Estimates | | | |
|---|---|---|---|
| **Effect** | **Point Estimate** | **95% Wald Confidence Limits** | |
| **firstdose 1. PFIZER vs 2. MODERN** | 1.309 | 1.021 | 1.678 |
| **sex2 F vs M** | 0.378 | 0.289 | 0.493 |
| **RACE4 BLACK vs WHITE** | 1.230 | 0.822 | 1.838 |
| **RACE4 OTHERS vs WHITE** | 0.910 | 0.641 | 1.292 |
| **RACE4 UNKNOWN vs WHITE** | 0.485 | 0.239 | 0.985 |
| **ETHNICITY3 HISPANIC vs NOT HISPANIC** | 0.476 | 0.329 | 0.689 |
| **ETHNICITY3 UNKNOWN vs NOT HISPANIC** | 1.796 | 1.313 | 2.457 |
| **decile_sviscore1** | 1.106 | 1.047 | 1.168 |
| **decile_sviscore4** | 0.996 | 0.946 | 1.049 |
| **mortalityratepct** | <0.001 | <0.001 | >999.999 |
| **Vaccinationsite CHD OR DOH vs PUBLIX OR CVS OR WALGREENS** | 0.757 | 0.371 | 1.543 |

#### The LOGISTIC Procedure

##### agenew=18-39 years

| Odds Ratio Estimates | | | |
|---|---|---|---|
| Effect | Point Estimate | 95% Wald Confidence Limits | |
| Vaccinationsite DOCTOR CLINIC vs PUBLIX OR CVS OR WALGREENS | 1.263 | 0.884 | 1.803 |
| Vaccinationsite FIRE RESQUE OR EMS vs PUBLIX OR CVS OR WALGREENS | <0.001 | <0.001 | >999.999 |
| Vaccinationsite FQHC OR RHC OR CHC vs PUBLIX OR CVS OR WALGREENS | 1.215 | 0.384 | 3.841 |
| Vaccinationsite HOSPITAL CLINIC vs PUBLIX OR CVS OR WALGREENS | 0.694 | 0.284 | 1.695 |
| Vaccinationsite HOSPITAL ER vs PUBLIX OR CVS OR WALGREENS | 2.028 | 0.283 | 14.528 |
| Vaccinationsite OTHER PHARMACIES vs PUBLIX OR CVS OR WALGREENS | 1.222 | 0.737 | 2.026 |
| Vaccinationsite OTHERS OR MISC vs PUBLIX OR CVS OR WALGREENS | 0.246 | 0.091 | 0.664 |

| Association of Predicted Probabilities and Observed Responses | | | |
|---|---|---|---|
| Percent Concordant | 71.0 | Somers' D | 0.419 |
| Percent Discordant | 29.0 | Gamma | 0.420 |
| Percent Tied | 0.0 | Tau-a | 0.000 |
| Pairs | 125026420 | c | 0.710 |

#### The LOGISTIC Procedure

**agenew=40-59 years**

| Model Information | |
|---|---|
| **Data Set** | WORK.MATCHEDCOHORT |
| **Response Variable** | NONCOVIDDEATHPLUS1YRD1 |
| **Number of Response Levels** | 2 |
| **Model** | binary logit |
| **Optimization Technique** | Fisher's scoring |

| | |
|---|---|
| **Number of Observations Read** | 634102 |
| **Number of Observations Used** | 634102 |

| Response Profile | | |
|---|---|---|
| **Ordered Value** | **NONCOVIDDEATHPLUS1YRD1** | **Total Frequency** |
| **1** | 0 | 632818 |
| **2** | 1 | 1284 |

**Probability modeled is NONCOVIDDEATHPLUS1YRD1=1.**

| Class Level Information | | | | | | | | | |
|---|---|---|---|---|---|---|---|---|---|
| **Class** | **Value** | **Design Variables** | | | | | | | |
| **agenew** | **40-59 years** | | | | | | | | |
| **firstdose** | **1. PFIZER** | 1 | | | | | | | |
| | **2. MODERN** | 0 | | | | | | | |
| **sex2** | **F** | 1 | | | | | | | |
| | **M** | 0 | | | | | | | |
| **RACE4** | **BLACK** | 1 | 0 | 0 | | | | | |
| | **OTHERS** | 0 | 1 | 0 | | | | | |
| | **UNKNOWN** | 0 | 0 | 1 | | | | | |
| | **WHITE** | 0 | 0 | 0 | | | | | |
| **ETHNICITY3** | **HISPANIC** | 1 | 0 | | | | | | |
| | **NOT HISPANIC** | 0 | 0 | | | | | | |
| | **UNKNOWN** | 0 | 1 | | | | | | |
| **Vaccinationsite** | **CHD OR DOH** | 1 | 0 | 0 | 0 | 0 | 0 | 0 | 0 |
| | **DOCTOR CLINIC** | 0 | 1 | 0 | 0 | 0 | 0 | 0 | 0 |
| | **FIRE RESQUE OR EMS** | 0 | 0 | 1 | 0 | 0 | 0 | 0 | 0 |
| | **FQHC OR RHC OR CHC** | 0 | 0 | 0 | 1 | 0 | 0 | 0 | 0 |
| | **HOSPITAL CLINIC** | 0 | 0 | 0 | 0 | 1 | 0 | 0 | 0 |
| | **HOSPITAL ER** | 0 | 0 | 0 | 0 | 0 | 1 | 0 | 0 |

#### The LOGISTIC Procedure

**agenew=40-59 years**

| Class Level Information | | | | | | | | | |
|---|---|---|---|---|---|---|---|---|---|
| Class | Value | Design Variables | | | | | | | |
| | OTHER PHARMACIES | 0 | 0 | 0 | 0 | 0 | 0 | 1 | 0 |
| | OTHERS OR MISC | 0 | 0 | 0 | 0 | 0 | 0 | 0 | 1 |
| | PUBLIX OR CVS OR WALGREENS | 0 | 0 | 0 | 0 | 0 | 0 | 0 | 0 |

| Model Convergence Status |
|---|
| Convergence criterion (GCONV=1E-8) satisfied. |

| Model Fit Statistics | | |
|---|---|---|
| Criterion | Intercept Only | Intercept and Covariates |
| AIC | 18494.724 | 17807.274 |
| SC | 18506.084 | 18023.114 |
| -2 Log L | 18492.724 | 17769.274 |

| Testing Global Null Hypothesis: BETA=0 | | | |
|---|---|---|---|
| Test | Chi-Square | DF | Pr > ChiSq |
| Likelihood Ratio | 723.4498 | 18 | <.0001 |
| Score | 736.6710 | 18 | <.0001 |
| Wald | 693.3421 | 18 | <.0001 |

| Type 3 Analysis of Effects | | | |
|---|---|---|---|
| Effect | DF | Wald Chi-Square | Pr > ChiSq |
| firstdose | 1 | 4.8331 | 0.0279 |
| agenew | 0 | . | . |
| sex2 | 1 | 33.1082 | <.0001 |
| RACE4 | 3 | 11.8351 | 0.0080 |
| ETHNICITY3 | 2 | 141.7397 | <.0001 |
| decile_sviscore1 | 1 | 69.5370 | <.0001 |
| decile_sviscore4 | 1 | 0.0391 | 0.8432 |
| mortalityratepct | 1 | 154.7289 | <.0001 |
| Vaccinationsite | 8 | 129.5430 | <.0001 |

#### The LOGISTIC Procedure

**agenew=40-59 years**

| Analysis of Maximum Likelihood Estimates | | | | | | | |
|---|---|---|---|---|---|---|---|
| **Parameter** | | **DF** | **Estimate** | **Standard Error** | **Wald Chi-Square** | **Pr > ChiSq** | **Label** |
| **Intercept** | | 1 | -6.9546 | 0.0884 | 6189.9793 | <.0001 | Intercept: NONCOVIDDEATHPLUS1YRD1=1 |
| **firstdose** | **1. PFIZER** | 1 | 0.1231 | 0.0560 | 4.8331 | 0.0279 | firstdose 1. PFIZER |
| **sex2** | **F** | 1 | -0.3481 | 0.0605 | 33.1082 | <.0001 | sex2 F |
| **RACE4** | **BLACK** | 1 | -0.0200 | 0.0962 | 0.0431 | 0.8356 | RACE4 BLACK |
| **RACE4** | **OTHERS** | 1 | -0.3708 | 0.1110 | 11.1582 | 0.0008 | RACE4 OTHERS |
| **RACE4** | **UNKNOWN** | 1 | -0.1835 | 0.1481 | 1.5338 | 0.2155 | RACE4 UNKNOWN |
| **ETHNICITY3** | **HISPANIC** | 1 | -0.7103 | 0.0955 | 55.3104 | <.0001 | ETHNICITY3 HISPANIC |
| **ETHNICITY3** | **UNKNOWN** | 1 | 0.6274 | 0.0851 | 54.3914 | <.0001 | ETHNICITY3 UNKNOWN |
| **decile_sviscore1** | | 1 | 0.1053 | 0.0126 | 69.5370 | <.0001 | Rank for Variable SVISCORE1 |
| **decile_sviscore4** | | 1 | 0.00236 | 0.0119 | 0.0391 | 0.8432 | Rank for Variable SVISCORE4 |
| **mortalityratepct** | | 1 | 0.7415 | 0.0596 | 154.7289 | <.0001 | |
| **Vaccinationsite** | **CHD OR DOH** | 1 | -0.0633 | 0.1598 | 0.1567 | 0.6922 | Vaccinationsite CHD OR DOH |
| **Vaccinationsite** | **DOCTOR CLINIC** | 1 | 0.2694 | 0.0738 | 13.3152 | 0.0003 | Vaccinationsite DOCTOR CLINIC |
| **Vaccinationsite** | **FIRE RESQUE OR EMS** | 1 | -0.2772 | 0.7091 | 0.1528 | 0.6959 | Vaccinationsite FIRE RESQUE OR EMS |
| **Vaccinationsite** | **FQHC OR RHC OR CHC** | 1 | 0.3200 | 0.2105 | 2.3115 | 0.1284 | Vaccinationsite FQHC OR RHC OR CHC |
| **Vaccinationsite** | **HOSPITAL CLINIC** | 1 | 0.7607 | 0.1021 | 55.5115 | <.0001 | Vaccinationsite HOSPITAL CLINIC |
| **Vaccinationsite** | **HOSPITAL ER** | 1 | 0.1800 | 0.5024 | 0.1284 | 0.7201 | Vaccinationsite HOSPITAL ER |
| **Vaccinationsite** | **OTHER PHARMACIES** | 1 | 0.3920 | 0.1116 | 12.3343 | 0.0004 | Vaccinationsite OTHER PHARMACIES |
| **Vaccinationsite** | **OTHERS OR MISC** | 1 | -1.1357 | 0.1683 | 45.5616 | <.0001 | Vaccinationsite OTHERS OR MISC |

| Odds Ratio Estimates | | | |
|---|---|---|---|
| **Effect** | **Point Estimate** | **95% Wald Confidence Limits** | |
| **firstdose 1. PFIZER vs 2. MODERN** | 1.131 | 1.013 | 1.262 |
| **sex2 F vs M** | 0.706 | 0.627 | 0.795 |
| **RACE4 BLACK vs WHITE** | 0.980 | 0.812 | 1.184 |
| **RACE4 OTHERS vs WHITE** | 0.690 | 0.555 | 0.858 |
| **RACE4 UNKNOWN vs WHITE** | 0.832 | 0.623 | 1.113 |
| **ETHNICITY3 HISPANIC vs NOT HISPANIC** | 0.491 | 0.408 | 0.593 |
| **ETHNICITY3 UNKNOWN vs NOT HISPANIC** | 1.873 | 1.585 | 2.213 |
| **decile_sviscore1** | 1.111 | 1.084 | 1.139 |
| **decile_sviscore4** | 1.002 | 0.979 | 1.026 |
| **mortalityratepct** | 2.099 | 1.868 | 2.359 |
| **Vaccinationsite CHD OR DOH vs PUBLIX OR CVS OR WALGREENS** | 0.939 | 0.686 | 1.284 |

#### The LOGISTIC Procedure

**agenew=40-59 years**

| Odds Ratio Estimates | | | |
|---|---|---|---|
| Effect | Point Estimate | 95% Wald Confidence Limits | |
| Vaccinationsite DOCTOR CLINIC vs PUBLIX OR CVS OR WALGREENS | 1.309 | 1.133 | 1.513 |
| Vaccinationsite FIRE RESQUE OR EMS vs PUBLIX OR CVS OR WALGREENS | 0.758 | 0.189 | 3.042 |
| Vaccinationsite FQHC OR RHC OR CHC vs PUBLIX OR CVS OR WALGREENS | 1.377 | 0.912 | 2.080 |
| Vaccinationsite HOSPITAL CLINIC vs PUBLIX OR CVS OR WALGREENS | 2.140 | 1.752 | 2.614 |
| Vaccinationsite HOSPITAL ER vs PUBLIX OR CVS OR WALGREENS | 1.197 | 0.447 | 3.205 |
| Vaccinationsite OTHER PHARMACIES vs PUBLIX OR CVS OR WALGREENS | 1.480 | 1.189 | 1.842 |
| Vaccinationsite OTHERS OR MISC vs PUBLIX OR CVS OR WALGREENS | 0.321 | 0.231 | 0.447 |

| Association of Predicted Probabilities and Observed Responses | | | |
|---|---|---|---|
| Percent Concordant | 71.0 | Somers' D | 0.421 |
| Percent Discordant | 28.9 | Gamma | 0.421 |
| Percent Tied | 0.0 | Tau-a | 0.002 |
| Pairs | 812538312 | c | 0.711 |

#### The LOGISTIC Procedure

**agenew=60+ years**

| Model Information | |
|---|---|
| **Data Set** | WORK.MATCHEDCOHORT |
| **Response Variable** | NONCOVIDDEATHPLUS1YRD1 |
| **Number of Response Levels** | 2 |
| **Model** | binary logit |
| **Optimization Technique** | Fisher's scoring |

| | |
|---|---|
| **Number of Observations Read** | 836236 |
| **Number of Observations Used** | 836236 |

| Response Profile | | |
|---|---|---|
| **Ordered Value** | **NONCOVIDDEATHPLUS1YRD1** | **Total Frequency** |
| **1** | 0 | 826202 |
| **2** | 1 | 10034 |

**Probability modeled is NONCOVIDDEATHPLUS1YRD1=1.**

| Class Level Information | | | | | | | | | |
|---|---|---|---|---|---|---|---|---|---|
| **Class** | **Value** | **Design Variables** | | | | | | | |
| **agenew** | **60+ years** | | | | | | | | |
| **firstdose** | **1. PFIZER** | 1 | | | | | | | |
| | **2. MODERN** | 0 | | | | | | | |
| **sex2** | **F** | 1 | | | | | | | |
| | **M** | 0 | | | | | | | |
| **RACE4** | **BLACK** | 1 | 0 | 0 | | | | | |
| | **OTHERS** | 0 | 1 | 0 | | | | | |
| | **UNKNOWN** | 0 | 0 | 1 | | | | | |
| | **WHITE** | 0 | 0 | 0 | | | | | |
| **ETHNICITY3** | **HISPANIC** | 1 | 0 | | | | | | |
| | **NOT HISPANIC** | 0 | 0 | | | | | | |
| | **UNKNOWN** | 0 | 1 | | | | | | |
| **Vaccinationsite** | **CHD OR DOH** | 1 | 0 | 0 | 0 | 0 | 0 | 0 | 0 |
| | **DOCTOR CLINIC** | 0 | 1 | 0 | 0 | 0 | 0 | 0 | 0 |
| | **FIRE RESQUE OR EMS** | 0 | 0 | 1 | 0 | 0 | 0 | 0 | 0 |
| | **FQHC OR RHC OR CHC** | 0 | 0 | 0 | 1 | 0 | 0 | 0 | 0 |
| | **HOSPITAL CLINIC** | 0 | 0 | 0 | 0 | 1 | 0 | 0 | 0 |
| | **HOSPITAL ER** | 0 | 0 | 0 | 0 | 0 | 1 | 0 | 0 |

#### The LOGISTIC Procedure

**agenew=60+ years**

| Class Level Information | | | | | | | | | |
|---|---|---|---|---|---|---|---|---|---|
| **Class** | **Value** | **Design Variables** | | | | | | | |
| | **OTHER PHARMACIES** | 0 | 0 | 0 | 0 | 0 | 0 | 1 | 0 |
| | **OTHERS OR MISC** | 0 | 0 | 0 | 0 | 0 | 0 | 0 | 1 |
| | **PUBLIX OR CVS OR WALGREENS** | 0 | 0 | 0 | 0 | 0 | 0 | 0 | 0 |

| Model Convergence Status |
|---|
| Convergence criterion (GCONV=1E-8) satisfied. |

| Model Fit Statistics | | |
|---|---|---|
| **Criterion** | **Intercept Only** | **Intercept and Covariates** |
| **AIC** | 108708.51 | 101172.65 |
| **SC** | 108720.14 | 101393.75 |
| **-2 Log L** | 108706.51 | 101134.65 |

| Testing Global Null Hypothesis: BETA=0 | | | |
|---|---|---|---|
| **Test** | **Chi-Square** | **DF** | **Pr > ChiSq** |
| **Likelihood Ratio** | 7571.8542 | 18 | <.0001 |
| **Score** | 11608.8217 | 18 | <.0001 |
| **Wald** | 8122.4245 | 18 | <.0001 |

| Type 3 Analysis of Effects | | | |
|---|---|---|---|
| **Effect** | **DF** | **Wald Chi-Square** | **Pr > ChiSq** |
| **firstdose** | 1 | 248.9706 | <.0001 |
| **agenew** | 0 | . | . |
| **sex2** | 1 | 98.2073 | <.0001 |
| **RACE4** | 3 | 197.2481 | <.0001 |
| **ETHNICITY3** | 2 | 1357.5716 | <.0001 |
| **decile_sviscore1** | 1 | 43.7782 | <.0001 |
| **decile_sviscore4** | 1 | 8.0417 | 0.0046 |
| **mortalityratepct** | 1 | 5242.0343 | <.0001 |
| **Vaccinationsite** | 8 | 1111.5799 | <.0001 |

#### The LOGISTIC Procedure

##### agenew=60+ years

| Analysis of Maximum Likelihood Estimates | | | | | | | |
|---|---|---|---|---|---|---|---|
| Parameter | | DF | Estimate | Standard Error | Wald Chi-Square | Pr > ChiSq | Label |
| Intercept | | 1 | -5.0735 | 0.0313 | 26201.3531 | <.0001 | Intercept: NONCOVIDDEATHPLUS1YRD1=1 |
| firstdose | 1. PFIZER | 1 | 0.3245 | 0.0206 | 248.9706 | <.0001 | firstdose 1. PFIZER |
| sex2 | F | 1 | -0.2038 | 0.0206 | 98.2073 | <.0001 | sex2 F |
| RACE4 | BLACK | 1 | -0.1942 | 0.0537 | 13.0906 | 0.0003 | RACE4 BLACK |
| RACE4 | OTHERS | 1 | -0.5743 | 0.0420 | 187.2025 | <.0001 | RACE4 OTHERS |
| RACE4 | UNKNOWN | 1 | 0.00368 | 0.0595 | 0.0038 | 0.9507 | RACE4 UNKNOWN |
| ETHNICITY3 | HISPANIC | 1 | -0.5699 | 0.0421 | 183.4845 | <.0001 | ETHNICITY3 HISPANIC |
| ETHNICITY3 | UNKNOWN | 1 | 1.0539 | 0.0325 | 1052.8073 | <.0001 | ETHNICITY3 UNKNOWN |
| decile_sviscore1 | | 1 | 0.0298 | 0.00450 | 43.7782 | <.0001 | Rank for Variable SVISCORE1 |
| decile_sviscore4 | | 1 | 0.0120 | 0.00424 | 8.0417 | 0.0046 | Rank for Variable SVISCORE4 |
| mortalityratepct | | 1 | 0.1626 | 0.00225 | 5242.0343 | <.0001 | |
| Vaccinationsite | CHD OR DOH | 1 | -0.1585 | 0.0284 | 31.2204 | <.0001 | Vaccinationsite CHD OR DOH |
| Vaccinationsite | DOCTOR CLINIC | 1 | 0.2537 | 0.0276 | 84.3014 | <.0001 | Vaccinationsite DOCTOR CLINIC |
| Vaccinationsite | FIRE RESQUE OR EMS | 1 | -0.0234 | 0.0559 | 0.1746 | 0.6760 | Vaccinationsite FIRE RESQUE OR EMS |
| Vaccinationsite | FQHC OR RHC OR CHC | 1 | -0.3009 | 0.0977 | 9.4832 | 0.0021 | Vaccinationsite FQHC OR RHC OR CHC |
| Vaccinationsite | HOSPITAL CLINIC | 1 | 0.0589 | 0.0424 | 1.9298 | 0.1648 | Vaccinationsite HOSPITAL CLINIC |
| Vaccinationsite | HOSPITAL ER | 1 | 0.0551 | 0.1821 | 0.0914 | 0.7624 | Vaccinationsite HOSPITAL ER |
| Vaccinationsite | OTHER PHARMACIES | 1 | 0.2871 | 0.0669 | 18.4161 | <.0001 | Vaccinationsite OTHER PHARMACIES |
| Vaccinationsite | OTHERS OR MISC | 1 | -1.4002 | 0.0496 | 798.4651 | <.0001 | Vaccinationsite OTHERS OR MISC |

| Odds Ratio Estimates | | | |
|---|---|---|---|
| Effect | Point Estimate | 95% Wald Confidence Limits | |
| firstdose 1. PFIZER vs 2. MODERN | 1.383 | 1.329 | 1.440 |
| sex2 F vs M | 0.816 | 0.783 | 0.849 |
| RACE4 BLACK vs WHITE | 0.824 | 0.741 | 0.915 |
| RACE4 OTHERS vs WHITE | 0.563 | 0.519 | 0.611 |
| RACE4 UNKNOWN vs WHITE | 1.004 | 0.893 | 1.128 |
| ETHNICITY3 HISPANIC vs NOT HISPANIC | 0.566 | 0.521 | 0.614 |
| ETHNICITY3 UNKNOWN vs NOT HISPANIC | 2.869 | 2.692 | 3.057 |
| decile_sviscore1 | 1.030 | 1.021 | 1.039 |
| decile_sviscore4 | 1.012 | 1.004 | 1.021 |
| mortalityratepct | 1.177 | 1.171 | 1.182 |
| Vaccinationsite CHD OR DOH vs PUBLIX OR CVS OR WALGREENS | 0.853 | 0.807 | 0.902 |

#### The LOGISTIC Procedure

**agenew=60+ years**

| Odds Ratio Estimates | | | |
|---|---|---|---|
| Effect | Point Estimate | 95% Wald Confidence Limits | |
| Vaccinationsite DOCTOR CLINIC vs PUBLIX OR CVS OR WALGREENS | 1.289 | 1.221 | 1.361 |
| Vaccinationsite FIRE RESQUE OR EMS vs PUBLIX OR CVS OR WALGREENS | 0.977 | 0.876 | 1.090 |
| Vaccinationsite FQHC OR RHC OR CHC vs PUBLIX OR CVS OR WALGREENS | 0.740 | 0.611 | 0.896 |
| Vaccinationsite HOSPITAL CLINIC vs PUBLIX OR CVS OR WALGREENS | 1.061 | 0.976 | 1.153 |
| Vaccinationsite HOSPITAL ER vs PUBLIX OR CVS OR WALGREENS | 1.057 | 0.739 | 1.510 |
| Vaccinationsite OTHER PHARMACIES vs PUBLIX OR CVS OR WALGREENS | 1.333 | 1.169 | 1.519 |
| Vaccinationsite OTHERS OR MISC vs PUBLIX OR CVS OR WALGREENS | 0.247 | 0.224 | 0.272 |

| Association of Predicted Probabilities and Observed Responses | | | |
|---|---|---|---|
| Percent Concordant | 73.2 | Somers' D | 0.465 |
| Percent Discordant | 26.8 | Gamma | 0.465 |
| Percent Tied | 0.0 | Tau-a | 0.011 |
| Pairs | 8290110868 | c | 0.732 |

### Matched cohort Risk of death Dose 1 SUICIDE OR HOMICIDE deaths

#### The LOGISTIC Procedure

| Model Information | |
|---|---|
| **Data Set** | WORK.MATCHEDCOHORT |
| **Response Variable** | suicidehomicide1yrD1 |
| **Number of Response Levels** | 2 |
| **Model** | binary logit |
| **Optimization Technique** | Fisher's scoring |

| | |
|---|---|
| **Number of Observations Read** | 1962822 |
| **Number of Observations Used** | 1962822 |

| Response Profile | | |
|---|---|---|
| **Ordered Value** | **suicidehomicide1yrD1** | **Total Frequency** |
| **1** | 0 | 1962598 |
| **2** | 1 | 224 |

**Probability modeled is suicidehomicide1yrD1=1.**

| Class Level Information | | |
|---|---|---|
| **Class** | **Value** | **Design Variables** |
| **firstdose** | **1. PFIZER** | 1 |
| | **2. MODERN** | 0 |

| Model Convergence Status |
|---|
| Convergence criterion (GCONV=1E-8) satisfied. |

| Model Fit Statistics | | |
|---|---|---|
| **Criterion** | **Intercept Only** | **Intercept and Covariates** |
| **AIC** | 4517.029 | 4516.865 |
| **SC** | 4529.519 | 4541.845 |
| **-2 Log L** | 4515.029 | 4512.865 |

| Testing Global Null Hypothesis: BETA=0 | | | |
|---|---|---|---|
| **Test** | **Chi-Square** | **DF** | **Pr > ChiSq** |
| **Likelihood Ratio** | 2.1644 | 1 | 0.1412 |
| **Score** | 2.1610 | 1 | 0.1416 |
| **Wald** | 2.1539 | 1 | 0.1422 |

### Matched cohort Risk of death Dose 1 SUICIDE OR HOMICIDE deaths

#### The LOGISTIC Procedure

| Type 3 Analysis of Effects | | | |
|---|---|---|---|
| Effect | DF | Wald Chi-Square | Pr > ChiSq |
| **firstdose** | 1 | 2.1539 | 0.1422 |

| Analysis of Maximum Likelihood Estimates | | | | | | | |
|---|---|---|---|---|---|---|---|
| Parameter | | DF | Estimate | Standard Error | Wald Chi-Square | Pr > ChiSq | Label |
| **Intercept** | | 1 | -9.1815 | 0.0995 | 8513.5495 | <.0001 | Intercept: suicidehomicide1yrD1=1 |
| **firstdose** | **1. PFIZER** | 1 | 0.1971 | 0.1343 | 2.1539 | 0.1422 | firstdose 1. PFIZER |

| Odds Ratio Estimates | | | |
|---|---|---|---|
| Effect | Point Estimate | 95% Wald Confidence Limits | |
| **firstdose 1. PFIZER vs 2. MODERN** | 1.218 | 0.936 | 1.585 |

| Association of Predicted Probabilities and Observed Responses | | | |
|---|---|---|---|
| **Percent Concordant** | 27.5 | **Somers' D** | 0.049 |
| **Percent Discordant** | 22.5 | **Gamma** | 0.098 |
| **Percent Tied** | 50.0 | **Tau-a** | 0.000 |
| **Pairs** | 439621952 | **c** | 0.525 |

### SUICIDE OR HOMICIDE death adjusted (logistic regression)

#### The LOGISTIC Procedure

| Model Information | |
|---|---|
| Data Set | WORK.MATCHEDCOHORT |
| Response Variable | suicidehomicide1yrD1 |
| Number of Response Levels | 2 |
| Model | binary logit |
| Optimization Technique | Fisher's scoring |

| | |
|---|---|
| Number of Observations Read | 1962822 |
| Number of Observations Used | 1962822 |

| Response Profile | | |
|---|---|---|
| Ordered Value | suicidehomicide1yrD1 | Total Frequency |
| 1 | 0 | 1962598 |
| 2 | 1 | 224 |

**Probability modeled is suicidehomicide1yrD1=1.**

| Class Level Information | | | | | | | | | |
|---|---|---|---|---|---|---|---|---|---|
| Class | Value | Design Variables | | | | | | | |
| agenew | 18-39 | 0 | 0 | 0 | 0 | 0 | | | |
| | 40-49 | 1 | 0 | 0 | 0 | 0 | | | |
| | 50-59 | 0 | 1 | 0 | 0 | 0 | | | |
| | 60-69 | 0 | 0 | 1 | 0 | 0 | | | |
| | 70-79 | 0 | 0 | 0 | 1 | 0 | | | |
| | 80+ | 0 | 0 | 0 | 0 | 1 | | | |
| firstdose | 1. PFIZER | 1 | | | | | | | |
| | 2. MODERN | 0 | | | | | | | |
| sex2 | F | 1 | | | | | | | |
| | M | 0 | | | | | | | |
| RACE4 | BLACK | 1 | 0 | 0 | | | | | |
| | OTHERS | 0 | 1 | 0 | | | | | |
| | UNKNOWN | 0 | 0 | 1 | | | | | |
| | WHITE | 0 | 0 | 0 | | | | | |
| ETHNICITY3 | HISPANIC | 1 | 0 | | | | | | |
| | NOT HISPANIC | 0 | 0 | | | | | | |
| | UNKNOWN | 0 | 1 | | | | | | |
| Vaccinationsite | CHD OR DOH | 1 | 0 | 0 | 0 | 0 | 0 | 0 | 0 |

### SUICIDE OR HOMICIDE death adjusted (logistic regression)

#### The LOGISTIC Procedure

| Class Level Information | | | | | | | | | |
|---|---|---|---|---|---|---|---|---|---|
| Class | Value | Design Variables | | | | | | | |
| | DOCTOR CLINIC | 0 | 1 | 0 | 0 | 0 | 0 | 0 | 0 |
| | FIRE RESQUE OR EMS | 0 | 0 | 1 | 0 | 0 | 0 | 0 | 0 |
| | FQHC OR RHC OR CHC | 0 | 0 | 0 | 1 | 0 | 0 | 0 | 0 |
| | HOSPITAL CLINIC | 0 | 0 | 0 | 0 | 1 | 0 | 0 | 0 |
| | HOSPITAL ER | 0 | 0 | 0 | 0 | 0 | 1 | 0 | 0 |
| | OTHER PHARMACIES | 0 | 0 | 0 | 0 | 0 | 0 | 1 | 0 |
| | OTHERS OR MISC | 0 | 0 | 0 | 0 | 0 | 0 | 0 | 1 |
| | PUBLIX OR CVS OR WALGREENS | 0 | 0 | 0 | 0 | 0 | 0 | 0 | 0 |

| Model Convergence Status |
|---|
| Convergence criterion (GCONV=1E-8) satisfied. |

| Model Fit Statistics | | |
|---|---|---|
| Criterion | Intercept Only | Intercept and Covariates |
| AIC | 4517.029 | 4439.208 |
| SC | 4529.519 | 4738.966 |
| -2 Log L | 4515.029 | 4391.208 |

| Testing Global Null Hypothesis: BETA=0 | | | |
|---|---|---|---|
| Test | Chi-Square | DF | Pr > ChiSq |
| Likelihood Ratio | 123.8212 | 23 | <.0001 |
| Score | 121.7876 | 23 | <.0001 |
| Wald | 113.8191 | 23 | <.0001 |

| Type 3 Analysis of Effects | | | |
|---|---|---|---|
| Effect | DF | Wald Chi-Square | Pr > ChiSq |
| firstdose | 1 | 2.1540 | 0.1422 |
| agenew | 5 | 3.0043 | 0.6993 |
| sex2 | 1 | 39.9836 | <.0001 |
| RACE4 | 3 | 7.9392 | 0.0473 |
| ETHNICITY3 | 2 | 39.5707 | <.0001 |
| decile_sviscore1 | 1 | 4.5616 | 0.0327 |
| decile_sviscore4 | 1 | 0.2037 | 0.6518 |

### SUICIDE OR HOMICIDE death adjusted (logistic regression)

#### The LOGISTIC Procedure

| Type 3 Analysis of Effects | | | |
|---|---|---|---|
| Effect | DF | Wald Chi-Square | Pr > ChiSq |
| mortalityratepct | 1 | 0.0026 | 0.9591 |
| Vaccinationsite | 8 | 22.6993 | 0.0038 |

| Analysis of Maximum Likelihood Estimates | | | | | | | |
|---|---|---|---|---|---|---|---|
| Parameter | | DF | Estimate | Standard Error | Wald Chi-Square | Pr > ChiSq | Label |
| Intercept | | 1 | -8.8641 | 0.2158 | 1686.8390 | <.0001 | Intercept: suicidehomicide1yrD1=1 |
| firstdose | 1. PFIZER | 1 | 0.1971 | 0.1343 | 2.1540 | 0.1422 | firstdose 1. PFIZER |
| agenew | 40-49 | 1 | -0.1479 | 0.2503 | 0.3489 | 0.5547 | agenew 40-49 |
| agenew | 50-59 | 1 | 0.1023 | 0.2086 | 0.2403 | 0.6240 | agenew 50-59 |
| agenew | 60-69 | 1 | 0.1002 | 0.2199 | 0.2074 | 0.6488 | agenew 60-69 |
| agenew | 70-79 | 1 | 0.3431 | 0.2710 | 1.6037 | 0.2054 | agenew 70-79 |
| agenew | 80+ | 1 | 0.6529 | 0.5261 | 1.5396 | 0.2147 | agenew 80+ |
| sex2 | F | 1 | -0.9272 | 0.1466 | 39.9836 | <.0001 | sex2 F |
| RACE4 | BLACK | 1 | -0.8287 | 0.3701 | 5.0153 | 0.0251 | RACE4 BLACK |
| RACE4 | OTHERS | 1 | -0.3424 | 0.2259 | 2.2965 | 0.1297 | RACE4 OTHERS |
| RACE4 | UNKNOWN | 1 | -0.4782 | 0.3756 | 1.6214 | 0.2029 | RACE4 UNKNOWN |
| ETHNICITY3 | HISPANIC | 1 | -1.1685 | 0.2719 | 18.4661 | <.0001 | ETHNICITY3 HISPANIC |
| ETHNICITY3 | UNKNOWN | 1 | 0.7308 | 0.1928 | 14.3715 | 0.0002 | ETHNICITY3 UNKNOWN |
| decile_sviscore1 | | 1 | 0.0632 | 0.0296 | 4.5616 | 0.0327 | Rank for Variable SVISCORE1 |
| decile_sviscore4 | | 1 | -0.0127 | 0.0281 | 0.2037 | 0.6518 | Rank for Variable SVISCORE4 |
| mortalityratepct | | 1 | 0.00292 | 0.0570 | 0.0026 | 0.9591 | |
| Vaccinationsite | CHD OR DOH | 1 | -0.6266 | 0.2629 | 5.6821 | 0.0171 | Vaccinationsite CHD OR DOH |
| Vaccinationsite | DOCTOR CLINIC | 1 | -0.1698 | 0.2000 | 0.7212 | 0.3957 | Vaccinationsite DOCTOR CLINIC |
| Vaccinationsite | FIRE RESQUE OR EMS | 1 | -0.3673 | 0.5959 | 0.3799 | 0.5377 | Vaccinationsite FIRE RESQUE OR EMS |
| Vaccinationsite | FQHC OR RHC OR CHC | 1 | -0.3598 | 0.7158 | 0.2527 | 0.6151 | Vaccinationsite FQHC OR RHC OR CHC |
| Vaccinationsite | HOSPITAL CLINIC | 1 | 0.1862 | 0.2787 | 0.4462 | 0.5041 | Vaccinationsite HOSPITAL CLINIC |
| Vaccinationsite | HOSPITAL ER | 1 | 0.2921 | 1.0056 | 0.0844 | 0.7715 | Vaccinationsite HOSPITAL ER |
| Vaccinationsite | OTHER PHARMACIES | 1 | 0.6171 | 0.2775 | 4.9476 | 0.0261 | Vaccinationsite OTHER PHARMACIES |
| Vaccinationsite | OTHERS OR MISC | 1 | -0.9450 | 0.3017 | 9.8094 | 0.0017 | Vaccinationsite OTHERS OR MISC |

### SUICIDE OR HOMICIDE death adjusted (logistic regression)

#### The LOGISTIC Procedure

| Odds Ratio Estimates | | | |
|---|---|---|---|
| Effect | Point Estimate | 95% Wald Confidence Limits | |
| firstdose 1. PFIZER vs 2. MODERN | 1.218 | 0.936 | 1.585 |
| agenew 40-49 vs 18-39 | 0.863 | 0.528 | 1.409 |
| agenew 50-59 vs 18-39 | 1.108 | 0.736 | 1.667 |
| agenew 60-69 vs 18-39 | 1.105 | 0.718 | 1.701 |
| agenew 70-79 vs 18-39 | 1.409 | 0.829 | 2.397 |
| agenew 80+ vs 18-39 | 1.921 | 0.685 | 5.388 |
| sex2 F vs M | 0.396 | 0.297 | 0.527 |
| RACE4 BLACK vs WHITE | 0.437 | 0.211 | 0.902 |
| RACE4 OTHERS vs WHITE | 0.710 | 0.456 | 1.106 |
| RACE4 UNKNOWN vs WHITE | 0.620 | 0.297 | 1.294 |
| ETHNICITY3 HISPANIC vs NOT HISPANIC | 0.311 | 0.182 | 0.530 |
| ETHNICITY3 UNKNOWN vs NOT HISPANIC | 2.077 | 1.423 | 3.030 |
| decile_sviscore1 | 1.065 | 1.005 | 1.129 |
| decile_sviscore4 | 0.987 | 0.934 | 1.043 |
| mortalityratepct | 1.003 | 0.897 | 1.121 |
| Vaccinationsite CHD OR DOH vs PUBLIX OR CVS OR WALGREENS | 0.534 | 0.319 | 0.895 |
| Vaccinationsite DOCTOR CLINIC vs PUBLIX OR CVS OR WALGREENS | 0.844 | 0.570 | 1.249 |
| Vaccinationsite FIRE RESQUE OR EMS vs PUBLIX OR CVS OR WALGREENS | 0.693 | 0.215 | 2.227 |
| Vaccinationsite FQHC OR RHC OR CHC vs PUBLIX OR CVS OR WALGREENS | 0.698 | 0.172 | 2.838 |
| Vaccinationsite HOSPITAL CLINIC vs PUBLIX OR CVS OR WALGREENS | 1.205 | 0.698 | 2.080 |
| Vaccinationsite HOSPITAL ER vs PUBLIX OR CVS OR WALGREENS | 1.339 | 0.187 | 9.612 |
| Vaccinationsite OTHER PHARMACIES vs PUBLIX OR CVS OR WALGREENS | 1.854 | 1.076 | 3.193 |
| Vaccinationsite OTHERS OR MISC vs PUBLIX OR CVS OR WALGREENS | 0.389 | 0.215 | 0.702 |

| Association of Predicted Probabilities and Observed Responses | | | |
|---|---|---|---|
| Percent Concordant | 70.6 | Somers' D | 0.412 |
| Percent Discordant | 29.4 | Gamma | 0.412 |
| Percent Tied | 0.0 | Tau-a | 0.000 |
| Pairs | 439621952 | c | 0.706 |

### SUICIDE OR HOMICIDE deaths adjusted by Age

#### The LOGISTIC Procedure

**agenew=18-39 years**

| Model Information | |
|---|---|
| **Data Set** | WORK.MATCHEDCOHORT |
| **Response Variable** | suicidehomicide1yrD1 |
| **Number of Response Levels** | 2 |
| **Model** | binary logit |
| **Optimization Technique** | Fisher's scoring |

| | |
|---|---|
| **Number of Observations Read** | 492484 |
| **Number of Observations Used** | 492484 |

| Response Profile | | |
|---|---|---|
| **Ordered Value** | **suicidehomicide1yrD1** | **Total Frequency** |
| **1** | 0 | 492431 |
| **2** | 1 | 53 |

**Probability modeled is suicidehomicide1yrD1=1.**

| Class Level Information | | | | | | | | | |
|---|---|---|---|---|---|---|---|---|---|
| **Class** | **Value** | **Design Variables** | | | | | | | |
| **agenew** | **18-39 years** | | | | | | | | |
| **firstdose** | **1. PFIZER** | 1 | | | | | | | |
| | **2. MODERN** | 0 | | | | | | | |
| **sex2** | **F** | 1 | | | | | | | |
| | **M** | 0 | | | | | | | |
| **RACE4** | **BLACK** | 1 | 0 | 0 | | | | | |
| | **OTHERS** | 0 | 1 | 0 | | | | | |
| | **UNKNOWN** | 0 | 0 | 1 | | | | | |
| | **WHITE** | 0 | 0 | 0 | | | | | |
| **ETHNICITY3** | **HISPANIC** | 1 | 0 | | | | | | |
| | **NOT HISPANIC** | 0 | 0 | | | | | | |
| | **UNKNOWN** | 0 | 1 | | | | | | |
| **Vaccinationsite** | **CHD OR DOH** | 1 | 0 | 0 | 0 | 0 | 0 | 0 | 0 |
| | **DOCTOR CLINIC** | 0 | 1 | 0 | 0 | 0 | 0 | 0 | 0 |
| | **FIRE RESQUE OR EMS** | 0 | 0 | 1 | 0 | 0 | 0 | 0 | 0 |
| | **FQHC OR RHC OR CHC** | 0 | 0 | 0 | 1 | 0 | 0 | 0 | 0 |
| | **HOSPITAL CLINIC** | 0 | 0 | 0 | 0 | 1 | 0 | 0 | 0 |
| | **HOSPITAL ER** | 0 | 0 | 0 | 0 | 0 | 1 | 0 | 0 |

#### The LOGISTIC Procedure

**agenew=18-39 years**

| Class Level Information | | | | | | | | | |
|---|---|---|---|---|---|---|---|---|---|
| **Class** | **Value** | **Design Variables** | | | | | | | |
| | **OTHER PHARMACIES** | 0 | 0 | 0 | 0 | 0 | 0 | 1 | 0 |
| | **OTHERS OR MISC** | 0 | 0 | 0 | 0 | 0 | 0 | 0 | 1 |
| | **PUBLIX OR CVS OR WALGREENS** | 0 | 0 | 0 | 0 | 0 | 0 | 0 | 0 |

| Model Convergence Status |
|---|
| Convergence criterion (GCONV=1E-8) satisfied. |

| Model Fit Statistics | | |
|---|---|---|
| **Criterion** | **Intercept Only** | **Intercept and Covariates** |
| **AIC** | 1076.508 | 1086.253 |
| **SC** | 1087.616 | 1297.290 |
| **-2 Log L** | 1074.508 | 1048.253 |

| Testing Global Null Hypothesis: BETA=0 | | | |
|---|---|---|---|
| **Test** | **Chi-Square** | **DF** | **Pr > ChiSq** |
| **Likelihood Ratio** | 26.2554 | 18 | 0.0941 |
| **Score** | 25.2616 | 18 | 0.1179 |
| **Wald** | 22.8598 | 18 | 0.1960 |

| Type 3 Analysis of Effects | | | |
|---|---|---|---|
| **Effect** | **DF** | **Wald Chi-Square** | **Pr > ChiSq** |
| **firstdose** | 1 | 0.9167 | 0.3383 |
| **agenew** | 0 | . | . |
| **sex2** | 1 | 11.5992 | 0.0007 |
| **RACE4** | 3 | 0.8959 | 0.8264 |
| **ETHNICITY3** | 2 | 5.2314 | 0.0731 |
| **decile_sviscore1** | 1 | 1.2280 | 0.2678 |
| **decile_sviscore4** | 1 | 0.0557 | 0.8135 |
| **mortalityratepct** | 1 | 0.0000 | 0.9956 |
| **Vaccinationsite** | 8 | 2.5786 | 0.9580 |

#### The LOGISTIC Procedure

**agenew=18-39 years**

| Analysis of Maximum Likelihood Estimates | | | | | | | |
|---|---|---|---|---|---|---|---|
| Parameter | | DF | Estimate | Standard Error | Wald Chi-Square | Pr > ChiSq | Label |
| Intercept | | 1 | -9.2093 | 0.3804 | 586.1200 | <.0001 | Intercept: suicidehomicide1yrD1=1 |
| firstdose | 1. PFIZER | 1 | 0.2654 | 0.2772 | 0.9167 | 0.3383 | firstdose 1. PFIZER |
| sex2 | F | 1 | -1.0230 | 0.3004 | 11.5992 | 0.0007 | sex2 F |
| RACE4 | BLACK | 1 | 0.0264 | 0.4977 | 0.0028 | 0.9577 | RACE4 BLACK |
| RACE4 | OTHERS | 1 | 0.3056 | 0.3550 | 0.7412 | 0.3893 | RACE4 OTHERS |
| RACE4 | UNKNOWN | 1 | -0.1491 | 0.6693 | 0.0496 | 0.8237 | RACE4 UNKNOWN |
| ETHNICITY3 | HISPANIC | 1 | -0.7484 | 0.3987 | 3.5246 | 0.0605 | ETHNICITY3 HISPANIC |
| ETHNICITY3 | UNKNOWN | 1 | 0.2800 | 0.3720 | 0.5667 | 0.4516 | ETHNICITY3 UNKNOWN |
| decile_sviscore1 | | 1 | 0.0683 | 0.0616 | 1.2280 | 0.2678 | Rank for Variable SVISCORE1 |
| decile_sviscore4 | | 1 | 0.0139 | 0.0590 | 0.0557 | 0.8135 | Rank for Variable SVISCORE4 |
| mortalityratepct | | 1 | -13.0652 | 2365.7 | 0.0000 | 0.9956 | |
| Vaccinationsite | CHD OR DOH | 1 | -0.7566 | 1.0171 | 0.5534 | 0.4569 | Vaccinationsite CHD OR DOH |
| Vaccinationsite | DOCTOR CLINIC | 1 | 0.2195 | 0.4157 | 0.2789 | 0.5974 | Vaccinationsite DOCTOR CLINIC |
| Vaccinationsite | FIRE RESQUE OR EMS | 1 | -12.6617 | 2631.0 | 0.0000 | 0.9962 | Vaccinationsite FIRE RESQUE OR EMS |
| Vaccinationsite | FQHC OR RHC OR CHC | 1 | -12.5315 | 768.2 | 0.0003 | 0.9870 | Vaccinationsite FQHC OR RHC OR CHC |
| Vaccinationsite | HOSPITAL CLINIC | 1 | -0.4033 | 1.0178 | 0.1570 | 0.6920 | Vaccinationsite HOSPITAL CLINIC |
| Vaccinationsite | HOSPITAL ER | 1 | -12.6975 | 1783.0 | 0.0001 | 0.9943 | Vaccinationsite HOSPITAL ER |
| Vaccinationsite | OTHER PHARMACIES | 1 | 0.5827 | 0.4870 | 1.4317 | 0.2315 | Vaccinationsite OTHER PHARMACIES |
| Vaccinationsite | OTHERS OR MISC | 1 | 0.2260 | 0.5355 | 0.1782 | 0.6730 | Vaccinationsite OTHERS OR MISC |

| Odds Ratio Estimates | | | |
|---|---|---|---|
| Effect | Point Estimate | 95% Wald Confidence Limits | |
| firstdose 1. PFIZER vs 2. MODERN | 1.304 | 0.757 | 2.245 |
| sex2 F vs M | 0.360 | 0.200 | 0.648 |
| RACE4 BLACK vs WHITE | 1.027 | 0.387 | 2.724 |
| RACE4 OTHERS vs WHITE | 1.357 | 0.677 | 2.722 |
| RACE4 UNKNOWN vs WHITE | 0.861 | 0.232 | 3.199 |
| ETHNICITY3 HISPANIC vs NOT HISPANIC | 0.473 | 0.217 | 1.033 |
| ETHNICITY3 UNKNOWN vs NOT HISPANIC | 1.323 | 0.638 | 2.743 |
| decile_sviscore1 | 1.071 | 0.949 | 1.208 |
| decile_sviscore4 | 1.014 | 0.903 | 1.138 |
| mortalityratepct | <0.001 | <0.001 | >999.999 |
| Vaccinationsite CHD OR DOH vs PUBLIX OR CVS OR WALGREENS | 0.469 | 0.064 | 3.445 |

#### The LOGISTIC Procedure

##### agenew=18-39 years

| Odds Ratio Estimates | | | |
|---|---|---|---|
| Effect | Point Estimate | 95% Wald Confidence Limits | |
| Vaccinationsite DOCTOR CLINIC vs PUBLIX OR CVS OR WALGREENS | 1.245 | 0.551 | 2.813 |
| Vaccinationsite FIRE RESQUE OR EMS vs PUBLIX OR CVS OR WALGREENS | <0.001 | <0.001 | >999.999 |
| Vaccinationsite FQHC OR RHC OR CHC vs PUBLIX OR CVS OR WALGREENS | <0.001 | <0.001 | >999.999 |
| Vaccinationsite HOSPITAL CLINIC vs PUBLIX OR CVS OR WALGREENS | 0.668 | 0.091 | 4.912 |
| Vaccinationsite HOSPITAL ER vs PUBLIX OR CVS OR WALGREENS | <0.001 | <0.001 | >999.999 |
| Vaccinationsite OTHER PHARMACIES vs PUBLIX OR CVS OR WALGREENS | 1.791 | 0.690 | 4.651 |
| Vaccinationsite OTHERS OR MISC vs PUBLIX OR CVS OR WALGREENS | 1.254 | 0.439 | 3.581 |

| Association of Predicted Probabilities and Observed Responses | | | |
|---|---|---|---|
| Percent Concordant | 69.7 | Somers' D | 0.395 |
| Percent Discordant | 30.2 | Gamma | 0.395 |
| Percent Tied | 0.1 | Tau-a | 0.000 |
| Pairs | 26098843 | c | 0.697 |

### The LOGISTIC Procedure

**agenew=40-59 years**

| Model Information | |
|---|---|
| Data Set | WORK.MATCHEDCOHORT |
| Response Variable | suicidehomicide1yrD1 |
| Number of Response Levels | 2 |
| Model | binary logit |
| Optimization Technique | Fisher's scoring |

| | |
|---|---|
| Number of Observations Read | 634102 |
| Number of Observations Used | 634102 |

| Response Profile | | |
|---|---|---|
| Ordered Value | suicidehomicide1yrD1 | Total Frequency |
| 1 | 0 | 634034 |
| 2 | 1 | 68 |

**Probability modeled is suicidehomicide1yrD1=1.**

| Class Level Information | | | | | | | | | |
|---|---|---|---|---|---|---|---|---|---|
| Class | Value | Design Variables | | | | | | | |
| agenew | 40-59 years | | | | | | | | |
| firstdose | 1. PFIZER | 1 | | | | | | | |
| | 2. MODERN | 0 | | | | | | | |
| sex2 | F | 1 | | | | | | | |
| | M | 0 | | | | | | | |
| RACE4 | BLACK | 1 | 0 | 0 | | | | | |
| | OTHERS | 0 | 1 | 0 | | | | | |
| | UNKNOWN | 0 | 0 | 1 | | | | | |
| | WHITE | 0 | 0 | 0 | | | | | |
| ETHNICITY3 | HISPANIC | 1 | 0 | | | | | | |
| | NOT HISPANIC | 0 | 0 | | | | | | |
| | UNKNOWN | 0 | 1 | | | | | | |
| Vaccinationsite | CHD OR DOH | 1 | 0 | 0 | 0 | 0 | 0 | 0 | 0 |
| | DOCTOR CLINIC | 0 | 1 | 0 | 0 | 0 | 0 | 0 | 0 |
| | FIRE RESQUE OR EMS | 0 | 0 | 1 | 0 | 0 | 0 | 0 | 0 |
| | FQHC OR RHC OR CHC | 0 | 0 | 0 | 1 | 0 | 0 | 0 | 0 |
| | HOSPITAL CLINIC | 0 | 0 | 0 | 0 | 1 | 0 | 0 | 0 |
| | HOSPITAL ER | 0 | 0 | 0 | 0 | 0 | 1 | 0 | 0 |

#### The LOGISTIC Procedure

**agenew=40-59 years**

| Class Level Information | | | | | | | | | |
|---|---|---|---|---|---|---|---|---|---|
| **Class** | **Value** | **Design Variables** | | | | | | | |
| | **OTHER PHARMACIES** | 0 | 0 | 0 | 0 | 0 | 0 | 1 | 0 |
| | **OTHERS OR MISC** | 0 | 0 | 0 | 0 | 0 | 0 | 0 | 1 |
| | **PUBLIX OR CVS OR WALGREENS** | 0 | 0 | 0 | 0 | 0 | 0 | 0 | 0 |

| Model Convergence Status |
|---|
| Convergence criterion (GCONV=1E-8) satisfied. |

| Model Fit Statistics | | |
|---|---|---|
| **Criterion** | **Intercept Only** | **Intercept and Covariates** |
| **AIC** | 1381.095 | 1366.988 |
| **SC** | 1392.455 | 1582.827 |
| **-2 Log L** | 1379.095 | 1328.988 |

| Testing Global Null Hypothesis: BETA=0 | | | |
|---|---|---|---|
| **Test** | **Chi-Square** | **DF** | **Pr > ChiSq** |
| **Likelihood Ratio** | 50.1068 | 18 | <.0001 |
| **Score** | 46.7488 | 18 | 0.0002 |
| **Wald** | 39.9876 | 18 | 0.0021 |

| Type 3 Analysis of Effects | | | |
|---|---|---|---|
| **Effect** | **DF** | **Wald Chi-Square** | **Pr > ChiSq** |
| **firstdose** | 1 | 0.0599 | 0.8067 |
| **agenew** | 0 | . | . |
| **sex2** | 1 | 5.8900 | 0.0152 |
| **RACE4** | 3 | 4.3416 | 0.2269 |
| **ETHNICITY3** | 2 | 7.9363 | 0.0189 |
| **decile_sviscore1** | 1 | 0.2525 | 0.6153 |
| **decile_sviscore4** | 1 | 0.8642 | 0.3526 |
| **mortalityratepct** | 1 | 3.4836 | 0.0620 |
| **Vaccinationsite** | 8 | 11.6661 | 0.1667 |

### SUICIDE OR HOMICIDE deaths adjusted by Age

#### The LOGISTIC Procedure

**agenew=40-59 years**

| Analysis of Maximum Likelihood Estimates | | | | | | | |
|---|---|---|---|---|---|---|---|
| **Parameter** | | **DF** | **Estimate** | **Standard Error** | **Wald Chi-Square** | **Pr > ChiSq** | **Label** |
| **Intercept** | | 1 | -8.9883 | 0.3528 | 648.9371 | <.0001 | Intercept: suicidehomicide1yrD1=1 |
| **firstdose** | **1. PFIZER** | 1 | 0.0594 | 0.2427 | 0.0599 | 0.8067 | firstdose 1. PFIZER |
| **sex2** | **F** | 1 | -0.6577 | 0.2710 | 5.8900 | 0.0152 | sex2 F |
| **RACE4** | **BLACK** | 1 | -1.7470 | 1.0176 | 2.9476 | 0.0860 | RACE4 BLACK |
| **RACE4** | **OTHERS** | 1 | -0.5918 | 0.4852 | 1.4875 | 0.2226 | RACE4 OTHERS |
| **RACE4** | **UNKNOWN** | 1 | -0.0289 | 0.6013 | 0.0023 | 0.9616 | RACE4 UNKNOWN |
| **ETHNICITY3** | **HISPANIC** | 1 | -0.7084 | 0.4309 | 2.7024 | 0.1002 | ETHNICITY3 HISPANIC |
| **ETHNICITY3** | **UNKNOWN** | 1 | 0.7055 | 0.3715 | 3.6058 | 0.0576 | ETHNICITY3 UNKNOWN |
| **decile_sviscore1** | | 1 | -0.0270 | 0.0537 | 0.2525 | 0.6153 | Rank for Variable SVISCORE1 |
| **decile_sviscore4** | | 1 | 0.0473 | 0.0508 | 0.8642 | 0.3526 | Rank for Variable SVISCORE4 |
| **mortalityratepct** | | 1 | 0.4970 | 0.2663 | 3.4836 | 0.0620 | |
| **Vaccinationsite** | **CHD OR DOH** | 1 | -0.2347 | 0.7256 | 0.1047 | 0.7463 | Vaccinationsite CHD OR DOH |
| **Vaccinationsite** | **DOCTOR CLINIC** | 1 | -0.5341 | 0.4108 | 1.6909 | 0.1935 | Vaccinationsite DOCTOR CLINIC |
| **Vaccinationsite** | **FIRE RESQUE OR EMS** | 1 | -12.6625 | 1443.3 | 0.0001 | 0.9930 | Vaccinationsite FIRE RESQUE OR EMS |
| **Vaccinationsite** | **FQHC OR RHC OR CHC** | 1 | -12.4942 | 560.6 | 0.0005 | 0.9822 | Vaccinationsite FQHC OR RHC OR CHC |
| **Vaccinationsite** | **HOSPITAL CLINIC** | 1 | 0.8176 | 0.3876 | 4.4488 | 0.0349 | Vaccinationsite HOSPITAL CLINIC |
| **Vaccinationsite** | **HOSPITAL ER** | 1 | -12.8427 | 1274.9 | 0.0001 | 0.9920 | Vaccinationsite HOSPITAL ER |
| **Vaccinationsite** | **OTHER PHARMACIES** | 1 | 0.4255 | 0.4773 | 0.7947 | 0.3727 | Vaccinationsite OTHER PHARMACIES |
| **Vaccinationsite** | **OTHERS OR MISC** | 1 | -1.9316 | 1.0168 | 3.6085 | 0.0575 | Vaccinationsite OTHERS OR MISC |

| Odds Ratio Estimates | | | |
|---|---|---|---|
| **Effect** | **Point Estimate** | **95% Wald Confidence Limits** | |
| **firstdose 1. PFIZER vs 2. MODERN** | 1.061 | 0.660 | 1.707 |
| **sex2 F vs M** | 0.518 | 0.305 | 0.881 |
| **RACE4 BLACK vs WHITE** | 0.174 | 0.024 | 1.281 |
| **RACE4 OTHERS vs WHITE** | 0.553 | 0.214 | 1.432 |
| **RACE4 UNKNOWN vs WHITE** | 0.971 | 0.299 | 3.157 |
| **ETHNICITY3 HISPANIC vs NOT HISPANIC** | 0.492 | 0.212 | 1.146 |
| **ETHNICITY3 UNKNOWN vs NOT HISPANIC** | 2.025 | 0.978 | 4.194 |
| **decile_sviscore1** | 0.973 | 0.876 | 1.081 |
| **decile_sviscore4** | 1.048 | 0.949 | 1.158 |
| **mortalityratepct** | 1.644 | 0.975 | 2.770 |
| **Vaccinationsite CHD OR DOH vs PUBLIX OR CVS OR WALGREENS** | 0.791 | 0.191 | 3.278 |

#### The LOGISTIC Procedure

**agenew=40-59 years**

| Odds Ratio Estimates | | | |
|---|---|---|---|
| Effect | Point Estimate | 95% Wald Confidence Limits | |
| Vaccinationsite DOCTOR CLINIC vs PUBLIX OR CVS OR WALGREENS | 0.586 | 0.262 | 1.311 |
| Vaccinationsite FIRE RESQUE OR EMS vs PUBLIX OR CVS OR WALGREENS | <0.001 | <0.001 | >999.999 |
| Vaccinationsite FQHC OR RHC OR CHC vs PUBLIX OR CVS OR WALGREENS | <0.001 | <0.001 | >999.999 |
| Vaccinationsite HOSPITAL CLINIC vs PUBLIX OR CVS OR WALGREENS | 2.265 | 1.060 | 4.842 |
| Vaccinationsite HOSPITAL ER vs PUBLIX OR CVS OR WALGREENS | <0.001 | <0.001 | >999.999 |
| Vaccinationsite OTHER PHARMACIES vs PUBLIX OR CVS OR WALGREENS | 1.530 | 0.600 | 3.900 |
| Vaccinationsite OTHERS OR MISC vs PUBLIX OR CVS OR WALGREENS | 0.145 | 0.020 | 1.063 |

| Association of Predicted Probabilities and Observed Responses | | | |
|---|---|---|---|
| Percent Concordant | 73.4 | Somers' D | 0.468 |
| Percent Discordant | 26.6 | Gamma | 0.468 |
| Percent Tied | 0.0 | Tau-a | 0.000 |
| Pairs | 43114312 | c | 0.734 |

#### The LOGISTIC Procedure

**agenew=60+ years**

| Model Information | |
|---|---|
| **Data Set** | WORK.MATCHEDCOHORT |
| **Response Variable** | suicidehomicide1yrD1 |
| **Number of Response Levels** | 2 |
| **Model** | binary logit |
| **Optimization Technique** | Fisher's scoring |

| | |
|---|---|
| **Number of Observations Read** | 836236 |
| **Number of Observations Used** | 836236 |

| Response Profile | | |
|---|---|---|
| **Ordered Value** | **suicidehomicide1yrD1** | **Total Frequency** |
| **1** | 0 | 836133 |
| **2** | 1 | 103 |

**Probability modeled is suicidehomicide1yrD1=1.**

| Class Level Information | | | | | | | | | |
|---|---|---|---|---|---|---|---|---|---|
| **Class** | **Value** | **Design Variables** | | | | | | | |
| **agenew** | **60+ years** | | | | | | | | |
| **firstdose** | **1. PFIZER** | 1 | | | | | | | |
| | **2. MODERN** | 0 | | | | | | | |
| **sex2** | **F** | 1 | | | | | | | |
| | **M** | 0 | | | | | | | |
| **RACE4** | **BLACK** | 1 | 0 | 0 | | | | | |
| | **OTHERS** | 0 | 1 | 0 | | | | | |
| | **UNKNOWN** | 0 | 0 | 1 | | | | | |
| | **WHITE** | 0 | 0 | 0 | | | | | |
| **ETHNICITY3** | **HISPANIC** | 1 | 0 | | | | | | |
| | **NOT HISPANIC** | 0 | 0 | | | | | | |
| | **UNKNOWN** | 0 | 1 | | | | | | |
| **Vaccinationsite** | **CHD OR DOH** | 1 | 0 | 0 | 0 | 0 | 0 | 0 | 0 |
| | **DOCTOR CLINIC** | 0 | 1 | 0 | 0 | 0 | 0 | 0 | 0 |
| | **FIRE RESQUE OR EMS** | 0 | 0 | 1 | 0 | 0 | 0 | 0 | 0 |
| | **FQHC OR RHC OR CHC** | 0 | 0 | 0 | 1 | 0 | 0 | 0 | 0 |
| | **HOSPITAL CLINIC** | 0 | 0 | 0 | 0 | 1 | 0 | 0 | 0 |
| | **HOSPITAL ER** | 0 | 0 | 0 | 0 | 0 | 1 | 0 | 0 |

#### The LOGISTIC Procedure

**agenew=60+ years**

| Class Level Information | | | | | | | | | |
|---|---|---|---|---|---|---|---|---|---|
| **Class** | **Value** | **Design Variables** | | | | | | | |
| | **OTHER PHARMACIES** | 0 | 0 | 0 | 0 | 0 | 0 | 1 | 0 |
| | **OTHERS OR MISC** | 0 | 0 | 0 | 0 | 0 | 0 | 0 | 1 |
| | **PUBLIX OR CVS OR WALGREENS** | 0 | 0 | 0 | 0 | 0 | 0 | 0 | 0 |

| Model Convergence Status |
|---|
| Quasi-complete separation of data points detected. |

**Warning:** The maximum likelihood estimate may not exist.

**Warning:** The LOGISTIC procedure continues in spite of the above warning. Results shown are based on the last maximum likelihood iteration. Validity of the model fit is questionable.

| Model Fit Statistics | | |
|---|---|---|
| **Criterion** | **Intercept Only** | **Intercept and Covariates** |
| **AIC** | 2062.386 | 2007.054 |
| **SC** | 2074.023 | 2228.151 |
| **-2 Log L** | 2060.386 | 1969.054 |

| Testing Global Null Hypothesis: BETA=0 | | | |
|---|---|---|---|
| **Test** | **Chi-Square** | **DF** | **Pr > ChiSq** |
| **Likelihood Ratio** | 91.3323 | 18 | <.0001 |
| **Score** | 89.2087 | 18 | <.0001 |
| **Wald** | 68.1589 | 18 | <.0001 |

| Type 3 Analysis of Effects | | | |
|---|---|---|---|
| **Effect** | **DF** | **Wald Chi-Square** | **Pr > ChiSq** |
| **firstdose** | 1 | 1.6268 | 0.2021 |
| **agenew** | 0 | . | . |
| **sex2** | 1 | 19.3950 | <.0001 |
| **RACE4** | 3 | 6.6933 | 0.0823 |
| **ETHNICITY3** | 2 | 18.6880 | <.0001 |
| **decile_sviscore1** | 1 | 6.7870 | 0.0092 |
| **decile_sviscore4** | 1 | 1.9204 | 0.1658 |
| **mortalityratepct** | 1 | 1.9429 | 0.1634 |
| **Vaccinationsite** | 8 | 16.2155 | 0.0394 |

#### The LOGISTIC Procedure

**Warning:** The validity of the model fit is questionable.

**agenew=60+ years**

| Analysis of Maximum Likelihood Estimates | | | | | | | |
|---|---|---|---|---|---|---|---|
| Parameter | | DF | Estimate | Standard Error | Wald Chi-Square | Pr > ChiSq | Label |
| Intercept | | 1 | -8.7850 | 0.2907 | 913.4006 | <.0001 | Intercept: suicidehomicide1yrD1=1 |
| firstdose | 1. PFIZER | 1 | 0.2534 | 0.1987 | 1.6268 | 0.2021 | firstdose 1. PFIZER |
| sex2 | F | 1 | -0.9492 | 0.2155 | 19.3950 | <.0001 | sex2 F |
| RACE4 | BLACK | 1 | -1.2615 | 0.7281 | 3.0018 | 0.0832 | RACE4 BLACK |
| RACE4 | OTHERS | 1 | -0.5506 | 0.3957 | 1.9359 | 0.1641 | RACE4 OTHERS |
| RACE4 | UNKNOWN | 1 | -1.1218 | 0.7528 | 2.2205 | 0.1362 | RACE4 UNKNOWN |
| ETHNICITY3 | HISPANIC | 1 | -13.9188 | 279.1 | 0.0025 | 0.9602 | ETHNICITY3 HISPANIC |
| ETHNICITY3 | UNKNOWN | 1 | 1.1946 | 0.2763 | 18.6855 | <.0001 | ETHNICITY3 UNKNOWN |
| decile_sviscore1 | | 1 | 0.1127 | 0.0433 | 6.7870 | 0.0092 | Rank for Variable SVISCORE1 |
| decile_sviscore4 | | 1 | -0.0573 | 0.0413 | 1.9204 | 0.1658 | Rank for Variable SVISCORE4 |
| mortalityratepct | | 1 | 0.0481 | 0.0345 | 1.9429 | 0.1634 | |
| Vaccinationsite | CHD OR DOH | 1 | -0.6039 | 0.2975 | 4.1208 | 0.0424 | Vaccinationsite CHD OR DOH |
| Vaccinationsite | DOCTOR CLINIC | 1 | -0.0113 | 0.2767 | 0.0017 | 0.9676 | Vaccinationsite DOCTOR CLINIC |
| Vaccinationsite | FIRE RESQUE OR EMS | 1 | -0.0986 | 0.6008 | 0.0269 | 0.8696 | Vaccinationsite FIRE RESQUE OR EMS |
| Vaccinationsite | FQHC OR RHC OR CHC | 1 | 0.2065 | 0.7246 | 0.0812 | 0.7757 | Vaccinationsite FQHC OR RHC OR CHC |
| Vaccinationsite | HOSPITAL CLINIC | 1 | -0.1276 | 0.4392 | 0.0844 | 0.7715 | Vaccinationsite HOSPITAL CLINIC |
| Vaccinationsite | HOSPITAL ER | 1 | 0.9353 | 1.0138 | 0.8511 | 0.3562 | Vaccinationsite HOSPITAL ER |
| Vaccinationsite | OTHER PHARMACIES | 1 | 0.7602 | 0.4773 | 2.5364 | 0.1112 | Vaccinationsite OTHER PHARMACIES |
| Vaccinationsite | OTHERS OR MISC | 1 | -1.0493 | 0.3916 | 7.1804 | 0.0074 | Vaccinationsite OTHERS OR MISC |

| Odds Ratio Estimates | | | |
|---|---|---|---|
| Effect | Point Estimate | 95% Wald Confidence Limits | |
| firstdose 1. PFIZER vs 2. MODERN | 1.288 | 0.873 | 1.902 |
| sex2 F vs M | 0.387 | 0.254 | 0.591 |
| RACE4 BLACK vs WHITE | 0.283 | 0.068 | 1.180 |
| RACE4 OTHERS vs WHITE | 0.577 | 0.265 | 1.252 |
| RACE4 UNKNOWN vs WHITE | 0.326 | 0.074 | 1.424 |
| ETHNICITY3 HISPANIC vs NOT HISPANIC | <0.001 | <0.001 | >999.999 |
| ETHNICITY3 UNKNOWN vs NOT HISPANIC | 3.302 | 1.921 | 5.676 |
| decile_sviscore1 | 1.119 | 1.028 | 1.218 |
| decile_sviscore4 | 0.944 | 0.871 | 1.024 |
| mortalityratepct | 1.049 | 0.981 | 1.123 |
| Vaccinationsite CHD OR DOH vs PUBLIX OR CVS OR WALGREENS | 0.547 | 0.305 | 0.979 |

#### The LOGISTIC Procedure

**Warning:** The validity of the model fit is questionable.

**agenew=60+ years**

| Odds Ratio Estimates | | | |
|---|---|---|---|
| Effect | Point Estimate | 95% Wald Confidence Limits | |
| Vaccinationsite DOCTOR CLINIC vs PUBLIX OR CVS OR WALGREENS | 0.989 | 0.575 | 1.701 |
| Vaccinationsite FIRE RESQUE OR EMS vs PUBLIX OR CVS OR WALGREENS | 0.906 | 0.279 | 2.942 |
| Vaccinationsite FQHC OR RHC OR CHC vs PUBLIX OR CVS OR WALGREENS | 1.229 | 0.297 | 5.087 |
| Vaccinationsite HOSPITAL CLINIC vs PUBLIX OR CVS OR WALGREENS | 0.880 | 0.372 | 2.082 |
| Vaccinationsite HOSPITAL ER vs PUBLIX OR CVS OR WALGREENS | 2.548 | 0.349 | 18.584 |
| Vaccinationsite OTHER PHARMACIES vs PUBLIX OR CVS OR WALGREENS | 2.139 | 0.839 | 5.451 |
| Vaccinationsite OTHERS OR MISC vs PUBLIX OR CVS OR WALGREENS | 0.350 | 0.163 | 0.754 |

| Association of Predicted Probabilities and Observed Responses | | | |
|---|---|---|---|
| Percent Concordant | 75.4 | Somers' D | 0.507 |
| Percent Discordant | 24.6 | Gamma | 0.508 |
| Percent Tied | 0.0 | Tau-a | 0.000 |
| Pairs | 86121699 | c | 0.754 |

### Matched cohort Risk of death Dose 1 All-cause deaths

#### The LOGISTIC Procedure

| Model Information | |
|---|---|
| Data Set | WORK.DOSE1MATCHEDDATAFINAL |
| Response Variable | death1yrd1 |
| Number of Response Levels | 2 |
| Model | binary logit |
| Optimization Technique | Fisher's scoring |

| | |
|---|---|
| Number of Observations Read | 11288333 |
| Number of Observations Used | 11288333 |

| Response Profile | | |
|---|---|---|
| Ordered Value | death1yrd1 | Total Frequency |
| 1 | 0 | 11174068 |
| 2 | 1 | 114265 |

**Probability modeled is death1yrd1=1.**

| Class Level Information | | |
|---|---|---|
| Class | Value | Design Variables |
| firstdose | 1. PFIZER | 1 |
| | 2. MODERN | 0 |

| Model Convergence Status |
|---|
| Convergence criterion (GCONV=1E-8) satisfied. |

| Model Fit Statistics | | |
|---|---|---|
| Criterion | Intercept Only | Intercept and Covariates |
| AIC | 1277010.8 | 1276671.9 |
| SC | 1277025.0 | 1276700.3 |
| -2 Log L | 1277008.8 | 1276667.9 |

| Testing Global Null Hypothesis: BETA=0 | | | |
|---|---|---|---|
| Test | Chi-Square | DF | Pr > ChiSq |
| Likelihood Ratio | 340.9397 | 1 | <.0001 |
| Score | 342.8185 | 1 | <.0001 |
| Wald | 342.4875 | 1 | <.0001 |

### Matched cohort Risk of death Dose 1 All-cause deaths

#### The LOGISTIC Procedure

| Type 3 Analysis of Effects | | | |
|---|---|---|---|
| **Effect** | **DF** | **Wald Chi-Square** | **Pr > ChiSq** |
| **firstdose** | 1 | 342.4875 | <.0001 |

| Analysis of Maximum Likelihood Estimates | | | | | | | |
|---|---|---|---|---|---|---|---|
| **Parameter** | | **DF** | **Estimate** | **Standard Error** | **Wald Chi-Square** | **Pr > ChiSq** | **Label** |
| **Intercept** | | 1 | -4.5198 | 0.00446 | 1026164.42 | <.0001 | Intercept: death1yrd1=1 |
| **firstdose** | **1. PFIZER** | 1 | -0.1107 | 0.00598 | 342.4875 | <.0001 | firstdose 1. PFIZER |

| Odds Ratio Estimates | | | |
|---|---|---|---|
| **Effect** | **Point Estimate** | **95% Wald Confidence Limits** | |
| **firstdose 1. PFIZER vs 2. MODERN** | 0.895 | 0.885 | 0.906 |

| Association of Predicted Probabilities and Observed Responses | | | |
|---|---|---|---|
| **Percent Concordant** | 25.9 | **Somers' D** | 0.027 |
| **Percent Discordant** | 23.2 | **Gamma** | 0.055 |
| **Percent Tied** | 50.9 | **Tau-a** | 0.001 |
| **Pairs** | 1.2768049E12 | **c** | 0.514 |

### All-cause death adjusted (logistic regression)

#### The LOGISTIC Procedure

| Model Information | |
|---|---|
| **Data Set** | WORK.DOSE1MATCHEDDATAFINAL |
| **Response Variable** | death1yr |
| **Number of Response Levels** | 2 |
| **Model** | binary logit |
| **Optimization Technique** | Fisher's scoring |

| | |
|---|---|
| **Number of Observations Read** | 11288333 |
| **Number of Observations Used** | 11073697 |

| Response Profile | | |
|---|---|---|
| **Ordered Value** | **death1yr** | **Total Frequency** |
| **1** | 0 | 10948189 |
| **2** | 1 | 125508 |

**Probability modeled is death1yr=1.**

**Note:** 214636 observations were deleted due to missing values for the response or explanatory variables.

| Class Level Information | | | | | | | | | |
|---|---|---|---|---|---|---|---|---|---|
| **Class** | **Value** | **Design Variables** | | | | | | | |
| **agenew** | **18-39** | 0 | 0 | 0 | 0 | 0 | | | |
| | **40-49** | 1 | 0 | 0 | 0 | 0 | | | |
| | **50-59** | 0 | 1 | 0 | 0 | 0 | | | |
| | **60-69** | 0 | 0 | 1 | 0 | 0 | | | |
| | **70-79** | 0 | 0 | 0 | 1 | 0 | | | |
| | **80+** | 0 | 0 | 0 | 0 | 1 | | | |
| **firstdose** | **1. PFIZER** | 1 | | | | | | | |
| | **2. MODERN** | 0 | | | | | | | |
| **sex2** | **F** | 1 | | | | | | | |
| | **M** | 0 | | | | | | | |
| **RACE4** | **BLACK** | 1 | 0 | 0 | | | | | |
| | **OTHERS** | 0 | 1 | 0 | | | | | |
| | **UNKNOWN** | 0 | 0 | 1 | | | | | |
| | **WHITE** | 0 | 0 | 0 | | | | | |
| **ETHNICITY3** | **HISPANIC** | 1 | 0 | | | | | | |
| | **NOT HISPANIC** | 0 | 0 | | | | | | |
| | **UNKNOWN** | 0 | 1 | | | | | | |
| **Vaccinationsite** | **CHD OR DOH** | 1 | 0 | 0 | 0 | 0 | 0 | 0 | 0 |

#### The LOGISTIC Procedure

| Class Level Information | | | | | | | | | |
|---|---|---|---|---|---|---|---|---|---|
| **Class** | **Value** | **Design Variables** | | | | | | | |
| | **DOCTOR CLINIC** | 0 | 1 | 0 | 0 | 0 | 0 | 0 | 0 |
| | **FIRE RESQUE OR EMS** | 0 | 0 | 1 | 0 | 0 | 0 | 0 | 0 |
| | **FQHC OR RHC OR CHC** | 0 | 0 | 0 | 1 | 0 | 0 | 0 | 0 |
| | **HOSPITAL CLINIC** | 0 | 0 | 0 | 0 | 1 | 0 | 0 | 0 |
| | **HOSPITAL ER** | 0 | 0 | 0 | 0 | 0 | 1 | 0 | 0 |
| | **OTHER PHARMACIES** | 0 | 0 | 0 | 0 | 0 | 0 | 1 | 0 |
| | **OTHERS OR MISC** | 0 | 0 | 0 | 0 | 0 | 0 | 0 | 1 |
| | **PUBLIX OR CVS OR WALGREENS** | 0 | 0 | 0 | 0 | 0 | 0 | 0 | 0 |

| Model Convergence Status |
|---|
| Convergence criterion (GCONV=1E-8) satisfied. |

| Model Fit Statistics | | |
|---|---|---|
| **Criterion** | **Intercept Only** | **Intercept and Covariates** |
| **AIC** | 1374131.3 | 1134042.4 |
| **SC** | 1374145.6 | 1134383.7 |
| **-2 Log L** | 1374129.3 | 1133994.4 |

| Testing Global Null Hypothesis: BETA=0 | | | |
|---|---|---|---|
| **Test** | **Chi-Square** | **DF** | **Pr > ChiSq** |
| **Likelihood Ratio** | 240134.940 | 23 | <.0001 |
| **Score** | 381524.600 | 23 | <.0001 |
| **Wald** | 192459.224 | 23 | <.0001 |

| Type 3 Analysis of Effects | | | |
|---|---|---|---|
| **Effect** | **DF** | **Wald Chi-Square** | **Pr > ChiSq** |
| **firstdose** | 1 | 2697.6917 | <.0001 |
| **agenew** | 5 | 68338.2011 | <.0001 |
| **sex2** | 1 | 3318.2261 | <.0001 |
| **RACE4** | 3 | 1240.9571 | <.0001 |
| **ETHNICITY3** | 2 | 13525.3252 | <.0001 |
| **decile_sviscore1** | 1 | 1482.7708 | <.0001 |
| **decile_sviscore4** | 1 | 287.7230 | <.0001 |

### All-cause death adjusted (logistic regression)

#### The LOGISTIC Procedure

| Type 3 Analysis of Effects | | | |
|---|---|---|---|
| Effect | DF | Wald Chi-Square | Pr > ChiSq |
| mortalityratepct | 1 | 6869.3112 | <.0001 |
| Vaccinationsite | 8 | 5925.9229 | <.0001 |

| Analysis of Maximum Likelihood Estimates | | | | | | | |
|---|---|---|---|---|---|---|---|
| Parameter | | DF | Estimate | Standard Error | Wald Chi-Square | Pr > ChiSq | Label |
| Intercept | | 1 | -7.3945 | 0.0226 | 106877.196 | <.0001 | Intercept: death1yr=1 |
| firstdose | 1. PFIZER | 1 | 0.3252 | 0.00626 | 2697.6917 | <.0001 | firstdose 1. PFIZER |
| agenew | 40-49 | 1 | 0.8741 | 0.0276 | 1001.0035 | <.0001 | agenew 40-49 |
| agenew | 50-59 | 1 | 1.6771 | 0.0235 | 5093.3277 | <.0001 | agenew 50-59 |
| agenew | 60-69 | 1 | 2.4726 | 0.0221 | 12545.6553 | <.0001 | agenew 60-69 |
| agenew | 70-79 | 1 | 3.2098 | 0.0218 | 21682.0343 | <.0001 | agenew 70-79 |
| agenew | 80+ | 1 | 4.1402 | 0.0226 | 33652.4581 | <.0001 | agenew 80+ |
| sex2 | F | 1 | -0.3396 | 0.00590 | 3318.2261 | <.0001 | sex2 F |
| RACE4 | BLACK | 1 | -0.0447 | 0.0114 | 15.5076 | <.0001 | RACE4 BLACK |
| RACE4 | OTHERS | 1 | -0.2899 | 0.00877 | 1093.0177 | <.0001 | RACE4 OTHERS |
| RACE4 | UNKNOWN | 1 | -0.1989 | 0.0116 | 291.7276 | <.0001 | RACE4 UNKNOWN |
| ETHNICITY3 | HISPANIC | 1 | -0.6457 | 0.0102 | 3972.9788 | <.0001 | ETHNICITY3 HISPANIC |
| ETHNICITY3 | UNKNOWN | 1 | 0.6593 | 0.00792 | 6936.1063 | <.0001 | ETHNICITY3 UNKNOWN |
| decile_sviscore1 | | 1 | 0.0476 | 0.00124 | 1482.7708 | <.0001 | Rank for Variable SVISCORE1 |
| decile_sviscore4 | | 1 | 0.0203 | 0.00120 | 287.7230 | <.0001 | Rank for Variable SVISCORE4 |
| mortalityratepct | | 1 | 0.0609 | 0.000735 | 6869.3112 | <.0001 | |
| Vaccinationsite | CHD OR DOH | 1 | -0.3317 | 0.00844 | 1544.5653 | <.0001 | Vaccinationsite CHD OR DOH |
| Vaccinationsite | DOCTOR CLINIC | 1 | -0.00467 | 0.00866 | 0.2909 | 0.5897 | Vaccinationsite DOCTOR CLINIC |
| Vaccinationsite | FIRE RESQUE OR EMS | 1 | -0.4815 | 0.0140 | 1186.0068 | <.0001 | Vaccinationsite FIRE RESQUE OR EMS |
| Vaccinationsite | FQHC OR RHC OR CHC | 1 | -0.1732 | 0.0178 | 94.2970 | <.0001 | Vaccinationsite FQHC OR RHC OR CHC |
| Vaccinationsite | HOSPITAL CLINIC | 1 | -0.0961 | 0.0120 | 64.4926 | <.0001 | Vaccinationsite HOSPITAL CLINIC |
| Vaccinationsite | HOSPITAL ER | 1 | -0.2583 | 0.0357 | 52.3887 | <.0001 | Vaccinationsite HOSPITAL ER |
| Vaccinationsite | OTHER PHARMACIES | 1 | 0.3161 | 0.0130 | 592.9255 | <.0001 | Vaccinationsite OTHER PHARMACIES |
| Vaccinationsite | OTHERS OR MISC | 1 | -0.8099 | 0.0161 | 2525.4592 | <.0001 | Vaccinationsite OTHERS OR MISC |

#### The LOGISTIC Procedure

| Odds Ratio Estimates | | | |
|---|---|---|---|
| Effect | Point Estimate | 95% Wald Confidence Limits | |
| firstdose 1. PFIZER vs 2. MODERN | 1.384 | 1.367 | 1.401 |
| agenew 40-49 vs 18-39 | 2.397 | 2.270 | 2.530 |
| agenew 50-59 vs 18-39 | 5.350 | 5.109 | 5.602 |
| agenew 60-69 vs 18-39 | 11.853 | 11.351 | 12.378 |
| agenew 70-79 vs 18-39 | 24.773 | 23.737 | 25.854 |
| agenew 80+ vs 18-39 | 62.813 | 60.095 | 65.654 |
| sex2 F vs M | 0.712 | 0.704 | 0.720 |
| RACE4 BLACK vs WHITE | 0.956 | 0.935 | 0.978 |
| RACE4 OTHERS vs WHITE | 0.748 | 0.736 | 0.761 |
| RACE4 UNKNOWN vs WHITE | 0.820 | 0.801 | 0.839 |
| ETHNICITY3 HISPANIC vs NOT HISPANIC | 0.524 | 0.514 | 0.535 |
| ETHNICITY3 UNKNOWN vs NOT HISPANIC | 1.933 | 1.904 | 1.964 |
| decile_sviscore1 | 1.049 | 1.046 | 1.051 |
| decile_sviscore4 | 1.020 | 1.018 | 1.023 |
| mortalityratepct | 1.063 | 1.061 | 1.064 |
| Vaccinationsite CHD OR DOH vs PUBLIX OR CVS OR WALGREENS | 0.718 | 0.706 | 0.730 |
| Vaccinationsite DOCTOR CLINIC vs PUBLIX OR CVS OR WALGREENS | 0.995 | 0.979 | 1.012 |
| Vaccinationsite FIRE RESQUE OR EMS vs PUBLIX OR CVS OR WALGREENS | 0.618 | 0.601 | 0.635 |
| Vaccinationsite FQHC OR RHC OR CHC vs PUBLIX OR CVS OR WALGREENS | 0.841 | 0.812 | 0.871 |
| Vaccinationsite HOSPITAL CLINIC vs PUBLIX OR CVS OR WALGREENS | 0.908 | 0.887 | 0.930 |
| Vaccinationsite HOSPITAL ER vs PUBLIX OR CVS OR WALGREENS | 0.772 | 0.720 | 0.828 |
| Vaccinationsite OTHER PHARMACIES vs PUBLIX OR CVS OR WALGREENS | 1.372 | 1.337 | 1.407 |
| Vaccinationsite OTHERS OR MISC vs PUBLIX OR CVS OR WALGREENS | 0.445 | 0.431 | 0.459 |

| Association of Predicted Probabilities and Observed Responses | | | |
|---|---|---|---|
| Percent Concordant | 83.1 | Somers' D | 0.694 |
| Percent Discordant | 13.7 | Gamma | 0.717 |
| Percent Tied | 3.2 | Tau-a | 0.016 |
| Pairs | 1.3740853E12 | c | 0.847 |

### All-cause deaths adjusted by Age

#### The LOGISTIC Procedure

**agenew=18-39 years**

| Model Information | |
|---|---|
| **Data Set** | WORK.DOSE1MATCHEDDATAFINAL |
| **Response Variable** | death1yr |
| **Number of Response Levels** | 2 |
| **Model** | binary logit |
| **Optimization Technique** | Fisher's scoring |

| | |
|---|---|
| **Number of Observations Read** | 2896312 |
| **Number of Observations Used** | 2843581 |

| Response Profile | | |
|---|---|---|
| **Ordered Value** | **death1yr** | **Total Frequency** |
| **1** | 0 | 2841269 |
| **2** | 1 | 2312 |

**Probability modeled is death1yr=1.**

**Note:** 52731 observations were deleted due to missing values for the response or explanatory variables.

| Class Level Information | | | | | | | | | |
|---|---|---|---|---|---|---|---|---|---|
| **Class** | **Value** | **Design Variables** | | | | | | | |
| **agenew** | **18-39 years** | | | | | | | | |
| **firstdose** | **1. PFIZER** | 1 | | | | | | | |
| | **2. MODERN** | 0 | | | | | | | |
| **sex2** | **F** | 1 | | | | | | | |
| | **M** | 0 | | | | | | | |
| **RACE4** | **BLACK** | 1 | 0 | 0 | | | | | |
| | **OTHERS** | 0 | 1 | 0 | | | | | |
| | **UNKNOWN** | 0 | 0 | 1 | | | | | |
| | **WHITE** | 0 | 0 | 0 | | | | | |
| **ETHNICITY3** | **HISPANIC** | 1 | 0 | | | | | | |
| | **NOT HISPANIC** | 0 | 0 | | | | | | |
| | **UNKNOWN** | 0 | 1 | | | | | | |
| **Vaccinationsite** | **CHD OR DOH** | 1 | 0 | 0 | 0 | 0 | 0 | 0 | 0 |
| | **DOCTOR CLINIC** | 0 | 1 | 0 | 0 | 0 | 0 | 0 | 0 |
| | **FIRE RESQUE OR EMS** | 0 | 0 | 1 | 0 | 0 | 0 | 0 | 0 |
| | **FQHC OR RHC OR CHC** | 0 | 0 | 0 | 1 | 0 | 0 | 0 | 0 |
| | **HOSPITAL CLINIC** | 0 | 0 | 0 | 0 | 1 | 0 | 0 | 0 |

### All-cause deaths adjusted by Age

#### The LOGISTIC Procedure

**agenew=18-39 years**

| Class Level Information | | | | | | | | | |
|---|---|---|---|---|---|---|---|---|---|
| Class | Value | Design Variables | | | | | | | |
| | HOSPITAL ER | 0 | 0 | 0 | 0 | 0 | 1 | 0 | 0 |
| | OTHER PHARMACIES | 0 | 0 | 0 | 0 | 0 | 0 | 1 | 0 |
| | OTHERS OR MISC | 0 | 0 | 0 | 0 | 0 | 0 | 0 | 1 |
| | PUBLIX OR CVS OR WALGREENS | 0 | 0 | 0 | 0 | 0 | 0 | 0 | 0 |

| Model Convergence Status |
|---|
| Convergence criterion (GCONV=1E-8) satisfied. |

| Model Fit Statistics | | |
|---|---|---|
| Criterion | Intercept Only | Intercept and Covariates |
| AIC | 37522.523 | 36683.883 |
| SC | 37535.383 | 36928.234 |
| -2 Log L | 37520.523 | 36645.883 |

| Testing Global Null Hypothesis: BETA=0 | | | |
|---|---|---|---|
| Test | Chi-Square | DF | Pr > ChiSq |
| Likelihood Ratio | 874.6397 | 18 | <.0001 |
| Score | 842.4060 | 18 | <.0001 |
| Wald | 799.3461 | 18 | <.0001 |

| Type 3 Analysis of Effects | | | |
|---|---|---|---|
| Effect | DF | Wald Chi-Square | Pr > ChiSq |
| firstdose | 1 | 7.5637 | 0.0060 |
| agenew | 0 | . | . |
| sex2 | 1 | 225.3408 | <.0001 |
| RACE4 | 3 | 73.5996 | <.0001 |
| ETHNICITY3 | 2 | 261.9132 | <.0001 |
| decile_sviscore1 | 1 | 71.6390 | <.0001 |
| decile_sviscore4 | 1 | 0.2152 | 0.6427 |
| mortalityratepct | 1 | 0.6252 | 0.4291 |
| Vaccinationsite | 8 | 48.2173 | <.0001 |

#### The LOGISTIC Procedure

**agenew=18-39 years**

| Analysis of Maximum Likelihood Estimates | | | | | | | |
|---|---|---|---|---|---|---|---|
| **Parameter** | | **DF** | **Estimate** | **Standard Error** | **Wald Chi-Square** | **Pr > ChiSq** | **Label** |
| **Intercept** | | 1 | -6.9445 | 0.0660 | 11069.4528 | <.0001 | Intercept: death1yr=1 |
| **firstdose** | **1. PFIZER** | 1 | -0.1290 | 0.0469 | 7.5637 | 0.0060 | firstdose 1. PFIZER |
| **sex2** | **F** | 1 | -0.6472 | 0.0431 | 225.3408 | <.0001 | sex2 F |
| **RACE4** | **BLACK** | 1 | 0.2605 | 0.0604 | 18.6173 | <.0001 | RACE4 BLACK |
| **RACE4** | **OTHERS** | 1 | -0.3744 | 0.0620 | 36.5216 | <.0001 | RACE4 OTHERS |
| **RACE4** | **UNKNOWN** | 1 | -0.2005 | 0.0782 | 6.5763 | 0.0103 | RACE4 UNKNOWN |
| **ETHNICITY3** | **HISPANIC** | 1 | -0.8800 | 0.0621 | 200.5983 | <.0001 | ETHNICITY3 HISPANIC |
| **ETHNICITY3** | **UNKNOWN** | 1 | 0.2263 | 0.0565 | 16.0721 | <.0001 | ETHNICITY3 UNKNOWN |
| **decile_sviscore1** | | 1 | 0.0772 | 0.00913 | 71.6390 | <.0001 | Rank for Variable SVISCORE1 |
| **decile_sviscore4** | | 1 | -0.00408 | 0.00879 | 0.2152 | 0.6427 | Rank for Variable SVISCORE4 |
| **mortalityratepct** | | 1 | 0.2390 | 0.3023 | 0.6252 | 0.4291 | |
| **Vaccinationsite** | **CHD OR DOH** | 1 | -0.00904 | 0.0804 | 0.0126 | 0.9105 | Vaccinationsite CHD OR DOH |
| **Vaccinationsite** | **DOCTOR CLINIC** | 1 | 0.1162 | 0.0615 | 3.5656 | 0.0590 | Vaccinationsite DOCTOR CLINIC |
| **Vaccinationsite** | **FIRE RESQUE OR EMS** | 1 | 0.1006 | 0.1886 | 0.2842 | 0.5940 | Vaccinationsite FIRE RESQUE OR EMS |
| **Vaccinationsite** | **FQHC OR RHC OR CHC** | 1 | 0.2082 | 0.1104 | 3.5580 | 0.0593 | Vaccinationsite FQHC OR RHC OR CHC |
| **Vaccinationsite** | **HOSPITAL CLINIC** | 1 | 0.1523 | 0.0864 | 3.1050 | 0.0781 | Vaccinationsite HOSPITAL CLINIC |
| **Vaccinationsite** | **HOSPITAL ER** | 1 | -0.5473 | 0.2610 | 4.3973 | 0.0360 | Vaccinationsite HOSPITAL ER |
| **Vaccinationsite** | **OTHER PHARMACIES** | 1 | 0.2993 | 0.0718 | 17.3698 | <.0001 | Vaccinationsite OTHER PHARMACIES |
| **Vaccinationsite** | **OTHERS OR MISC** | 1 | -0.5325 | 0.1381 | 14.8605 | 0.0001 | Vaccinationsite OTHERS OR MISC |

| Odds Ratio Estimates | | | |
|---|---|---|---|
| **Effect** | **Point Estimate** | **95% Wald Confidence Limits** | |
| **firstdose 1. PFIZER vs 2. MODERN** | 0.879 | 0.802 | 0.964 |
| **sex2 F vs M** | 0.524 | 0.481 | 0.570 |
| **RACE4 BLACK vs WHITE** | 1.298 | 1.153 | 1.461 |
| **RACE4 OTHERS vs WHITE** | 0.688 | 0.609 | 0.776 |
| **RACE4 UNKNOWN vs WHITE** | 0.818 | 0.702 | 0.954 |
| **ETHNICITY3 HISPANIC vs NOT HISPANIC** | 0.415 | 0.367 | 0.468 |
| **ETHNICITY3 UNKNOWN vs NOT HISPANIC** | 1.254 | 1.123 | 1.401 |
| **decile_sviscore1** | 1.080 | 1.061 | 1.100 |
| **decile_sviscore4** | 0.996 | 0.979 | 1.013 |
| **mortalityratepct** | 1.270 | 0.702 | 2.297 |
| **Vaccinationsite CHD OR DOH vs PUBLIX OR CVS OR WALGREENS** | 0.991 | 0.847 | 1.160 |

### All-cause deaths adjusted by Age

#### The LOGISTIC Procedure

**agenew=18-39 years**

| Odds Ratio Estimates | | | |
|---|---|---|---|
| Effect | Point Estimate | 95% Wald Confidence Limits | |
| Vaccinationsite DOCTOR CLINIC vs PUBLIX OR CVS OR WALGREENS | 1.123 | 0.996 | 1.267 |
| Vaccinationsite FIRE RESQUE OR EMS vs PUBLIX OR CVS OR WALGREENS | 1.106 | 0.764 | 1.600 |
| Vaccinationsite FQHC OR RHC OR CHC vs PUBLIX OR CVS OR WALGREENS | 1.232 | 0.992 | 1.529 |
| Vaccinationsite HOSPITAL CLINIC vs PUBLIX OR CVS OR WALGREENS | 1.165 | 0.983 | 1.380 |
| Vaccinationsite HOSPITAL ER vs PUBLIX OR CVS OR WALGREENS | 0.579 | 0.347 | 0.965 |
| Vaccinationsite OTHER PHARMACIES vs PUBLIX OR CVS OR WALGREENS | 1.349 | 1.172 | 1.553 |
| Vaccinationsite OTHERS OR MISC vs PUBLIX OR CVS OR WALGREENS | 0.587 | 0.448 | 0.770 |

| Association of Predicted Probabilities and Observed Responses | | | |
|---|---|---|---|
| Percent Concordant | 67.2 | Somers' D | 0.345 |
| Percent Discordant | 32.7 | Gamma | 0.345 |
| Percent Tied | 0.0 | Tau-a | 0.001 |
| Pairs | 6569013928 | c | 0.672 |

#### The LOGISTIC Procedure

**agenew=40-59 years**

| Model Information | |
|---|---|
| **Data Set** | WORK.DOSE1MATCHEDDATAFINAL |
| **Response Variable** | death1yr |
| **Number of Response Levels** | 2 |
| **Model** | binary logit |
| **Optimization Technique** | Fisher's scoring |

| | |
|---|---|
| **Number of Observations Read** | 3575201 |
| **Number of Observations Used** | 3510311 |

| Response Profile | | |
|---|---|---|
| **Ordered Value** | **death1yr** | **Total Frequency** |
| **1** | 0 | 3498781 |
| **2** | 1 | 11530 |

**Probability modeled is death1yr=1.**

**Note:** 64890 observations were deleted due to missing values for the response or explanatory variables.

| Class Level Information | | | | | | | | | |
|---|---|---|---|---|---|---|---|---|---|
| **Class** | **Value** | **Design Variables** | | | | | | | |
| **agenew** | **40-59 years** | | | | | | | | |
| **firstdose** | **1. PFIZER** | 1 | | | | | | | |
| | **2. MODERN** | 0 | | | | | | | |
| **sex2** | **F** | 1 | | | | | | | |
| | **M** | 0 | | | | | | | |
| **RACE4** | **BLACK** | 1 | 0 | 0 | | | | | |
| | **OTHERS** | 0 | 1 | 0 | | | | | |
| | **UNKNOWN** | 0 | 0 | 1 | | | | | |
| | **WHITE** | 0 | 0 | 0 | | | | | |
| **ETHNICITY3** | **HISPANIC** | 1 | 0 | | | | | | |
| | **NOT HISPANIC** | 0 | 0 | | | | | | |
| | **UNKNOWN** | 0 | 1 | | | | | | |
| **Vaccinationsite** | **CHD OR DOH** | 1 | 0 | 0 | 0 | 0 | 0 | 0 | 0 |
| | **DOCTOR CLINIC** | 0 | 1 | 0 | 0 | 0 | 0 | 0 | 0 |
| | **FIRE RESQUE OR EMS** | 0 | 0 | 1 | 0 | 0 | 0 | 0 | 0 |
| | **FQHC OR RHC OR CHC** | 0 | 0 | 0 | 1 | 0 | 0 | 0 | 0 |
| | **HOSPITAL CLINIC** | 0 | 0 | 0 | 0 | 1 | 0 | 0 | 0 |

### All-cause deaths adjusted by Age

#### The LOGISTIC Procedure

**agenew=40-59 years**

| Class Level Information | | | | | | | | | |
|---|---|---|---|---|---|---|---|---|---|
| **Class** | **Value** | **Design Variables** | | | | | | | |
| | **HOSPITAL ER** | 0 | 0 | 0 | 0 | 0 | 1 | 0 | 0 |
| | **OTHER PHARMACIES** | 0 | 0 | 0 | 0 | 0 | 0 | 1 | 0 |
| | **OTHERS OR MISC** | 0 | 0 | 0 | 0 | 0 | 0 | 0 | 1 |
| | **PUBLIX OR CVS OR WALGREENS** | 0 | 0 | 0 | 0 | 0 | 0 | 0 | 0 |

| Model Convergence Status |
|---|
| Convergence criterion (GCONV=1E-8) satisfied. |

| Model Fit Statistics | | |
|---|---|---|
| **Criterion** | **Intercept Only** | **Intercept and Covariates** |
| **AIC** | 154892.87 | 148894.65 |
| **SC** | 154905.94 | 149143.00 |
| **-2 Log L** | 154890.87 | 148856.65 |

| Testing Global Null Hypothesis: BETA=0 | | | |
|---|---|---|---|
| **Test** | **Chi-Square** | **DF** | **Pr > ChiSq** |
| **Likelihood Ratio** | 6034.2223 | 18 | <.0001 |
| **Score** | 6195.5357 | 18 | <.0001 |
| **Wald** | 5754.7368 | 18 | <.0001 |

| Type 3 Analysis of Effects | | | |
|---|---|---|---|
| **Effect** | **DF** | **Wald Chi-Square** | **Pr > ChiSq** |
| **firstdose** | 1 | 25.7566 | <.0001 |
| **agenew** | 0 | . | . |
| **sex2** | 1 | 369.7140 | <.0001 |
| **RACE4** | 3 | 108.2520 | <.0001 |
| **ETHNICITY3** | 2 | 1127.1257 | <.0001 |
| **decile_sviscore1** | 1 | 539.6544 | <.0001 |
| **decile_sviscore4** | 1 | 16.7178 | <.0001 |
| **mortalityratepct** | 1 | 1334.5738 | <.0001 |
| **Vaccinationsite** | 8 | 405.9031 | <.0001 |

### All-cause deaths adjusted by Age

#### The LOGISTIC Procedure

**agenew=40-59 years**

| Analysis of Maximum Likelihood Estimates | | | | | | | |
|---|---|---|---|---|---|---|---|
| Parameter | | DF | Estimate | Standard Error | Wald Chi-Square | Pr > ChiSq | Label |
| Intercept | | 1 | -6.1877 | 0.0326 | 35995.5354 | <.0001 | Intercept: death1yr=1 |
| firstdose | 1. PFIZER | 1 | -0.1051 | 0.0207 | 25.7566 | <.0001 | firstdose 1. PFIZER |
| sex2 | F | 1 | -0.3837 | 0.0200 | 369.7140 | <.0001 | sex2 F |
| RACE4 | BLACK | 1 | -0.0561 | 0.0282 | 3.9589 | 0.0466 | RACE4 BLACK |
| RACE4 | OTHERS | 1 | -0.3116 | 0.0309 | 101.4286 | <.0001 | RACE4 OTHERS |
| RACE4 | UNKNOWN | 1 | -0.1579 | 0.0355 | 19.7315 | <.0001 | RACE4 UNKNOWN |
| ETHNICITY3 | HISPANIC | 1 | -0.8612 | 0.0302 | 813.9516 | <.0001 | ETHNICITY3 HISPANIC |
| ETHNICITY3 | UNKNOWN | 1 | 0.2827 | 0.0266 | 113.1382 | <.0001 | ETHNICITY3 UNKNOWN |
| decile_sviscore1 | | 1 | 0.0964 | 0.00415 | 539.6544 | <.0001 | Rank for Variable SVISCORE1 |
| decile_sviscore4 | | 1 | 0.0163 | 0.00398 | 16.7178 | <.0001 | Rank for Variable SVISCORE4 |
| mortalityratepct | | 1 | 0.6450 | 0.0177 | 1334.5738 | <.0001 | |
| Vaccinationsite | CHD OR DOH | 1 | -0.1088 | 0.0348 | 9.7537 | 0.0018 | Vaccinationsite CHD OR DOH |
| Vaccinationsite | DOCTOR CLINIC | 1 | 0.1117 | 0.0267 | 17.4938 | <.0001 | Vaccinationsite DOCTOR CLINIC |
| Vaccinationsite | FIRE RESQUE OR EMS | 1 | -0.2631 | 0.0799 | 10.8521 | 0.0010 | Vaccinationsite FIRE RESQUE OR EMS |
| Vaccinationsite | FQHC OR RHC OR CHC | 1 | 0.0651 | 0.0483 | 1.8133 | 0.1781 | Vaccinationsite FQHC OR RHC OR CHC |
| Vaccinationsite | HOSPITAL CLINIC | 1 | 0.4869 | 0.0331 | 216.6635 | <.0001 | Vaccinationsite HOSPITAL CLINIC |
| Vaccinationsite | HOSPITAL ER | 1 | -0.3593 | 0.0963 | 13.9364 | 0.0002 | Vaccinationsite HOSPITAL ER |
| Vaccinationsite | OTHER PHARMACIES | 1 | 0.2494 | 0.0334 | 55.6424 | <.0001 | Vaccinationsite OTHER PHARMACIES |
| Vaccinationsite | OTHERS OR MISC | 1 | -0.3678 | 0.0554 | 44.0312 | <.0001 | Vaccinationsite OTHERS OR MISC |

| Odds Ratio Estimates | | | |
|---|---|---|---|
| Effect | Point Estimate | 95% Wald Confidence Limits | |
| firstdose 1. PFIZER vs 2. MODERN | 0.900 | 0.864 | 0.938 |
| sex2 F vs M | 0.681 | 0.655 | 0.709 |
| RACE4 BLACK vs WHITE | 0.945 | 0.895 | 0.999 |
| RACE4 OTHERS vs WHITE | 0.732 | 0.689 | 0.778 |
| RACE4 UNKNOWN vs WHITE | 0.854 | 0.797 | 0.916 |
| ETHNICITY3 HISPANIC vs NOT HISPANIC | 0.423 | 0.398 | 0.448 |
| ETHNICITY3 UNKNOWN vs NOT HISPANIC | 1.327 | 1.259 | 1.398 |
| decile_sviscore1 | 1.101 | 1.092 | 1.110 |
| decile_sviscore4 | 1.016 | 1.009 | 1.024 |
| mortalityratepct | 1.906 | 1.841 | 1.973 |
| Vaccinationsite CHD OR DOH vs PUBLIX OR CVS OR WALGREENS | 0.897 | 0.838 | 0.960 |

#### The LOGISTIC Procedure

**agenew=40-59 years**

| Odds Ratio Estimates | | | |
|---|---|---|---|
| Effect | Point Estimate | 95% Wald Confidence Limits | |
| Vaccinationsite DOCTOR CLINIC vs PUBLIX OR CVS OR WALGREENS | 1.118 | 1.061 | 1.178 |
| Vaccinationsite FIRE RESQUE OR EMS vs PUBLIX OR CVS OR WALGREENS | 0.769 | 0.657 | 0.899 |
| Vaccinationsite FQHC OR RHC OR CHC vs PUBLIX OR CVS OR WALGREENS | 1.067 | 0.971 | 1.173 |
| Vaccinationsite HOSPITAL CLINIC vs PUBLIX OR CVS OR WALGREENS | 1.627 | 1.525 | 1.736 |
| Vaccinationsite HOSPITAL ER vs PUBLIX OR CVS OR WALGREENS | 0.698 | 0.578 | 0.843 |
| Vaccinationsite OTHER PHARMACIES vs PUBLIX OR CVS OR WALGREENS | 1.283 | 1.202 | 1.370 |
| Vaccinationsite OTHERS OR MISC vs PUBLIX OR CVS OR WALGREENS | 0.692 | 0.621 | 0.772 |

| Association of Predicted Probabilities and Observed Responses | | | |
|---|---|---|---|
| Percent Concordant | 70.1 | Somers' D | 0.402 |
| Percent Discordant | 29.9 | Gamma | 0.402 |
| Percent Tied | 0.0 | Tau-a | 0.003 |
| Pairs | 40340944930 | c | 0.701 |

### The LOGISTIC Procedure

**agenew=60+ years**

| Model Information | |
|---|---|
| **Data Set** | WORK.DOSE1MATCHEDDATAFINAL |
| **Response Variable** | death1yr |
| **Number of Response Levels** | 2 |
| **Model** | binary logit |
| **Optimization Technique** | Fisher's scoring |

| | |
|---|---|
| **Number of Observations Read** | 4816820 |
| **Number of Observations Used** | 4719805 |

| Response Profile | | |
|---|---|---|
| **Ordered Value** | **death1yr** | **Total Frequency** |
| **1** | 0 | 4608139 |
| **2** | 1 | 111666 |

**Probability modeled is death1yr=1.**

**Note:** 97015 observations were deleted due to missing values for the response or explanatory variables.

| Class Level Information | | | | | | | | | |
|---|---|---|---|---|---|---|---|---|---|
| **Class** | **Value** | **Design Variables** | | | | | | | |
| **agenew** | **60+ years** | | | | | | | | |
| **firstdose** | **1. PFIZER** | 1 | | | | | | | |
| | **2. MODERN** | 0 | | | | | | | |
| **sex2** | **F** | 1 | | | | | | | |
| | **M** | 0 | | | | | | | |
| **RACE4** | **BLACK** | 1 | 0 | 0 | | | | | |
| | **OTHERS** | 0 | 1 | 0 | | | | | |
| | **UNKNOWN** | 0 | 0 | 1 | | | | | |
| | **WHITE** | 0 | 0 | 0 | | | | | |
| **ETHNICITY3** | **HISPANIC** | 1 | 0 | | | | | | |
| | **NOT HISPANIC** | 0 | 0 | | | | | | |
| | **UNKNOWN** | 0 | 1 | | | | | | |
| **Vaccinationsite** | **CHD OR DOH** | 1 | 0 | 0 | 0 | 0 | 0 | 0 | 0 |
| | **DOCTOR CLINIC** | 0 | 1 | 0 | 0 | 0 | 0 | 0 | 0 |
| | **FIRE RESQUE OR EMS** | 0 | 0 | 1 | 0 | 0 | 0 | 0 | 0 |
| | **FQHC OR RHC OR CHC** | 0 | 0 | 0 | 1 | 0 | 0 | 0 | 0 |
| | **HOSPITAL CLINIC** | 0 | 0 | 0 | 0 | 1 | 0 | 0 | 0 |

#### The LOGISTIC Procedure

**agenew=60+ years**

| Class Level Information | | | | | | | | | |
|---|---|---|---|---|---|---|---|---|---|
| Class | Value | Design Variables | | | | | | | |
| | HOSPITAL ER | 0 | 0 | 0 | 0 | 0 | 1 | 0 | 0 |
| | OTHER PHARMACIES | 0 | 0 | 0 | 0 | 0 | 0 | 1 | 0 |
| | OTHERS OR MISC | 0 | 0 | 0 | 0 | 0 | 0 | 0 | 1 |
| | PUBLIX OR CVS OR WALGREENS | 0 | 0 | 0 | 0 | 0 | 0 | 0 | 0 |

| Model Convergence Status |
|---|
| Convergence criterion (GCONV=1E-8) satisfied. |

| Model Fit Statistics | | |
|---|---|---|
| Criterion | Intercept Only | Intercept and Covariates |
| AIC | 1056828.4 | 972807.53 |
| SC | 1056841.7 | 973061.51 |
| -2 Log L | 1056826.4 | 972769.53 |

| Testing Global Null Hypothesis: BETA=0 | | | |
|---|---|---|---|
| Test | Chi-Square | DF | Pr > ChiSq |
| Likelihood Ratio | 84056.8273 | 18 | <.0001 |
| Score | 133989.529 | 18 | <.0001 |
| Wald | 83070.6501 | 18 | <.0001 |

| Type 3 Analysis of Effects | | | |
|---|---|---|---|
| Effect | DF | Wald Chi-Square | Pr > ChiSq |
| firstdose | 1 | 2944.2529 | <.0001 |
| agenew | 0 | . | . |
| sex2 | 1 | 879.0080 | <.0001 |
| RACE4 | 3 | 1486.6487 | <.0001 |
| ETHNICITY3 | 2 | 14606.9832 | <.0001 |
| decile_sviscore1 | 1 | 624.1636 | <.0001 |
| decile_sviscore4 | 1 | 383.2833 | <.0001 |
| mortalityratepct | 1 | 60258.8024 | <.0001 |
| Vaccinationsite | 8 | 4413.7747 | <.0001 |

#### The LOGISTIC Procedure

**agenew=60+ years**

| Analysis of Maximum Likelihood Estimates | | | | | | | |
|---|---|---|---|---|---|---|---|
| **Parameter** | | **DF** | **Estimate** | **Standard Error** | **Wald Chi-Square** | **Pr > ChiSq** | **Label** |
| **Intercept** | | 1 | -4.4623 | 0.0100 | 197556.678 | <.0001 | Intercept: death1yr=1 |
| **firstdose** | **1. PFIZER** | 1 | 0.3604 | 0.00664 | 2944.2529 | <.0001 | firstdose 1. PFIZER |
| **sex2** | **F** | 1 | -0.1842 | 0.00621 | 879.0080 | <.0001 | sex2 F |
| **RACE4** | **BLACK** | 1 | -0.2224 | 0.0127 | 304.5433 | <.0001 | RACE4 BLACK |
| **RACE4** | **OTHERS** | 1 | -0.3166 | 0.00971 | 1062.1796 | <.0001 | RACE4 OTHERS |
| **RACE4** | **UNKNOWN** | 1 | -0.2569 | 0.0125 | 425.2459 | <.0001 | RACE4 UNKNOWN |
| **ETHNICITY3** | **HISPANIC** | 1 | -0.7818 | 0.0118 | 4397.6254 | <.0001 | ETHNICITY3 HISPANIC |
| **ETHNICITY3** | **UNKNOWN** | 1 | 0.7318 | 0.00836 | 7658.1965 | <.0001 | ETHNICITY3 UNKNOWN |
| **decile_sviscore1** | | 1 | 0.0328 | 0.00131 | 624.1636 | <.0001 | Rank for Variable SVISCORE1 |
| **decile_sviscore4** | | 1 | 0.0249 | 0.00127 | 383.2833 | <.0001 | Rank for Variable SVISCORE4 |
| **mortalityratepct** | | 1 | 0.1438 | 0.000586 | 60258.8024 | <.0001 | |
| **Vaccinationsite** | **CHD OR DOH** | 1 | -0.2827 | 0.00876 | 1042.0840 | <.0001 | Vaccinationsite CHD OR DOH |
| **Vaccinationsite** | **DOCTOR CLINIC** | 1 | -0.00438 | 0.00932 | 0.2216 | 0.6378 | Vaccinationsite DOCTOR CLINIC |
| **Vaccinationsite** | **FIRE RESQUE OR EMS** | 1 | -0.4072 | 0.0145 | 791.6414 | <.0001 | Vaccinationsite FIRE RESQUE OR EMS |
| **Vaccinationsite** | **FQHC OR RHC OR CHC** | 1 | -0.2076 | 0.0195 | 113.5646 | <.0001 | Vaccinationsite FQHC OR RHC OR CHC |
| **Vaccinationsite** | **HOSPITAL CLINIC** | 1 | -0.1727 | 0.0132 | 172.5448 | <.0001 | Vaccinationsite HOSPITAL CLINIC |
| **Vaccinationsite** | **HOSPITAL ER** | 1 | -0.3263 | 0.0388 | 70.5996 | <.0001 | Vaccinationsite HOSPITAL ER |
| **Vaccinationsite** | **OTHER PHARMACIES** | 1 | 0.2480 | 0.0144 | 298.1898 | <.0001 | Vaccinationsite OTHER PHARMACIES |
| **Vaccinationsite** | **OTHERS OR MISC** | 1 | -0.8143 | 0.0172 | 2247.1367 | <.0001 | Vaccinationsite OTHERS OR MISC |

| Odds Ratio Estimates | | | |
|---|---|---|---|
| **Effect** | **Point Estimate** | **95% Wald Confidence Limits** | |
| **firstdose 1. PFIZER vs 2. MODERN** | 1.434 | 1.415 | 1.453 |
| **sex2 F vs M** | 0.832 | 0.822 | 0.842 |
| **RACE4 BLACK vs WHITE** | 0.801 | 0.781 | 0.821 |
| **RACE4 OTHERS vs WHITE** | 0.729 | 0.715 | 0.743 |
| **RACE4 UNKNOWN vs WHITE** | 0.773 | 0.755 | 0.793 |
| **ETHNICITY3 HISPANIC vs NOT HISPANIC** | 0.458 | 0.447 | 0.468 |
| **ETHNICITY3 UNKNOWN vs NOT HISPANIC** | 2.079 | 2.045 | 2.113 |
| **decile_sviscore1** | 1.033 | 1.031 | 1.036 |
| **decile_sviscore4** | 1.025 | 1.023 | 1.028 |
| **mortalityratepct** | 1.155 | 1.153 | 1.156 |
| **Vaccinationsite CHD OR DOH vs PUBLIX OR CVS OR WALGREENS** | 0.754 | 0.741 | 0.767 |

#### The LOGISTIC Procedure

**agenew=60+ years**

| Odds Ratio Estimates | | | |
|---|---|---|---|
| Effect | Point Estimate | 95% Wald Confidence Limits | |
| Vaccinationsite DOCTOR CLINIC vs PUBLIX OR CVS OR WALGREENS | 0.996 | 0.978 | 1.014 |
| Vaccinationsite FIRE RESQUE OR EMS vs PUBLIX OR CVS OR WALGREENS | 0.666 | 0.647 | 0.685 |
| Vaccinationsite FQHC OR RHC OR CHC vs PUBLIX OR CVS OR WALGREENS | 0.813 | 0.782 | 0.844 |
| Vaccinationsite HOSPITAL CLINIC vs PUBLIX OR CVS OR WALGREENS | 0.841 | 0.820 | 0.863 |
| Vaccinationsite HOSPITAL ER vs PUBLIX OR CVS OR WALGREENS | 0.722 | 0.669 | 0.779 |
| Vaccinationsite OTHER PHARMACIES vs PUBLIX OR CVS OR WALGREENS | 1.281 | 1.246 | 1.318 |
| Vaccinationsite OTHERS OR MISC vs PUBLIX OR CVS OR WALGREENS | 0.443 | 0.428 | 0.458 |

| Association of Predicted Probabilities and Observed Responses | | | |
|---|---|---|---|
| Percent Concordant | 74.4 | Somers' D | 0.488 |
| Percent Discordant | 25.6 | Gamma | 0.488 |
| Percent Tied | 0.0 | Tau-a | 0.023 |
| Pairs | 514572449574 | c | 0.744 |
